# Strain-resolved analysis in a randomized trial of antibiotic pretreatment and maintenance dose delivery mode with fecal microbiota transplant for ulcerative colitis

**DOI:** 10.1101/2021.08.07.21261556

**Authors:** Byron J. Smith, Yvette Piceno, Martin Zydek, Bing Zhang, Lara Aboud Syriani, Jonathan P. Terdiman, Zain Kassam, Averil Ma, Susan V. Lynch, Katherine S. Pollard, Najwa El-Nachef

**Affiliations:** The Gladstone Institute of Data Science and Biotechnology, San Francisco, CA; Department of Epidemiology and Biostatistics, University of California, San Francisco, CA; Symbiome, Inc., San Francisco, CA; Division of Gastroenterology, University of California, San Francisco, CA; College of Osteopathic Medicine of the Pacific, Western University of Health Sciences, Pomona, CA; Finch Therapeutics, Somerville, MA; Department of Medicine, University of California, San Francisco, CA; Benioff Center for Microbiome Medicine, University of California, San Francisco, CA; Chan-Zuckerberg Biohub, San Francisco, CA

## Abstract

Fecal microbiota transplant is a promising therapy for ulcerative colitis. Parameters maximizing effectiveness and tolerability are not yet clear, and it is not known how import the transmission of donor microbes to patients is. Here (clinicaltrails.gov: NCT03006809) we have tested the effects of antibiotic pretreatment and compared two modes of maintenance dose delivery, capsules versus enema, in a randomized, pilot, open-label, 2×2 factorial design with 22 patients analyzed with mild to moderate UC. Clinically, the treatment was well-tolerated with favorable safety profile. Of patients who received antibiotic pretreatment, 6 of 11 experienced remission after six weeks of treatment, versus 2 of 11 non-pretreated patients (odds ratio: 1.69, 95% confidence interval: -0.25 to 3.62). No significant differences were found between maintenance dosing via capsules versus enema. In exploratory analyses, microbiome turnover at both the species and strain levels was extensive and significantly more pronounced in the pretreated patients. Associations were also revealed between taxonomic turnover and changes in the composition of primary and secondary bile acids. Together these findings suggest that antibiotic pretreatment contributes to microbiome engraftment and possibly clinical effectiveness, and validate longitudinal strain tracking as a powerful way to monitor the dynamics and impact of microbiota transfer.

## Introduction

Fecal microbiota transplant (FMT) is recognized as a promising therapy for inflammatory bowel disease, in particular ulcerative colitis (UC) [1,2]. According to meta-analyses, half of the UC patients receiving FMT display a response to treatment, and one third achieve clinical remission of their symptoms [3–6]. Across four randomized, placebo-controlled studies (RCTs) [7–10], 28% of FMT and only 9% of placebo recipients achieved remission [4]. Despite this potential, the mechanism by which FMT improves gut health is not known, which hinders attempts to increase its efficacy. In addition, the role of the gut microbiome in the etiology of UC is unsettled; the composition of the gut microbiome is known to differ in UC cases compared to healthy controls but it remains unclear if these differences are a major cause of the disease or a result of underlying inflammation [11,12]. Understanding how FMT facilitates remission may shed light on the underlying biology of UC and potentially enable the development of novel therapies.

Parameters maximizing the efficacy of FMT for UC have not yet been established, and extensive study-to-study variation in treatment protocols has made comparisons challenging. Many studies use antibiotic pretreatment to diminish the patients’ own baseline microbiomes before FMT (e.g. [13,14]), and a recent meta-analysis has found higher efficacy in studies using this approach [6], but none of the four published RCTs used antibiotics in this way [7–10].

Protocols also vary greatly in the number and method of FMT application. Most include an initial dose via colonoscopy, and many follow up with a maintenance dose regimen ranging from just once three weeks after the initial dose [7] to five times a week for two months [9]. Maintenance doses may improve efficacy [6], but their cost and tolerability are substantial challenges. The recent development of FMT capsule formulations may make maintenance dosing logistically easier and more acceptable to patients [15–17]. However, it is unclear if FMT capsules, administered orally, are as effective as other delivery modes for transferring the donor’s microbiotia. Digestive enzymes, pH fluctuations, bile salts, and other environmental stresses may affect which taxa survive transit through the small bowel. Conversely, donor material may reach the upper colon more effectively when applied by capsule than by enema. While capsules have been shown to be no less effective for the treatment of recurrent *Clostridioides difficile* infections [15,18], it is not yet clear whether they have equivalent efficacy for UC.

Many optimizations of FMT for UC—antibiotic pretreatment, repeated maintenance doses, anaerobic preparation, etc.—assume that effectiveness depends on the sustained transfer of microbial taxa and their functional capacities [19], although this has not been confirmed. Likewise, it is not clear whether a subset of donor communities might be particularly effective. In fact, features of the microbiome, in either patients or their donors, that predict FMT efficacy have not been convincingly identified [19–21]. An important challenge for understanding the role of engraftment is in distinguishing taxa originating in donors’ or recipients’ microbiomes [22,23]. While statistical approaches like SourceTracker, which models samples as blends of taxa from multiple sources, have been effectively applied to both 16S rRNA gene [24] and metagenomic surveys [25], most tools for taxonomic analysis of microbial communities cannot differentiate between donor and patient populations of the same species, due to limited taxonomic resolution [26]. The application of recently developed approaches to identifying and tracking populations at the level of strains rather than species would both increase the detection of transfer events and help elucidate the relationship between community turnover and recovery from UC.

Furthermore, recent work has attempted to identify a link between the metabolic function of the gut microbiota and UC [27]. Numerous results indicate involvement of bile acids (BAs) in IBD and recovery, in particular their transformation to secondary BAs by microbes [28–36]. Paired BA metabolomics with metagenomic sequencing before, during, and after FMT might help to elucidate the importance of these signalling compounds in UC treatment [1,27].

Here we experimentally test the impacts of both antibiotic pretreatment and maintenance dosing protocols on the microbiome and the efficacy of FMT therapy in patients with mild to moderate UC. Along with clinical features, we analyze a longitudinal, multimodal dataset describing the taxonomic and functional gene composition of the microbiota, as well as primary and secondary BAs in stool. Our data comprise a novel resource pairing strain-level taxonomic resolution and BA profiles over a sustained FMT protocol. We find that antibiotic pretreatment substantially increases the transfer of host bacteria to patients and is weakly associated with greater FMT efficacy. Notably, rates of engraftment and clinical remission were similar with both capsules and enemas as maintenance dosing methods. Our results will contribute to optimized protocols for the transfer of donor microbes to patients, as well as improved understanding of the role of the gut microbiome in UC.

## Results

### Study design and patient demographics

We conducted a prospective trial of FMT in patients with active, mild to moderate UC (illustrated in Fig. 1A). Patients were randomized into arms receiving antibiotic pretreatment (ABX+) or not (ABX-), and maintenance doses via either enema (ENMA) or capsules (CAPS). Detailed demographics across arms are available in Supplementary Table S1. For both the initial FMT and maintenance doses, each participant received screened, prepared stool from just one of four healthy donors. Two colonoscopies were performed to assess disease severity and location, one concurrent with the initial FMT application (D0), and the second two weeks after the last maintenance dose (F1).

**Figure 1:**
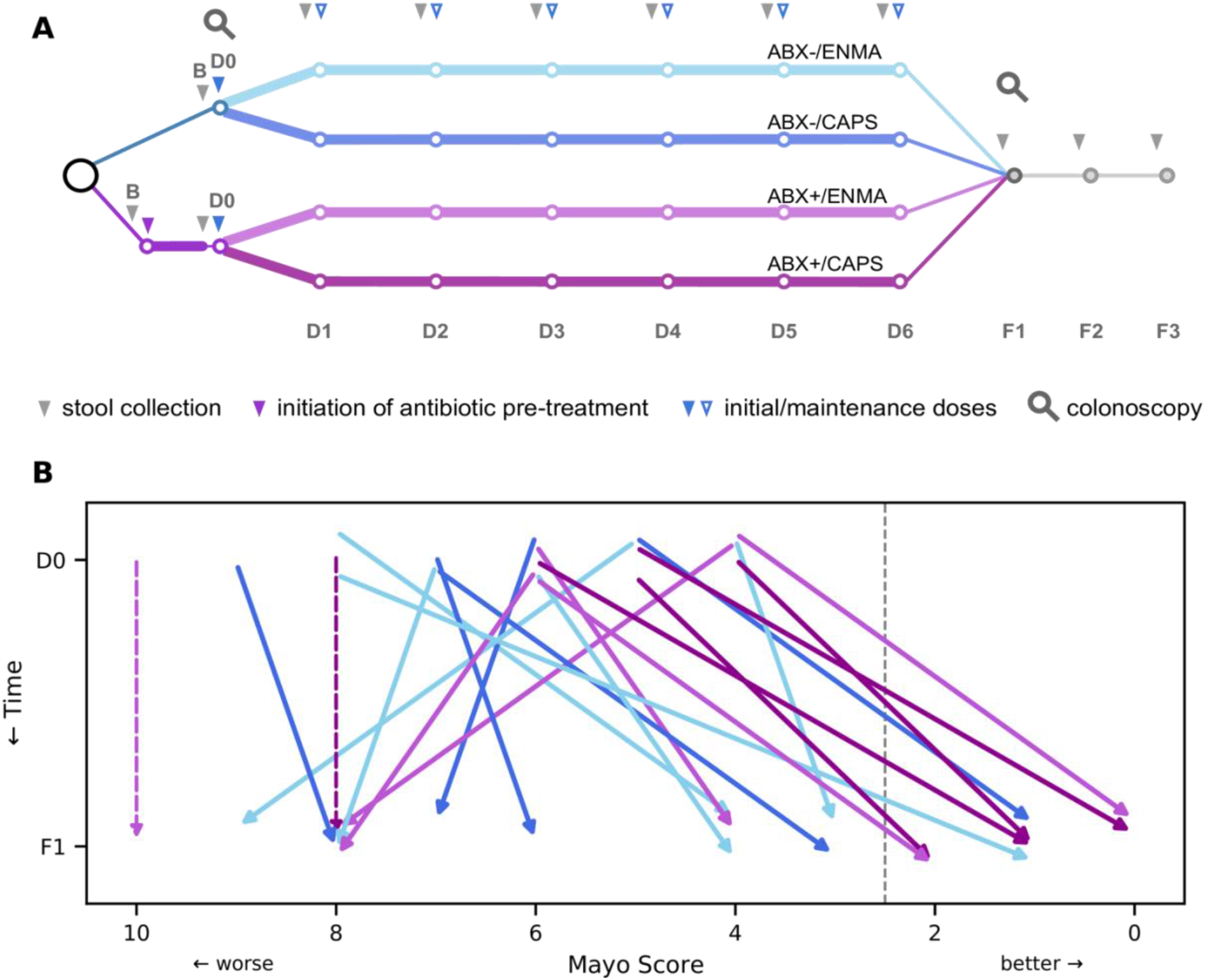
Study design and primary efficacy endpoint of an experimental FMT treatment for UC, showing improved symptoms at follow-up, particularly among patients receiving antibiotics. In our 2×2 factorial design, each patient either received antibiotic pretreatment (ABX+) or not (ABX-), and either received capsules (CAPS) or enema (ENMA) for maintenance dosing. **(A)** Major events during the longitudinal study, including baseline (B) and subsequent fecal sample collections (grey arrowheads), colonoscopies (magnifying glasses), initial FMT (D0, solid blue arrowhead), six weekly maintenance doses (D1-D6, open blue arrowheads), and three follow-up appointments (F1-F3). The start of oral antibiotics is represented (purple arrowhead). **(B)** Change in Mayo scores of each patient between D0 (top) and F1 (bottom). Dashed arrows show the D0 Mayo score of patients who withdrew from the study due to worsening symptoms. Patient arrows are colored by arm and shifted vertically by a small, arbitrary amount to increase visual distinction. The grey dotted line just left of Mayo score 2 marks the remission threshold. All patients with a Mayo score ≤2 at F1 also had an endoscopy sub-score that decreased by at least 1 point; therefore, all were considered to be in remission.

Patients sampled their own stools before the initiation of therapy (baseline sample, B), after the end of antibiotic pretreatment, before each of the six maintenance doses (D1-6), and before each of the three follow-up appointments (F1-3). Maintenance doses were administered weekly, and follow-up appointments scheduled approximately 2 weeks, 6 weeks, and 14 weeks after the last maintenance dose.

Patients were interviewed for the occurrence of solicited adverse events during each study visit and up to 12 months after completion of the study. Minor events included self-limited abdominal pain, constipation, diarrhea, vomiting, abdominal discomfort, excessive flatulence and fever. Notably, of the three occurrences of vomiting in the CAPS arms, all were linked to the same donor (D0485) and happened within a few hours after administration of capsules. No aspiration events occurred. Five patients experienced worsening UC during the study period. Two withdrew due to a need for escalation of therapy. One UC flare constituted a serious adverse event as it required hospitalization but clinicians identified the withdrawal of steroids as a more likely cause then the study treatment. Adverse events are detailed in Supplementary Table S2.

### Antibiotic pretreatment may increase FMT efficacy

We evaluated the clinical impact of treatment using the Mayo score, which compounds subscores including the severity of rectal bleeding, stool frequency, and an endoscopic assessment of epithelial inflammation [37]. Clinical remission, defined as a total Mayo score ≤2 at the first follow-up (time point F1) and endoscopic improvement by ≥1, was reached in a total of 8 patients out of the 20 who received a follow-up colonoscopy, consistent with past studies of FMT for UC [6]. Of these, 6 of 9 (67%) ABX+ patients achieved remission, while only 2 of 11 (18%) did in the ABX-arms (odds ratio: 2.08, 95% confidence interval: 0.00 - 4.16, p=0.065 by two-sided Fisher exact test). While short of a traditional significance threshold, our results nonetheless suggest increased remission in patients receiving antibiotic pretreatment.

Two additional patients, both in ABX+ arms, dropped out of the study due to a flare of UC symptoms before F1. If these patients are included in statistical analyses, the ABX+ remission rate drops to 55% (6 of 11), which is not significantly different from ABX- (OR: 1.69, 95% CI: - 0.25 - 3.62, p=0.18). Tests of other treatment covariates—maintenance method and donor— showed no significant effects, and more complicated models (e.g. those involving multiple covariates and interactions) could not be fit given the small sample size and large number of covariates. Patients were classified as “responders”, a less stringent designation than remission, if their total Mayo score decreased by 3 or more points. Ten of 22 patients were responders, with no statistical association with pretreatment, maintenance method, or donor, tested individually by logistic regression.

Overall, Mayo scores decreased from initial FMT (time point D0) to first follow up (F1; p=0.015 by Wilcoxon signed-rank test, Fig. 1B). However, the magnitude of this change did not appear to be related to any of the treatment covariates. For instance, ABX+ patients did not experience a greater reduction in Mayo score than did ABX-patients, despite the trend towards a higher rate of remission reported above. Since two of four Mayo subscores—for stool frequency and rectal bleeding—were assessed at multiple time points, we analyzed the effect of treatment parameters on each of these during maintenance and follow-up under a repeated measures framework. We found no effect of antibiotics or maintenance methods on either sub-score. There was a significant donor effect on rectal bleeding (p<1e-3 by LRT in a General Estimating Equations framework), but not on stool frequency (p=0.12). Larger studies may be necessary to detect subtle effects of treatment on Mayo scores.

### Metagenomics and metabolomics generate complementary representations of patients’ microbiomes before, during, and after treatment

In order to document the impacts of FMT on the gut microbiome, we longitudinally profiled the taxonomic composition and functional capacity of the fecal community in a subset of patients using several complementary methods. Surveys of the 16S rRNA taxonomic marker gene were paired with shotgun metagenomic sequencing to establish species-level, strain-level, and functional gene composition. In addition, primary and secondary BA profiles were collected via untargeted metabolomics. Details of which samples and profiles were collected for each patient are available in Supplementary Tables S3-6.

Sequencing surveys of the 16S rRNA gene revealed 857 unique amplicon sequence variants (ASVs). In addition, shotgun metagenomic libraries metagenotyped with GT-PRO [38] identified 371 species. To increase the resolution of the taxonomic data, we leveraged single nucleotide polymorphisms (SNPs) as a means to distinguish strains within the broader species categories. Using Strain Finder [22], we identified 3846 strains with detectable abundance in at least two samples. The larger number of strains than ASVs or metagenomic species suggests that strain-level resolution can better differentiate the taxa transferred from donors from those present in patients before FMT.

Functional gene composition was characterized by annotating shotgun metagenomic reads with KEGG gene families (KOs), which identified 7587 unique annotations. Metabolomics identified and quantified a total of 51 primary and secondary BAs across all samples, reflecting the remarkable chemical diversity of these molecules.

### Microbiome features cluster by both patient and donor after FMT

To determine the extent to which FMT modified the patients’ microbiomes, we examined the clustering of genomic and metabolic profiles by patient and by donor during and after FMT treatment (see Fig. 2).

**Figure 2:**
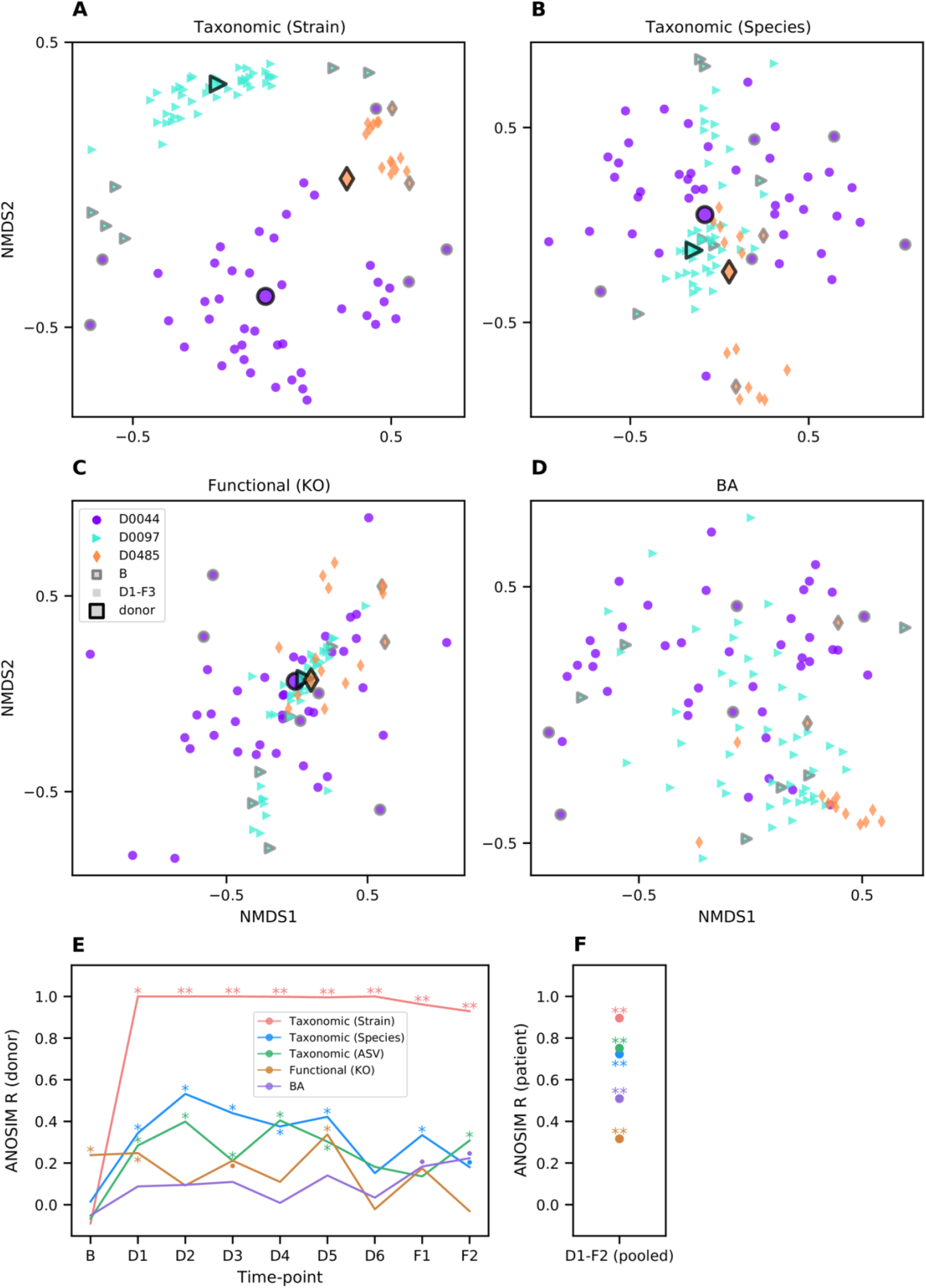
Samples cluster within patient and donor groups during and after FMT treatment. **(A-D)** Non-metric multidimensional scalings were calculated from pairwise sample comparisons based on BC dissimilarity of taxonomic profiles from shotgun metagenomes at strain (A), and species resolution (B), and on cosine dissimilarity of functional gene (C) and BA profiles (D). The orientations and scales of axes are arbitrary, and proximity on a plot reflects similarity. Markers represent individual fecal samples from patients (no black outline) or the mean of all samples from a single donor (black outlines). Shapes and colors are matched between patients and their respective donors. Patients’ baseline samples are outlined in grey. Identical ordinations colored by individual subjects are available in Supplementary Fig. S1. **(E, F)** ANOSIM R scores, an index of clustering strength based on pairwise sample dissimilarities, for the four profiles from (A-D) as well as an additional taxonomic profile based on 16S rRNA gene ASVs. Larger R values indicate stronger clustering by donor at each time point (E) or by patient across pooled time points (F). Significance, as assessed by ANOSIM permutation test (n=9999): p≤0.1 (•), p≤0.05 (*), p≤0.001 (**).

Clustering by donor was detected for several of the profiles (Fig. 2A-E), demonstrating the impacts of FMT. Specifically, strain profiles clustered robustly by donor at every time point after the initial colonoscopy, while—unsurprisingly—baseline samples showed no such clustering. Species profiles, both metagenomic and ASV based, exhibited weaker but also significant clustering by donor during maintenance and follow-up time points. For functional gene annotations, donor clustering was significant, though far less dramatic. However, since this clustering was seen even with baseline samples (ANOSIM R=0.24, p=0.038), clustering at D1 (R=0.25, p=0.048), D3 (R=0.21, p=0.072), and D5 (R=0.34, p=0.017) might be spurious. The limited clustering of functional gene profiles by donor suggests that these are either highly similar across donors or less effectively transferred to patients.

BA profiles showed some signs of clustering by donor at the F1 and F2 time points (R=0.18, 0.22, and p=0.094, 0.060 respectively), raising the interesting possibility that FMT treatment could have a donor-dependent impact on patients’ BA profiles. However, this is not a substantial or significant effect; given the small sample size, it is not clear that it reflects a reproducible result. Other sources of variation in microbiome profiles may reduce the power of this analysis to detect clustering by donor. For example, BA metabolism is known to vary by subject sex [39] (although we did not detect this in our study; see Supplementary Fig. S2).

Despite the perturbation of FMT treatment, clustering by patient across time points was observed for all four microbiome profiles (Fig. 2F, Supplementary Fig. S1). This effect was strongest at the strain level (R=0.90, p<0.001), followed by species level (R=0.72, p<0.001). Intriguingly, both functional annotations (R=0.32, p<0.001) and BA profiles (R=0.52, p<0.001) clustered significantly by patient, suggesting that FMT was unable to uniformly affect these aspects of patients’ microbiomes, and therefore patient-specific effects may modulate the impacts of FMT.

### Taxonomic analysis reveals rapid and extensive transfer of donor species and strains to patients

The clustering of taxonomic profiles by donor indicates effective colonization of patients during FMT. To quantify this transfer, we tracked bacteria species that were specific to patients, specific to their respective donor, or shared by both. Patients had a median of 77 donor species at F1 (representing a median 25% of their total community relative abundance) and 56 (12.3%) at F2. The sensitivity of this approach is limited, however, by the preponderance of shared species, which make up most of the baseline community: median 56% of species and 73% of relative abundance in baseline samples. The fractions of shared taxa estimated from 16S rRNA gene amplicon surveys is nearly identical: 55% of ASVs comprising 71% of relative abundance.

By contrast, only a median of 24% of strains were shared at baseline. Using strains therefore creates more opportunities to infer transfer and engraftment and greatly improves the taxonomic resolution of our analyses (see Fig. 3, Supplementary Fig. S3, and Supplementary Fig. S4). Overall, patients became more populated with donor strains over the course of the study (Fig. 3C). Patients had a median of 260 donor strains at F1 (57% of relative abundance), and 232 (50%) at F2, about six weeks after the end of treatment. The introduction of donor strains is concomitant with patient-specific strains dropping from 87% of total community abundance at baseline (the remaining 13% shared with their donor) to 12% and 20% of the community at F1 and F2, respectively (Fig. 3A). Patients’ BC similarity (1 - BC dissimilarity) to donors based on strain profiles increased from a median of 0.01 at baseline to 0.17 at F1 (p<0.001 by MWU test, (Fig. 3B). Notably, patients’ communities were more similar to their respective donor than to their original composition, with a BC similarity to their own baseline of 0.09 by F1 (Fig. 3D), indicating that FMT profoundly affects the taxonomic composition of patients’ gut microbiomes.

**Figure 3:**
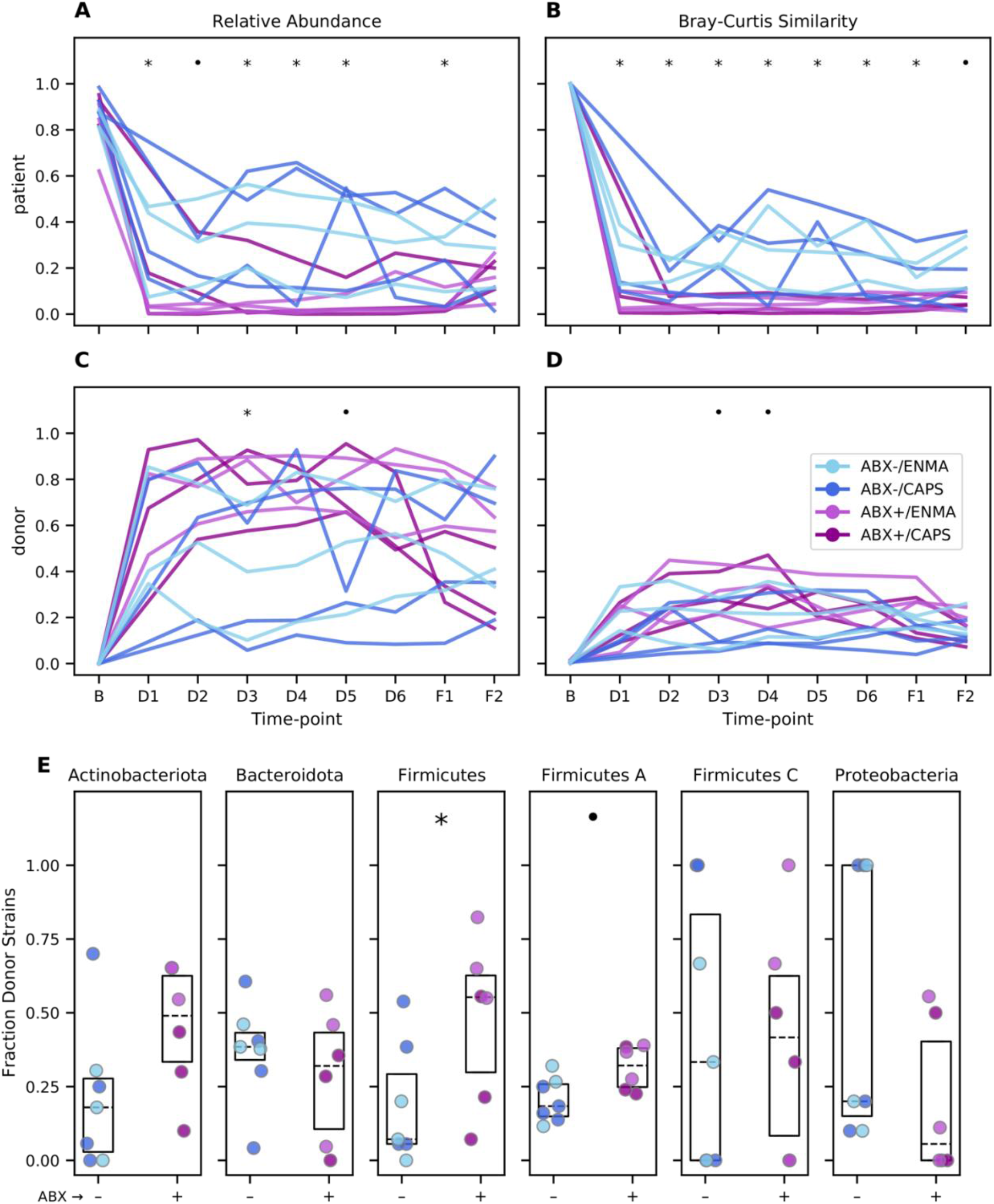
Greater community perturbation and engraftment of donor strains following FMT treatment in antibiotic-pretreated patients. **(A, C)** Abundance of strains specific to each patient’s baseline sample (A) or specific to each donor sample (C) relative to the patient’s total strain populations over the course of treatment. **(B, D)** BC similarity (1 - BC dissimilarity) between each patient’s sample and that patient’s own baseline (B) or their assigned donor (D). **(E)** Fraction of donor strains detected in each patient’s fecal samples at F1. Each dot represents a patient, colors are as in (A-D). Boxes span the interquartile range with the median also marked (dotted line). In all panels, symbols indicate individual time points (A-D) and taxa (E) with p-values less than 0.1 (•) or 0.05 (*) by MWU test for differences between patients who did or did not receive antibiotic pretreatment.

Donor strains were already detected in patients’ fecal community profiles at the D1 time point, prior to the first maintenance dose and just one week after the initial FMT (Fig. 3C). Interestingly, the relative abundance of donor strains, or the BC similarity to the donor community (Fig. 3D), did not substantially increase at subsequent time points (p>0.05 for all comparisons of D1 to D2-F2 by Wilcoxon rank-sum test), suggesting that repeated maintenance doses may have only limited importance in ensuring strain transfer.

Not all bacterial clades were transferred at the same rate from donors to patients. We estimated the overall strain transfer rate as the fraction of all donor-specific strains, summed across all donor/patient pairs, that were subsequently found in patients at F1 (Fig. 3E). This transferability index differed between some of the most abundant phyla—here defined as in the Genome Taxonomy Database (GTDB) [40,41]—from 29% for Actinobacteriota (98 of 339 opportunities) and 26% for Firmicutes A (1138 of 4305 opportunities) to 34% for Bacteroidota (362 of 1062 opportunities, significantly different from Firmicutes A, p<0.001, by two-sided Fisher exact test). Interestingly, the Verrucomicrobiota, represented exclusively by strains of *Akkermansia muciniphila*, had by far the lowest transfer rate at 2.9% (1 of 35 opportunities). Even taking into account the small number of donor-specific strains from the species, this was significantly below the rate for other phyla (p=0.002, 0.035, and 0.011 for Actinobacteriota, Firmicutes A, and Proteobacteria, respectively, and <0.001 for Bacteroidota). *A. muciniphila* has been shown to be depleted in UC patients relative to healthy subjects [42], and to ameliorate colitis in a chemically induced mouse model [43]. While we are hesitant to speculate based on this observation alone, FMT protocols optimized to transfer *A. muciniphila* might show increased remissions rates if the species indeed plays a role in recovery.

Past studies have applied strain tracking data to explore the rules of bacterial engraftment after FMT [22] and examples of coexisting donor and patient strains have been noted [44]. Consistent with this finding, we noted several examples of donor and patient strain coexistence at time point F1 (see Supplementary Fig. S5). A detailed characterization of which strains transfer, coexist, or are excluded from donor communities will be important to explore in future work.

### Antibiotics modulate effects of FMT on the microbiome

We next examined whether maintenance method, donor, and antibiotic pretreatment affected the outcomes of FMT.

Testing each treatment covariate while accounting for this repeated measures design, we found that samples taken during maintenance and follow-up time points from patients who had received antibiotic pretreatment had lower BC similarity to their own baseline (p<0.001 by LRT in the GEE framework), had a smaller total relative abundance of patients’ baseline strains (p<0.001), were more similar to donors (p=0.015), and were composed of a larger relative abundance of their donors’ strains (p=0.015). Similarly, donor was also a significant predictor of all four outcomes: BC similarity to baseline (p=0.021), BC similarity to donor (p<0.001), relative abundance of baseline strains (p<0.001), and relative abundance of donor strains (p<0.001). When testing the effect of antibiotic pretreatment while also controlling for donor, only the BC similarity to baseline and fraction of baseline strains remained significant (p=0.002 and p=0.005, respectively), likely due to the small size of the data and the imbalance in the distribution of treatment arms across donors. Comparing repeated samples from patients that got different maintenance dose formulations showed a small effect (p=0.044) with slightly greater BC similarity to donor for individuals who received enemas, but this effect was not sustained when controlling for donor. Tests carried out at individual time points showed similar patterns (Fig. 3A-D), however, this approach does not harness the increased statistical power of combining multiple observations for each patient.

Overall, a slightly larger fraction of donor-specific bacterial strains was found at F1 in patients receiving antibiotic pretreatment, but this effect did not rise to the level of statistical significance (p=0.134 by MWU). Interestingly, antibiotic pretreatment did not affect the engraftment of all bacterial clades equally (Fig. 3E). When broken down at the phylum level, strains classified as Firmicutes and Firmicutes A—distinct phyla in the GTDB—both showed higher transmission from donors to patients in ABX+ arms (p=0.032 and 0.054, respectively). The phylum Actinobacteriota also shared this trend, although it did not approach statistical significance (p=0.133), and we did not detect an effect for other abundant phyla. Comparisons between maintenance methods (CAPS vs. ENMA) did not reveal differences in strain engraftment for any of the abundant phyla. In total, these results suggest that antibiotic pretreatment increases the engraftment of donor strains, and is likely an effective strategy to optimize for this outcome after FMT.

### Changes in taxonomic profiles after FMT correlate with changes in functional gene and BA profiles

Given the dramatic transfer of donor taxa during FMT, we investigated the extent to which this taxonomic perturbation was concomitant with perturbations of functional gene and BA profiles. To do this, we performed pairwise Mantel tests (see Fig. 4A) among intra-patient dissimilarity matrices comparing taxonomic community composition at family, species, and strain levels to gene family coverages and BA profiles.

**Figure 4:**
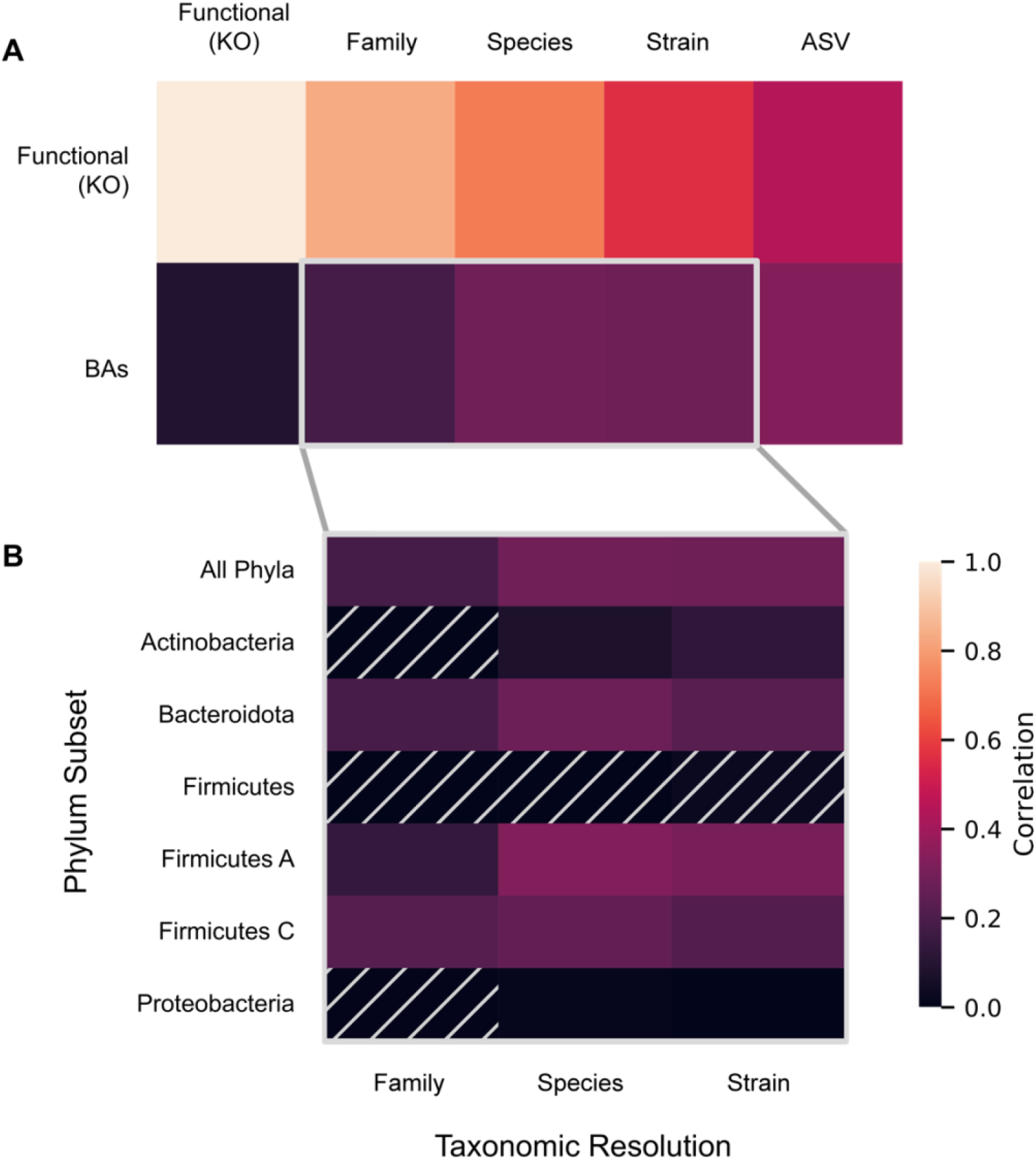
Changes across taxonomic, functional, and BA profiles are correlated. Heatmap tiles depict Pearson correlation coefficients between all within-patient pairwise dissimilarity scores, controlling for time, with brighter colors indicating a stronger association between profiles. Comparisons are for **(A)** each pairwise comparison between taxonomic and functional gene profiles with BA and functional gene profiles, or **(B)** BAs, specifically, compared across taxonomic profiles at different resolutions (X-axis) and phylum-level subsets (Y-axis). Hatched cells indicate comparisons where p-values are >0.05 by Mantel permutation test (n=9999); all other comparisons are significant at or below this threshold.

All correlations with functional gene profiles were statistically significant with partial correlation coefficients greater than 0.5 after controlling for days since FMT. Likewise, the three taxonomic profiles also correlated with BA profiles. Changes in functional gene profiles were only weakly correlated with BA profiles (partial R = 0.09, p=0.045). Although outside the scope of this study, we speculate that this correlation might be higher for the subset of KOs that are related to BA metabolism. Nonetheless, all four correlations together demonstrate that various changes to the microbiome are interrelated during and after FMT.

While the correlation between taxonomic and functional gene profiles increased at higher taxonomic levels (family > species > strain), the opposite was found for BA profiles (family < species ≈ strain). A partial Mantel test did not find a significant correlation with strains after controlling for species (partial R = 0.07, p=0.119). The fact that family-level associations were weaker than associations at either species or strain levels suggests that intra-family differences in association with BAs are likely to be important.

To narrow down aspects of the microbiome with the strongest potential impact on BA profiles, we subsetted the taxonomic composition at the family, species, and strain levels for the six bacterial phyla with the highest average abundance across donors and patients at baseline: the Firmicutes A (54.0%), Bacteroidota (18.3%), Actinobacteriota (10.4%), Firmicutes (8.1%), Proteobacteria (3.6%), and Firmicutes C (2.8%). We then compared changes in each microbial community fraction to changes in BAs while controlling for time (Fig. 4B). We found that at each level most taxonomic subsets were significantly correlated with BA-profile dissimilarities. Interestingly, at the species level the strongest correlation with BAs was found with the highly abundant Firmicutes A fraction. This is consistent with the limited phylogenetic distribution of 7α-dehydroxylation function to this clade [45], understood to be a key step in bacterial BA transformation. However, while for the Firmicutes A, Firmicutes C, Firmicutes, and Bacteroidota fractions the association is *not* stronger at strain-level resolution, the correlation for the Actinobacteriota fraction rose from 0.075 (p = 0.051) at the species resolution to 0.126 (p = 0.013) at strain resolution. Consistent with this, there was a correlation between shifts in BA profiles and shifts in Actinobacteriota strain profiles even after controlling for the Actinobacteriota species profile (partial Mantel R = 0.125, p = 0.038), which could potentially be explained by greater intraspecific variation in BA transformation activity in this phylum, perhaps due to a heterogeneous distribution of bile salt hydrolases in the clade [46]. This result further highlights the importance of strain-level analysis for understanding the drivers of functional differences in microbiomes during FMT.

## Discussion

Here we have experimentally tested the effects of antibiotic pretreatment and maintenance dosing modalities on the clinical and microbiome impacts of FMT for UC. We collected and analyzed a comprehensive, longitudinal, multi-omics dataset, including clinical measurements, taxonomic and functional metagenomics, and BAs from fecal samples taken before, during, and after treatment. While not statistically significant at a traditional p-value threshold, we find a trend towards increased remission rates after FMT in patients receiving antibiotic pretreatment. Similarly, we find significantly increased transmission of donor microbiota after FMT in pretreated patients, and the possibility that pretreatment could result in a greater transfer of microbial functions.

These results contribute to a growing literature on potential optimizations for FMT that increase efficacy, safety, and tolerability of the therapy. Importantly, in this study, maintenance dosing via capsules versus enemas lead to similar strain transmission and remission rates, consistent with previous findings on FMT to treat *C. difficile* infection [15,18,47]. Meta-analyses have shown an association between maintenance dosing and increased remission [6], although the frequency and duration has varied considerably, and previous studies with only two maintenance doses have also demonstrated the effectiveness of FMT therapy for UC [10]. We observe rapid conversion of patients’ microbiomes early in treatment, raising the interesting possibility that a shorter maintenance regimen may be sufficient to elicit sustained engraftment, although this will need to be tested experimentally in future work.

While our findings suggest that antibiotic pretreatment facilitates the sustained transfer of donor microbiota, and this was concomitant with a possible increase in efficacy, we did not find compelling support for the widely assumed importance of this microbial transfer for treatment efficacy. We also did not find clear evidence for differences in the effectiveness of material from different donors (e.g. as described in [48]), although our study was not designed to find subtle effects of this type if they do exist.

This pilot study was limited by the small number of participants completing the full protocol, as well as the large number of primary and exploratory analyses. Without a no-treatment control group, it is also not possible to directly establish the importance of FMT itself for the amelioration of symptoms and microbiome turnover; indeed, it is conceivable that other components of the treatment (e.g. pre-FMT antibiotics and purgatives) independently impacted symptoms and/or strain engraftment. However, the remission rate that we observed is consistent with past FMT trials and substantially larger than with placebo alone [6]. Since donors were assigned based on material availability, some treatment effects were difficult to disentangle from possible donor effects. In addition, reported statistical significance was not corrected for multiple testing. Given these limitations, many of the patterns identified here should be validated in larger designs. Of particular importance, future studies will need mechanisms to account for patients discontinuing the experimental treatment due to worsening symptoms. Interpretation of the metabolomic and metagenomic results presented here is limited—necessarily—by the inclusion of only those patients with a follow-up colonoscopy. Likewise, while our microbiome analyses focused on stool samples, the mucosa-adherent microbiota may be more directly linked to host immunological state [49]. Additional work is needed to analyze patterns of engraftment in the mucosa and across localized sites in the gut. Despite these shortcomings, the application of computational methods designed to maximize the value of our comprehensive, longitudinal dataset enabled the detection of differences between study arms in taxonomic and functional turnover, among other novel insights.

We find that the use of strain haplotypes inferred from SNP profiles [22] greatly improved the sensitivity of our transmission analysis. While this was expected given the large number of shared taxa at the species and 16S rRNA gene amplicon SNV levels, it is surprising that this improvement was possible without tuning hyper-parameters of the Strain Finder algorithm. Independent validation of the haplotypes and strain abundance estimates would be technically challenging, and is outside this study’s scope. However, the conclusions detailed here—most of which result from comparisons of identically analyzed samples—are likely robust to variable accuracy of strain inferences. The SNP deconvolution approach was of particular value in this study, since other available methods for strain tracking (reviewed in [50]) either require high quality strain reference sets [51], or use heuristics based on dissimilarities between metagenotypes that assume species have one dominant strain per sample [52] or ignore the possibility of rare strains (e.g. less than 10% in [53]). None of these alternative approaches is optimal for the tracking of discrete strains in a study where donor and recipient communities are intentionally mixed through FMT. While we focused on the identification of individual strains transmitting from donors to recipients, alternative methods exist [24,25], and we expect that the increased taxonomic resolution offered by SNP deconvolution can contribute greatly to the sensitivity of these as well. Continued development of haplotype deconvolution methods will empower further studies of strain-specific functions and ecological dynamics in FMT.

The conversion of primary bile salts into a diverse assortment of secondary BAs is a microbial process of particular interest in UC, among other conditions. Two findings from this study contribute to a better understanding of how BA profiles are affected by FMT. First, profiles clustered by patient but not significantly by donor. Subject-level clustering after FMT can be observed in prior work [54], and indicates that both host and microbial processing of BAs are remarkably stable; despite robust transfer of donor microbes, our protocol was unable to systematically modulate the concentrations of these metabolites. Second, we nonetheless saw that the magnitude of changes to BA profiles correlated with the extent of taxonomic turnover. These observations suggest that the donor microbiome indeed impacts the patient’s BA profile, but that the direction of this effect may depend on properties of the patient and their microbiome. Together these results invite further inquiry into the role of BAs and other metabolites in UC, and how FMT might be harnessed to modulate them.

## Conclusions

Here we have shown evidence for a potential increase in the remission of UC and the transmission of donor microbes during FMT in patients receiving antibiotic pretreatment. By contrast, we do not find a difference in efficacy or transmission due to the maintenance dosing protocol, suggesting that capsules may be a viable alternative to enemas. We observe patterns consistent with the hypothesis that increased transmission may result in improved outcomes, potentially due to changes in the composition and activity of the gut microbiome. This work demonstrates the increased sensitivity of strain-resolved metagenomic surveys in tracking transmission of donor microbiota, and presents a longitudinal, multi-modal characterization of the microbiome that can inform better-powered clinical trials designed to identify optimal treatment protocols for clinical use.

## Methods

### Patient recruitment, ethics approval, and concurrent therapies

Patients aged 18-64 years with a history of UC confirmed by endoscopy and pathology were recruited at a single academic center and were considered eligible if they had mild to moderate disease activity—a total Mayo score of 4-9 with endoscopic subscore of 1 or 2 assessed by flexible sigmoidoscopy or colonoscopy within 12 months of enrollment and reassessed at the time of initial colonoscopy for FMT delivery. Patients with prior colectomy, severe immunodeficiency, indeterminate colitis, severe UC or history of inflammation limited to distal proctitis (distal 5cm) were excluded from participation. See Supplementary Methods for full inclusion and exclusion criteria. Disease activity was assessed again at time of initial colonoscopy.

All participants were recruited between March 2017 and March 2020 at the University of California, San Francisco, and provided written informed consent for voluntary participation in this protocol which was approved by the UCSF Committee on Human Research (study number: 16-20066). FMT was approved for use for this indication under FDA Investigational New Drug application (IND 16467). This study was registered at Clinicaltrials.gov (NCT03006809, first registered 2016-12-30). All methods were performed in accordance with the relevant guidelines and regulations.

Concurrent therapies were allowed during the course of the trial as long as doses were stable (mesalamine x 4 weeks, immunomodulators x 3 months and biologics x 3 months). However, steroids were minimized to an equivalent dose of no more than 10 mg prednisone/day with forced weaning of 2.5 mg/week during the study period. Additionally, rectal therapy was discontinued 30 days prior to study treatment and probiotics were held six weeks prior to administration of the first FMT dose.

### Study design and clinical details

Participants were randomized into one of four arms by study coordinators by arbitrary selection of unlabeled paper envelopes containing assignments. Unlike study arms, donor material was assigned non-randomly based on availability. Study treatments were administered from September 2017 to March 2020, and the safety follow-up period continued for one year after the end of treatment. Patients assigned to ABX+ arms were pretreated with neomycin, vancomycin and metronidazole 500 mg twice-daily for 5 days, followed by a one-day wash-out period. Patients then underwent colonoscopy to confirm eligibility, receiving their first FMT dose (250 ml) during the procedure. Starting one week later, and over the next six weeks, patients received maintenance doses, 30 capsules weekly for patients in the CAPS arm, or a 60-mL enema weekly for patients in the ENMA arm. Donor stool was provided by OpenBiome, whose screening methods have been previously described [55]. Participants were assigned a single donor for all doses throughout the study period. Deviations from the prescribed design are detailed in Supplementary Table S7.

To minimize invasiveness, baseline endoscopy for patients in ABX+ arms was performed after patients received antibiotics. Past studies have considered antibiotics as a treatment for UC, and conceivably these patients may have already experienced amelioration of symptoms after their enrollment. This possible confounding seems unlikely, however, both because past work has largely not supported the effectiveness of antibiotics [56–59], and due to the short duration of antibiotic treatment in this study.

Baseline data including serum inflammatory markers, infectious stool studies and fecal calprotectin were obtained prior to D0 (the initial FMT) and F1. A full list of clinical assessments is available in Supplementary Table S8. Colonoscopy for endoscopic restaging and repeat biopsies was performed at D0. Adverse events were solicited the day after colonoscopy, weekly during the course of maintenance therapy and then monthly until 6 months after initial FMT and again at 12 months after initial FMT. Remission at F1 was defined as a total Mayo score ≤2 and endoscopic improvement by ≥1 point.

Detailed descriptions of fecal sample collection and processing are included in the Supplementary Methods. Lists of which samples were collected and used for 16S rRNA gene V4 region and shotgun metagenomic library sequencing, as well as untargeted metabolomics are listed in Supplementary Tables S3-S6, respectively.

### Data processing and reproducibility

#### Environment and pipeline

Sequence and metabolite data were analyzed using a reproducible pipeline implemented with the Snakemake workflow manager [60]. Our computational environment is available as a Docker container <https://hub.docker.com/repository/docker/bsmith89/compbio> [61] and uses Conda [62] for most software installations. Final analyses were performed and visualized in Python and R using the Jupyter Notebook environment [63]. Where randomization was used, random number generators were seeded with a fixed value for reproducibility. Detailed descriptions of the data analysis pipeline are included in the Supplementary Methods.

#### Taxonomic profiling

We applied GT-PRO [38] to count the occurences of reads containing exact k-mers representative of previously identified, per-species, bi-allelic positions in the UHGG [64], a comprehensive database of human gut bacterial reference genomes. The coverage (sum of major and minor allele counts) of these SNP sites was then used to estimate the per-sample abundance of each species as the mean of all position coverages observed in any sample, after discarding the 5% highest and lowest coverage positions. This trimmed mean makes our coverage estimate robust to anomalously high or low coverage positions. Relative abundances were then calculated as the coverage of each species divided by the total coverage of all species estimated for that sample.

The GT-PRO metagenotypes were also used to estimate the abundance of haplotypes (referred to as strains above) in each species. This was accomplished using the tool Strain Finder [22], run on the major and minor allele coverages at 100 randomly selected positions. Importantly, due to the scope of this study and computational limitations, we did not rigorously estimate the optimal number of strains in each sample and limited Strain Finder runtime to 60 minutes per species. We therefore may not have reached a global optimum in all cases. Instead we allowed for a maximum of 20 haplotypes to be fit for each species. Fitting the model in this way could result in inferred haplotypes grouping or splitting true strain abundances more than if we had also optimized the number of haplotypes and run the random search algorithm for longer. While this may have reduced the sensitivity of our analyses, we do not believe that it limited the veracity of any of our conclusions, which were primarily based on overall dissimilarity scores between samples and tracking of strains in donor/patient pairs. Fractional abundances of each strain inferred by strain-finder were then scaled by the previously estimated, per-sample species coverages to produce an estimate of strain coverage in each sample.

We demonstrate improvements in sensitivity to transfer events using this approach in the Supplementary Materials.

### Statistical analysis

#### Patient/sample exclusion and efficacy statistics

For all analyses, as many patients and samples were included as possible. For instance, engraftment comparisons at individual time points include all available samples, ignoring patients with missing samples. Metagenomic data for one sample with fewer than 1e6 reads was dropped from analysis, as this is much less than all others. Detailed lists of which microbiome profiles were collected for each patient are available as Supplementary Tables S3-6.

Patients were included in remission comparisons if they had both baseline and follow-up colonoscopies/Mayo scores. Two additional patients that had withdrawn without follow-up colonoscopies were also included as “non-remissions” because they withdrew due to worsening symptoms. Differences in remission rates between treatment groups were calculated for patients pooled into ABX-/ABX+ or CAPS/ENMA groups, and tested using a two-sided Fisher exact test.

Given the small sample size and exploratory analyses, statistical significance was not corrected for multiple testing, throughout.

#### Profile comparisons: Dissimilarities, ordination, clustering analysis

Donor taxonomic and functional profiles were calculated by summing coverage across all samples obtained from that donor—10, 31, and 1 samples with metagenomes for D0044, D0097, and D0485, respectively. For microbial taxonomic profiles, inferred strain coverages in each sample were normalized to sum to one. Then, for higher taxonomic levels, strain relative abundances were summed within assignments provided by the UHGG database, which are based on the GTDB [40,41]. Likewise, for phylum-specific analyses, taxonomic profiles were partitioned based on the UHGG assignment and then renormalized to sum to one. Ordination and cluster analyses were performed on pairwise dissimilarities between sample profiles. For all taxonomic profiles the BC dissimilarity was used, while for functional gene and BA profiles— neither of which is strictly compositional—we used the cosine dissimilarity instead. Ordinations were performed using non-metric multidimensional scaling as implemented in the Scikit-Learn package for Python [65]. Clustering of profiles by donor and patient was tested with ANOSIM using 9999 permutations to calculate a p-value.

#### Longitudinal data analysis

For response variables with repeated measures on the same patients, the effects of treatment parameters were tested under the general estimating equations framework (implemented in the geepack package for R [66]) using a robust, autoregressive covariance structure parameterized by the temporal order of samples. Individual tests were carried out for the effects of antibiotic pretreatment, maintenance dosing method, and donor identity against a null model that included only weeks since initial FMT. Where indicated, donor identity was also included in the null model to test for effects of treatment parameters above and beyond this sometimes-confounding random effect.

#### Mantel tests

Correlations between dissimilarities for pairs of profiles were tested with the Mantel and partial Mantel tests. Specifically, dissimilarity matrices were calculated for baseline, maintenance, and follow-up samples from each patient (post-antibiotic samples were excluded). These matrices were combined as a block-diagonal matrix; no values were included for inter-patient comparisons. Mantel tests were performed using pearson correlations or partial correlations, and p-values were calculated from 9999 permutations. For tests controlling for time, partial correlations were based on the square-root of the time in days between samples being collected. While shortcomings of the Mantel test have been documented [67], the method has been applied in past studies linking different features of the microbiome [68]. In addition, spatial and temporal autocorrelation is expected to result in decreased power, making this approach a conservative measure of associations across data types.

## Supporting information

Code Notebook

Self Sampling Instructions

CONSORT Checklist

## Data Availability

Amplicon sequences from the 16S rRNA gene and metagenomic sequences with human reads removed were uploaded to the SRA under BioProject PRJNA737472 and will be released upon publication. All code and metadata needed to reproduce our results are available at doi:10.5281/zenodo.5851803.

https://github.com/bsmith89/ucfmt2

https://www.ncbi.nlm.nih.gov/bioproject/PRJNA737472

https://doi.org/10.5281/zenodo.5851803

## Additional Information

### Author Contributions

- BJS: conceptualization, data curation, formal analysis, methodology, software, visualization, writing - original draft, writing - review & editing.
- YMP: methodology, investigation, writing - review & editing.
- KSP: conceptualization, methodology, writing - review & editing, funding acquisition, supervision
- LAS: project administration, investigation, resources, writing - review and editing
- NE: conceptualization, patient recruitment/enrollment, methodology, supervision, funding acquisition, writing-review and editing
- MZ: project administration, investigation, data collection, data curation, writing - review and editing
- ZK: conceptualization, methodology, writing - review and editing
- JPT: patient recruitment, review and editing
- AM: funding acquisition, conceptualization
- BZ: data curation, methodology, formal analysis, writing - original draft, writing - review and editing
- SVL: funding acquisition, conceptualization, supervision, writing - review and editing

## Acknowledgements

- BJS and BZ are supported by funding from the National Institutes of Health grant number 5T32DK007007.
- KSP and BJS were supported by funding from National Science Foundation grant number 1563159, Chan Zuckerberg Biohub, and Gladstone Institutes.
- NE, MZ and YMP were supported by funding from the Kenneth Rainin Foundation
- Kole Lynch and Doug Fadrosh prepared libraries and sequenced 16S rRNA gene amplicons.
- Françoise Chanut provided editorial support.

## Data availability

Amplicon sequences from the 16S rRNA gene and metagenomic sequences with human reads removed were uploaded to the SRA under BioProject PRJNA737472. All code and metadata needed to reproduce our results are available at <https://doi.org/10.5281/zenodo.5851803>.

