## Supplementary Materials for "Strain-resolved analysis in a randomized trial of antibiotic pretreatment and maintenance dose delivery mode with fecal microbiota transplant for ulcerative colitis"

### Supplementary Note: Analysis Notebook

Static rendering of Jupyter notebook with analysis/plotting of relevant results.

CodeNotebook.html

### Supplementary Results: Patient Demographics

**Table S1**: Summary of demographics for 22 patients included in efficacy statistics

SupplementaryTablesS1-8.xlsx

### Supplementary Results: Adverse Events

**Table S2**: Summary of adverse events for all patients enrolled. Numbers indicate number of patients experiencing one or more events of each type. Where events are on a three-point scale, higher values mean more severe events.

SupplementaryTablesS1-8.xlsx

### Supplementary Table: Samples and Microbiome Profiles Collected by Patient

**Table S3**: List of fecal samples collected.

SupplementaryTablesS1-8.xlsx

**Table S4**: List of 16S rRNA gene libraries sequenced.

SupplementaryTablesS1-8.xlsx

**Table S5**: List of shotgun metagenomic libraries sequenced.

SupplementaryTablesS1-8.xlsx

**Table S6**: List of metabolomic profiles collected.

SupplementaryTablesS1-8.xlsx

### Supplementary Results: Patients Deviating from Initial Study Design

**Table S7**: Summary of relevant deviations from main protocol.

SupplementaryTablesS1-8.xlsx

### Supplementary Results: Ordinations of microbiome profiles colored by patient

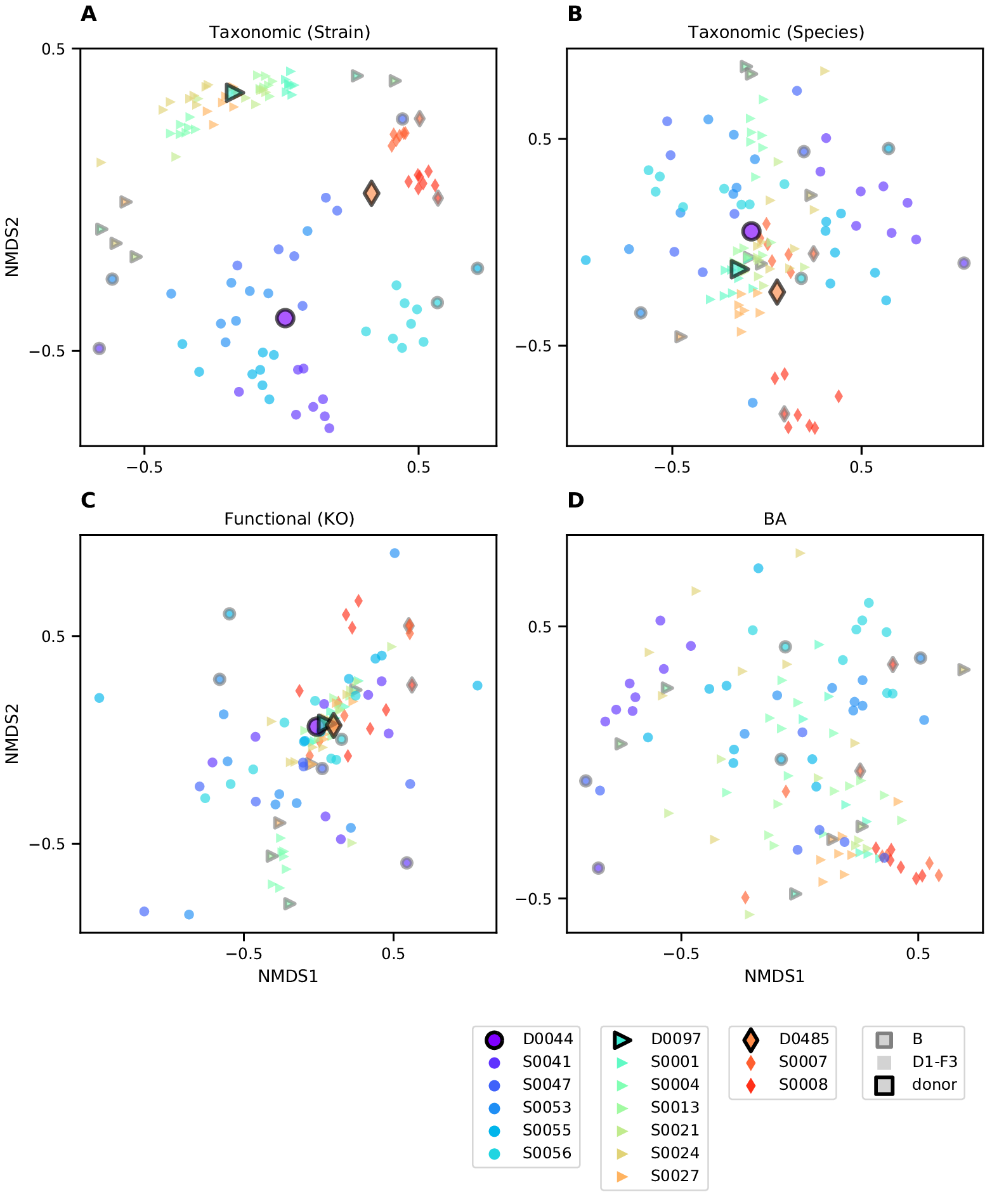

**Figure S1**: Clustering of taxonomic, functional, and BA profiles across time points. Ordinations are calculated and plotted as in Fig. 2. For **(A-C)** datatypes where donor samples were also profiled, larger points with black outlines represent the mean of all samples from that donor. Samples from each subject are differentiated by color and shape as indicated in the legend (bottom left). Patients’ baseline samples are outlined in black. The same four ordinations colored by assigned donors are available as Fig. 2A-D.

### Supplementary Results: Profile clustering by patient sex

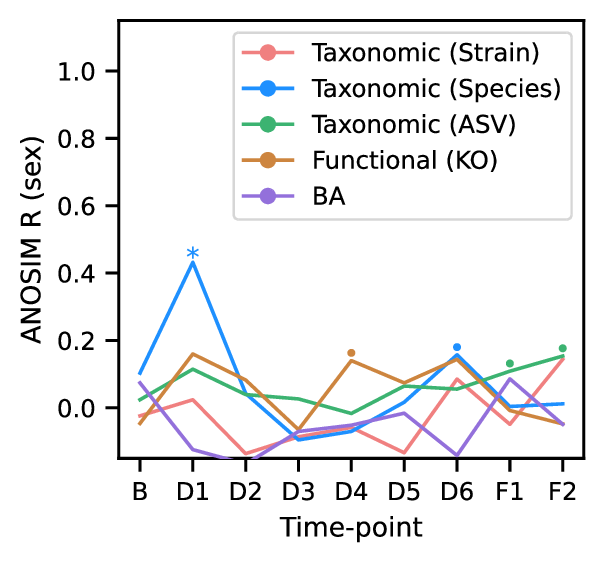

**Figure S2**: Minimal clustering of samples by sex across microbiome profiles. Plot of clustering over time points, as in Fig. 2E, but comparing samples by subject sex instead of donor. ANOSIM R scores, an index of clustering strength based on pairwise sample dissimilarities, are shown for four microbiome profiles as in Fig. 2E. Larger R values indicate stronger clustering by sex at each time point. Significance, as assessed by ANOSIM permutation test (n=9999): p≤0.1 (•), p≤0.05 (*), p≤0.001 (**).

### Supplementary Results: Shared ASVs/Species/Strains across Donors

Comparing taxa across donors presents one way to evaluate the effect of taxonomic resolution on the sensitivity of tracking transfers between individuals. That very few strains are found to be shared across donor communities (see Supplementary Fig. S1) indicates that false positives—non-transfer of a donor strain incorrectly inferred to be a transfer—are infrequent.

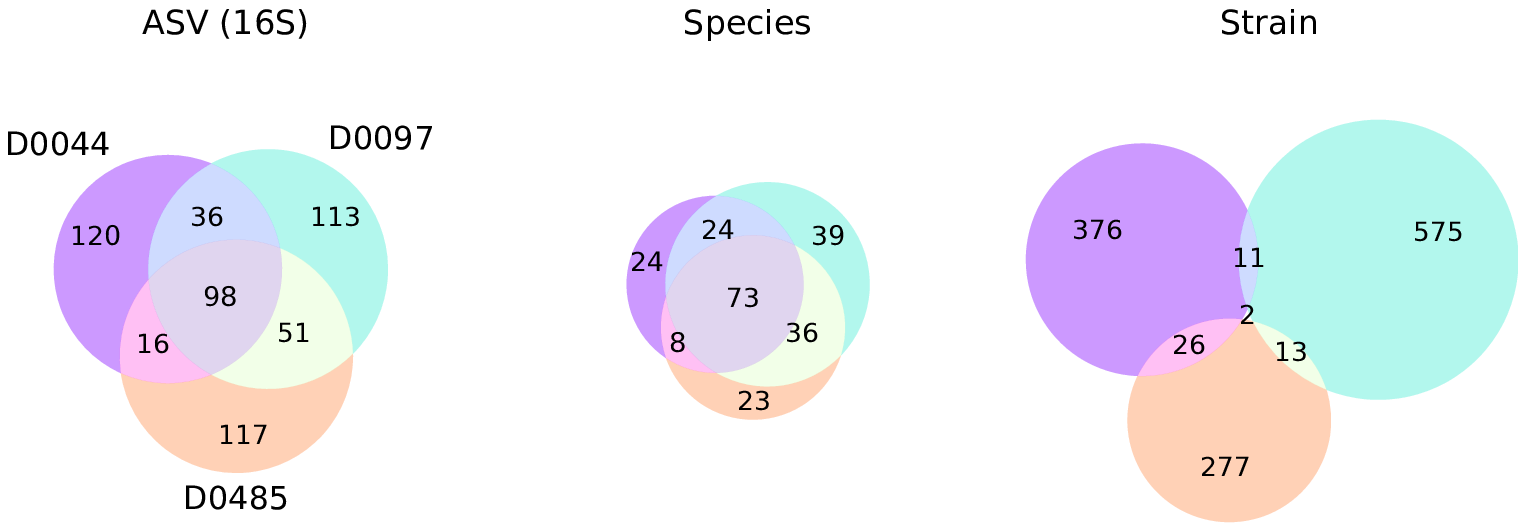

**Figure S3**: Overlap between taxa detected in three donors shows that strain-level taxonomic resolution increases sensitivity and specificity of engraftment detection. Venn diagrams depict relationships between sets of taxa detected in donor samples, and numbers indicate the size of the respective sets. Taxa were considered to be detected in a donor if their mean relative abundance across that donor’s samples was greater than 0.01%. Circles are colored by donor as in Fig. 2. Species composition was estimated from metagenomes based on the mean coverage reported by GT-PRO [38], and was further partitioned into strain composition based on haplotype deconvolution with Strain Finder [22].

### Supplementary Results: Extended Engraftment Analysis

Besides strains identified as specific to a patient’s baseline sample or their donor’s samples, we also categorized strains found in both (“shared”) or not found in either (referred to as “other” strains). This last category represents the possible engraftment of microbes from a patient’s environment. To explore our sensitivity to novel microbes, as well as our ability to differentiate between strains from different sources, we further subdivided these into taxa found in other donors’ samples (“other-donor”), and those not found in the patients baseline nor any of the donors (“other-nodonor”).

Repeating the categorization for taxa surveyed by 16S rRNA gene sequencing or in metagenomc data at the species level, we can compare the three approaches (see Fig. S4). Besides more sensitivity to donor and subject specific taxa, we also find that strain-level analysis assigns a higher fraction of the total relative abundance to “other” strains. Interestingly, most of these are not found in any of the other donors, indicating that our simple approach to identifying engraftment events has a low rate of false-positives.

Interestingly, the subject with the highest relative abundance of “other-donor” strains at time points F1 and F2, in fact received two maintenance doses from a different donor (D0485) instead of their assigned donor (D0044) (see Supplementary Table S7). This anecdote seems to further demonstrate the sensitivity of strains for detecting engraftment due to FMT.

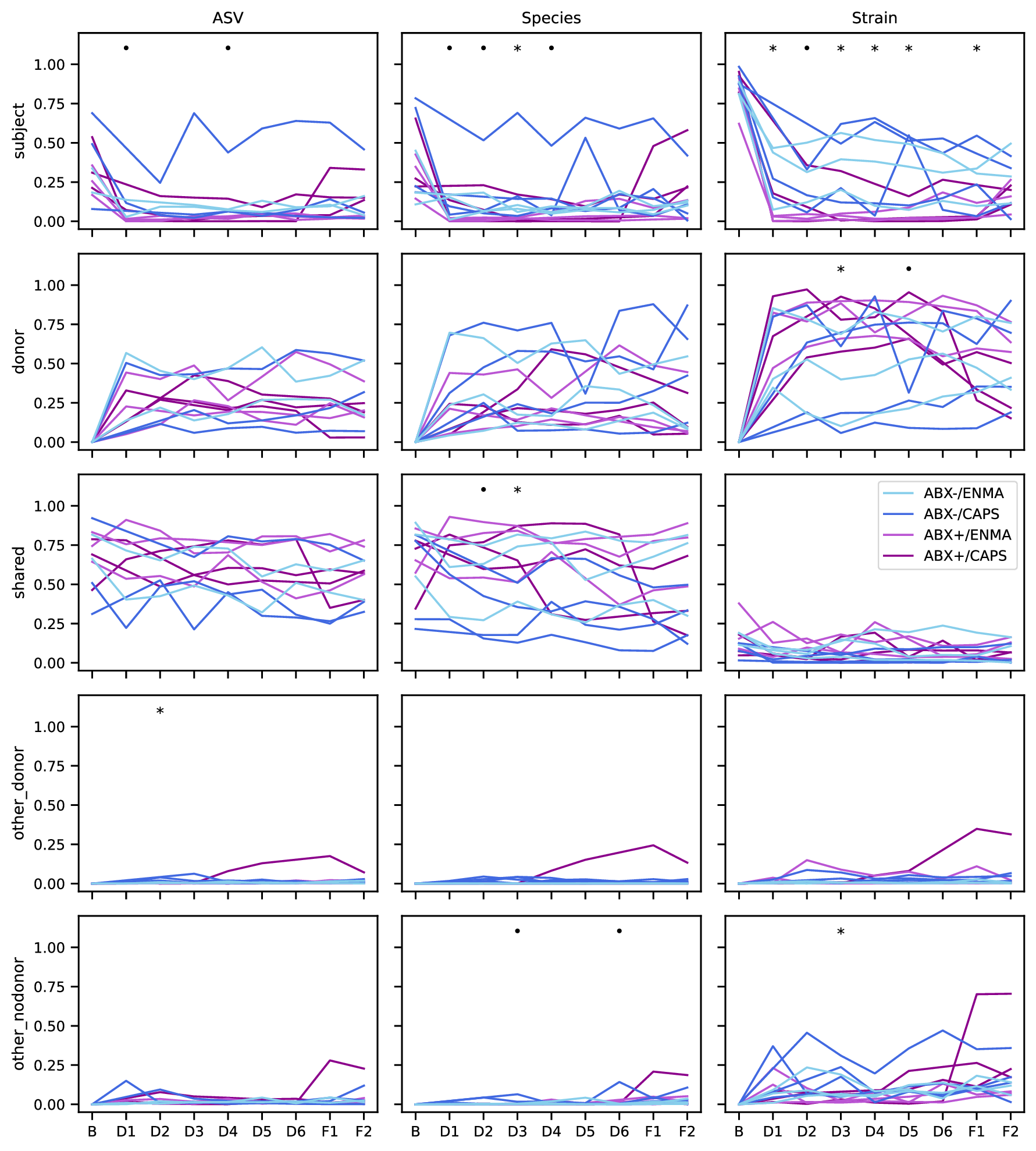

**Figure S4**: Abundance of taxa classified by their presence in each patients baseline as well as the donor samples. Panels in each column share a classification method—16S (ASVs), metagenomic species, or strains within species—and rows share a taxon category: specific to each patient’s baseline sample (“subject” taxa), specific to the assigned donor’s samples (“donor”), found in both (“shared”), found in neither but detected in a different donor (“other_donor”), or not found in the patients baseline nor any of the donors’ samples (“other_nodonor”). Total relative abundance across taxa classified into each type are plotted as in Fig. 3 panels A and C, with a line in each panel representing an individual patient and colored by study arm. In all panels, symbols indicate individual time points with p-values less than 0.1 (•) or 0.05 (*) by MWU test for differences between patients who did or did not receive antibiotic pretreatment.

### Supplementary Results: Donor/Recipient Strain Coexistence

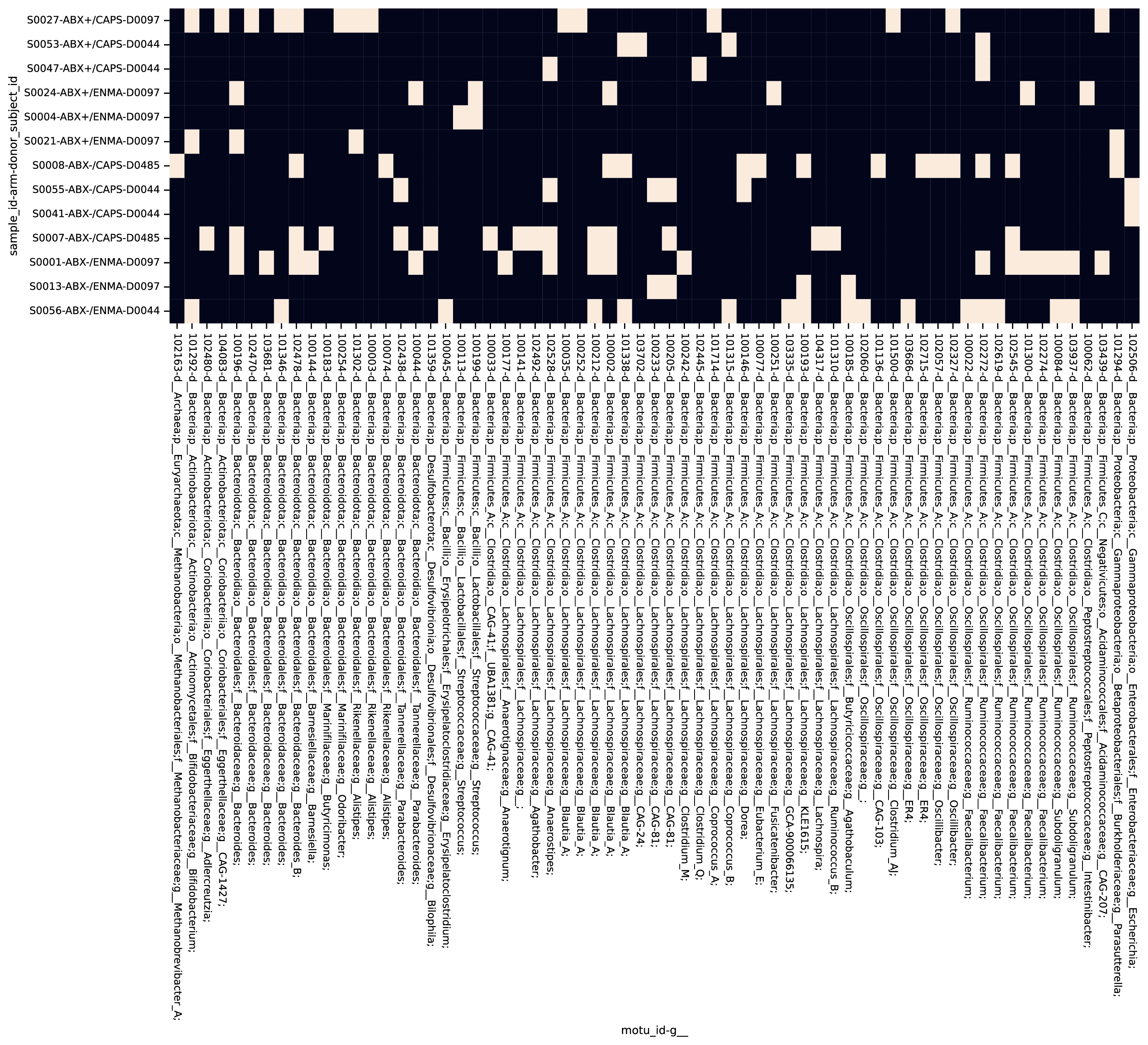

**Figure S5**: Identification of possible co-existence at follow-up of both donor and subject strains. Patient samples collected at F1 with both donor and patient strains of a single species detected are indicated (white tiles). Rows reflect individual patients and labels include subject ID, study arm, and donor. Column labels indicate the species and include a GTDB taxonomy up to the genus level. Species without examples of co-existence are excluded.

### Supplementary Methods: Detailed Patient Inclusion/Exclusion Criteria

#### Inclusion Criteria

- Patients with history of mild to moderate Ulcerative Colitis confirmed by endoscopy and pathology.
- Total Mayo score 4-9, endoscopic subscore ≥1; patients who have not had endoscopic evaluation within one year of enrollment will have flexible sigmoidoscopy for evaluation.
- Age 18-64 and deemed otherwise healthy at the discretion of the investigator.
- Concurrent therapies with mesalamine (stable x 4 weeks), immunomodulators (stable x 3 months), and biologic agents (stable x 3 months) will be allowed to continue during study.
- Prednisone must be ≤ 10 mg/day at the time of treatment and will be weaned by 2.5 mg/week during the study period.

#### Exclusion Criteria

- Severe or refractory UC defined as Mayo score ≥10, endoscopic disease activity score 3
- Untreated enteric infection (positive stool test for any of the following: Clostridium difficile, Salmonella, Shigella, Yersinia, Campylobacter, enteropathogenic E. coli or other enteric infection at the discretion of the investigator.
- History of colectomy
- Disease limited to distal proctitis (distal 5 cm)
- Patients taking probiotics within six weeks of planned FMT therapy.
- Severe immunodeficiency, inherited or acquired (e.g. HIV, chemotherapy or radiation therapy)
- Patients with the following laboratory abnormalities: absolute neutrophil count (ANC) < 1000 /µl, platelets <50 x 10^9 /L, hemoglobin <6.5 g/dL.
- History of anaphylaxis (severe allergic reaction) to food allergens (e.g. tree nuts, shellfish)
- Dysphagia (orophyaryngeal, esophageal, functional, neuromuscular)
- History of recurrent aspiration episodes
- Documented severe gastroparesis
- Active intestinal obstruction
- Patients with renal insufficiency (GFR < 50 ml/min)
- Allergy to the following generally regarded as safe ingredients (GRAS): glycerol, acid resistant HPMC, gellan gum, cocoa butter, titanium dioxide
- Adverse event attributable to any previous FMT
- Allergy/intolerance to proton pump inhibitor therapy
- Allergy/intolerance to vancomycin, metronidazole, or neomycin.
- Non-steroidal inflammatory medications (NSAIDs) as long-term treatment, defined as use for at least 4 days a week each month.
- Cholestyramine use
- Any condition in which the investigator thinks the FMT treatment may pose a health risk (e.g. severely immunocompromised)
- Simultaneous participation in another interventional clinical trial
- Patients who are pregnant, breast feeding or planning pregnancy during study trial period.
- During the trial period until one week after the trial end: Non-use of appropriate contraceptives in females of childbearing potential (e.g. condoms, intrauterine device (IUD), hormonal contraception, or other means considered adequate by the responsible investigator) or in males with a child-fathering potential (condoms, or other means considered adequate by the responsible investigator during treatment) or well-founded doubt about the patient’s cooperation
- Patients with any other significant medical condition that could confound or interfere with evaluation of safety, tolerability or prevent compliance with the study protocol at the discretion of the investigator
- Life expectancy <6 months

### Supplementary Methods: Stool Sampling Instructions to Patients

Instructions to patients for stool self-sampling.

SelfSampling.pdf

### Supplementary Methods: Clinical Laboratory Assessments

**Table S8**: List of clinical laboratory assessments

SupplementaryTablesS1-8.xlsx

### Supplementary Methods: Sample Processing

#### Sample collection and DNA extraction

Patients were instructed to collect a stool sample into provided sample vials (Sarstedt faeces tubes cat. no. 80.734.311) before each study visit and either bring it refrigerated to our clinic in-person or send it with a frozen cool pack via overnight courier. Upon delivery, stool specimens were stored at -80 °C until their analysis.

DNA extraction from fecal samples was performed using the modified cetyltrimethylammonium bromide (CTAB) as previously described [69]. In brief, ~0.3 g aliquot was taken from each frozen stool sample and suspended in 500 µL CTAB extraction buffer in a Lysing Matrix E tube (MP Biomedicals) by vortexing, followed by incubation for 15 minutes at 65 °C. After adding 500 µL phenol:chloroform:isoamyl alcohol (25:24:1), the solution underwent bead-beating (5.5 m/s for 30 seconds), followed by centrifugation (16,000 g for 5 minutes at 4 °C). The resulting aqueous phase (approximately 400 µL) was transferred to a new 2 mL 96-well plate. An additional 500 µL CTAB extraction buffer was added to the fecal aliquot and both the bead-beating and centrifugation steps were repeated, resulting in approximately 800 µL. An equal volume of chloroform was added and the solution was mixed and centrifuged (3,000 g for 10 minutes). The aqueous phase (approximately 600 µl) was transferred to another 2 mL 96-well plate, combined with 2-volume polyethylene glycol and stored at 4 °C overnight to precipitate DNA. Samples were then centrifuged (3,000 g for 60 minutes) to pellet DNA, washed twice with cold 70% EtOH, resuspended in sterile water and diluted to 10 ng DNA/µL (Qubit dsDNA BR Assay Kit; ThermoFisher Scientific, MA)

#### Sequencing

For 16S rRNA gene profiling, The V4 region was amplified as previously described [69]. PCR reactions were performed with 0.625 U Hot Start ExTaq and 1x buffer, 200 µM dNTPs, 0.56 µL/µL BSA, 0.4 µM each forward (F515) and reverse (R806) primers in triplicate 25 µL reactions containing 10ng of template gDNA. Thermal cycling was set at: 98 °C for 2 minutes, 30 rounds of 98 °C for 20 sec, 50 °C for 30 sec, 72 °C for 45 sec, and a final extension at 72 °C for 10 minutes. Amplicons were normalized (SequalPrep Normalization Plate Kit; ThermoFisher Scientific, MA), quantified (Qubit dsDNA BR Assay Kit; ThermoFisher Scientific, MA), pooled in equimolar concentrations, purified (Agencourt AMPure XP System; Beckman-Coulter), quantified (KAPA Library Quantification Kit; KAPA Biosystems), and diluted to 2 nM. Equimolar PhiX spike-in control was added at 40% final volume, and the samples were sequenced on an Illumina NextSeq 500 Platform.

For metagenomic sequencing, an aliquot of the extracted DNA was sent to QB3 at the University of California, Berkeley <<https://qb3.berkeley.edu/>> for sequencing on the NovaSeq 6000 platform using the 150PE Flow Cell S4 format.

#### Metabolomics

For metabolomics profiling, 112 fecal samples (200 mg) were provided to Metabolon (Durham, NC) who performed Ultrahigh Performance Liquid Chromotography/Tandem Mass Spectrometry (UPLC-MS/MS) and Gas Chromatography-Mass Spectrometry (GC-MS) based on standardized published protocol <<http://www.metabolon.com/>>. Detected molecules were identified through Metabolon’s library of purified standards, which encompasses >3,300 commercially available compounds. This yielded 1050 distinct metabolites among the samples, from which the 51 metabolites identified as primary and secondary bile acids were analyzed for this study. For each identified BA, peak intensities were normalized by the root mean squared intensity of that peak across samples before downstream analysis.

### Supplementary Methods: Microbiome profiling

#### 16S rRNA gene amplicon analysis

Raw amplicon sequencing data was processed as in [69]. Briefly, sequencer output was converted to fastq with bcl2fastq v2.16.0.10 <<https://support.illumina.com/sequencing/sequencing_software/bcl2fastq-conversion-software.html>> and demultiplexed by barcode with QIIME [70]. Quality-filtering via DADA2 [71] was performed using the recommended settings with the following adjustments: maximum expected errors allowed ≤ 3, truncation length of 150 bases for R1 and 140 for R2; chimeric sequences were found using minFoldParentOverAbundance = 8. Taxonomy was assigned with SILVA database V132 [72,73] to amplicon sequence variants (ASV). Contaminant ASV were identified with the decontam [74] package for R. ASV with >2% of total read sums contributed by controls and those with < 0.001% total read count across all samples were excluded. A phylogenetic tree was constructed using the phangorn [75,76], msa [77], and DECIPHER [78] packages for R. Amplicon sequencing resulted in an average of 3.4e5 paired reads per sample after processing.

#### Metagenomic reads pre-processing

Metagenomic sequences were deduplicated using FastUniq [79], any contaminating adapter sequence removed using Scythe [80], and then quality trimmed with Sickle [81]. Any cleaned reads that mapped to the human reference genome (GRCh38 [82]) using Bowtie2 [83] were removed. Shotgun metagenomic sequencing resulted in 4.9e7 paired reads per sample after processing.

#### Functional gene profiling

Reads were annotated with presumed functions by first identifying homology to the UHGP-50 (50% identity clusters, [64]) reference database using DIAMOND [84], a fast implementation of the BLASTX algorithm [85]. All top hits to this database were tallied for each sample. UHGP-50 hits were subsequently annotated with KEGG Orthology (KO) numbers based on assignments previously generated for the UHGP using EggNOG-mapper [86]. Tallies of reads mapping to these annotations were not corrected for gene length.
