## Supplementary material for "Strain-resolved analysis in a randomized trial of antibiotic pretreatment and maintenance dose delivery mode with fecal microbiota transplant for ulcerative colitis": Code Notebook

consolidated\_manuscript\_analyses


### Preamble¶

In [ ]:

```
%load_ext autoreload
%autoreload 2
```

In [ ]:

```
%load_ext rpy2.ipython
```

In [ ]:

```
import pandas as pd
import sqlite3
import seaborn as sns
import matplotlib as mpl
import matplotlib.pyplot as plt
from lib.pandas import idxwhere
import numpy as np
from tqdm import tqdm
import lib.project_data
import lib.plot
import lib.engraftment
import lib.project_style
from lib.dissimilarity import (
    dmatrix,
    align_dmatrices,
    mask_dmatrix,
    triu_stack,
    diss_table,
    partial_mantel_test,
    mantel_test,
)
import scipy as sp
import scipy.stats
import statsmodels.formula.api as smf
from statsmodels import api as sm
from warnings import warn
from itertools import product

np.random.seed(1)


def _print_visual_separator(n=100):
    print("\n" + "-" * n + "\n")


sns.set_context("paper")
# mpl.rcParams["figure.dpi"] = 200  # For publication images
mpl.rcParams["figure.dpi"] = 100  # For notebook use images
```

### Load Data¶

In [ ]:

```
con = sqlite3.connect("sdata/database.db")
```

#### Subject / Visit Data¶

In [ ]:

```
subject_x_visit_type = lib.project_data.load_subject_x_visit_type_table(con)
print(
    subject_x_visit_type.days_post_fmt.unstack().loc[
        :, lib.project_data.VISIT_TYPE_ORDER
    ]
)
```

```
visit_type  colonoscopy_1  maintenance_1  maintenance_2  maintenance_3  \
subject_id                                                               
S0001                 0.0            7.0           14.0           22.0   
S0004                 0.0            6.0           13.0           20.0   
S0007                 0.0            6.0           14.0           21.0   
S0008                 0.0            7.0           13.0           21.0   
S0013                 0.0            7.0           16.0           23.0   
S0017                 0.0            7.0           14.0           21.0   
S0021                 0.0            6.0           14.0           20.0   
S0024                 0.0            7.0           12.0           21.0   
S0027                 0.0            5.0           10.0           17.0   
S0029                 0.0            8.0           13.0           21.0   
S0031                 0.0            7.0           13.0           21.0   
S0037                 0.0            7.0           14.0           21.0   
S0041                 0.0            7.0           14.0           21.0   
S0043                 0.0            6.0           15.0           20.0   
S0047                 0.0            6.0           13.0           20.0   
S0053                 0.0            4.0           11.0           18.0   
S0055                 0.0            7.0           14.0           21.0   
S0056                 0.0            5.0           12.0           19.0   
S0058                 0.0            NaN            NaN            NaN   
S0059                 0.0            6.0           12.0           21.0   
S0060                -1.0            4.0           11.0           18.0   
S0061                 0.0            4.0           11.0           19.0   
S0062                 0.0            7.0           14.0           21.0   
S0063                 0.0            4.0           13.0           20.0   
S0064                 0.0            4.0           16.0           23.0   

visit_type  maintenance_4  maintenance_5  maintenance_6  colonoscopy_2  \
subject_id                                                               
S0001                28.0           34.0           42.0           56.0   
S0004                27.0           33.0           41.0           56.0   
S0007                28.0           35.0           42.0           56.0   
S0008                28.0           35.0           42.0           53.0   
S0013                29.0           36.0           41.0           56.0   
S0017                28.0           35.0            NaN           42.0   
S0021                27.0           36.0            NaN           49.0   
S0024                28.0           35.0           42.0           56.0   
S0027                24.0           31.0           38.0           56.0   
S0029                28.0           35.0            NaN            NaN   
S0031                 NaN            NaN            NaN           54.0   
S0037                35.0           61.0            NaN            NaN   
S0041                28.0           35.0           42.0           47.0   
S0043                27.0           35.0           42.0            NaN   
S0047                27.0           33.0           41.0           84.0   
S0053                25.0           32.0           39.0           61.0   
S0055                29.0           36.0           42.0           56.0   
S0056                28.0           33.0           39.0           56.0   
S0058                 NaN            NaN            NaN            NaN   
S0059                27.0           34.0           40.0           54.0   
S0060                22.0           29.0           36.0           48.0   
S0061                25.0           32.0           39.0           56.0   
S0062                28.0           35.0           40.0           49.0   
S0063                27.0           34.0           46.0           56.0   
S0064                28.0           37.0            NaN           51.0   

visit_type  followup_1_month  followup_3_month  
subject_id                                      
S0001                   90.0             154.0  
S0004                   95.0             152.0  
S0007                   93.0             160.0  
S0008                  112.0             197.0  
S0013                  103.0             184.0  
S0017                   71.0               NaN  
S0021                  102.0               NaN  
S0024                   87.0             153.0  
S0027                   95.0             216.0  
S0029                   97.0             127.0  
S0031                    NaN             126.0  
S0037                   91.0               NaN  
S0041                   89.0             115.0  
S0043                    NaN               NaN  
S0047                  118.0               NaN  
S0053                  105.0             153.0  
S0055                   91.0             153.0  
S0056                   94.0               NaN  
S0058                    NaN               NaN  
S0059                    NaN               NaN  
S0060                  104.0               NaN  
S0061                   88.0               NaN  
S0062                   87.0               NaN  
S0063                    NaN               NaN  
S0064                    NaN               NaN
```

In [ ]:

```
subject = lib.project_data.load_subject_table(con)
recipient_x_sample_type = lib.project_data.load_recipient_x_sample_type_table(con)
recipient = subject.assign(
    remission_=lambda x: x.remission.where(~x.withdrawal_due_to_failure, np.nan),
    responder_=lambda x: x.responder.where(~x.withdrawal_due_to_failure, np.nan),
    mayo_endo_improved_=lambda x: x.mayo_endo_improved.where(
        ~x.withdrawal_due_to_failure, np.nan
    ),
)[lambda x: x.recipient]

recipient.sort_values(["arm"])[lib.project_data.RECIPIENT_SUMMARIZE_COLUMNS]
```

Out[ ]:

|  | arm | sex | donor\_subject\_id | withdrawal\_due\_to\_failure | mayo\_total\_start | mayo\_total\_end | remission | responder |
| --- | --- | --- | --- | --- | --- | --- | --- | --- |
| subject\_id |  |  |  |  |  |  |  |  |
| S0047 | ABX+/CAPS | male | D0044 | False | 4.0 | 1.0 | True | True |
| S0068 | ABX+/CAPS | female | D0485 | <NA> | NaN | NaN | <NA> | <NA> |
| S0066 | ABX+/CAPS | male | D0485 | <NA> | NaN | NaN | <NA> | <NA> |
| S0063 | ABX+/CAPS | female | D0065 | False | 5.0 | 2.0 | True | True |
| S0053 | ABX+/CAPS | male | D0044 | False | 6.0 | 1.0 | True | True |
| S0027 | ABX+/CAPS | female | D0097 | False | 5.0 | 0.0 | True | True |
| S0037 | ABX+/CAPS | male | D0485 | True | 8.0 | NaN | False | False |
| S0064 | ABX+/ENMA | male | D0485 | False | 6.0 | 2.0 | True | True |
| S0060 | ABX+/ENMA | male | D0044 | False | 6.0 | 8.0 | False | False |
| S0043 | ABX+/ENMA | female | D0097 | True | 10.0 | NaN | False | False |
| S0070 | ABX+/ENMA | female | D0097 | <NA> | NaN | NaN | <NA> | <NA> |
| S0024 | ABX+/ENMA | male | D0097 | False | 6.0 | 4.0 | False | False |
| S0004 | ABX+/ENMA | male | D0097 | False | 4.0 | 0.0 | True | True |
| S0021 | ABX+/ENMA | female | D0097 | False | 4.0 | 8.0 | False | False |
| S0041 | ABX-/CAPS | female | D0044 | False | 7.0 | 6.0 | False | False |
| S0055 | ABX-/CAPS | female | D0044 | False | 9.0 | 8.0 | False | False |
| S0067 | ABX-/CAPS | male | D0485 | <NA> | NaN | NaN | <NA> | <NA> |
| S0007 | ABX-/CAPS | male | D0485 | False | 6.0 | 7.0 | False | False |
| S0059 | ABX-/CAPS | male | D0044 | False | 7.0 | 3.0 | False | True |
| S0031 | ABX-/CAPS | female | D0485 | <NA> | 6.0 | NaN | <NA> | <NA> |
| S0008 | ABX-/CAPS | male | D0485 | False | 5.0 | 1.0 | True | True |
| S0029 | ABX-/ENMA | female | D0097 | <NA> | 5.0 | NaN | <NA> | <NA> |
| S0013 | ABX-/ENMA | male | D0097 | False | 4.0 | 3.0 | False | False |
| S0017 | ABX-/ENMA | female | D0097 | True | 5.0 | 9.0 | False | False |
| S0061 | ABX-/ENMA | male | D0044 | False | 6.0 | 4.0 | False | False |
| S0058 | ABX-/ENMA | female | D0044 | <NA> | 9.0 | NaN | <NA> | <NA> |
| S0056 | ABX-/ENMA | female | D0044 | False | 7.0 | 8.0 | False | False |
| S0069 | ABX-/ENMA | female | D0097 | <NA> | NaN | NaN | <NA> | <NA> |
| S0062 | ABX-/ENMA | male | D0485 | False | 8.0 | 1.0 | True | True |
| S0001 | ABX-/ENMA | female | D0097 | False | 8.0 | 4.0 | False | True |

In [ ]:

```
recipient.shape
```

Out[ ]:

```
(30, 21)
```

#### Microbiome Data¶

In [ ]:

```
donor_means_list = ["D0044_mean", "D0097_mean", "D0485_mean"]
```

In [ ]:

```
sample_x_rotu_cvrg = lib.project_data.load_sample_x_rotu_cvrg_table(con)
sample_x_rotu = lib.project_data.load_sample_x_rotu_rabund_table(con)

low_mgen_coverage_samples = ["SS01011"]  # See data_checkup.ipynb

sample_x_motu = lib.project_data.load_sample_x_motu_rabund_table(con).drop(
    low_mgen_coverage_samples, errors="ignore"
)
sample_x_sotu = lib.project_data.load_sample_x_sotu_rabund_table(con).drop(
    low_mgen_coverage_samples, errors="ignore"
)
```

In [ ]:

```
sample_x_ko_cvrg = lib.project_data.load_sample_x_ko_table(con).drop(
    low_mgen_coverage_samples, errors="ignore"
)
```

In [ ]:

```
sample_x_chem_ba = lib.project_data.load_sample_x_chem_ba_table(con)
sample_x_chem_ba_std2 = sample_x_chem_ba.apply(lambda x: x / np.sqrt(np.mean(x ** 2)))
```

In [ ]:

```
sample = lib.project_data.load_sample_table(con)
recipient_has_mgen_data_list = sample.loc[
    list(set(recipient_x_sample_type["sample_id"]) & set(sample_x_ko_cvrg.index))
].subject_id.unique()
```

In [ ]:

```
sample_x_motu[100022].sort_values(ascending=False).head(20)
```

Out[ ]:

```
sample_id
DS0044_006    0.152174
DS0044_009    0.103879
SS01148       0.088235
SS01145       0.080646
SS01105       0.077779
D0044_mean    0.068596
SS01033       0.063600
SS01139       0.060779
SS01134       0.060040
DS0097_006    0.057485
SS01142       0.055676
DS0097_022    0.053872
SS01027       0.052720
DS0097_034    0.051043
SS01151       0.049468
DS0097_025    0.046021
DS0097_009    0.045934
DS0097_024    0.045610
SS01169       0.044627
DS0097_031    0.044512
Name: 100022, dtype: float64
```

In [ ]:

```
sample_x_sotu.loc[["SS01009", "SS01057"], "100022":"100023"].sum(1)
```

Out[ ]:

```
sample_id
SS01009    0.000196
SS01057    0.002239
dtype: float64
```

\_sample = pd.read\_table('meta/sample.tsv', index\_col='sample\_id')
sample.join(\_sample[['plate\_well', 'source\_sample\_id']])[sample.has\_mgen][['source\_sample\_id', 'sample\_notes', 'plate\_well']].to\_csv('all\_sequenced\_samples.csv')

In [ ]:

```
donor_order = ["D0044", "D0097", "D0485"]

subject_has_mgen_list = []

for donor_subject_id in donor_order:
    subject_has_mgen_list.append(donor_subject_id)
    _donor_subject_list = idxwhere(
        (recipient.donor_subject_id == donor_subject_id)
        & recipient.index.to_series().isin(recipient_has_mgen_data_list)
    )
    subject_has_mgen_list.extend(_donor_subject_list)

subject_has_mgen_order = (
    subject.loc[subject_has_mgen_list]
    .sort_values(["donor_subject_id", "recipient", "subject_id"], ascending=True)
    .index.to_list()
)

recipient_has_mgen_order = [
    subject_id for subject_id in subject_has_mgen_order if subject_id not in donor_order
]


# recipient_has_mgen_order = idxwhere((
#     recipient_x_sample_type
#     .sample_id
#     .unstack('sample_type')
#     .reindex(columns=lib.project_data.SAMPLE_TYPE_ORDER)
# ).isin(sample_x_sotu.index).sum(1) > 2)


subject_has_mgen_order, recipient_has_mgen_order
```

Out[ ]:

```
(['D0044',
  'S0041',
  'S0047',
  'S0053',
  'S0055',
  'S0056',
  'D0097',
  'S0001',
  'S0004',
  'S0013',
  'S0021',
  'S0024',
  'S0027',
  'D0485',
  'S0007',
  'S0008'],
 ['S0041',
  'S0047',
  'S0053',
  'S0055',
  'S0056',
  'S0001',
  'S0004',
  'S0013',
  'S0021',
  'S0024',
  'S0027',
  'S0007',
  'S0008'])
```

In [ ]:

```
subject_color_palette = lib.plot.construct_ordered_pallete(
    subject_has_mgen_order,
    cm="rainbow",
)

lib.plot.demo_pallete(subject_color_palette)
```

```
   D0044     S0041     S0047     S0053     S0055     S0056     D0097  \
0    0.5  0.374510  0.249020  0.123529  0.001961  0.127451  0.252941   
1    0.0  0.195845  0.384106  0.557489  0.709281  0.833602  0.925638   
2    1.0  0.995147  0.980635  0.956604  0.923289  0.881012  0.830184   
3    1.0  1.000000  1.000000  1.000000  1.000000  1.000000  1.000000   

      S0001     S0004     S0013     S0021     S0024     S0027     D0485  \
0  0.378431  0.503922  0.629412  0.754902  0.880392  1.000000  1.000000   
1  0.981823  0.999981  0.979410  0.920906  0.826734  0.700543  0.547220   
2  0.771298  0.704926  0.631711  0.552365  0.467658  0.378411  0.285492   
3  1.000000  1.000000  1.000000  1.000000  1.000000  1.000000  1.000000   

      S0007     S0008  
0  1.000000  1.000000  
1  0.372702  0.183750  
2  0.189801  0.092268  
3  1.000000  1.000000
```

##### Taxonomy¶

In [ ]:

```
motu_to_taxonomy = lib.project_data.load_motu_to_taxonomy_table(con)
```

In [ ]:

```
sotu_to_motu = sample_x_sotu.columns.to_series(name="motu_id").str[:6].astype(int)
sotu_to_taxonomy = (
    sotu_to_motu.astype(str)
    .to_frame()
    .join(motu_to_taxonomy.set_index(motu_to_taxonomy.index.astype(str)), on="motu_id")
    .assign(motu_id=lambda x: x.motu_id.astype(int))
)
sotu_to_taxonomy.head(2)
```

Out[ ]:

|  | motu\_id | d\_\_ | p\_\_ | c\_\_ | o\_\_ | f\_\_ | g\_\_ | s\_\_ |
| --- | --- | --- | --- | --- | --- | --- | --- | --- |
| sotu\_id |  |  |  |  |  |  |  |  |
| 100002-other | 100002 | d\_\_Bacteria; | d\_\_Bacteria;p\_\_Firmicutes\_A; | d\_\_Bacteria;p\_\_Firmicutes\_A;c\_\_Clostridia; | d\_\_Bacteria;p\_\_Firmicutes\_A;c\_\_Clostridia;o\_\_L... | d\_\_Bacteria;p\_\_Firmicutes\_A;c\_\_Clostridia;o\_\_L... | d\_\_Bacteria;p\_\_Firmicutes\_A;c\_\_Clostridia;o\_\_L... | d\_\_Bacteria;p\_\_Firmicutes\_A;c\_\_Clostridia;o\_\_L... |
| 100002-s001 | 100002 | d\_\_Bacteria; | d\_\_Bacteria;p\_\_Firmicutes\_A; | d\_\_Bacteria;p\_\_Firmicutes\_A;c\_\_Clostridia; | d\_\_Bacteria;p\_\_Firmicutes\_A;c\_\_Clostridia;o\_\_L... | d\_\_Bacteria;p\_\_Firmicutes\_A;c\_\_Clostridia;o\_\_L... | d\_\_Bacteria;p\_\_Firmicutes\_A;c\_\_Clostridia;o\_\_L... | d\_\_Bacteria;p\_\_Firmicutes\_A;c\_\_Clostridia;o\_\_L... |

###### Slice/Merge Taxa¶

In [ ]:

```
sample_x_sotu_bacteroidota = sample_x_sotu.loc[
    :,
    sample_x_sotu.columns.isin(
        idxwhere(sotu_to_taxonomy.p__.isin(["d__Bacteria;p__Bacteroidota;"]))
    ),
].apply(lib.stats.normalize, axis=1)
sample_x_sotu_firmicutes_A = sample_x_sotu.loc[
    :,
    sample_x_sotu.columns.isin(
        idxwhere(sotu_to_taxonomy.p__.isin(["d__Bacteria;p__Firmicutes_A;"]))
    ),
].apply(lib.stats.normalize, axis=1)
sample_x_sotu_firmicutes_C = sample_x_sotu.loc[
    :,
    sample_x_sotu.columns.isin(
        idxwhere(sotu_to_taxonomy.p__.isin(["d__Bacteria;p__Firmicutes_C;"]))
    ),
].apply(lib.stats.normalize, axis=1)
sample_x_sotu_firmicutes = sample_x_sotu.loc[
    :,
    sample_x_sotu.columns.isin(
        idxwhere(sotu_to_taxonomy.p__.isin(["d__Bacteria;p__Firmicutes;"]))
    ),
].apply(lib.stats.normalize, axis=1)
sample_x_sotu_actinobacteria = sample_x_sotu.loc[
    :,
    sample_x_sotu.columns.isin(
        idxwhere(sotu_to_taxonomy.p__.isin(["d__Bacteria;p__Actinobacteriota;"]))
    ),
].apply(lib.stats.normalize, axis=1)
sample_x_sotu_proteobacteria = sample_x_sotu.loc[
    :,
    sample_x_sotu.columns.isin(
        idxwhere(sotu_to_taxonomy.p__.isin(["d__Bacteria;p__Proteobacteria;"]))
    ),
].apply(lib.stats.normalize, axis=1)
```

sample\_x\_sotu.loc[sample.sample\_type.isin(['donor\_mean', 'baseline'])].groupby(sotu\_to\_taxonomy.p\_\_, axis='columns').sum().mean().sort\_values()

In [ ]:

```
sample_x_family = sample_x_sotu.groupby(sotu_to_taxonomy.f__, axis="columns").sum()
sample_x_family_bacteroidota = sample_x_sotu_bacteroidota.groupby(
    sotu_to_taxonomy.f__, axis="columns"
).sum()
sample_x_family_firmicutes = sample_x_sotu_firmicutes.groupby(
    sotu_to_taxonomy.f__, axis="columns"
).sum()
sample_x_family_firmicutes_A = sample_x_sotu_firmicutes_A.groupby(
    sotu_to_taxonomy.f__, axis="columns"
).sum()
sample_x_family_firmicutes_C = sample_x_sotu_firmicutes_C.groupby(
    sotu_to_taxonomy.f__, axis="columns"
).sum()
sample_x_family_actinobacteria = sample_x_sotu_actinobacteria.groupby(
    sotu_to_taxonomy.f__, axis="columns"
).sum()
sample_x_family_proteobacteria = sample_x_sotu_proteobacteria.groupby(
    sotu_to_taxonomy.f__, axis="columns"
).sum()
```

In [ ]:

```
sample_x_motu = sample_x_sotu.groupby(sotu_to_taxonomy.motu_id, axis="columns").sum()
sample_x_motu_bacteroidota = sample_x_sotu_bacteroidota.groupby(
    sotu_to_taxonomy.motu_id, axis="columns"
).sum()
sample_x_motu_firmicutes = sample_x_sotu_firmicutes.groupby(
    sotu_to_taxonomy.motu_id, axis="columns"
).sum()
sample_x_motu_firmicutes_A = sample_x_sotu_firmicutes_A.groupby(
    sotu_to_taxonomy.motu_id, axis="columns"
).sum()
sample_x_motu_firmicutes_C = sample_x_sotu_firmicutes_C.groupby(
    sotu_to_taxonomy.motu_id, axis="columns"
).sum()
sample_x_motu_actinobacteria = sample_x_sotu_actinobacteria.groupby(
    sotu_to_taxonomy.motu_id, axis="columns"
).sum()
sample_x_motu_proteobacteria = sample_x_sotu_proteobacteria.groupby(
    sotu_to_taxonomy.motu_id, axis="columns"
).sum()
```

In [ ]:

```
sample_x_family = sample_x_motu.groupby(motu_to_taxonomy.f__, axis="columns").sum()
sample_x_phylum = sample_x_motu.groupby(motu_to_taxonomy.p__, axis="columns").sum()
```

#### Sample Metadata¶

In [ ]:

```
sample = (
    lib.project_data.load_sample_table(con)
    .assign(
        has_mgen=lambda x: x.index.isin(idxwhere(sample_x_ko_cvrg.sum(1) > 1e6)),
        has_chem=lambda x: x.index.isin(sample_x_chem_ba.index),
        has_rrs=lambda x: x.index.isin(sample_x_rotu.index),
    )
    .sort_values(["subject_id", "days_post_fmt"])
)
```

In [ ]:

```
recipient_x_sample_type = lib.project_data.load_recipient_x_sample_type_table(con)

print(recipient_x_sample_type["sample_id"].unstack())
```

```
sample_type baseline followup_1 followup_2 followup_3 post_antibiotic  \
subject_id                                                              
S0001        SS01002    SS01026    SS01048    SS01070             NaN   
S0004        SS01000    SS01018    SS01042    SS01067         SS01001   
S0007        SS01020    SS01046    SS01062    SS01093             NaN   
S0008        SS01038    SS01063    SS01087    SS01119             NaN   
S0013        SS01013    SS01037    SS01053    SS01089             NaN   
S0017        SS01021        NaN    SS00999        NaN             NaN   
S0021        SS01068    SS01086    SS01105        NaN         SS01069   
S0024        SS01057    SS01088    SS01098    SS01124         SS01056   
S0027        SS01090    SS01108    SS01125    SS01132         SS01091   
S0029        SS01058        NaN    SS01106        NaN             NaN   
S0031        SS01041    SS01061        NaN        NaN             NaN   
S0037        SS01095        NaN        NaN        NaN         SS01096   
S0041        SS01117    SS01131    SS01133        NaN             NaN   
S0043        SS01099        NaN        NaN        NaN         SS01101   
S0047        SS01135    SS01153    SS01165        NaN         SS01136   
S0053        SS01134    SS01148    SS01162    SS01175         SS01137   
S0055        SS01150    SS01161    SS01166    SS01181             NaN   
S0056        SS01164    SS01172    SS01185        NaN             NaN   
S0058        SS01178        NaN        NaN        NaN             NaN   
S0059        SS01173    SS01199        NaN        NaN             NaN   
S0060        SS01180    SS01207        NaN        NaN         SS01184   
S0061        SS01179        NaN        NaN        NaN             NaN   
S0062        SS01198        NaN        NaN        NaN             NaN   

sample_type pre_maintenance_1 pre_maintenance_2 pre_maintenance_3  \
subject_id                                                          
S0001                 SS01007           SS01008           SS01012   
S0004                 SS01004           SS01005           SS01006   
S0007                     NaN               NaN           SS01032   
S0008                     NaN           SS01047           SS01054   
S0013                 SS01017           SS01023           SS01027   
S0017                 SS01025           SS01029           SS01031   
S0021                 SS01072           SS01076           SS01080   
S0024                 SS01060           SS01066           SS01073   
S0027                     NaN           SS01092           SS01094   
S0029                 SS01064           SS01065               NaN   
S0031                 SS01045           SS01049           SS01050   
S0037                 SS01104           SS01110           SS01111   
S0041                 SS01120           SS01123           SS01126   
S0043                 SS01107           SS01109           SS01115   
S0047                 SS01140               NaN           SS01149   
S0053                 SS01138           SS01139           SS01141   
S0055                 SS01154           SS01155           SS01156   
S0056                 SS01163           SS01169           SS01168   
S0058                     NaN           SS01187           SS01189   
S0059                 SS01176           SS01177           SS01182   
S0060                 SS01183           SS01188           SS01194   
S0061                     NaN           SS01191           SS01197   
S0062                 SS01201           SS01205           SS01210   

sample_type pre_maintenance_4 pre_maintenance_5 pre_maintenance_6  
subject_id                                                         
S0001                 SS01016           SS01019           SS01022  
S0004                 SS01009           SS01011           SS01015  
S0007                 SS01034           SS01039           SS01043  
S0008                 SS01052           SS01055           SS01059  
S0013                 SS01030           SS01033           SS01036  
S0017                     NaN           SS01035               NaN  
S0021                 SS01083           SS01084               NaN  
S0024                 SS01075           SS01078           SS01081  
S0027                 SS01097           SS01100           SS01103  
S0029                 SS01071               NaN               NaN  
S0031                     NaN               NaN               NaN  
S0037                 SS01114           SS01122               NaN  
S0041                 SS01127           SS01128           SS01130  
S0043                 SS01113           SS01116           SS01118  
S0047                 SS01146           SS01147               NaN  
S0053                 SS01142           SS01145           SS01151  
S0055                 SS01160           SS01157           SS01158  
S0056                 SS01167           SS01170           SS01171  
S0058                 SS01196               NaN               NaN  
S0059                     NaN           SS01200               NaN  
S0060                 SS01195           SS01204          SS01109a  
S0061                 SS01203           SS01208           SS01206  
S0062                 SS01211               NaN               NaN
```

### Results¶

#### Subsection: Study design and subject demographics¶

In [ ]:

```
# TODO: Build demographics table?
```

In [ ]:

```
recipient_has_efficacy_list = idxwhere(recipient.remission.notna())

(
    subject_x_visit_type.loc[recipient_has_efficacy_list]
    .groupby(level="visit_type")
    .days_post_fmt.quantile([0.0, 0.25, 0.5, 0.75, 1.0])
    .unstack()
)
```

Out[ ]:

|  | 0.00 | 0.25 | 0.50 | 0.75 | 1.00 |
| --- | --- | --- | --- | --- | --- |
| visit\_type |  |  |  |  |  |
| colonoscopy\_1 | -1.0 | 0.00 | 0.0 | 0.00 | 0.0 |
| colonoscopy\_2 | 42.0 | 50.50 | 56.0 | 56.00 | 84.0 |
| followup\_1\_month | 71.0 | 89.25 | 93.5 | 102.75 | 118.0 |
| followup\_3\_month | 115.0 | 153.00 | 153.5 | 178.00 | 216.0 |
| maintenance\_1 | 4.0 | 5.00 | 6.0 | 7.00 | 7.0 |
| maintenance\_2 | 10.0 | 12.00 | 13.5 | 14.00 | 16.0 |
| maintenance\_3 | 17.0 | 20.00 | 21.0 | 21.00 | 23.0 |
| maintenance\_4 | 22.0 | 27.00 | 28.0 | 28.00 | 35.0 |
| maintenance\_5 | 29.0 | 33.00 | 35.0 | 35.00 | 61.0 |
| maintenance\_6 | 36.0 | 39.25 | 41.0 | 42.00 | 46.0 |

In [ ]:

```
(
    sample[lambda x: x.subject_id.isin(recipient_has_efficacy_list)]
    .groupby("sample_type")
    .days_post_fmt.quantile([0.0, 0.25, 0.5, 0.75, 1.0])
    .unstack()
)
```

Out[ ]:

|  | 0.00 | 0.25 | 0.50 | 0.75 | 1.00 |
| --- | --- | --- | --- | --- | --- |
| sample\_type |  |  |  |  |  |
| baseline | -15.0 | -7.25 | -5.0 | -1.75 | 0.0 |
| followup\_1 | 46.0 | 48.00 | 53.0 | 55.00 | 82.0 |
| followup\_2 | 71.0 | 90.50 | 95.0 | 102.00 | 118.0 |
| followup\_3 | 152.0 | 153.00 | 153.0 | 160.00 | 196.0 |
| post\_antibiotic | -2.0 | -2.00 | -2.0 | -1.00 | -1.0 |
| pre\_maintenance\_1 | 4.0 | 5.00 | 5.5 | 6.25 | 7.0 |
| pre\_maintenance\_2 | 10.0 | 11.00 | 12.5 | 14.00 | 16.0 |
| pre\_maintenance\_3 | 16.0 | 19.00 | 19.5 | 20.00 | 23.0 |
| pre\_maintenance\_4 | 22.0 | 26.00 | 27.0 | 28.00 | 34.0 |
| pre\_maintenance\_5 | 29.0 | 32.00 | 34.0 | 35.00 | 60.0 |
| pre\_maintenance\_6 | 36.0 | 39.25 | 40.5 | 42.00 | 42.0 |

In [ ]:

```
(
    sample[lambda x: x.subject_id.isin(recipient_has_efficacy_list)]
    .groupby("sample_type")
    .days_post_fmt.quantile([0.0, 0.25, 0.5, 0.75, 1.0])
    .unstack()
)
```

Out[ ]:

|  | 0.00 | 0.25 | 0.50 | 0.75 | 1.00 |
| --- | --- | --- | --- | --- | --- |
| sample\_type |  |  |  |  |  |
| baseline | -15.0 | -7.25 | -5.0 | -1.75 | 0.0 |
| followup\_1 | 46.0 | 48.00 | 53.0 | 55.00 | 82.0 |
| followup\_2 | 71.0 | 90.50 | 95.0 | 102.00 | 118.0 |
| followup\_3 | 152.0 | 153.00 | 153.0 | 160.00 | 196.0 |
| post\_antibiotic | -2.0 | -2.00 | -2.0 | -1.00 | -1.0 |
| pre\_maintenance\_1 | 4.0 | 5.00 | 5.5 | 6.25 | 7.0 |
| pre\_maintenance\_2 | 10.0 | 11.00 | 12.5 | 14.00 | 16.0 |
| pre\_maintenance\_3 | 16.0 | 19.00 | 19.5 | 20.00 | 23.0 |
| pre\_maintenance\_4 | 22.0 | 26.00 | 27.0 | 28.00 | 34.0 |
| pre\_maintenance\_5 | 29.0 | 32.00 | 34.0 | 35.00 | 60.0 |
| pre\_maintenance\_6 | 36.0 | 39.25 | 40.5 | 42.00 | 42.0 |

In [ ]:

```
d0 = subject_x_visit_type.days_post_fmt.unstack()[lib.project_data.VISIT_TYPE_ORDER]
d1 = pd.DataFrame(
    d0[lib.project_data.VISIT_TYPE_ORDER[1:]].values
    - d0[lib.project_data.VISIT_TYPE_ORDER[:-1]].values,
    index=d0.index,
    columns=lib.project_data.VISIT_TYPE_ORDER[1:],
)
print(d1)
```

```
            maintenance_1  maintenance_2  maintenance_3  maintenance_4  \
subject_id                                                               
S0001                 7.0            7.0            8.0            6.0   
S0004                 6.0            7.0            7.0            7.0   
S0007                 6.0            8.0            7.0            7.0   
S0008                 7.0            6.0            8.0            7.0   
S0013                 7.0            9.0            7.0            6.0   
S0017                 7.0            7.0            7.0            7.0   
S0021                 6.0            8.0            6.0            7.0   
S0024                 7.0            5.0            9.0            7.0   
S0027                 5.0            5.0            7.0            7.0   
S0029                 8.0            5.0            8.0            7.0   
S0031                 7.0            6.0            8.0            NaN   
S0037                 7.0            7.0            7.0           14.0   
S0041                 7.0            7.0            7.0            7.0   
S0043                 6.0            9.0            5.0            7.0   
S0047                 6.0            7.0            7.0            7.0   
S0053                 4.0            7.0            7.0            7.0   
S0055                 7.0            7.0            7.0            8.0   
S0056                 5.0            7.0            7.0            9.0   
S0058                 NaN            NaN            NaN            NaN   
S0059                 6.0            6.0            9.0            6.0   
S0060                 5.0            7.0            7.0            4.0   
S0061                 4.0            7.0            8.0            6.0   
S0062                 7.0            7.0            7.0            7.0   
S0063                 4.0            9.0            7.0            7.0   
S0064                 4.0           12.0            7.0            5.0   

            maintenance_5  maintenance_6  colonoscopy_2  followup_1_month  \
subject_id                                                                  
S0001                 6.0            8.0           14.0              34.0   
S0004                 6.0            8.0           15.0              39.0   
S0007                 7.0            7.0           14.0              37.0   
S0008                 7.0            7.0           11.0              59.0   
S0013                 7.0            5.0           15.0              47.0   
S0017                 7.0            NaN            NaN              29.0   
S0021                 9.0            NaN            NaN              53.0   
S0024                 7.0            7.0           14.0              31.0   
S0027                 7.0            7.0           18.0              39.0   
S0029                 7.0            NaN            NaN               NaN   
S0031                 NaN            NaN            NaN               NaN   
S0037                26.0            NaN            NaN               NaN   
S0041                 7.0            7.0            5.0              42.0   
S0043                 8.0            7.0            NaN               NaN   
S0047                 6.0            8.0           43.0              34.0   
S0053                 7.0            7.0           22.0              44.0   
S0055                 7.0            6.0           14.0              35.0   
S0056                 5.0            6.0           17.0              38.0   
S0058                 NaN            NaN            NaN               NaN   
S0059                 7.0            6.0           14.0               NaN   
S0060                 7.0            7.0           12.0              56.0   
S0061                 7.0            7.0           17.0              32.0   
S0062                 7.0            5.0            9.0              38.0   
S0063                 7.0           12.0           10.0               NaN   
S0064                 9.0            NaN            NaN               NaN   

            followup_3_month  
subject_id                    
S0001                   64.0  
S0004                   57.0  
S0007                   67.0  
S0008                   85.0  
S0013                   81.0  
S0017                    NaN  
S0021                    NaN  
S0024                   66.0  
S0027                  121.0  
S0029                   30.0  
S0031                    NaN  
S0037                    NaN  
S0041                   26.0  
S0043                    NaN  
S0047                    NaN  
S0053                   48.0  
S0055                   62.0  
S0056                    NaN  
S0058                    NaN  
S0059                    NaN  
S0060                    NaN  
S0061                    NaN  
S0062                    NaN  
S0063                    NaN  
S0064                    NaN
```

#### Subsection: FMT efficacy and impacts of treatment¶

In [ ]:

```
recipient[~recipient["withdrawal_due_to_failure"].isna()][
    lib.project_data.RECIPIENT_SUMMARIZE_COLUMNS
]
```

Out[ ]:

|  | arm | sex | donor\_subject\_id | withdrawal\_due\_to\_failure | mayo\_total\_start | mayo\_total\_end | remission | responder |
| --- | --- | --- | --- | --- | --- | --- | --- | --- |
| subject\_id |  |  |  |  |  |  |  |  |
| S0001 | ABX-/ENMA | female | D0097 | False | 8.0 | 4.0 | False | True |
| S0004 | ABX+/ENMA | male | D0097 | False | 4.0 | 0.0 | True | True |
| S0007 | ABX-/CAPS | male | D0485 | False | 6.0 | 7.0 | False | False |
| S0008 | ABX-/CAPS | male | D0485 | False | 5.0 | 1.0 | True | True |
| S0013 | ABX-/ENMA | male | D0097 | False | 4.0 | 3.0 | False | False |
| S0017 | ABX-/ENMA | female | D0097 | True | 5.0 | 9.0 | False | False |
| S0021 | ABX+/ENMA | female | D0097 | False | 4.0 | 8.0 | False | False |
| S0024 | ABX+/ENMA | male | D0097 | False | 6.0 | 4.0 | False | False |
| S0027 | ABX+/CAPS | female | D0097 | False | 5.0 | 0.0 | True | True |
| S0037 | ABX+/CAPS | male | D0485 | True | 8.0 | NaN | False | False |
| S0041 | ABX-/CAPS | female | D0044 | False | 7.0 | 6.0 | False | False |
| S0043 | ABX+/ENMA | female | D0097 | True | 10.0 | NaN | False | False |
| S0047 | ABX+/CAPS | male | D0044 | False | 4.0 | 1.0 | True | True |
| S0053 | ABX+/CAPS | male | D0044 | False | 6.0 | 1.0 | True | True |
| S0055 | ABX-/CAPS | female | D0044 | False | 9.0 | 8.0 | False | False |
| S0056 | ABX-/ENMA | female | D0044 | False | 7.0 | 8.0 | False | False |
| S0059 | ABX-/CAPS | male | D0044 | False | 7.0 | 3.0 | False | True |
| S0060 | ABX+/ENMA | male | D0044 | False | 6.0 | 8.0 | False | False |
| S0061 | ABX-/ENMA | male | D0044 | False | 6.0 | 4.0 | False | False |
| S0062 | ABX-/ENMA | male | D0485 | False | 8.0 | 1.0 | True | True |
| S0063 | ABX+/CAPS | female | D0065 | False | 5.0 | 2.0 | True | True |
| S0064 | ABX+/ENMA | male | D0485 | False | 6.0 | 2.0 | True | True |

In [ ]:

```
(
    recipient
    [
        lambda x: True
        & (~x.withdrawal_due_to_failure.isna())
        # & x.index.isin(sample[sample.has_chem].subject_id.unique())
    ]
    [lib.project_data.RECIPIENT_SUMMARIZE_COLUMNS]
    .groupby(['donor_subject_id', 'sex'])
    .apply(len)
    .unstack('sex', fill_value=0)
)

# D0485 got only male participants. So there's certainly some confounding of donor by sex.
# This is relevant particularly in the bile-acid analysis, because there are known sex differences in BA
# profiles.
```

Out[ ]:

| sex | female | male |
| --- | --- | --- |
| donor\_subject\_id |  |  |
| D0044 | 3 | 5 |
| D0065 | 1 | 0 |
| D0097 | 5 | 3 |
| D0485 | 0 | 5 |

In [ ]:

```
(
    recipient
    [
        lambda x: True
        & (~x.withdrawal_due_to_failure.isna())
        & x.index.isin(sample[sample.has_chem].subject_id.unique())
    ]
    [lib.project_data.RECIPIENT_SUMMARIZE_COLUMNS]
    .groupby(['donor_subject_id', 'sex'])
    .apply(len)
    .unstack('sex', fill_value=0)
)
```

Out[ ]:

| sex | female | male |
| --- | --- | --- |
| donor\_subject\_id |  |  |
| D0044 | 3 | 2 |
| D0097 | 3 | 3 |
| D0485 | 0 | 2 |

In [ ]:

```
recipient[recipient["withdrawal_due_to_failure"]][
    lib.project_data.RECIPIENT_SUMMARIZE_COLUMNS
]
```

Out[ ]:

|  | arm | sex | donor\_subject\_id | withdrawal\_due\_to\_failure | mayo\_total\_start | mayo\_total\_end | remission | responder |
| --- | --- | --- | --- | --- | --- | --- | --- | --- |
| subject\_id |  |  |  |  |  |  |  |  |
| S0017 | ABX-/ENMA | female | D0097 | True | 5.0 | 9.0 | False | False |
| S0037 | ABX+/CAPS | male | D0485 | True | 8.0 | NaN | False | False |
| S0043 | ABX+/ENMA | female | D0097 | True | 10.0 | NaN | False | False |

##### Remission and responder status¶

In [ ]:

```
# 'remission_' and 'responder_' are both strict definitions,
# excluding participants that did not complete the trial.
for endpoint in [
    "remission",
    "remission_",
    "responder",
    "responder_",
    "mayo_endo_improved",
]:

    print(recipient.groupby([endpoint]).apply(len).fillna(0).astype(int))
    _print_visual_separator()
```

```
remission
False    14
True      8
dtype: int64

----------------------------------------------------------------------------------------------------

remission_
False    11
True      8
dtype: int64

----------------------------------------------------------------------------------------------------

responder
False    12
True     10
dtype: int64

----------------------------------------------------------------------------------------------------

responder_
False     9
True     10
dtype: int64

----------------------------------------------------------------------------------------------------

mayo_endo_improved
False    11
True     11
dtype: int64

----------------------------------------------------------------------------------------------------
```

print(recipient[[
'treatment\_abx\_pre',
'maintenance\_',
'donor\_subject\_id',
'remission',
'responder',
'withdrawal\_due\_to\_failure',
]].to\_markdown())

###### Antibiotics¶

In [ ]:

```
# 'remission_' and 'responder_' are both strict definitions,
# excluding participants that did not complete the trial.
for endpoint in [
    "remission",
    "remission_",
    "responder",
    "responder_",
    "mayo_endo_improved",
]:
    contingency = (
        recipient.groupby(["antibiotics_", endpoint])
        .apply(len)
        .unstack()
        .fillna(0)
        .astype(int)
    )

    print(contingency)
    print(sp.stats.fisher_exact(contingency))
    _print_visual_separator()
```

```
remission     False  True
antibiotics_             
ABX+              5     6
ABX-              9     2
(0.18518518518518517, 0.1826625386996899)

----------------------------------------------------------------------------------------------------

remission_    False  True
antibiotics_             
ABX+              3     6
ABX-              8     2
(0.125, 0.0697785186949274)

----------------------------------------------------------------------------------------------------

responder     False  True
antibiotics_             
ABX+              5     6
ABX-              7     4
(0.47619047619047616, 0.6699214098594914)

----------------------------------------------------------------------------------------------------

responder_    False  True
antibiotics_             
ABX+              3     6
ABX-              6     4
(0.3333333333333333, 0.36984996427720923)

----------------------------------------------------------------------------------------------------

mayo_endo_improved  False  True
antibiotics_                   
ABX+                    4     7
ABX-                    7     4
(0.32653061224489793, 0.39485591807573345)

----------------------------------------------------------------------------------------------------
```

In [ ]:

```
d = recipient.assign(
    **{
        endpoint: recipient[endpoint].astype(
            float
        )  # Use float instead of pd.Boolean dtype.
        for endpoint in [
            "remission",
            "remission_",
            "responder",
            "responder_",
            "mayo_endo_improved",
            "mayo_endo_improved_",
        ]
    }
)

for formula in [
    "remission ~ antibiotics_",
    "remission_ ~ antibiotics_",
    "responder ~ antibiotics_",
    "responder_ ~ antibiotics_",
    "mayo_endo_improved ~ antibiotics_",
    "mayo_endo_improved_ ~ antibiotics_",
]:
    fit = smf.logit(formula, data=d).fit()
    print(fit.summary())
    print(fit.conf_int())
    _print_visual_separator()
```

```
Optimization terminated successfully.
         Current function value: 0.581574
         Iterations 5
                           Logit Regression Results                           
==============================================================================
Dep. Variable:              remission   No. Observations:                   22
Model:                          Logit   Df Residuals:                       20
Method:                           MLE   Df Model:                            1
Date:                Fri, 14 Jan 2022   Pseudo R-squ.:                  0.1128
Time:                        13:15:46   Log-Likelihood:                -12.795
converged:                       True   LL-Null:                       -14.421
Covariance Type:            nonrobust   LLR p-value:                   0.07134
========================================================================================
                           coef    std err          z      P>|z|      [0.025      0.975]
----------------------------------------------------------------------------------------
Intercept                0.1823      0.606      0.301      0.763      -1.004       1.369
antibiotics_[T.ABX-]    -1.6864      0.989     -1.705      0.088      -3.624       0.252
========================================================================================
                             0         1
Intercept            -1.004496  1.369139
antibiotics_[T.ABX-] -3.624463  0.251665

----------------------------------------------------------------------------------------------------

Optimization terminated successfully.
         Current function value: 0.564876
         Iterations 5
                           Logit Regression Results                           
==============================================================================
Dep. Variable:             remission_   No. Observations:                   19
Model:                          Logit   Df Residuals:                       17
Method:                           MLE   Df Model:                            1
Date:                Fri, 14 Jan 2022   Pseudo R-squ.:                  0.1701
Time:                        13:15:46   Log-Likelihood:                -10.733
converged:                       True   LL-Null:                       -12.932
Covariance Type:            nonrobust   LLR p-value:                   0.03597
========================================================================================
                           coef    std err          z      P>|z|      [0.025      0.975]
----------------------------------------------------------------------------------------
Intercept                0.6931      0.707      0.980      0.327      -0.693       2.079
antibiotics_[T.ABX-]    -2.0794      1.061     -1.961      0.050      -4.158      -0.001
========================================================================================
                             0         1
Intercept            -0.692757  2.079051
antibiotics_[T.ABX-] -4.158297 -0.000586

----------------------------------------------------------------------------------------------------

Optimization terminated successfully.
         Current function value: 0.672246
         Iterations 4
                           Logit Regression Results                           
==============================================================================
Dep. Variable:              responder   No. Observations:                   22
Model:                          Logit   Df Residuals:                       20
Method:                           MLE   Df Model:                            1
Date:                Fri, 14 Jan 2022   Pseudo R-squ.:                 0.02433
Time:                        13:15:46   Log-Likelihood:                -14.789
converged:                       True   LL-Null:                       -15.158
Covariance Type:            nonrobust   LLR p-value:                    0.3904
========================================================================================
                           coef    std err          z      P>|z|      [0.025      0.975]
----------------------------------------------------------------------------------------
Intercept                0.1823      0.606      0.301      0.763      -1.004       1.369
antibiotics_[T.ABX-]    -0.7419      0.872     -0.851      0.395      -2.450       0.966
========================================================================================
                             0         1
Intercept            -1.004496  1.369139
antibiotics_[T.ABX-] -2.450059  0.966184

----------------------------------------------------------------------------------------------------

Optimization terminated successfully.
         Current function value: 0.655723
         Iterations 4
                           Logit Regression Results                           
==============================================================================
Dep. Variable:             responder_   No. Observations:                   19
Model:                          Logit   Df Residuals:                       17
Method:                           MLE   Df Model:                            1
Date:                Fri, 14 Jan 2022   Pseudo R-squ.:                 0.05210
Time:                        13:15:46   Log-Likelihood:                -12.459
converged:                       True   LL-Null:                       -13.143
Covariance Type:            nonrobust   LLR p-value:                    0.2419
========================================================================================
                           coef    std err          z      P>|z|      [0.025      0.975]
----------------------------------------------------------------------------------------
Intercept                0.6931      0.707      0.980      0.327      -0.693       2.079
antibiotics_[T.ABX-]    -1.0986      0.957     -1.147      0.251      -2.975       0.778
========================================================================================
                             0         1
Intercept            -0.692757  2.079051
antibiotics_[T.ABX-] -2.975135  0.777910

----------------------------------------------------------------------------------------------------

Optimization terminated successfully.
         Current function value: 0.655482
         Iterations 4
                           Logit Regression Results                           
==============================================================================
Dep. Variable:     mayo_endo_improved   No. Observations:                   22
Model:                          Logit   Df Residuals:                       20
Method:                           MLE   Df Model:                            1
Date:                Fri, 14 Jan 2022   Pseudo R-squ.:                 0.05434
Time:                        13:15:46   Log-Likelihood:                -14.421
converged:                       True   LL-Null:                       -15.249
Covariance Type:            nonrobust   LLR p-value:                    0.1980
========================================================================================
                           coef    std err          z      P>|z|      [0.025      0.975]
----------------------------------------------------------------------------------------
Intercept                0.5596      0.627      0.893      0.372      -0.669       1.788
antibiotics_[T.ABX-]    -1.1192      0.886     -1.263      0.207      -2.857       0.618
========================================================================================
                             0         1
Intercept            -0.668857  1.788088
antibiotics_[T.ABX-] -2.856554  0.618091

----------------------------------------------------------------------------------------------------

Optimization terminated successfully.
         Current function value: 0.605130
         Iterations 5
                            Logit Regression Results                           
===============================================================================
Dep. Variable:     mayo_endo_improved_   No. Observations:                   19
Model:                           Logit   Df Residuals:                       17
Method:                            MLE   Df Model:                            1
Date:                 Fri, 14 Jan 2022   Pseudo R-squ.:                  0.1109
Time:                         13:15:46   Log-Likelihood:                -11.497
converged:                        True   LL-Null:                       -12.932
Covariance Type:             nonrobust   LLR p-value:                   0.09030
========================================================================================
                           coef    std err          z      P>|z|      [0.025      0.975]
----------------------------------------------------------------------------------------
Intercept                1.2528      0.802      1.562      0.118      -0.319       2.824
antibiotics_[T.ABX-]    -1.6582      1.029     -1.611      0.107      -3.676       0.359
========================================================================================
                             0         1
Intercept            -0.318704  2.824230
antibiotics_[T.ABX-] -3.675681  0.359225

----------------------------------------------------------------------------------------------------
```

In [ ]:

###### Enema vs. Capsules¶

In [ ]:

```
# 'remission_' and 'responder_' are both strict definitions,
# excluding participants that did not complete the trial.
for endpoint in [
    "remission",
    "remission_",
    "responder",
    "responder_",
    "mayo_endo_improved",
]:
    contingency = (
        recipient.groupby(["maintenance_", endpoint])
        .apply(len)
        .unstack()
        .fillna(0)
        .astype(int)
    )

    print(contingency)
    print(sp.stats.fisher_exact(contingency.values))
    _print_visual_separator()
```

```
remission     False  True
maintenance_             
CAPS              5     5
ENMA              9     3
(0.3333333333333333, 0.37770897832817296)

----------------------------------------------------------------------------------------------------

remission_    False  True
maintenance_             
CAPS              4     5
ENMA              7     3
(0.34285714285714286, 0.36984996427720895)

----------------------------------------------------------------------------------------------------

responder     False  True
maintenance_             
CAPS              4     6
ENMA              8     4
(0.3333333333333333, 0.39128363896165874)

----------------------------------------------------------------------------------------------------

responder_    False  True
maintenance_             
CAPS              3     6
ENMA              6     4
(0.3333333333333333, 0.36984996427720923)

----------------------------------------------------------------------------------------------------

mayo_endo_improved  False  True
maintenance_                   
CAPS                    4     6
ENMA                    7     5
(0.47619047619047616, 0.6699214098594928)

----------------------------------------------------------------------------------------------------
```

In [ ]:

```
d = recipient.assign(
    **{
        endpoint: recipient[endpoint].astype(
            float
        )  # Use float instead of pd.Boolean dtype.
        for endpoint in [
            "remission",
            "remission_",
            "responder",
            "responder_",
            "mayo_endo_improved",
            "mayo_endo_improved_",
        ]
    }
)

for formula in [
    "remission ~ maintenance_",
    "remission_ ~ maintenance_",
    "responder ~ maintenance_",
    "responder_ ~ maintenance_",
    "mayo_endo_improved ~ maintenance_",
    "mayo_endo_improved_ ~ maintenance_",
]:
    fit = smf.logit(formula, data=d).fit()
    print(fit.summary())
    print(fit.conf_int())
    _print_visual_separator()
```

```
Optimization terminated successfully.
         Current function value: 0.621795
         Iterations 5
                           Logit Regression Results                           
==============================================================================
Dep. Variable:              remission   No. Observations:                   22
Model:                          Logit   Df Residuals:                       20
Method:                           MLE   Df Model:                            1
Date:                Fri, 14 Jan 2022   Pseudo R-squ.:                 0.05139
Time:                        13:15:46   Log-Likelihood:                -13.679
converged:                       True   LL-Null:                       -14.421
Covariance Type:            nonrobust   LLR p-value:                    0.2234
========================================================================================
                           coef    std err          z      P>|z|      [0.025      0.975]
----------------------------------------------------------------------------------------
Intercept            -2.668e-16      0.632  -4.22e-16      1.000      -1.240       1.240
maintenance_[T.ENMA]    -1.0986      0.919     -1.196      0.232      -2.900       0.702
========================================================================================
                             0        1
Intercept            -1.239590  1.23959
maintenance_[T.ENMA] -2.899695  0.70247

----------------------------------------------------------------------------------------------------

Optimization terminated successfully.
         Current function value: 0.646910
         Iterations 5
                           Logit Regression Results                           
==============================================================================
Dep. Variable:             remission_   No. Observations:                   19
Model:                          Logit   Df Residuals:                       17
Method:                           MLE   Df Model:                            1
Date:                Fri, 14 Jan 2022   Pseudo R-squ.:                 0.04954
Time:                        13:15:47   Log-Likelihood:                -12.291
converged:                       True   LL-Null:                       -12.932
Covariance Type:            nonrobust   LLR p-value:                    0.2577
========================================================================================
                           coef    std err          z      P>|z|      [0.025      0.975]
----------------------------------------------------------------------------------------
Intercept                0.2231      0.671      0.333      0.739      -1.092       1.538
maintenance_[T.ENMA]    -1.0704      0.962     -1.112      0.266      -2.957       0.816
========================================================================================
                             0         1
Intercept            -1.091640  1.537927
maintenance_[T.ENMA] -2.956687  0.815804

----------------------------------------------------------------------------------------------------

Optimization terminated successfully.
         Current function value: 0.653104
         Iterations 4
                           Logit Regression Results                           
==============================================================================
Dep. Variable:              responder   No. Observations:                   22
Model:                          Logit   Df Residuals:                       20
Method:                           MLE   Df Model:                            1
Date:                Fri, 14 Jan 2022   Pseudo R-squ.:                 0.05211
Time:                        13:15:47   Log-Likelihood:                -14.368
converged:                       True   LL-Null:                       -15.158
Covariance Type:            nonrobust   LLR p-value:                    0.2088
========================================================================================
                           coef    std err          z      P>|z|      [0.025      0.975]
----------------------------------------------------------------------------------------
Intercept                0.4055      0.645      0.628      0.530      -0.860       1.671
maintenance_[T.ENMA]    -1.0986      0.890     -1.235      0.217      -2.843       0.645
========================================================================================
                             0         1
Intercept            -0.859686  1.670616
maintenance_[T.ENMA] -2.842503  0.645278

----------------------------------------------------------------------------------------------------

Optimization terminated successfully.
         Current function value: 0.655723
         Iterations 4
                           Logit Regression Results                           
==============================================================================
Dep. Variable:             responder_   No. Observations:                   19
Model:                          Logit   Df Residuals:                       17
Method:                           MLE   Df Model:                            1
Date:                Fri, 14 Jan 2022   Pseudo R-squ.:                 0.05210
Time:                        13:15:47   Log-Likelihood:                -12.459
converged:                       True   LL-Null:                       -13.143
Covariance Type:            nonrobust   LLR p-value:                    0.2419
========================================================================================
                           coef    std err          z      P>|z|      [0.025      0.975]
----------------------------------------------------------------------------------------
Intercept                0.6931      0.707      0.980      0.327      -0.693       2.079
maintenance_[T.ENMA]    -1.0986      0.957     -1.147      0.251      -2.975       0.778
========================================================================================
                             0         1
Intercept            -0.692757  2.079051
maintenance_[T.ENMA] -2.975135  0.777910

----------------------------------------------------------------------------------------------------

Optimization terminated successfully.
         Current function value: 0.676383
         Iterations 4
                           Logit Regression Results                           
==============================================================================
Dep. Variable:     mayo_endo_improved   No. Observations:                   22
Model:                          Logit   Df Residuals:                       20
Method:                           MLE   Df Model:                            1
Date:                Fri, 14 Jan 2022   Pseudo R-squ.:                 0.02418
Time:                        13:15:47   Log-Likelihood:                -14.880
converged:                       True   LL-Null:                       -15.249
Covariance Type:            nonrobust   LLR p-value:                    0.3904
========================================================================================
                           coef    std err          z      P>|z|      [0.025      0.975]
----------------------------------------------------------------------------------------
Intercept                0.4055      0.645      0.628      0.530      -0.860       1.671
maintenance_[T.ENMA]    -0.7419      0.872     -0.851      0.395      -2.450       0.966
========================================================================================
                             0         1
Intercept            -0.859686  1.670616
maintenance_[T.ENMA] -2.450059  0.966184

----------------------------------------------------------------------------------------------------

Optimization terminated successfully.
         Current function value: 0.666321
         Iterations 4
                            Logit Regression Results                           
===============================================================================
Dep. Variable:     mayo_endo_improved_   No. Observations:                   19
Model:                           Logit   Df Residuals:                       17
Method:                            MLE   Df Model:                            1
Date:                 Fri, 14 Jan 2022   Pseudo R-squ.:                 0.02102
Time:                         13:15:47   Log-Likelihood:                -12.660
converged:                        True   LL-Null:                       -12.932
Covariance Type:             nonrobust   LLR p-value:                    0.4609
========================================================================================
                           coef    std err          z      P>|z|      [0.025      0.975]
----------------------------------------------------------------------------------------
Intercept                0.6931      0.707      0.980      0.327      -0.693       2.079
maintenance_[T.ENMA]    -0.6931      0.949     -0.731      0.465      -2.553       1.166
========================================================================================
                             0         1
Intercept            -0.692757  2.079051
maintenance_[T.ENMA] -2.552532  1.166238

----------------------------------------------------------------------------------------------------
```

###### Donor¶

In [ ]:

```
d = recipient[
    lambda x: x.donor_subject_id != "D0065"
].assign(  # Drop the single subject who received donor 65.
    **{
        endpoint: recipient[endpoint].astype(
            float
        )  # Use float instead of pd.Boolean dtype.
        for endpoint in [
            "remission",
            "remission_",
            "responder",
            "responder_",
            "mayo_endo_improved",
            "mayo_endo_improved_",
        ]
    }
)

for formula in [
    "remission ~ donor_subject_id",
    "remission_ ~ donor_subject_id",
    "responder ~ donor_subject_id",
    "responder_ ~ donor_subject_id",
    "mayo_endo_improved ~ donor_subject_id",
    "mayo_endo_improved_ ~ donor_subject_id",
]:
    print(smf.logit(formula, data=d).fit().summary())
    _print_visual_separator()
```

```
Optimization terminated successfully.
         Current function value: 0.588687
         Iterations 5
                           Logit Regression Results                           
==============================================================================
Dep. Variable:              remission   No. Observations:                   21
Model:                          Logit   Df Residuals:                       18
Method:                           MLE   Df Model:                            2
Date:                Fri, 14 Jan 2022   Pseudo R-squ.:                 0.07514
Time:                        13:15:47   Log-Likelihood:                -12.362
converged:                       True   LL-Null:                       -13.367
Covariance Type:            nonrobust   LLR p-value:                    0.3663
=============================================================================================
                                coef    std err          z      P>|z|      [0.025      0.975]
---------------------------------------------------------------------------------------------
Intercept                    -1.0986      0.816     -1.346      0.178      -2.699       0.502
donor_subject_id[T.D0097]  3.146e-16      1.155   2.72e-16      1.000      -2.263       2.263
donor_subject_id[T.D0485]     1.5041      1.225      1.228      0.219      -0.896       3.905
=============================================================================================

----------------------------------------------------------------------------------------------------

Optimization terminated successfully.
         Current function value: 0.587061
         Iterations 5
                           Logit Regression Results                           
==============================================================================
Dep. Variable:             remission_   No. Observations:                   18
Model:                          Logit   Df Residuals:                       15
Method:                           MLE   Df Model:                            2
Date:                Fri, 14 Jan 2022   Pseudo R-squ.:                  0.1215
Time:                        13:15:47   Log-Likelihood:                -10.567
converged:                       True   LL-Null:                       -12.028
Covariance Type:            nonrobust   LLR p-value:                    0.2319
=============================================================================================
                                coef    std err          z      P>|z|      [0.025      0.975]
---------------------------------------------------------------------------------------------
Intercept                    -1.0986      0.816     -1.346      0.178      -2.699       0.502
donor_subject_id[T.D0097]     0.4055      1.190      0.341      0.733      -1.927       2.738
donor_subject_id[T.D0485]     2.1972      1.414      1.554      0.120      -0.575       4.969
=============================================================================================

----------------------------------------------------------------------------------------------------

Optimization terminated successfully.
         Current function value: 0.664289
         Iterations 4
                           Logit Regression Results                           
==============================================================================
Dep. Variable:              responder   No. Observations:                   21
Model:                          Logit   Df Residuals:                       18
Method:                           MLE   Df Model:                            2
Date:                Fri, 14 Jan 2022   Pseudo R-squ.:                 0.02726
Time:                        13:15:47   Log-Likelihood:                -13.950
converged:                       True   LL-Null:                       -14.341
Covariance Type:            nonrobust   LLR p-value:                    0.6764
=============================================================================================
                                coef    std err          z      P>|z|      [0.025      0.975]
---------------------------------------------------------------------------------------------
Intercept                    -0.5108      0.730     -0.699      0.484      -1.942       0.921
donor_subject_id[T.D0097]  2.765e-16      1.033   2.68e-16      1.000      -2.024       2.024
donor_subject_id[T.D0485]     0.9163      1.169      0.784      0.433      -1.375       3.208
=============================================================================================

----------------------------------------------------------------------------------------------------

Optimization terminated successfully.
         Current function value: 0.650041
         Iterations 5
                           Logit Regression Results                           
==============================================================================
Dep. Variable:             responder_   No. Observations:                   18
Model:                          Logit   Df Residuals:                       15
Method:                           MLE   Df Model:                            2
Date:                Fri, 14 Jan 2022   Pseudo R-squ.:                 0.06219
Time:                        13:15:47   Log-Likelihood:                -11.701
converged:                       True   LL-Null:                       -12.477
Covariance Type:            nonrobust   LLR p-value:                    0.4603
=============================================================================================
                                coef    std err          z      P>|z|      [0.025      0.975]
---------------------------------------------------------------------------------------------
Intercept                    -0.5108      0.730     -0.699      0.484      -1.942       0.921
donor_subject_id[T.D0097]     0.5108      1.095      0.466      0.641      -1.636       2.658
donor_subject_id[T.D0485]     1.6094      1.366      1.178      0.239      -1.068       4.287
=============================================================================================

----------------------------------------------------------------------------------------------------

Optimization terminated successfully.
         Current function value: 0.676321
         Iterations 4
                           Logit Regression Results                           
==============================================================================
Dep. Variable:     mayo_endo_improved   No. Observations:                   21
Model:                          Logit   Df Residuals:                       18
Method:                           MLE   Df Model:                            2
Date:                Fri, 14 Jan 2022   Pseudo R-squ.:                 0.02268
Time:                        13:15:47   Log-Likelihood:                -14.203
converged:                       True   LL-Null:                       -14.532
Covariance Type:            nonrobust   LLR p-value:                    0.7193
=============================================================================================
                                coef    std err          z      P>|z|      [0.025      0.975]
---------------------------------------------------------------------------------------------
Intercept                 -1.205e-17      0.707   -1.7e-17      1.000      -1.386       1.386
donor_subject_id[T.D0097]    -0.5108      1.017     -0.503      0.615      -2.503       1.482
donor_subject_id[T.D0485]     0.4055      1.155      0.351      0.725      -1.858       2.669
=============================================================================================

----------------------------------------------------------------------------------------------------

Optimization terminated successfully.
         Current function value: 0.664078
         Iterations 5
                            Logit Regression Results                           
===============================================================================
Dep. Variable:     mayo_endo_improved_   No. Observations:                   18
Model:                           Logit   Df Residuals:                       15
Method:                            MLE   Df Model:                            2
Date:                 Fri, 14 Jan 2022   Pseudo R-squ.:                 0.03331
Time:                         13:15:47   Log-Likelihood:                -11.953
converged:                        True   LL-Null:                       -12.365
Covariance Type:             nonrobust   LLR p-value:                    0.6624
=============================================================================================
                                coef    std err          z      P>|z|      [0.025      0.975]
---------------------------------------------------------------------------------------------
Intercept                 -1.991e-16      0.707  -2.82e-16      1.000      -1.386       1.386
donor_subject_id[T.D0097]  2.166e-16      1.080   2.01e-16      1.000      -2.117       2.117
donor_subject_id[T.D0485]     1.0986      1.354      0.811      0.417      -1.555       3.752
=============================================================================================

----------------------------------------------------------------------------------------------------
```

##### Mayo Score Improvement¶

In [ ]:

```
fig, ax = plt.subplots(figsize=(8, 3))

d0 = recipient.assign(
    withdrawal_due_to_failure=lambda x: x.withdrawal_due_to_failure.astype(float)
).assign(jitter=lambda x: np.linspace(-0.1, 0.1, num=len(x)))
# d0 = recipient.dropna(subset=['mayo_total_start', 'mayo_total_end'])
# x_shift_range = np.linspace(-0.001, 0.001, num=d0.shape[0])
# d0 = d0.sort_values(['mayo_total_start', 'mayo_total_end'], ascending=False).assign(offset=x_shift_range)

plot_kwsA = dict(alpha=1.0)
for subject_id, d1 in d0.iterrows():
    if np.isnan(d1.mayo_total_start):
        continue
    elif d1.withdrawal_due_to_failure == 1.0:
        plot_kwsB = dict(linestyle="--", lw=1.5)
        d1["mayo_total_end"] = d1["mayo_total_start"]
    elif np.isnan(d1.mayo_total_end):
        continue
    else:
        plot_kwsB = dict(linestyle="-", lw=2)
    ax.annotate(
        "",
        xytext=(d1["mayo_total_start"], 0 + d1["jitter"]),
        xy=(d1["mayo_total_end"], 1 + d1["jitter"]),
        arrowprops=dict(
            arrowstyle="-|>",
            color=lib.project_style.DEFAULT_COLOR_PALETTE[d1["arm"]],
            **plot_kwsA,
            **plot_kwsB,
        ),
        # #         marker=lib.project_style.DEFAULT_MARKER_PALETTE[d1['donor_subject_id']],
        # #         marker='o',
        #         color=lib.project_style.DEFAULT_COLOR_PALETTE[d1['arm']],
        #         label='__nolegend__',
        #         **plot_kwsA,
        #         **plot_kwsB,
    )

for arm in lib.project_style.ARM_ORDER:
    ax.plot(
        [],
        [],
        color=lib.project_style.DEFAULT_COLOR_PALETTE[arm],
        label=arm,
        lw=2,
        **plot_kwsA,
    )
# for donor_subject_id in ['D0044', 'D0097', 'D0485', 'D0065']:
#     ax.plot([], [], linestyle='none', marker=lib.project_style.DEFAULT_MARKER_PALETTE[donor_subject_id], color='grey', label=donor_subject_id)

# ax.legend(bbox_to_anchor=(1, 1))
ax.axvline(
    2.5,
    lw=1,
    linestyle="--",
    color="grey",
    zorder=0,
)
ax.set_xlabel("Mayo Score")
ax.set_yticks([0, 1])
ax.set_ylim(-0.2, 1.2)
ax.set_xlim(-0.5, 10.5)
ax.set_yticklabels(["D0", "F1"])
ax.invert_xaxis()
ax.invert_yaxis()

ax.set_ylabel("← Time")
ax.text(0.2, 0.0, "← worse", fontsize=8, transform=fig.transFigure)
ax.text(0.77, 0.0, "better →", fontsize=8, transform=fig.transFigure)

plt.text(
    -0.05, 1.05, "B", fontsize=12, fontweight="heavy", transform=plt.gca().transAxes
)

fig.savefig("fig/mayo_endpoint_arrows.pdf", bbox_inches="tight")
```

In [ ]:

```
d = recipient.dropna(subset=["mayo_total_start", "mayo_total_end"])

sp.stats.wilcoxon(
    d.mayo_total_start, d.mayo_total_end, "pratt", correction=True, mode="exact"
)
```

Out[ ]:

```
WilcoxonResult(statistic=41.0, pvalue=0.01531219482421875)
```

###### GEE with Partial Mayo Scores¶

In [ ]:

```
## TODO: See "strain_species_analysis.ipynb" GEE sections.
```

In [ ]:

In [ ]:

```
efficacy_gee_data = (
    subject_x_visit_type.reset_index()
    .set_index("visit_id")
    .join(recipient, on="subject_id")
    .assign(
        remission=lambda x: x.remission.astype(float),
        responder=lambda x: x.responder.astype(float),
        mayo_endo_improved=lambda x: x.mayo_endo_improved.astype(float),
        weeks_post_fmt=lambda x: x.days_post_fmt / 7,
    )
    .drop(
        columns=[
            "treatment_abx_pre",
            "withdrawal_due_to_failure",
            "remission_",
            "responder_",
            "mayo_endo_improved_",
        ]
    )[lambda x: x.visit_class.isin(["maintenance", "followup"])]
    .sort_values(["subject_id", "weeks_post_fmt"])
)
```

In [ ]:

```
%%R -i efficacy_gee_data

library("geepack")

variables = c("weeks_post_fmt", "antibiotics_", "donor_subject_id", "maintenance_", "status_mayo_score_stool_frequency", "status_mayo_score_rectal_bleeding")
efficacy_gee_data$subject_id <- as.factor(efficacy_gee_data$subject_id)

efficacy_gee_data <- efficacy_gee_data[complete.cases(efficacy_gee_data[variables]),]

confint.geeglm <- function(object, parm, level = 0.95) {
    cc <- coef(summary(object))
    mult <- qnorm((1+level)/2)
    citab <- with(as.data.frame(cc),
                  cbind(lwr=Estimate-mult*Std.err,
                        upr=Estimate+mult*Std.err))
    rownames(citab) <- rownames(cc)
    citab[parm,]
}
# engraftment_gee_data_sr <- engraftment_gee_data[complete.cases(engraftment_gee_data[c('subject_rabund', variables)]),]
# engraftment_gee_data_db <- engraftment_gee_data[complete.cases(engraftment_gee_data[c('donor_bc', variables)]),]
# engraftment_gee_data_sb <- engraftment_gee_data[complete.cases(engraftment_gee_data[c('baseline_bc', variables)]),]
```

###### Null Model¶

In [ ]:

```
%%R

# WARNING: d$subject_id (or whatever "id" is, must be sorted)

lm0_mayo_sf <- geeglm(
    status_mayo_score_stool_frequency ~ weeks_post_fmt,
    id=subject_id,
    data=efficacy_gee_data,
    corstr="ar1",
    waves=efficacy_gee_data$weeks_post_fmt,
)
print(summary(lm0_mayo_sf))
print('--------')

lm0_mayo_rb <- geeglm(
    status_mayo_score_rectal_bleeding ~ weeks_post_fmt,
    id=subject_id,
    data=efficacy_gee_data,
    corstr="ar1",
    waves=efficacy_gee_data$weeks_post_fmt,
)
print(summary(lm0_mayo_rb))
print('--------')
```

```
Call:
geeglm(formula = status_mayo_score_stool_frequency ~ weeks_post_fmt, 
    data = efficacy_gee_data, id = subject_id, waves = efficacy_gee_data$weeks_post_fmt, 
    corstr = "ar1")

 Coefficients:
               Estimate  Std.err  Wald Pr(>|W|)    
(Intercept)     1.64747  0.18061 83.21  < 2e-16 ***
weeks_post_fmt -0.04386  0.01313 11.15 0.000839 ***
---
Signif. codes:  0 ‘***’ 0.001 ‘**’ 0.01 ‘*’ 0.05 ‘.’ 0.1 ‘ ’ 1

Correlation structure = ar1 
Estimated Scale Parameters:

            Estimate Std.err
(Intercept)   0.7742  0.1232
  Link = identity 

Estimated Correlation Parameters:
      Estimate Std.err
alpha        0       0
Number of clusters:   24  Maximum cluster size: 9 
[1] "--------"

Call:
geeglm(formula = status_mayo_score_rectal_bleeding ~ weeks_post_fmt, 
    data = efficacy_gee_data, id = subject_id, waves = efficacy_gee_data$weeks_post_fmt, 
    corstr = "ar1")

 Coefficients:
               Estimate  Std.err Wald Pr(>|W|)    
(Intercept)     0.80939  0.16067 25.4  4.7e-07 ***
weeks_post_fmt -0.03512  0.00758 21.5  3.6e-06 ***
---
Signif. codes:  0 ‘***’ 0.001 ‘**’ 0.01 ‘*’ 0.05 ‘.’ 0.1 ‘ ’ 1

Correlation structure = ar1 
Estimated Scale Parameters:

            Estimate Std.err
(Intercept)    0.583   0.106
  Link = identity 

Estimated Correlation Parameters:
      Estimate Std.err
alpha        0       0
Number of clusters:   24  Maximum cluster size: 9 
[1] "--------"
```

In [ ]:

```
%%R

# WARNING: d$subject_id (or whatever "id" is, must be sorted)

lm_abx_mayo_sf <- geeglm(
    status_mayo_score_stool_frequency ~ weeks_post_fmt + antibiotics_,
    id=subject_id,
    data=efficacy_gee_data,
    corstr="ar1",
    waves=efficacy_gee_data$weeks_post_fmt,
)
print(summary(lm_abx_mayo_sf))
print(confint.geeglm(lm_abx_mayo_sf))
print(anova(lm0_mayo_sf, lm_abx_mayo_sf))
print('--------')

lm_abx_mayo_rb <- geeglm(
    status_mayo_score_rectal_bleeding ~ weeks_post_fmt + antibiotics_,
    id=subject_id,
    data=efficacy_gee_data,
    corstr="ar1",
    waves=efficacy_gee_data$weeks_post_fmt,
)
print(summary(lm_abx_mayo_rb))
print(confint.geeglm(lm_abx_mayo_rb))
print(anova(lm0_mayo_rb, lm_abx_mayo_rb))
print('--------')
```

```
Call:
geeglm(formula = status_mayo_score_stool_frequency ~ weeks_post_fmt + 
    antibiotics_, data = efficacy_gee_data, id = subject_id, 
    waves = efficacy_gee_data$weeks_post_fmt, corstr = "ar1")

 Coefficients:
                 Estimate Std.err  Wald Pr(>|W|)    
(Intercept)        1.5221  0.2026 56.44  5.8e-14 ***
weeks_post_fmt    -0.0426  0.0129 10.99  0.00092 ***
antibiotics_ABX+   0.2705  0.2998  0.81  0.36679    
---
Signif. codes:  0 ‘***’ 0.001 ‘**’ 0.01 ‘*’ 0.05 ‘.’ 0.1 ‘ ’ 1

Correlation structure = ar1 
Estimated Scale Parameters:

            Estimate Std.err
(Intercept)    0.756   0.112
  Link = identity 

Estimated Correlation Parameters:
      Estimate Std.err
alpha        0       0
Number of clusters:   24  Maximum cluster size: 9 
                     lwr     upr
(Intercept)       1.1250  1.9191
weeks_post_fmt   -0.0679 -0.0174
antibiotics_ABX+ -0.3170  0.8581
Analysis of 'Wald statistic' Table

Model 1 status_mayo_score_stool_frequency ~ weeks_post_fmt + antibiotics_ 
Model 2 status_mayo_score_stool_frequency ~ weeks_post_fmt
  Df    X2 P(>|Chi|)
1  1 0.815      0.37
[1] "--------"

Call:
geeglm(formula = status_mayo_score_rectal_bleeding ~ weeks_post_fmt + 
    antibiotics_, data = efficacy_gee_data, id = subject_id, 
    waves = efficacy_gee_data$weeks_post_fmt, corstr = "ar1")

 Coefficients:
                 Estimate  Std.err  Wald Pr(>|W|)    
(Intercept)       0.92477  0.20726 19.91  8.1e-06 ***
weeks_post_fmt   -0.03624  0.00737 24.20  8.7e-07 ***
antibiotics_ABX+ -0.24892  0.24001  1.08      0.3    
---
Signif. codes:  0 ‘***’ 0.001 ‘**’ 0.01 ‘*’ 0.05 ‘.’ 0.1 ‘ ’ 1

Correlation structure = ar1 
Estimated Scale Parameters:

            Estimate Std.err
(Intercept)    0.568  0.0988
  Link = identity 

Estimated Correlation Parameters:
      Estimate Std.err
alpha        0       0
Number of clusters:   24  Maximum cluster size: 9 
                     lwr     upr
(Intercept)       0.5185  1.3310
weeks_post_fmt   -0.0507 -0.0218
antibiotics_ABX+ -0.7193  0.2215
Analysis of 'Wald statistic' Table

Model 1 status_mayo_score_rectal_bleeding ~ weeks_post_fmt + antibiotics_ 
Model 2 status_mayo_score_rectal_bleeding ~ weeks_post_fmt
  Df   X2 P(>|Chi|)
1  1 1.08       0.3
[1] "--------"
```

In [ ]:

```
%%R

# WARNING: d$subject_id (or whatever "id" is, must be sorted)

lm_dnr_mayo_sf <- geeglm(
    status_mayo_score_stool_frequency ~ weeks_post_fmt + donor_subject_id,
    id=subject_id,
    data=efficacy_gee_data,
    corstr="ar1",
    waves=efficacy_gee_data$weeks_post_fmt,
)
print(summary(lm_dnr_mayo_sf))
print(confint.geeglm(lm_dnr_mayo_sf))
print(anova(lm0_mayo_sf, lm_dnr_mayo_sf))
print('--------')

lm_dnr_mayo_rb <- geeglm(
    status_mayo_score_rectal_bleeding ~ weeks_post_fmt + donor_subject_id,
    id=subject_id,
    data=efficacy_gee_data,
    corstr="ar1",
    waves=efficacy_gee_data$weeks_post_fmt,
)
print(summary(lm_dnr_mayo_rb))
print(confint.geeglm(lm_dnr_mayo_rb))
print(anova(lm0_mayo_rb, lm_dnr_mayo_rb))
print('--------')
```

```
Call:
geeglm(formula = status_mayo_score_stool_frequency ~ weeks_post_fmt + 
    donor_subject_id, data = efficacy_gee_data, id = subject_id, 
    waves = efficacy_gee_data$weeks_post_fmt, corstr = "ar1")

 Coefficients:
                      Estimate Std.err  Wald Pr(>|W|)    
(Intercept)             1.9417  0.2038 90.75   <2e-16 ***
weeks_post_fmt         -0.0396  0.0132  9.04   0.0026 ** 
donor_subject_idD0065  -0.0657  0.1778  0.14   0.7119    
donor_subject_idD0097  -0.6214  0.3031  4.20   0.0404 *  
donor_subject_idD0485  -0.3449  0.3360  1.05   0.3047    
---
Signif. codes:  0 ‘***’ 0.001 ‘**’ 0.01 ‘*’ 0.05 ‘.’ 0.1 ‘ ’ 1

Correlation structure = ar1 
Estimated Scale Parameters:

            Estimate Std.err
(Intercept)    0.701   0.119
  Link = identity 

Estimated Correlation Parameters:
      Estimate Std.err
alpha        0       0
Number of clusters:   24  Maximum cluster size: 9 
                          lwr     upr
(Intercept)            1.5422  2.3412
weeks_post_fmt        -0.0655 -0.0138
donor_subject_idD0065 -0.4142  0.2828
donor_subject_idD0097 -1.2155 -0.0273
donor_subject_idD0485 -1.0035  0.3136
Analysis of 'Wald statistic' Table

Model 1 status_mayo_score_stool_frequency ~ weeks_post_fmt + donor_subject_id 
Model 2 status_mayo_score_stool_frequency ~ weeks_post_fmt
  Df   X2 P(>|Chi|)
1  3 5.86      0.12
[1] "--------"

Call:
geeglm(formula = status_mayo_score_rectal_bleeding ~ weeks_post_fmt + 
    donor_subject_id, data = efficacy_gee_data, id = subject_id, 
    waves = efficacy_gee_data$weeks_post_fmt, corstr = "ar1")

 Coefficients:
                      Estimate  Std.err  Wald Pr(>|W|)    
(Intercept)            1.07657  0.23040 21.83  3.0e-06 ***
weeks_post_fmt        -0.03619  0.00798 20.59  5.7e-06 ***
donor_subject_idD0065 -0.92885  0.21752 18.23  2.0e-05 ***
donor_subject_idD0097 -0.25744  0.29267  0.77    0.379    
donor_subject_idD0485 -0.57403  0.26312  4.76    0.029 *  
---
Signif. codes:  0 ‘***’ 0.001 ‘**’ 0.01 ‘*’ 0.05 ‘.’ 0.1 ‘ ’ 1

Correlation structure = ar1 
Estimated Scale Parameters:

            Estimate Std.err
(Intercept)     0.52     0.1
  Link = identity 

Estimated Correlation Parameters:
      Estimate Std.err
alpha        0       0
Number of clusters:   24  Maximum cluster size: 9 
                          lwr     upr
(Intercept)            0.6250  1.5281
weeks_post_fmt        -0.0518 -0.0206
donor_subject_idD0065 -1.3552 -0.5025
donor_subject_idD0097 -0.8311  0.3162
donor_subject_idD0485 -1.0897 -0.0583
Analysis of 'Wald statistic' Table

Model 1 status_mayo_score_rectal_bleeding ~ weeks_post_fmt + donor_subject_id 
Model 2 status_mayo_score_rectal_bleeding ~ weeks_post_fmt
  Df   X2 P(>|Chi|)    
1  3 30.5   1.1e-06 ***
---
Signif. codes:  0 ‘***’ 0.001 ‘**’ 0.01 ‘*’ 0.05 ‘.’ 0.1 ‘ ’ 1
[1] "--------"
```

In [ ]:

```
%%R

# WARNING: d$subject_id (or whatever "id" is, must be sorted)

lm_mnt_mayo_sf <- geeglm(
    status_mayo_score_stool_frequency ~ weeks_post_fmt + maintenance_,
    id=subject_id,
    data=efficacy_gee_data,
    corstr="ar1",
    waves=efficacy_gee_data$weeks_post_fmt,
)
print(summary(lm_mnt_mayo_sf))
print(confint.geeglm(lm_mnt_mayo_sf))
print(anova(lm0_mayo_sf, lm_mnt_mayo_sf))
print('--------')

lm_mnt_mayo_rb <- geeglm(
    status_mayo_score_rectal_bleeding ~ weeks_post_fmt + maintenance_,
    id=subject_id,
    data=efficacy_gee_data,
    corstr="ar1",
    waves=efficacy_gee_data$weeks_post_fmt,
)
print(summary(lm_mnt_mayo_rb))
print(confint.geeglm(lm_mnt_mayo_rb))
print(anova(lm0_mayo_rb, lm_mnt_mayo_rb))
print('--------')
```

```
Call:
geeglm(formula = status_mayo_score_stool_frequency ~ weeks_post_fmt + 
    maintenance_, data = efficacy_gee_data, id = subject_id, 
    waves = efficacy_gee_data$weeks_post_fmt, corstr = "ar1")

 Coefficients:
                 Estimate Std.err  Wald Pr(>|W|)    
(Intercept)        1.7362  0.1975 77.32  < 2e-16 ***
weeks_post_fmt    -0.0443  0.0133 11.09  0.00087 ***
maintenance_ENMA  -0.1582  0.2788  0.32  0.57028    
---
Signif. codes:  0 ‘***’ 0.001 ‘**’ 0.01 ‘*’ 0.05 ‘.’ 0.1 ‘ ’ 1

Correlation structure = ar1 
Estimated Scale Parameters:

            Estimate Std.err
(Intercept)    0.768   0.124
  Link = identity 

Estimated Correlation Parameters:
      Estimate Std.err
alpha        0       0
Number of clusters:   24  Maximum cluster size: 9 
                     lwr     upr
(Intercept)       1.3492  2.1232
weeks_post_fmt   -0.0703 -0.0182
maintenance_ENMA -0.7046  0.3881
Analysis of 'Wald statistic' Table

Model 1 status_mayo_score_stool_frequency ~ weeks_post_fmt + maintenance_ 
Model 2 status_mayo_score_stool_frequency ~ weeks_post_fmt
  Df    X2 P(>|Chi|)
1  1 0.322      0.57
[1] "--------"

Call:
geeglm(formula = status_mayo_score_rectal_bleeding ~ weeks_post_fmt + 
    maintenance_, data = efficacy_gee_data, id = subject_id, 
    waves = efficacy_gee_data$weeks_post_fmt, corstr = "ar1")

 Coefficients:
                 Estimate  Std.err  Wald Pr(>|W|)    
(Intercept)       0.72725  0.19519 13.88  0.00019 ***
weeks_post_fmt   -0.03475  0.00725 22.96  1.7e-06 ***
maintenance_ENMA  0.14644  0.24866  0.35  0.55592    
---
Signif. codes:  0 ‘***’ 0.001 ‘**’ 0.01 ‘*’ 0.05 ‘.’ 0.1 ‘ ’ 1

Correlation structure = ar1 
Estimated Scale Parameters:

            Estimate Std.err
(Intercept)    0.578   0.102
  Link = identity 

Estimated Correlation Parameters:
      Estimate Std.err
alpha        0       0
Number of clusters:   24  Maximum cluster size: 9 
                    lwr     upr
(Intercept)       0.345  1.1098
weeks_post_fmt   -0.049 -0.0205
maintenance_ENMA -0.341  0.6338
Analysis of 'Wald statistic' Table

Model 1 status_mayo_score_rectal_bleeding ~ weeks_post_fmt + maintenance_ 
Model 2 status_mayo_score_rectal_bleeding ~ weeks_post_fmt
  Df    X2 P(>|Chi|)
1  1 0.347      0.56
[1] "--------"
```

#### Subsection: Multi-modal longitudinal characterization of the microbiome¶

##### Data sizes¶

In [ ]:

```
d = sample_x_rotu_cvrg.drop(donor_means_list)

d.sum().sum() / len(d.index)
```

Out[ ]:

```
336683.22222222225
```

In [ ]:

```
# 16S ASVs
d = sample_x_rotu.drop(donor_means_list)

thresh = 1e-3

((d > thresh).sum() >= 2).sum()
```

Out[ ]:

```
857
```

In [ ]:

```
# UHGG Species
d = sample_x_motu.drop(donor_means_list)

thresh = 1e-3

((d > thresh).sum() >= 2).sum()
```

Out[ ]:

```
371
```

In [ ]:

```
# Hybrid species/StrainFinder haplotypes

# TODO: Figure out why I'm not seeing any strain columns for species that were not
# deconvolved.

thresh = 1e-3

d = sample_x_sotu.drop(donor_means_list)

((d > thresh).sum() >= 1).sum()
```

Out[ ]:

```
3846
```

In [ ]:

```
# KO Annotations

d = sample_x_ko_cvrg.drop(donor_means_list)
((d >= 1).sum() >= 2).sum()
```

Out[ ]:

```
7587
```

In [ ]:

```
len(sample_x_chem_ba.columns)
```

Out[ ]:

```
51
```

#### Subsection: Subject and donor clustering within datasets¶

sample\_x\_rotu.join(m, how='left').groupby(['subject\_id', 'sample\_type']).apply(len).unstack()

In [ ]:

```
m = (
    sample.join(subject, on="subject_id", lsuffix="_")
    .assign(subject_type=lambda x: x.recipient.map({True: "recipient", False: "donor"}))
    .assign(zorder=lambda x: x.recipient.map({True: 0, False: 1}))
)

s = m.sample_type.isin(lib.project_data.SAMPLE_TYPE_ORDER_SIMPLE + ["donor_mean"])
edgecolor_palette = {
    "donor": "black",
    "baseline": "grey",
    "maintenance": "none",
    "followup": "none",
}

markersize_palette = {
    "recipient": 30,
    "donor": 90,
}

fig, axs = plt.subplots(2, 2, figsize=(8, 8))
for (name, d, metric, panel), ax in zip(
    [
        ("sotu", sample_x_sotu, "braycurtis", "A"),
        ("motu", sample_x_motu, "braycurtis", "B"),
        #     ('eggnog', sample_x_eggnog_cvrg, 'cosine'),
        ("ko", sample_x_ko_cvrg, "cosine", "C"),
        ("chem_ba", sample_x_chem_ba_std2, "cosine", "D"),
    ],
    axs.flatten(),
):
    ax, _ = lib.plot.ordination_plot(
        data=d,
        meta=m,
        subset=s,
        ordin_kws=dict(metric=metric),
        xy=("PC1", "PC2"),
        colorby="subject_id",
        color_palette=subject_color_palette,
        colorby_order=list(subject_color_palette),
        edgecolorby="sample_class",
        edgecolor_palette=edgecolor_palette,
        edgecolorby_order=list(edgecolor_palette),
        markerby="donor_subject_id",
        marker_palette=lib.project_style.DEFAULT_MARKER_PALETTE,
        markersizeby="subject_type",
        markersize_palette=markersize_palette,
        markersizeby_order=list(markersize_palette),
        zorderby="zorder",
        scatter_kws=dict(lw=2, alpha=0.65),
        frac_explained_label=False,
        ax=ax,
        fill_legend=False,
    )
    ax.set_title(lib.project_style.OMICS_NAME[name])
    ax.get_legend().set_visible(False)
    ax.set_xticks([-0.5, 0.5])
    ax.set_yticks([-0.5, 0.5])
    ax.text(-0.0, 1.07, panel, fontsize=12, fontweight="heavy", transform=ax.transAxes)
    ax.set_ylabel("")
    ax.set_xlabel("")

for ax in axs[-1, :]:
    ax.set_xlabel("NMDS1")
for ax in axs[:, 0]:
    ax.set_ylabel("NMDS2")

fig.tight_layout()
fig.savefig("fig/multiomics_ordination_by_subject.pdf", bbox_inches="tight")


# Legend
fig, axs = plt.subplots(1, 4, figsize=(5, 0.5))

donor_ax = {"D0044": 0, "D0097": 1, "D0485": 2, "other": 3}

for subject_id, d in subject.loc[subject_has_mgen_order].iterrows():
    s = {True: 40, False: 80}[d.recipient]
    marker = lib.project_style.DEFAULT_MARKER_PALETTE[d.donor_subject_id]
    lw, edgecolor = {True: (0, "none"), False: (2, "black")}[d.recipient]
    c = subject_color_palette[subject_id]
    axs[donor_ax[d.donor_subject_id]].scatter(
        [], [], c=[c], s=s, marker=marker, lw=lw, edgecolor=edgecolor, label=subject_id
    )

for sample_class, label in {"baseline": "B", "maintenance": "D1-F3"}.items():
    s = 60
    marker = "s"
    lw, edgecolor = 2, edgecolor_palette[sample_class]
    c = "lightgrey"
    axs[donor_ax["other"]].scatter(
        [], [], c=[c], s=s, marker=marker, lw=lw, edgecolor=edgecolor, label=label
    )

for subject_type in ["donor"]:
    s = markersize_palette[subject_type]
    marker = "s"
    c = "lightgrey"
    edgecolor = edgecolor_palette[subject_type]
    axs[donor_ax["other"]].scatter(
        [], [], c=[c], lw=2, edgecolor=edgecolor, s=s, marker=marker, label=subject_type
    )

for ax in axs:
    ax.legend()
    for spines in ["top", "right", "bottom", "left"]:
        ax.spines[spines].set_visible(False)
    ax.set_xticks([])
    ax.set_yticks([])
fig.tight_layout()
fig.savefig("fig/ordination_by_subject_legend.pdf", bbox_inches="tight")

# Legend
fig, ax = plt.subplots(figsize=(0.5, 0.5))

donor_ax = {"D0044": 0, "D0097": 1, "D0485": 2, "other": 3}

for subject_id, d in subject.loc[subject_has_mgen_order].iterrows():
    s = {True: 40, False: 80}[d.recipient]
    marker = lib.project_style.DEFAULT_MARKER_PALETTE[d.donor_subject_id]
    lw, edgecolor = {True: (0, "none"), False: (2, "black")}[d.recipient]
    c = subject_color_palette[subject_id]
    ax.scatter(
        [],
        [],
        c=[c],
        s=s,
        marker=marker,
        lw=lw,
        edgecolor=edgecolor,
        label=subject_id,
        alpha=0.65,
    )

for sample_class, label in {"baseline": "B", "maintenance": "D1-F3"}.items():
    s = 60
    marker = "s"
    lw, edgecolor = 2, edgecolor_palette[sample_class]
    c = "lightgrey"
    ax.scatter(
        [], [], c=[c], s=s, marker=marker, lw=lw, edgecolor=edgecolor, label=label
    )

for subject_type in ["donor"]:
    s = markersize_palette[subject_type]
    marker = "s"
    c = "lightgrey"
    edgecolor = edgecolor_palette[subject_type]
    ax.scatter(
        [], [], c=[c], lw=2, edgecolor=edgecolor, s=s, marker=marker, label=subject_type
    )

ax.legend()
for spines in ["top", "right", "bottom", "left"]:
    ax.spines[spines].set_visible(False)
ax.set_xticks([])
ax.set_yticks([])
fig.tight_layout()
fig.savefig("fig/ordination_by_subject_legend_vertical.pdf", bbox_inches="tight")
```

```
No artists with labels found to put in legend.  Note that artists whose label start with an underscore are ignored when legend() is called with no argument.
No artists with labels found to put in legend.  Note that artists whose label start with an underscore are ignored when legend() is called with no argument.
No artists with labels found to put in legend.  Note that artists whose label start with an underscore are ignored when legend() is called with no argument.
No artists with labels found to put in legend.  Note that artists whose label start with an underscore are ignored when legend() is called with no argument.
/var/folders/vv/8wpk7cln4lgb9qyqchjvcsmh0000gq/T/ipykernel_17922/1673015093.py:109: UserWarning: Tight layout not applied. The bottom and top margins cannot be made large enough to accommodate all axes decorations.
  fig.tight_layout()
/var/folders/vv/8wpk7cln4lgb9qyqchjvcsmh0000gq/T/ipykernel_17922/1673015093.py:157: UserWarning: Tight layout not applied. The left and right margins cannot be made large enough to accommodate all axes decorations.
  fig.tight_layout()
```

In [ ]:

```
m = (
    sample.join(subject, on="subject_id", lsuffix="_")
    .assign(subject_type=lambda x: x.recipient.map({True: "recipient", False: "donor"}))
    .assign(zorder=lambda x: x.recipient.map({True: 0, False: 1}))
)

s = m.sample_type.isin(lib.project_data.SAMPLE_TYPE_ORDER_SIMPLE + ["donor_mean"])
edgecolor_palette = {
    "donor": "black",
    "baseline": "grey",
    "maintenance": "none",
    "followup": "none",
}

markersize_palette = {
    "recipient": 30,
    "donor": 90,
}

fig, axs = plt.subplots(2, 2, figsize=(8, 8))
for (name, d, metric, panel), ax in zip(
    [
        ("sotu", sample_x_sotu, "braycurtis", "A"),
        ("motu", sample_x_motu, "braycurtis", "B"),
        #     ('eggnog', sample_x_eggnog_cvrg, 'cosine'),
        ("ko", sample_x_ko_cvrg, "cosine", "C"),
        ("chem_ba", sample_x_chem_ba_std2, "cosine", "D"),
    ],
    axs.flatten(),
):
    ax, _ = lib.plot.ordination_plot(
        data=d,
        meta=m,
        subset=s,
        ordin_kws=dict(metric=metric),
        xy=("PC1", "PC2"),
        colorby="donor_subject_id",
        color_palette=subject_color_palette,
        colorby_order=list(subject_color_palette),
        edgecolorby="sample_class",
        edgecolor_palette=edgecolor_palette,
        edgecolorby_order=list(edgecolor_palette),
        markerby="donor_subject_id",
        marker_palette=lib.project_style.DEFAULT_MARKER_PALETTE,
        markersizeby="subject_type",
        markersize_palette=markersize_palette,
        markersizeby_order=list(markersize_palette),
        zorderby="zorder",
        scatter_kws=dict(lw=2, alpha=0.75),
        frac_explained_label=False,
        ax=ax,
        fill_legend=False,
    )
    ax.set_title(lib.project_style.OMICS_NAME[name])
    ax.get_legend().set_visible(False)
    ax.set_xticks([-0.5, 0.5])
    ax.set_yticks([-0.5, 0.5])
    ax.text(-0.0, 1.07, panel, fontsize=12, fontweight="heavy", transform=ax.transAxes)
    ax.set_ylabel("")
    ax.set_xlabel("")

for ax in axs[-1, :]:
    ax.set_xlabel("NMDS1")
for ax in axs[:, 0]:
    ax.set_ylabel("NMDS2")

fig.tight_layout()
fig.savefig("fig/multiomics_ordination_by_donor.pdf", bbox_inches="tight")


# Legend as a figure
fig, ax = plt.subplots(1, 1, figsize=(1, 0.5))

for donor_subject_id in donor_order:
    s = markersize_palette["recipient"]
    marker = lib.project_style.DEFAULT_MARKER_PALETTE[donor_subject_id]
    c = subject_color_palette[donor_subject_id]
    ax.scatter([], [], c=[c], s=s, marker=marker, label=donor_subject_id)

for sample_class, label in {"baseline": "B", "maintenance": "D1-F3"}.items():
    s = markersize_palette["recipient"]
    marker = "s"
    lw, edgecolor = 2, edgecolor_palette[sample_class]
    c = "lightgrey"
    ax.scatter(
        [], [], c=[c], s=s, marker=marker, lw=lw, edgecolor=edgecolor, label=label
    )

for subject_type in ["donor"]:
    s = markersize_palette[subject_type]
    marker = "s"
    c = "lightgrey"
    edgecolor = edgecolor_palette[subject_type]
    ax.scatter(
        [], [], c=[c], lw=2, edgecolor=edgecolor, s=s, marker=marker, label=subject_type
    )

ax.legend(ncol=6)
for spines in ["top", "right", "bottom", "left"]:
    ax.spines[spines].set_visible(False)
ax.set_xticks([])
ax.set_yticks([])
fig.savefig("fig/ordination_by_donor_legend_horizontal.pdf", bbox_inches="tight")


# Legend as a figure
fig, ax = plt.subplots(1, 1, figsize=(1, 0.5))

for donor_subject_id in donor_order:
    s = markersize_palette["recipient"]
    marker = lib.project_style.DEFAULT_MARKER_PALETTE[donor_subject_id]
    c = subject_color_palette[donor_subject_id]
    ax.scatter([], [], c=[c], s=s, marker=marker, label=donor_subject_id)

for sample_class, label in {"baseline": "B", "maintenance": "D1-F3"}.items():
    s = markersize_palette["recipient"]
    marker = "s"
    lw, edgecolor = 2, edgecolor_palette[sample_class]
    c = "lightgrey"
    ax.scatter(
        [], [], c=[c], s=s, marker=marker, lw=lw, edgecolor=edgecolor, label=label
    )

for subject_type in ["donor"]:
    s = markersize_palette[subject_type]
    marker = "s"
    c = "lightgrey"
    edgecolor = edgecolor_palette[subject_type]
    ax.scatter(
        [], [], c=[c], lw=2, edgecolor=edgecolor, s=s, marker=marker, label=subject_type
    )

ax.legend(ncol=1)
for spines in ["top", "right", "bottom", "left"]:
    ax.spines[spines].set_visible(False)
ax.set_xticks([])
ax.set_yticks([])
fig.savefig("fig/ordination_by_donor_legend_vertical.pdf", bbox_inches="tight")
```

```
No artists with labels found to put in legend.  Note that artists whose label start with an underscore are ignored when legend() is called with no argument.
No artists with labels found to put in legend.  Note that artists whose label start with an underscore are ignored when legend() is called with no argument.
No artists with labels found to put in legend.  Note that artists whose label start with an underscore are ignored when legend() is called with no argument.
No artists with labels found to put in legend.  Note that artists whose label start with an underscore are ignored when legend() is called with no argument.
```

In [ ]:

```
grouping = "donor_subject_id"

donor_anosim_results = []
for dname, d, metric in tqdm(
    [
        ("sotu", sample_x_sotu, "braycurtis"),
        ("motu", sample_x_motu, "braycurtis"),
        ("rotu", sample_x_rotu, "braycurtis"),
        #     ('eggnog', sample_x_eggnog_cvrg, 'cosine'),
        ("ko", sample_x_ko_cvrg, "cosine"),
        #     ('keggmodule', sample_x_keggmodule_cvrg, 'cosine'),
        #     ('chem', sample_x_chem_std2, 'cosine'),
        ("chem_ba", sample_x_chem_ba_std2, "cosine"),
    ],
    ascii=True,
):
    for sample_type in lib.project_data.SAMPLE_TYPE_ORDER_SIMPLE:
        m = sample.join(subject, on="subject_id", lsuffix="_").assign(
            subject_type=lambda x: x.recipient.map({True: "recipient", False: "donor"})
        )
        d, m = lib.pandas.align_indexes(d, m)
        dmat = lib.dissimilarity.dmatrix(d, metric=metric)
        donor_anosim_results.append(
            (
                dname,
                sample_type,
                *lib.stats.anosim(
                    dmat, m[grouping], subset=m.sample_type.isin([sample_type]), n=9999
                )[["test statistic", "p-value"]],
            )
        )

donor_anosim_results = pd.DataFrame(
    donor_anosim_results, columns=["dataset", "sample_type", "r", "p"]
)

donor_anosim_r = donor_anosim_results.set_index(["sample_type", "dataset"])[
    "r"
].unstack()
donor_anosim_p = donor_anosim_results.set_index(["sample_type", "dataset"])[
    "p"
].unstack()

donor_anosim_r
```

```
100%|##########################################################################################################################| 5/5 [00:30<00:00,  6.10s/it]
```

Out[ ]:

| dataset | chem\_ba | ko | motu | rotu | sotu |
| --- | --- | --- | --- | --- | --- |
| sample\_type |  |  |  |  |  |
| baseline | -0.051775 | 0.238166 | 0.014793 | -0.094482 | -0.090237 |
| followup\_1 | 0.183432 | 0.174556 | 0.334320 | 0.136054 | 0.961538 |
| followup\_2 | 0.223373 | -0.031065 | 0.180473 | 0.240870 | 0.928994 |
| pre\_maintenance\_1 | 0.088000 | 0.248000 | 0.344000 | 0.284382 | 1.000000 |
| pre\_maintenance\_2 | 0.095238 | 0.092437 | 0.532213 | 0.399111 | 1.000000 |
| pre\_maintenance\_3 | 0.109467 | 0.211538 | 0.439349 | 0.211462 | 1.000000 |
| pre\_maintenance\_4 | 0.008876 | 0.109467 | 0.375740 | 0.404755 | 0.998521 |
| pre\_maintenance\_5 | 0.140533 | 0.337566 | 0.422222 | 0.302871 | 0.995767 |
| pre\_maintenance\_6 | 0.034056 | -0.021672 | 0.151703 | 0.181818 | 1.000000 |

In [ ]:

```
donor_anosim_p
```

Out[ ]:

| dataset | chem\_ba | ko | motu | rotu | sotu |
| --- | --- | --- | --- | --- | --- |
| sample\_type |  |  |  |  |  |
| baseline | 0.6146 | 0.0390 | 0.4453 | 0.6787 | 0.7454 |
| followup\_1 | 0.0922 | 0.1084 | 0.0126 | 0.1125 | 0.0002 |
| followup\_2 | 0.0599 | 0.5375 | 0.0889 | 0.0478 | 0.0001 |
| pre\_maintenance\_1 | 0.2464 | 0.0462 | 0.0491 | 0.0298 | 0.0077 |
| pre\_maintenance\_2 | 0.2360 | 0.2647 | 0.0040 | 0.0140 | 0.0009 |
| pre\_maintenance\_3 | 0.1991 | 0.0701 | 0.0022 | 0.0372 | 0.0001 |
| pre\_maintenance\_4 | 0.4307 | 0.1988 | 0.0052 | 0.0027 | 0.0001 |
| pre\_maintenance\_5 | 0.1403 | 0.0168 | 0.0029 | 0.0173 | 0.0004 |
| pre\_maintenance\_6 | 0.3786 | 0.5140 | 0.1665 | 0.1327 | 0.0001 |

In [ ]:

```
def _p_to_s(p):
    if p < 0.05:
        return 40
    elif p < 0.10:
        return 15
    else:
        return 0


sample_type_order_idx = pd.Series(
    {v: i for i, v in enumerate(lib.project_data.SAMPLE_TYPE_ORDER_SIMPLE)}
)
sample_type_tick_labels = [
    lib.project_style.SAMPLE_TYPE_LABELS[sample_type]
    for sample_type in lib.project_data.SAMPLE_TYPE_ORDER_SIMPLE
]

assert len(sample_type_tick_labels) == len(sample_type_order_idx)

datatypes = ["sotu", "motu", "rotu", "ko", "chem_ba"]
zorder_palette = {"sotu": 0, "motu": 1, "rotu": 2, "ko": 3, "chem_ba": 4}

fig, ax = plt.subplots(figsize=(3, 2.9))

for col in datatypes:
    d0 = pd.DataFrame(
        dict(
            r=donor_anosim_r[col],
            p=donor_anosim_p[col],
            s=donor_anosim_p[col].map(_p_to_s),
            x=sample_type_order_idx,
        )
    ).sort_values("x")
    ax.plot(
        "x",
        "r",
        data=d0,
        label="__nolegend__",
        c=lib.project_style.DEFAULT_COLOR_PALETTE[col],
        alpha=1.0,
        zorder=zorder_palette[col],
    )
    ax.plot(
        [],
        [],
        marker="o",
        linestyle="-",
        label=lib.project_style.OMICS_NAME[col],
        c=lib.project_style.DEFAULT_COLOR_PALETTE[col],
        alpha=1.0,
    )
    for _, d1 in d0.iterrows():
        annotation = lib.project_style.pvalue_to_annotation(d1.p)
        if annotation == "∙":
            yoffset = 0.02
        elif annotation == "**":
            yoffset = 0.01
        elif annotation == "*":
            yoffset = 0.01
        elif annotation == "":
            yoffset = 0
        ax.annotate(
            annotation,
            xy=(d1.x, d1.r + yoffset),
            color=lib.project_style.DEFAULT_COLOR_PALETTE[col],
            ha="center",
            va="center",
            zorder=zorder_palette[col],
        )
#     plt.scatter('x', 'r', data=d, s='s', label='__nolegend__', vmin=0, vmax=1, marker='o')
#     plt.xticks(ticks=d.x, labels=idxwhere(sample_type_order_idx.isin(d.x)), rotation=45, ha='right')

# # Dummy points for legend:
# plt.plot([], [], markersize=_p_to_s(0.04)/5, color='grey', linestyle='-', marker='o', label='p < 0.05')
# plt.plot([], [], markersize=_p_to_s(0.09)/5, color='grey', linestyle='-', marker='o', label='p < 0.10')
# # plt.plot([], [], markersize=_p_to_s(0.051)/6, color='grey', linestyle='-', marker='o', label='p>=0.05')


ax.set_ylim(-0.15, 1.15)
# ax.set_yticklabels([])

# ax.legend(bbox_to_anchor=(0.4, 0.465))
ax.set_ylabel("ANOSIM R (donor)")
ax.set_xlabel("Time-point")
plt.xticks(range(len(sample_type_tick_labels)), sample_type_tick_labels)
# ax.set_title('By Donor')
ax.text(-0.0, 1.05, "F", fontsize=12, fontweight="heavy", transform=plt.gca().transAxes)
fig.savefig("fig/multiomics_anosim_by_donor.pdf", bbox_inches="tight")

# Legend
fig, ax = plt.subplots(figsize=(1, 1))
for col in datatypes:
    ax.plot(
        [],
        [],
        marker="o",
        linestyle="-",
        label=lib.project_style.OMICS_NAME[col],
        c=lib.project_style.DEFAULT_COLOR_PALETTE[col],
        alpha=0.8,
    )
ax.legend()
ax.legend(ncol=1)
for spines in ["top", "right", "bottom", "left"]:
    ax.spines[spines].set_visible(False)
ax.set_xticks([])
ax.set_yticks([])

donor_anosim_p
fig.savefig("fig/multiomics_line_point_legend.pdf", bbox_inches="tight")
```

```
/var/folders/vv/8wpk7cln4lgb9qyqchjvcsmh0000gq/T/ipykernel_17922/4114870097.py:34: RuntimeWarning: Second argument 'r' is ambiguous: could be a format string but is in 'data'; using as data.  If it was intended as data, set the format string to an empty string to suppress this warning.  If it was intended as a format string, explicitly pass the x-values as well.  Alternatively, rename the entry in 'data'.
  ax.plot(
/var/folders/vv/8wpk7cln4lgb9qyqchjvcsmh0000gq/T/ipykernel_17922/4114870097.py:34: RuntimeWarning: Second argument 'r' is ambiguous: could be a format string but is in 'data'; using as data.  If it was intended as data, set the format string to an empty string to suppress this warning.  If it was intended as a format string, explicitly pass the x-values as well.  Alternatively, rename the entry in 'data'.
  ax.plot(
/var/folders/vv/8wpk7cln4lgb9qyqchjvcsmh0000gq/T/ipykernel_17922/4114870097.py:34: RuntimeWarning: Second argument 'r' is ambiguous: could be a format string but is in 'data'; using as data.  If it was intended as data, set the format string to an empty string to suppress this warning.  If it was intended as a format string, explicitly pass the x-values as well.  Alternatively, rename the entry in 'data'.
  ax.plot(
/var/folders/vv/8wpk7cln4lgb9qyqchjvcsmh0000gq/T/ipykernel_17922/4114870097.py:34: RuntimeWarning: Second argument 'r' is ambiguous: could be a format string but is in 'data'; using as data.  If it was intended as data, set the format string to an empty string to suppress this warning.  If it was intended as a format string, explicitly pass the x-values as well.  Alternatively, rename the entry in 'data'.
  ax.plot(
/var/folders/vv/8wpk7cln4lgb9qyqchjvcsmh0000gq/T/ipykernel_17922/4114870097.py:34: RuntimeWarning: Second argument 'r' is ambiguous: could be a format string but is in 'data'; using as data.  If it was intended as data, set the format string to an empty string to suppress this warning.  If it was intended as a format string, explicitly pass the x-values as well.  Alternatively, rename the entry in 'data'.
  ax.plot(
```

In [ ]:

```
grouping = "sex"

_anosim_results = []
for dname, d, metric in tqdm(
    [
        ("sotu", sample_x_sotu, "braycurtis"),
        ("motu", sample_x_motu, "braycurtis"),
        ("rotu", sample_x_rotu, "braycurtis"),
        #     ('eggnog', sample_x_eggnog_cvrg, 'cosine'),
        ("ko", sample_x_ko_cvrg, "cosine"),
        #     ('keggmodule', sample_x_keggmodule_cvrg, 'cosine'),
        #     ('chem', sample_x_chem_std2, 'cosine'),
        ("chem_ba", sample_x_chem_ba_std2, "cosine"),
    ],
    ascii=True,
):
    for sample_type in lib.project_data.SAMPLE_TYPE_ORDER_SIMPLE:
        m = sample.join(subject, on="subject_id", lsuffix="_").assign(
            subject_type=lambda x: x.recipient.map({True: "recipient", False: "donor"})
        )
        d, m = lib.pandas.align_indexes(d, m)
        dmat = lib.dissimilarity.dmatrix(d, metric=metric)
        _anosim_results.append(
            (
                dname,
                sample_type,
                *lib.stats.anosim(
                    dmat, m[grouping], subset=m.sample_type.isin([sample_type]), n=9999
                )[["test statistic", "p-value"]],
            )
        )

_anosim_results = pd.DataFrame(
    _anosim_results, columns=["dataset", "sample_type", "r", "p"]
)

sex_anosim_r = _anosim_results.set_index(["sample_type", "dataset"])[
    "r"
].unstack()
sex_anosim_p = _anosim_results.set_index(["sample_type", "dataset"])[
    "p"
].unstack()

sex_anosim_r
```

```
100%|##########################################################################################################################| 5/5 [00:27<00:00,  5.49s/it]
```

Out[ ]:

| dataset | chem\_ba | ko | motu | rotu | sotu |
| --- | --- | --- | --- | --- | --- |
| sample\_type |  |  |  |  |  |
| baseline | 0.074074 | -0.046296 | 0.103175 | 0.024052 | -0.023810 |
| followup\_1 | 0.085979 | -0.007937 | 0.003968 | 0.108844 | -0.048942 |
| followup\_2 | -0.048942 | -0.047619 | 0.011905 | 0.153790 | 0.144180 |
| pre\_maintenance\_1 | -0.124000 | 0.160000 | 0.432000 | 0.114742 | 0.024000 |
| pre\_maintenance\_2 | -0.166667 | 0.082667 | 0.042667 | 0.039444 | -0.136000 |
| pre\_maintenance\_3 | -0.070106 | -0.064815 | -0.095238 | 0.026476 | -0.085979 |
| pre\_maintenance\_4 | -0.051587 | 0.140212 | -0.070106 | -0.016741 | -0.058201 |
| pre\_maintenance\_5 | -0.015873 | 0.074074 | 0.016667 | 0.064732 | -0.133333 |
| pre\_maintenance\_6 | -0.141333 | 0.144000 | 0.157333 | 0.055556 | 0.085333 |

In [ ]:

```
def _p_to_s(p):
    if p < 0.05:
        return 40
    elif p < 0.10:
        return 15
    else:
        return 0


sample_type_order_idx = pd.Series(
    {v: i for i, v in enumerate(lib.project_data.SAMPLE_TYPE_ORDER_SIMPLE)}
)
sample_type_tick_labels = [
    lib.project_style.SAMPLE_TYPE_LABELS[sample_type]
    for sample_type in lib.project_data.SAMPLE_TYPE_ORDER_SIMPLE
]

assert len(sample_type_tick_labels) == len(sample_type_order_idx)

datatypes = ["sotu", "motu", "rotu", "ko", "chem_ba"]
zorder_palette = {"sotu": 0, "motu": 1, "rotu": 2, "ko": 3, "chem_ba": 4}

fig, ax = plt.subplots(figsize=(3, 2.9))

for col in datatypes:
    d0 = pd.DataFrame(
        dict(
            r=sex_anosim_r[col],
            p=sex_anosim_p[col],
            s=sex_anosim_p[col].map(_p_to_s),
            x=sample_type_order_idx,
        )
    ).sort_values("x")
    ax.plot(
        "x",
        "r",
        data=d0,
        label="__nolegend__",
        c=lib.project_style.DEFAULT_COLOR_PALETTE[col],
        alpha=1.0,
        zorder=zorder_palette[col],
    )
    ax.plot(
        [],
        [],
        marker="o",
        linestyle="-",
        label=lib.project_style.OMICS_NAME[col],
        c=lib.project_style.DEFAULT_COLOR_PALETTE[col],
        alpha=1.0,
    )
    for _, d1 in d0.iterrows():
        annotation = lib.project_style.pvalue_to_annotation(d1.p)
        if annotation == "∙":
            yoffset = 0.02
        elif annotation == "**":
            yoffset = 0.01
        elif annotation == "*":
            yoffset = 0.01
        elif annotation == "":
            yoffset = 0
        ax.annotate(
            annotation,
            xy=(d1.x, d1.r + yoffset),
            color=lib.project_style.DEFAULT_COLOR_PALETTE[col],
            ha="center",
            va="center",
            zorder=zorder_palette[col],
        )
#     plt.scatter('x', 'r', data=d, s='s', label='__nolegend__', vmin=0, vmax=1, marker='o')
#     plt.xticks(ticks=d.x, labels=idxwhere(sample_type_order_idx.isin(d.x)), rotation=45, ha='right')

# # Dummy points for legend:
# plt.plot([], [], markersize=_p_to_s(0.04)/5, color='grey', linestyle='-', marker='o', label='p < 0.05')
# plt.plot([], [], markersize=_p_to_s(0.09)/5, color='grey', linestyle='-', marker='o', label='p < 0.10')
# # plt.plot([], [], markersize=_p_to_s(0.051)/6, color='grey', linestyle='-', marker='o', label='p>=0.05')


ax.set_ylim(-0.15, 1.15)
# ax.set_yticklabels([])

# ax.legend(bbox_to_anchor=(0.4, 0.465))
ax.set_ylabel("ANOSIM R (sex)")
ax.set_xlabel("Time-point")
plt.xticks(range(len(sample_type_tick_labels)), sample_type_tick_labels)


# for col in datatypes:
#     ax.plot(
#         [],
#         [],
#         marker="o",
#         linestyle="-",
#         label=lib.project_style.OMICS_NAME[col],
#         c=lib.project_style.DEFAULT_COLOR_PALETTE[col],
#         alpha=0.8,
#     )
ax.legend()

# ax.set_title('By Donor')
# ax.text(-0.0, 1.05, "F", fontsize=12, fontweight="heavy", transform=plt.gca().transAxes)
fig.savefig("doc/static/multiomics_anosim_by_sex_figure.svg", bbox_inches="tight")
```

```
/var/folders/vv/8wpk7cln4lgb9qyqchjvcsmh0000gq/T/ipykernel_17922/1964846424.py:34: RuntimeWarning: Second argument 'r' is ambiguous: could be a format string but is in 'data'; using as data.  If it was intended as data, set the format string to an empty string to suppress this warning.  If it was intended as a format string, explicitly pass the x-values as well.  Alternatively, rename the entry in 'data'.
  ax.plot(
/var/folders/vv/8wpk7cln4lgb9qyqchjvcsmh0000gq/T/ipykernel_17922/1964846424.py:34: RuntimeWarning: Second argument 'r' is ambiguous: could be a format string but is in 'data'; using as data.  If it was intended as data, set the format string to an empty string to suppress this warning.  If it was intended as a format string, explicitly pass the x-values as well.  Alternatively, rename the entry in 'data'.
  ax.plot(
/var/folders/vv/8wpk7cln4lgb9qyqchjvcsmh0000gq/T/ipykernel_17922/1964846424.py:34: RuntimeWarning: Second argument 'r' is ambiguous: could be a format string but is in 'data'; using as data.  If it was intended as data, set the format string to an empty string to suppress this warning.  If it was intended as a format string, explicitly pass the x-values as well.  Alternatively, rename the entry in 'data'.
  ax.plot(
/var/folders/vv/8wpk7cln4lgb9qyqchjvcsmh0000gq/T/ipykernel_17922/1964846424.py:34: RuntimeWarning: Second argument 'r' is ambiguous: could be a format string but is in 'data'; using as data.  If it was intended as data, set the format string to an empty string to suppress this warning.  If it was intended as a format string, explicitly pass the x-values as well.  Alternatively, rename the entry in 'data'.
  ax.plot(
/var/folders/vv/8wpk7cln4lgb9qyqchjvcsmh0000gq/T/ipykernel_17922/1964846424.py:34: RuntimeWarning: Second argument 'r' is ambiguous: could be a format string but is in 'data'; using as data.  If it was intended as data, set the format string to an empty string to suppress this warning.  If it was intended as a format string, explicitly pass the x-values as well.  Alternatively, rename the entry in 'data'.
  ax.plot(
```

In [ ]:

```
grouping = "subject_id"
m = sample.join(subject, on="subject_id", lsuffix="_").assign(
    subject_type=lambda x: x.recipient.map({True: "recipient", False: "donor"})
)
subset = m.sample_type.isin(lib.project_data.SAMPLE_TYPE_ORDER_SIMPLE)

subject_anosim_results = []
for dname, d, metric in [
    ("sotu", sample_x_sotu, "braycurtis"),
    ("motu", sample_x_motu, "braycurtis"),
    ("rotu", sample_x_rotu, "braycurtis"),
    #     ('eggnog', sample_x_eggnog_cvrg, 'cosine'),
    ("ko", sample_x_ko_cvrg, "cosine"),
    #     ('keggmodule', sample_x_keggmodule_cvrg, 'cosine'),
    ("chem_ba", sample_x_chem_ba_std2, "cosine"),
    #     ('chem', sample_x_chem_std2, 'cosine'),
]:
    d, m = lib.pandas.align_indexes(d, m)
    dmat = lib.dissimilarity.dmatrix(d, metric="braycurtis")
    subject_anosim_results.append(
        (
            dname,
            *lib.stats.anosim(dmat, m[grouping], subset=subset)[
                ["test statistic", "p-value"]
            ],
        )
    )

subject_anosim_results = pd.DataFrame(
    subject_anosim_results, columns=["dataset", "r", "p"]
)
subject_anosim_results
```

Out[ ]:

|  | dataset | r | p |
| --- | --- | --- | --- |
| 0 | sotu | 0.895606 | 0.001 |
| 1 | motu | 0.723178 | 0.001 |
| 2 | rotu | 0.736131 | 0.001 |
| 3 | ko | 0.330117 | 0.001 |
| 4 | chem\_ba | 0.509164 | 0.001 |

In [ ]:

```
fig, ax = plt.subplots(figsize=(0.5, 2.9))

datatypes = ["sotu", "motu", "rotu", "ko", "chem_ba"]

d0 = subject_anosim_results.set_index("dataset")

for col in datatypes:
    d1 = d0.loc[col]
    ax.scatter(
        [0],
        d1.r,
        marker="o",
        s=20,
        label=lib.project_style.OMICS_NAME[col],
        c=lib.project_style.DEFAULT_COLOR_PALETTE[col],
        alpha=1.0,
    )
    annotation = lib.project_style.pvalue_to_annotation(d1.p)
    if annotation == "∙":
        yoffset = 0.03
    elif annotation == "**":
        yoffset = 0.02
    elif annotation == "*":
        yoffset = 0.02
    elif annotation == "":
        yoffset = 0
    ax.annotate(
        annotation,
        xy=(0, d1.r + yoffset),
        color=lib.project_style.DEFAULT_COLOR_PALETTE[col],
        ha="center",
        va="center",
        alpha=1.0,
    )

# ax.set_yticks([])
ax.set_ylim(-0.15, 1.15)
ax.set_xticks([0])
ax.set_xticklabels(["D1-F2\n(pooled)"])
plt.ylabel("ANOSIM R (subject)")
# plt.title('By Subject')
plt.text(
    -0.0, 1.05, "G", fontsize=12, fontweight="heavy", transform=plt.gca().transAxes
)
fig.savefig("fig/multiomics_anosim_by_subject.pdf", bbox_inches="tight")
```

In [ ]:

```
import skbio as skb

m = (
    sample.join(subject, on="subject_id", lsuffix="_")
    .assign(subject_type=lambda x: x.recipient.map({True: "recipient", False: "donor"}))
    .assign(zorder=lambda x: x.recipient.map({True: 0, False: 1}))
)

tree = skb.io.read(
    "sraw/2020-12-30-Dose_Finding_Study_Box_mirror/16S data files/dada2_optim_tree_OTUtips_FMT_VED_May2019.tre",
    into=skb.TreeNode,
).root_at_midpoint()

s = m.sample_type.isin(lib.project_data.SAMPLE_TYPE_ORDER_SIMPLE + ["donor_mean"])
edgecolor_palette = {
    "donor": "black",
    "baseline": "black",
    "maintenance": "none",
    "followup": "grey",
}

markersize_palette = {
    "recipient": 30,
    "donor": 80,
}

fig, ax = plt.subplots()
name, d, metric, panel, ax = (
    "rotu",
    sample_x_rotu_cvrg.rename(columns=lambda s: s.replace("_", " ")),
    "braycurtis",
    "X",
    ax,
)

ax, _ = lib.plot.ordination_plot(
    data=d,
    meta=m,
    subset=s,
    ordin_kws=dict(
        pdm_func=lambda d: skb.diversity.beta_diversity(
            "weighted_unifrac",
            counts=d,
            ids=d.index,
            otu_ids=d.columns,
            tree=tree,
            normalized=True,
        ).to_data_frame()
    ),
    xy=("PC1", "PC2"),
    colorby="subject_id",
    color_palette=subject_color_palette,
    colorby_order=list(subject_color_palette),
    edgecolorby="sample_class",
    edgecolor_palette=edgecolor_palette,
    edgecolorby_order=list(edgecolor_palette),
    markerby="donor_subject_id",
    marker_palette=lib.project_style.DEFAULT_MARKER_PALETTE,
    markersizeby="subject_type",
    markersize_palette=markersize_palette,
    markersizeby_order=list(markersize_palette),
    zorderby="zorder",
    scatter_kws=dict(lw=2, alpha=0.65),
    frac_explained_label=False,
    ax=ax,
    fill_legend=False,
)
ax.set_title(lib.project_style.OMICS_NAME[name])
ax.get_legend().set_visible(False)
ax.set_xticks([-0.5, 0.5])
ax.set_yticks([-0.5, 0.5])
ax.text(-0.0, 1.07, panel, fontsize=12, fontweight="heavy", transform=ax.transAxes)
# ax.set_ylabel('')
# ax.set_xlabel('')
```

```
No artists with labels found to put in legend.  Note that artists whose label start with an underscore are ignored when legend() is called with no argument.
```

Out[ ]:

```
Text(-0.0, 1.07, 'X')
```

In [ ]:

```
m = (
    sample.join(subject, on="subject_id", lsuffix="_")
    .assign(subject_type=lambda x: x.recipient.map({True: "recipient", False: "donor"}))
    .assign(zorder=lambda x: x.recipient.map({True: 0, False: 1}))
)

s = m.sample_type.isin(lib.project_data.SAMPLE_TYPE_ORDER_SIMPLE + ["donor_mean"])
edgecolor_palette = {
    "donor": "black",
    "baseline": "grey",
    "maintenance": "none",
    "followup": "none",
}

markersize_palette = {
    "recipient": 30,
    "donor": 90,
}

fig, axs = plt.subplots(2, 3, figsize=(9, 6))
for (name, d, metric, panel, colorby), ax in zip(
    [
        ("sotu", sample_x_sotu, "braycurtis", "A", "donor_subject_id"),
        ("motu", sample_x_motu, "braycurtis", "B", "donor_subject_id"),
        #     ('eggnog', sample_x_eggnog_cvrg, 'cosine', 'donor_subject_id'),
        ("ko", sample_x_ko_cvrg, "cosine", "C", "donor_subject_id"),
        ("chem_ba", sample_x_chem_ba_std2, "cosine", "D", "donor_subject_id"),
        ("chem_ba", sample_x_chem_ba_std2, "cosine", "E", "subject_id"),
    ],
    axs.flatten(),
):
    ax, _ = lib.plot.ordination_plot(
        data=d,
        meta=m,
        subset=s,
        ordin_kws=dict(metric=metric),
        xy=("PC1", "PC2"),
        colorby=colorby,
        color_palette=subject_color_palette,
        colorby_order=list(subject_color_palette),
        edgecolorby="sample_class",
        edgecolor_palette=edgecolor_palette,
        edgecolorby_order=list(edgecolor_palette),
        markerby="donor_subject_id",
        marker_palette=lib.project_style.DEFAULT_MARKER_PALETTE,
        markersizeby="subject_type",
        markersize_palette=markersize_palette,
        markersizeby_order=list(markersize_palette),
        zorderby="zorder",
        scatter_kws=dict(lw=2, alpha=0.65),
        frac_explained_label=False,
        ax=ax,
        fill_legend=False,
    )
    ax.set_title(lib.project_style.OMICS_NAME[name])
    ax.get_legend().set_visible(False)
    ax.set_xticks([-0.5, 0.5])
    ax.set_yticks([-0.5, 0.5])
    ax.text(-0.0, 1.07, panel, fontsize=12, fontweight="heavy", transform=ax.transAxes)
    ax.set_ylabel("")
    ax.set_xlabel("")

for ax in axs[-1, :]:
    ax.set_xlabel("NMDS1")
for ax in axs[:, 0]:
    ax.set_ylabel("NMDS2")

# Remove visual evidence of bottom right axis.
axs[0, -1].set_xlabel("NMDS1")
for spines in ["top", "right", "bottom", "left"]:
    axs[-1, -1].spines[spines].set_visible(False)
axs[-1, -1].set_xticks([])
axs[-1, -1].set_yticks([])
axs[-1, -1].set_xlabel(None)
axs[-1, -1].set_ylabel(None)

fig.tight_layout()
fig.savefig("fig/multiomics_ordination_combo.pdf", bbox_inches="tight")
```

```
No artists with labels found to put in legend.  Note that artists whose label start with an underscore are ignored when legend() is called with no argument.
No artists with labels found to put in legend.  Note that artists whose label start with an underscore are ignored when legend() is called with no argument.
No artists with labels found to put in legend.  Note that artists whose label start with an underscore are ignored when legend() is called with no argument.
No artists with labels found to put in legend.  Note that artists whose label start with an underscore are ignored when legend() is called with no argument.
No artists with labels found to put in legend.  Note that artists whose label start with an underscore are ignored when legend() is called with no argument.
```

#### Subsection: Transfer and engraftment of donor taxa in patients¶

In [ ]:

```
subject_to_baseline_sample = (
    sample[sample.sample_type == "baseline"]
    .reset_index()
    .set_index("subject_id")
    .sample_id
)
donor_to_donor_mean = (
    sample[sample.sample_type == "donor_mean"]
    .reset_index()
    .set_index("subject_id")
    .sample_id.rename_axis("donor_subject_id")
)
all_donor_mean_samples = donor_to_donor_mean.to_list()

assert subject_to_baseline_sample.index.is_unique
assert donor_to_donor_mean.index.is_unique
subject_to_baseline_sample, donor_to_donor_mean
```

Out[ ]:

```
(subject_id
 S0001    SS01002
 S0004    SS01000
 S0007    SS01020
 S0008    SS01038
 S0013    SS01013
 S0017    SS01021
 S0021    SS01068
 S0024    SS01057
 S0027    SS01090
 S0029    SS01058
 S0031    SS01041
 S0037    SS01095
 S0041    SS01117
 S0043    SS01099
 S0047    SS01135
 S0053    SS01134
 S0055    SS01150
 S0056    SS01164
 S0058    SS01178
 S0059    SS01173
 S0060    SS01180
 S0061    SS01179
 S0062    SS01198
 Name: sample_id, dtype: object,
 donor_subject_id
 D0044    D0044_mean
 D0097    D0097_mean
 D0485    D0485_mean
 Name: sample_id, dtype: object)
```

##### ASV-level analysis¶

In [ ]:

```
d0 = sample_x_rotu
thresh = 1e-6  # Thresh used for jaccard presence/absence.

taxon_class = {}
taxon_class_rabund = []
taxon_class_count = []
baseline_bc = []
donor_bc = []
baseline_jacc = []
donor_jacc = []


for subject_id, d1 in d0.groupby(sample.subject_id):
    if not subject_id in recipient_has_mgen_order:
        continue

    donor_sample_id = donor_to_donor_mean[subject.loc[subject_id, "donor_subject_id"]]
    other_donor_samples = list(set(all_donor_mean_samples) - set([donor_sample_id]))
    donor_bc.append(
        pd.Series(
            sp.spatial.distance.cdist(
                d0.loc[[donor_sample_id]], d1, metric="braycurtis"
            ).squeeze(),
            index=d1.index,
        )
    )
    donor_jacc.append(
        pd.Series(
            sp.spatial.distance.cdist(
                d0.loc[[donor_sample_id]] > thresh, d1 > thresh, metric="jaccard"
            ).squeeze(),
            index=d1.index,
        )
    )

    if not subject_to_baseline_sample[subject_id] in d0.index:
        warn(f"No baseline sample for {subject_id}. Skipping this subject.")
        continue
    baseline_sample_id = subject_to_baseline_sample[subject_id]
    baseline_bc.append(
        pd.Series(
            sp.spatial.distance.cdist(
                d0.loc[[baseline_sample_id]], d1, metric="braycurtis"
            ).squeeze(),
            index=d1.index,
        )
    )
    baseline_jacc.append(
        pd.Series(
            sp.spatial.distance.cdist(
                d0.loc[[baseline_sample_id]] > thresh, d1 > thresh, metric="jaccard"
            ).squeeze(),
            index=d1.index,
        )
    )

    taxon_class[subject_id] = lib.engraftment.classify_taxon_sets(
        d0, baseline_sample_id, donor_sample_id, thresh=thresh, other_donor_samples=other_donor_samples,
    )
    taxon_class_rabund.append(d1.groupby(taxon_class[subject_id], axis="columns").sum())
    taxon_class_count.append((d1 > thresh).groupby(taxon_class[subject_id], axis="columns").sum())

taxon_class = pd.DataFrame(taxon_class).rename_axis(columns='subject_id').stack().rename('taxon_class')
taxon_class_count = pd.concat(taxon_class_count)
taxon_class_count_frac = taxon_class_count.apply(lib.stats.normalize, axis=1)
taxon_class_rabund = pd.concat(taxon_class_rabund)
braycurtis = pd.DataFrame(
    dict(donor=pd.concat(donor_bc), subject=pd.concat(baseline_bc))
)
jaccard = pd.DataFrame(
    dict(donor=pd.concat(donor_jacc), subject=pd.concat(baseline_jacc))
)

# Final variable naming
rotu_taxon_class = taxon_class
rotu_braycurtis = braycurtis
rotu_jaccard = jaccard
rotu_taxon_class_rabund =taxon_class_rabund
rotu_taxon_class_count = taxon_class_count
rotu_taxon_class_count_frac = taxon_class_count_frac
```

```
/var/folders/vv/8wpk7cln4lgb9qyqchjvcsmh0000gq/T/ipykernel_17922/2321219925.py:37: UserWarning: No baseline sample for S0041. Skipping this subject.
  warn(f"No baseline sample for {subject_id}. Skipping this subject.")
/var/folders/vv/8wpk7cln4lgb9qyqchjvcsmh0000gq/T/ipykernel_17922/2321219925.py:37: UserWarning: No baseline sample for S0056. Skipping this subject.
  warn(f"No baseline sample for {subject_id}. Skipping this subject.")
```

In [ ]:

```
(
    sample[sample.sample_type == "followup_1"].join(rotu_taxon_class_count).donor.median(),
    sample[sample.sample_type == "followup_1"].join(rotu_taxon_class_rabund).donor.median(),
)
```

Out[ ]:

```
(113.0, 0.2475699666927062)
```

In [ ]:

```
(
    sample[sample.sample_type == "followup_2"].join(rotu_taxon_class_count).donor.median(),
    sample[sample.sample_type == "followup_2"].join(rotu_taxon_class_rabund).donor.median(),
)
```

Out[ ]:

```
(103.0, 0.2019215934542398)
```

In [ ]:

```
(
    sample[sample.sample_type == "baseline"]
    .join(rotu_taxon_class_count_frac)
    .shared.median(),
    sample[sample.sample_type == "baseline"].join(rotu_taxon_class_rabund).shared.median(),
)
```

Out[ ]:

```
(0.5391304347826087, 0.6900857493828191)
```

In [ ]:

```
_taxon_class_rabund = rotu_taxon_class_rabund
_taxon_class_count_frac = rotu_taxon_class_count_frac
_braycurtis = rotu_braycurtis
_jaccard = rotu_jaccard

_taxon_class_pvalues = []

for score, d0 in dict(
    taxon_class_rabund=_taxon_class_rabund,
    taxon_class_count_frac=_taxon_class_count_frac,
    braycurtis=_braycurtis,
    jaccard=_jaccard,
).items():
    for taxon_class in lib.engraftment.TAXON_CLASS_ORDER:
        if taxon_class not in d0:
            continue
        d1 = (
            d0.join(sample[["subject_id", "sample_type"]])
            .set_index(["subject_id", "sample_type"])[taxon_class]
            .unstack()[lib.project_data.SAMPLE_TYPE_ORDER_SIMPLE]
            .join(subject[["antibiotics_", "maintenance_", "responder", "remission"]])
        )
        for treatment, reference in [
            ("antibiotics_", "ABX-"),
            ("maintenance_", "ENMA"),
            #            ('responder', False),
            ("remission", False),
        ]:
            for sample_type in lib.project_data.SAMPLE_TYPE_ORDER_SIMPLE:
                #                 print(d1.groupby(treatment)[sample_type].median())
                try:
                    mwu = lib.stats.mannwhitneyu(
                        treatment,
                        sample_type,
                        data=d1,
                        reference=reference,
                        alternative="two-sided",
                        na_action="drop",
                    )
                #                     if (score == 'braycurtis') and (taxon_class=='subject') and (treatment=='antibiotics_') and (sample_type=='pre_maintenance_1'):
                #                         print(d2)
                except ValueError:
                    continue
                _taxon_class_pvalues.append(
                    (taxon_class, score, treatment, sample_type, mwu[1])
                )

_taxon_class_pvalues = (
    pd.DataFrame(
        _taxon_class_pvalues,
        columns=["taxon_class", "score", "treatment", "sample_type", "pvalue"],
    )
    .set_index(["treatment", "taxon_class", "score", "sample_type"])
    .squeeze()
)

rotu_taxon_class_pvalues = _taxon_class_pvalues

(
    _taxon_class_pvalues.unstack("sample_type")[
        lib.project_data.SAMPLE_TYPE_ORDER_SIMPLE
    ]
    .stack("sample_type")
    .unstack("treatment")
)
```

Out[ ]:

|  |  | treatment | antibiotics\_ | maintenance\_ | remission |
| --- | --- | --- | --- | --- | --- |
| taxon\_class | score | sample\_type |  |  |  |
| donor | braycurtis | baseline | 0.792208 | 0.792208 | 0.177489 |
|  |  | pre\_maintenance\_1 | 0.150794 | 0.761905 | 0.516667 |
|  |  | pre\_maintenance\_2 | 1.000000 | 0.428571 | 0.230303 |
|  |  | pre\_maintenance\_3 | 0.628205 | 0.294872 | 0.354312 |
|  |  | pre\_maintenance\_4 | 1.000000 | 0.294872 | 0.284382 |
| ... | ... | ... | ... | ... | ... |
| subject | taxon\_class\_rabund | pre\_maintenance\_4 | 0.051948 | 0.792208 | 0.662338 |
|  |  | pre\_maintenance\_5 | 0.125541 | 0.662338 | 0.792208 |
|  |  | pre\_maintenance\_6 | 0.285714 | 0.904762 | 1.000000 |
|  |  | followup\_1 | 0.329004 | 0.051948 | 0.329004 |
|  |  | followup\_2 | 0.662338 | 0.177489 | 0.125541 |

126 rows × 3 columns

##### Species-level analysis¶

In [ ]:

```
d0 = sample_x_motu
thresh = 1e-6  # Thresh used for jaccard presence/absence.

taxon_class = {}
taxon_class_rabund = []
taxon_class_count = []
baseline_bc = []
donor_bc = []
baseline_jacc = []
donor_jacc = []


for subject_id, d1 in d0.groupby(sample.subject_id):
    if not subject_id in recipient_has_mgen_order:
        continue

    donor_sample_id = donor_to_donor_mean[subject.loc[subject_id, "donor_subject_id"]]
    other_donor_samples = list(set(all_donor_mean_samples) - set([donor_sample_id]))
    donor_bc.append(
        pd.Series(
            sp.spatial.distance.cdist(
                d0.loc[[donor_sample_id]], d1, metric="braycurtis"
            ).squeeze(),
            index=d1.index,
        )
    )
    donor_jacc.append(
        pd.Series(
            sp.spatial.distance.cdist(
                d0.loc[[donor_sample_id]] > thresh, d1 > thresh, metric="jaccard"
            ).squeeze(),
            index=d1.index,
        )
    )

    if not subject_to_baseline_sample[subject_id] in d0.index:
        warn(f"No baseline sample for {subject_id}. Skipping this subject.")
        continue
    baseline_sample_id = subject_to_baseline_sample[subject_id]
    baseline_bc.append(
        pd.Series(
            sp.spatial.distance.cdist(
                d0.loc[[baseline_sample_id]], d1, metric="braycurtis"
            ).squeeze(),
            index=d1.index,
        )
    )
    baseline_jacc.append(
        pd.Series(
            sp.spatial.distance.cdist(
                d0.loc[[baseline_sample_id]] > thresh, d1 > thresh, metric="jaccard"
            ).squeeze(),
            index=d1.index,
        )
    )

    taxon_class[subject_id] = lib.engraftment.classify_taxon_sets(
        d0, baseline_sample_id, donor_sample_id, thresh=thresh, other_donor_samples=other_donor_samples,
    )
    taxon_class_rabund.append(d1.groupby(taxon_class[subject_id], axis="columns").sum())
    taxon_class_count.append((d1 > thresh).groupby(taxon_class[subject_id], axis="columns").sum())

taxon_class = pd.DataFrame(taxon_class).rename_axis(columns='subject_id').stack().rename('taxon_class')
taxon_class_count = pd.concat(taxon_class_count)
taxon_class_count_frac = taxon_class_count.apply(lib.stats.normalize, axis=1)
taxon_class_rabund = pd.concat(taxon_class_rabund)
braycurtis = pd.DataFrame(
    dict(donor=pd.concat(donor_bc), subject=pd.concat(baseline_bc))
)
jaccard = pd.DataFrame(
    dict(donor=pd.concat(donor_jacc), subject=pd.concat(baseline_jacc))
)

# Final variable naming
motu_taxon_class = taxon_class
motu_braycurtis = braycurtis
motu_jaccard = jaccard
motu_taxon_class_rabund =taxon_class_rabund
motu_taxon_class_count = taxon_class_count
motu_taxon_class_count_frac = taxon_class_count_frac
```

In [ ]:

```
(
    sample[sample.sample_type == "followup_1"].join(motu_taxon_class_count).donor.median(),
    sample[sample.sample_type == "followup_1"].join(motu_taxon_class_rabund).donor.median(),
)
```

Out[ ]:

```
(77.0, 0.2519716884869385)
```

In [ ]:

```
(
    sample[sample.sample_type == "followup_2"].join(motu_taxon_class_count).donor.median(),
    sample[sample.sample_type == "followup_2"].join(motu_taxon_class_rabund).donor.median(),
)
```

Out[ ]:

```
(56.0, 0.12286527987976861)
```

In [ ]:

```
(
    sample[sample.sample_type == "baseline"]
    .join(motu_taxon_class_count_frac)
    .shared.median(),
    sample[sample.sample_type == "baseline"].join(motu_taxon_class_rabund).shared.median(),
)
```

Out[ ]:

```
(0.5615384615384615, 0.727468798871346)
```

In [ ]:

```
_taxon_class_rabund = motu_taxon_class_rabund
_taxon_class_count_frac = motu_taxon_class_count_frac
_braycurtis = motu_braycurtis
_jaccard = motu_jaccard

_taxon_class_pvalues = []

for score, d0 in dict(
    taxon_class_rabund=_taxon_class_rabund,
    taxon_class_count_frac=_taxon_class_count_frac,
    braycurtis=_braycurtis,
    jaccard=_jaccard,
).items():
    for taxon_class in lib.engraftment.TAXON_CLASS_ORDER:
        if taxon_class not in d0:
            continue
        d1 = (
            d0.join(sample[["subject_id", "sample_type"]])
            .set_index(["subject_id", "sample_type"])[taxon_class]
            .unstack()[lib.project_data.SAMPLE_TYPE_ORDER_SIMPLE]
            .join(subject[["antibiotics_", "maintenance_", "responder", "remission"]])
        )
        for treatment, reference in [
            ("antibiotics_", "ABX-"),
            ("maintenance_", "ENMA"),
            #            ('responder', False),
            ("remission", False),
        ]:
            for sample_type in lib.project_data.SAMPLE_TYPE_ORDER_SIMPLE:
                #                 print(d1.groupby(treatment)[sample_type].median())
                try:
                    mwu = lib.stats.mannwhitneyu(
                        treatment,
                        sample_type,
                        data=d1,
                        reference=reference,
                        alternative="two-sided",
                        na_action="drop",
                    )
                #                     if (score == 'braycurtis') and (taxon_class=='subject') and (treatment=='antibiotics_') and (sample_type=='pre_maintenance_1'):
                #                         print(d2)
                except ValueError:
                    continue
                _taxon_class_pvalues.append(
                    (taxon_class, score, treatment, sample_type, mwu[1])
                )

_taxon_class_pvalues = (
    pd.DataFrame(
        _taxon_class_pvalues,
        columns=["taxon_class", "score", "treatment", "sample_type", "pvalue"],
    )
    .set_index(["treatment", "taxon_class", "score", "sample_type"])
    .squeeze()
)

motu_taxon_class_pvalues = _taxon_class_pvalues

(
    _taxon_class_pvalues.unstack("sample_type")[
        lib.project_data.SAMPLE_TYPE_ORDER_SIMPLE
    ]
    .stack("sample_type")
    .unstack("treatment")
)
```

Out[ ]:

|  |  | treatment | antibiotics\_ | maintenance\_ | remission |
| --- | --- | --- | --- | --- | --- |
| taxon\_class | score | sample\_type |  |  |  |
| donor | braycurtis | baseline | 0.945221 | 0.533800 | 0.284382 |
|  |  | pre\_maintenance\_1 | 0.309524 | 0.171429 | 0.666667 |
|  |  | pre\_maintenance\_2 | 0.536797 | 0.536797 | 0.412121 |
|  |  | pre\_maintenance\_3 | 0.445221 | 0.294872 | 0.354312 |
|  |  | pre\_maintenance\_4 | 0.365967 | 0.445221 | 0.832945 |
| ... | ... | ... | ... | ... | ... |
| subject | taxon\_class\_rabund | pre\_maintenance\_4 | 0.073427 | 0.533800 | 0.523699 |
|  |  | pre\_maintenance\_5 | 0.267677 | 0.876263 | 0.808081 |
|  |  | pre\_maintenance\_6 | 0.315152 | 0.662338 | 0.787879 |
|  |  | followup\_1 | 0.365967 | 0.137529 | 0.832945 |
|  |  | followup\_2 | 0.445221 | 0.294872 | 0.065268 |

126 rows × 3 columns

##### Strain-level analysis¶

In [ ]:

```
d0 = sample_x_sotu
thresh = 1e-6  # Thresh used for jaccard presence/absence.

taxon_class = {}
taxon_class_rabund = []
taxon_class_count = []
baseline_bc = []
donor_bc = []
baseline_jacc = []
donor_jacc = []


for subject_id, d1 in d0.groupby(sample.subject_id):
    if not subject_id in recipient_has_mgen_order:
        continue

    donor_sample_id = donor_to_donor_mean[subject.loc[subject_id, "donor_subject_id"]]
    other_donor_samples = list(set(all_donor_mean_samples) - set([donor_sample_id]))
    donor_bc.append(
        pd.Series(
            sp.spatial.distance.cdist(
                d0.loc[[donor_sample_id]], d1, metric="braycurtis"
            ).squeeze(),
            index=d1.index,
        )
    )
    donor_jacc.append(
        pd.Series(
            sp.spatial.distance.cdist(
                d0.loc[[donor_sample_id]] > thresh, d1 > thresh, metric="jaccard"
            ).squeeze(),
            index=d1.index,
        )
    )

    if not subject_to_baseline_sample[subject_id] in d0.index:
        warn(f"No baseline sample for {subject_id}. Skipping this subject.")
        continue
    baseline_sample_id = subject_to_baseline_sample[subject_id]
    baseline_bc.append(
        pd.Series(
            sp.spatial.distance.cdist(
                d0.loc[[baseline_sample_id]], d1, metric="braycurtis"
            ).squeeze(),
            index=d1.index,
        )
    )
    baseline_jacc.append(
        pd.Series(
            sp.spatial.distance.cdist(
                d0.loc[[baseline_sample_id]] > thresh, d1 > thresh, metric="jaccard"
            ).squeeze(),
            index=d1.index,
        )
    )

    taxon_class[subject_id] = lib.engraftment.classify_taxon_sets(
        d0, baseline_sample_id, donor_sample_id, thresh=thresh, other_donor_samples=other_donor_samples,
    )
    taxon_class_rabund.append(d1.groupby(taxon_class[subject_id], axis="columns").sum())
    taxon_class_count.append((d1 > thresh).groupby(taxon_class[subject_id], axis="columns").sum())

taxon_class = pd.DataFrame(taxon_class).rename_axis(columns='subject_id').stack().rename('taxon_class')
taxon_class_count = pd.concat(taxon_class_count)
taxon_class_count_frac = taxon_class_count.apply(lib.stats.normalize, axis=1)
taxon_class_rabund = pd.concat(taxon_class_rabund)
braycurtis = pd.DataFrame(
    dict(donor=pd.concat(donor_bc), subject=pd.concat(baseline_bc))
)
jaccard = pd.DataFrame(
    dict(donor=pd.concat(donor_jacc), subject=pd.concat(baseline_jacc))
)

# Final variable naming
sotu_taxon_class = taxon_class
sotu_braycurtis = braycurtis
sotu_jaccard = jaccard
sotu_taxon_class_rabund =taxon_class_rabund
sotu_taxon_class_count = taxon_class_count
sotu_taxon_class_count_frac = taxon_class_count_frac
```

In [ ]:

```
taxon_class
```

Out[ ]:

```
sotu_id       subject_id
100002-other  S0001         other_nodonor
              S0004         other_nodonor
              S0007         other_nodonor
              S0008         other_nodonor
              S0013         other_nodonor
                                ...      
104644-s020   S0041         other_nodonor
              S0047         other_nodonor
              S0053         other_nodonor
              S0055         other_nodonor
              S0056         other_nodonor
Name: taxon_class, Length: 127010, dtype: object
```

In [ ]:

```
sotu_taxon_class
```

Out[ ]:

```
sotu_id       subject_id
100002-other  S0001         other_nodonor
              S0004         other_nodonor
              S0007         other_nodonor
              S0008         other_nodonor
              S0013         other_nodonor
                                ...      
104644-s020   S0041         other_nodonor
              S0047         other_nodonor
              S0053         other_nodonor
              S0055         other_nodonor
              S0056         other_nodonor
Name: taxon_class, Length: 127010, dtype: object
```

In [ ]:

```
sotu_taxon_class
```

Out[ ]:

```
sotu_id       subject_id
100002-other  S0001         other_nodonor
              S0004         other_nodonor
              S0007         other_nodonor
              S0008         other_nodonor
              S0013         other_nodonor
                                ...      
104644-s020   S0041         other_nodonor
              S0047         other_nodonor
              S0053         other_nodonor
              S0055         other_nodonor
              S0056         other_nodonor
Name: taxon_class, Length: 127010, dtype: object
```

In [ ]:

```
sample[sample.sample_type == "baseline"].join(sotu_taxon_class_count_frac).shared.median()
```

Out[ ]:

```
0.2398989898989899
```

In [ ]:

```
(
    sample[sample.sample_type == "followup_1"].join(sotu_taxon_class_count).donor.median(),
    sample[sample.sample_type == "followup_1"].join(sotu_taxon_class_rabund).donor.median(),
)
```

Out[ ]:

```
(260.0, 0.5730734793850354)
```

In [ ]:

```
(
    sample[sample.sample_type == "followup_2"].join(sotu_taxon_class_count).donor.median(),
    sample[sample.sample_type == "followup_2"].join(sotu_taxon_class_rabund).donor.median(),
)
```

Out[ ]:

```
(232.0, 0.5035589175479743)
```

In [ ]:

```
(
    sample[sample.sample_type == "baseline"].join(sotu_taxon_class_count).subject.median(),
    sample[sample.sample_type == "baseline"].join(sotu_taxon_class_rabund).subject.median(),
)
```

Out[ ]:

```
(222.0, 0.8758540681347675)
```

In [ ]:

```
(
    sample[sample.sample_type == "followup_1"].join(sotu_taxon_class_count).subject.median(),
    sample[sample.sample_type == "followup_1"]
    .join(sotu_taxon_class_rabund)
    .subject.median(),
)
```

Out[ ]:

```
(54.0, 0.11733366182659051)
```

In [ ]:

```
(
    sample[sample.sample_type == "followup_1"]
    .join(sotu_taxon_class_rabund)
    .subject.median(),
    sample[sample.sample_type == "followup_2"]
    .join(sotu_taxon_class_rabund)
    .subject.median(),
)
```

Out[ ]:

```
(0.11733366182659051, 0.19969051730761916)
```

In [ ]:

```
(
    sample[sample.sample_type == "baseline"].join(sotu_braycurtis).donor.median(),
    sample[sample.sample_type == "followup_1"].join(sotu_braycurtis).donor.median(),
)
```

Out[ ]:

```
(0.9941177512274132, 0.832998039506422)
```

In [ ]:

```
sp.stats.mannwhitneyu(
    sample[sample.sample_type == "baseline"].join(sotu_braycurtis).donor.dropna().values,
    sample[sample.sample_type == "followup_1"].join(sotu_braycurtis).donor.dropna().values,
)
```

Out[ ]:

```
MannwhitneyuResult(statistic=169.0, pvalue=1.6496665061862602e-05)
```

In [ ]:

```
sample[sample.sample_type == "followup_1"].join(sotu_braycurtis).subject.median()
```

Out[ ]:

```
0.9109994212214586
```

In [ ]:

```
wilcoxon_results = []

for sample_type_ref in lib.project_data.SAMPLE_TYPE_ORDER_SIMPLE:
    for score, d0 in dict(
        donor_rabund=sotu_taxon_class_rabund["donor"], donor_braycurtis=sotu_braycurtis["donor"]
    ).items():
        d1 = (
            d0.to_frame(name="score")
            .join(sample[["subject_id", "sample_type"]])
            .set_index(["subject_id", "sample_type"])
            .score.unstack()
        )
        for sample_type in lib.project_data.SAMPLE_TYPE_ORDER_SIMPLE:
            d2 = d1[[sample_type_ref, sample_type]].dropna()
            try:
                wilcoxon_w, wilcoxon_p = sp.stats.wilcoxon(
                    d2[sample_type_ref], d2[sample_type], mode="exact"
                )
            except ValueError:
                wilcoxon_w, wilcoxon_p = np.nan, np.nan
            wilcoxon_results.append(
                (score, sample_type_ref, sample_type, wilcoxon_w, wilcoxon_p)
            )
#             if (sample_type_ref, sample_type) == ('baseline', 'pre_maintenance_1'):
#                 print(d2)

wilcoxon_results = pd.DataFrame(
    wilcoxon_results, columns=["score", "sample_type_ref", "sample_type", "w", "p"]
).set_index(["score", "sample_type_ref", "sample_type"])
```

In [ ]:

```
print(wilcoxon_results.xs("donor_rabund", level="score").p.unstack())
```

```
sample_type        baseline  followup_1  followup_2  pre_maintenance_1  \
sample_type_ref                                                          
baseline                NaN    0.000244    0.000244           0.001953   
followup_1         0.000244         NaN    0.190918           1.000000   
followup_2         0.000244    0.190918         NaN           0.275391   
pre_maintenance_1  0.001953    1.000000    0.275391                NaN   
pre_maintenance_2  0.000977    0.577148    0.206055           0.300781   
pre_maintenance_3  0.000244    0.892578    0.839355           0.845703   
pre_maintenance_4  0.000244    0.414307    0.216309           0.375000   
pre_maintenance_5  0.000488    0.423828    0.518555           0.652344   
pre_maintenance_6  0.000977    0.764648    0.764648           0.460938   

sample_type        pre_maintenance_2  pre_maintenance_3  pre_maintenance_4  \
sample_type_ref                                                              
baseline                    0.000977           0.000244           0.000244   
followup_1                  0.577148           0.892578           0.414307   
followup_2                  0.206055           0.839355           0.216309   
pre_maintenance_1           0.300781           0.845703           0.375000   
pre_maintenance_2                NaN           0.174805           0.965820   
pre_maintenance_3           0.174805                NaN           0.094238   
pre_maintenance_4           0.965820           0.094238                NaN   
pre_maintenance_5           0.556641           0.339355           0.909668   
pre_maintenance_6           1.000000           0.123047           0.965820   

sample_type        pre_maintenance_5  pre_maintenance_6  
sample_type_ref                                          
baseline                    0.000488           0.000977  
followup_1                  0.423828           0.764648  
followup_2                  0.518555           0.764648  
pre_maintenance_1           0.652344           0.460938  
pre_maintenance_2           0.556641           1.000000  
pre_maintenance_3           0.339355           0.123047  
pre_maintenance_4           0.909668           0.965820  
pre_maintenance_5                NaN           0.375000  
pre_maintenance_6           0.375000                NaN
```

In [ ]:

```
print(wilcoxon_results.xs("donor_braycurtis", level="score").p.unstack())
```

```
sample_type        baseline  followup_1  followup_2  pre_maintenance_1  \
sample_type_ref                                                          
baseline                NaN    0.000244    0.000244           0.001953   
followup_1         0.000244         NaN    0.146484           0.492188   
followup_2         0.000244    0.146484         NaN           0.232422   
pre_maintenance_1  0.001953    0.492188    0.232422                NaN   
pre_maintenance_2  0.000977    0.174805    0.041992           0.074219   
pre_maintenance_3  0.000244    0.216309    0.068115           0.193359   
pre_maintenance_4  0.000244    0.127197    0.039795           0.064453   
pre_maintenance_5  0.000488    0.109863    0.077148           0.300781   
pre_maintenance_6  0.000977    0.365234    0.240234           0.382812   

sample_type        pre_maintenance_2  pre_maintenance_3  pre_maintenance_4  \
sample_type_ref                                                              
baseline                    0.000977           0.000244           0.000244   
followup_1                  0.174805           0.216309           0.127197   
followup_2                  0.041992           0.068115           0.039795   
pre_maintenance_1           0.074219           0.193359           0.064453   
pre_maintenance_2                NaN           0.898438           0.577148   
pre_maintenance_3           0.898438                NaN           0.127197   
pre_maintenance_4           0.577148           0.127197                NaN   
pre_maintenance_5           0.625000           1.000000           0.077148   
pre_maintenance_6           0.556641           0.637695           0.764648   

sample_type        pre_maintenance_5  pre_maintenance_6  
sample_type_ref                                          
baseline                    0.000488           0.000977  
followup_1                  0.109863           0.365234  
followup_2                  0.077148           0.240234  
pre_maintenance_1           0.300781           0.382812  
pre_maintenance_2           0.625000           0.556641  
pre_maintenance_3           1.000000           0.637695  
pre_maintenance_4           0.077148           0.764648  
pre_maintenance_5                NaN           1.000000  
pre_maintenance_6           1.000000                NaN
```

In [ ]:

```
_taxon_class_rabund = sotu_taxon_class_rabund
_taxon_class_count_frac = sotu_taxon_class_count_frac
_braycurtis = sotu_braycurtis
_jaccard = sotu_jaccard

_taxon_class_pvalues = []

for score, d0 in dict(
    taxon_class_rabund=_taxon_class_rabund,
    taxon_class_count_frac=_taxon_class_count_frac,
    braycurtis=_braycurtis,
    jaccard=_jaccard,
).items():
    for taxon_class in lib.engraftment.TAXON_CLASS_ORDER:
        if taxon_class not in d0:
            continue
        d1 = (
            d0.join(sample[["subject_id", "sample_type"]])
            .set_index(["subject_id", "sample_type"])[taxon_class]
            .unstack()[lib.project_data.SAMPLE_TYPE_ORDER_SIMPLE]
            .join(subject[["antibiotics_", "maintenance_", "responder", "remission"]])
        )
        for treatment, reference in [
            ("antibiotics_", "ABX-"),
            ("maintenance_", "ENMA"),
            #            ('responder', False),
            ("remission", False),
        ]:
            for sample_type in lib.project_data.SAMPLE_TYPE_ORDER_SIMPLE:
                #                 print(d1.groupby(treatment)[sample_type].median())
                try:
                    mwu = lib.stats.mannwhitneyu(
                        treatment,
                        sample_type,
                        data=d1,
                        reference=reference,
                        alternative="two-sided",
                        na_action="drop",
                    )
                #                     if (score == 'braycurtis') and (taxon_class=='subject') and (treatment=='antibiotics_') and (sample_type=='pre_maintenance_1'):
                #                         print(d2)
                except ValueError:
                    continue
                _taxon_class_pvalues.append(
                    (taxon_class, score, treatment, sample_type, mwu[1])
                )

_taxon_class_pvalues = (
    pd.DataFrame(
        _taxon_class_pvalues,
        columns=["taxon_class", "score", "treatment", "sample_type", "pvalue"],
    )
    .set_index(["treatment", "taxon_class", "score", "sample_type"])
    .squeeze()
)

sotu_taxon_class_pvalues = _taxon_class_pvalues

(
    _taxon_class_pvalues.unstack("sample_type")[
        lib.project_data.SAMPLE_TYPE_ORDER_SIMPLE
    ]
    .stack("sample_type")
    .unstack("treatment")
)
```

Out[ ]:

|  |  | treatment | antibiotics\_ | maintenance\_ | remission |
| --- | --- | --- | --- | --- | --- |
| taxon\_class | score | sample\_type |  |  |  |
| donor | braycurtis | baseline | 0.365967 | 0.365967 | 0.523699 |
|  |  | pre\_maintenance\_1 | 1.000000 | 0.476190 | 0.516667 |
|  |  | pre\_maintenance\_2 | 0.536797 | 0.792208 | 0.527273 |
|  |  | pre\_maintenance\_3 | 0.073427 | 0.445221 | 0.724165 |
|  |  | pre\_maintenance\_4 | 0.051282 | 0.533800 | 0.943279 |
| ... | ... | ... | ... | ... | ... |
| subject | taxon\_class\_rabund | pre\_maintenance\_4 | 0.013986 | 0.835664 | 0.523699 |
|  |  | pre\_maintenance\_5 | 0.017677 | 0.530303 | 0.569697 |
|  |  | pre\_maintenance\_6 | 0.163636 | 0.930736 | 0.527273 |
|  |  | followup\_1 | 0.013986 | 0.730769 | 0.435120 |
|  |  | followup\_2 | 0.365967 | 0.835664 | 0.724165 |

126 rows × 3 columns

In [ ]:

```
fig, axs = plt.subplots(2, 2, figsize=(8, 5), sharex=True, sharey=True)
pvalue_for = "antibiotics_"
colorby = "antibiotics_"
colorby_order = lib.project_style.ANTIBIOTICS_ORDER
colorby_pallete = lib.project_style.DEFAULT_COLOR_PALETTE

for (score, (score_title, d0)), ax_col, panel_col in zip(
    dict(
        taxon_class_rabund=("Relative Abundance", sotu_taxon_class_rabund),
        braycurtis=("Bray-Curtis Similarity", 1 - sotu_braycurtis),
    ).items(),
    axs.T,
    [["A", "C"], ["B", "D"]],
):
    d1 = (
        d0.join(sample)
        .join(subject, on="subject_id")
        .join(lib.project_style.SAMPLE_TYPE_INDEX.rename("x"), on="sample_type")
    )
    ax_col[0].set_title(score_title)
    for taxon_class, ax, panel in zip(
        lib.engraftment.TAXON_CLASS_ORDER, ax_col, panel_col
    ):
        for colorby_feat, d2 in d1.groupby(colorby):
            for subject_id, d3 in d2.groupby("subject_id"):
                if taxon_class not in d3:
                    ax.set_visible(False)
                    continue
                ax.plot(
                    "x",
                    taxon_class,
                    data=d3.sort_values("x"),
                    color=lib.project_style.DEFAULT_COLOR_PALETTE[colorby_feat],
                    label="__nolegend__",
                )
        for sample_type, x in lib.project_style.SAMPLE_TYPE_INDEX.to_dict().items():
            try:
                annotation = lib.project_style.pvalue_to_annotation(
                    sotu_taxon_class_pvalues[(pvalue_for, taxon_class, score, sample_type)]
                )
                if annotation == "∙":
                    y = 1.101
                elif annotation == "**":
                    y = 1.08
                elif annotation == "*":
                    y = 1.08
                elif annotation == "":
                    y = 1.08
                ax.annotate(annotation, xy=(x, y), ha="center", va="center")
            except KeyError:
                continue
        ax.text(
            -0.0, 1.07, panel, fontsize=12, fontweight="heavy", transform=ax.transAxes
        )
    ax.set_xticks(
        lib.project_style.SAMPLE_TYPE_INDEX,
    )
    ax.set_xticklabels(lib.project_style.SAMPLE_TYPE_INDEX.index)
    lib.plot.rotate_xticklabels(ax=ax)
    ax.set_ylim(-0.05, 1.22)

for taxon_class, ax in zip(lib.engraftment.TAXON_CLASS_ORDER, axs[:, 0]):
    ax.set_ylabel(taxon_class)

ax = axs[1, 1]
for colorby_feat in colorby_order:
    ax.plot([], [], color=colorby_pallete[colorby_feat], label=colorby_feat)
ax.legend(bbox_to_anchor=(1, 1))
fig.tight_layout()
# fig.savefig('fig/engraftment_profile_antibiotics.png', dpi=250, bbox_inches='tight')
# fig.savefig('fig/engraftment_profile_antibiotics.pdf', dpi=250, bbox_inches='tight')
```

In [ ]:

```
fig, axs = plt.subplots(2, 2, figsize=(8, 5), sharex=True, sharey=True)
pvalue_for = "maintenance_"
colorby = "maintenance_"
colorby_order = lib.project_style.MAINTENANCE_ORDER
colorby_pallete = lib.project_style.DEFAULT_COLOR_PALETTE

for (score, (score_title, d0)), ax_col in zip(
    dict(
        taxon_class_rabund=("Relative Abundance", sotu_taxon_class_rabund),
        braycurtis=("Bray-Curtis imilarity", 1 - sotu_braycurtis),
    ).items(),
    axs.T,
):
    d1 = (
        d0.join(sample)
        .join(subject, on="subject_id")
        .join(lib.project_style.SAMPLE_TYPE_INDEX.rename("x"), on="sample_type")
    )
    ax_col[0].set_title(score_title)
    for taxon_class, ax in zip(lib.engraftment.TAXON_CLASS_ORDER, ax_col):
        for colorby_feat, d2 in d1.groupby(colorby):
            for subject_id, d3 in d2.groupby("subject_id"):
                if taxon_class not in d3:
                    ax.set_visible(False)
                    continue
                ax.plot(
                    "x",
                    taxon_class,
                    data=d3.sort_values("x"),
                    color=lib.project_style.DEFAULT_COLOR_PALETTE[colorby_feat],
                    label="__nolegend__",
                )
        for sample_type, x in lib.project_style.SAMPLE_TYPE_INDEX.to_dict().items():
            try:
                annotation = lib.project_style.pvalue_to_annotation(
                    sotu_taxon_class_pvalues[(pvalue_for, taxon_class, score, sample_type)]
                )
                if annotation == "∙":
                    y = 1.101
                elif annotation == "**":
                    y = 1.08
                elif annotation == "*":
                    y = 1.08
                elif annotation == "":
                    y = 1.08
                ax.annotate(annotation, xy=(x, y), ha="center", va="center")
            except KeyError:
                continue
    ax.set_xticks(
        lib.project_style.SAMPLE_TYPE_INDEX,
    )
    ax.set_xticklabels(lib.project_style.SAMPLE_TYPE_INDEX.index)
    lib.plot.rotate_xticklabels(ax=ax)
    ax.set_ylim(-0.05, 1.22)

for taxon_class, ax in zip(lib.engraftment.TAXON_CLASS_ORDER, axs[:, 0]):
    ax.set_ylabel(taxon_class)

ax = axs[0, -1]
for colorby_feat in colorby_order:
    ax.plot([], [], color=colorby_pallete[colorby_feat], label=colorby_feat)
ax.legend(bbox_to_anchor=(1, 1))

fig.savefig(
    "fig/engraftment_profile_maintenance_method.pdf", dpi=250, bbox_inches="tight"
)
```

In [ ]:

```
fig, axs = plt.subplots(2, 2, figsize=(8, 6), sharex=True, sharey=True)
pvalue_for = "antibiotics_"
colorby = "arm"
colorby_order = lib.project_style.ARM_ORDER
colorby_pallete = lib.project_style.DEFAULT_COLOR_PALETTE

for (score, (score_title, d0)), ax_col, panel_col in zip(
    dict(
        taxon_class_rabund=("Relative Abundance", sotu_taxon_class_rabund),
        braycurtis=("Bray-Curtis Similarity", 1 - sotu_braycurtis),
    ).items(),
    axs.T,
    [["A", "C"], ["B", "D"]],
):
    d1 = (
        d0.join(sample)
        .join(subject, on="subject_id")
        .join(lib.project_style.SAMPLE_TYPE_INDEX.rename("x"), on="sample_type")
    )
    ax_col[0].set_title(score_title)
    for taxon_class, ax, panel in zip(
        lib.engraftment.TAXON_CLASS_ORDER, ax_col, panel_col
    ):
        for colorby_feat, d2 in d1.groupby(colorby):
            for subject_id, d3 in d2.groupby("subject_id"):
                if taxon_class not in d3:
                    ax.set_visible(False)
                    continue
                ax.plot(
                    "x",
                    taxon_class,
                    data=d3.sort_values("x"),
                    lw=2,
                    color=lib.project_style.DEFAULT_COLOR_PALETTE[colorby_feat],
                    label="__nolegend__",
                    alpha=0.8,
                )
        for sample_type, x in lib.project_style.SAMPLE_TYPE_INDEX.to_dict().items():
            try:
                annotation = lib.project_style.pvalue_to_annotation(
                    sotu_taxon_class_pvalues[(pvalue_for, taxon_class, score, sample_type)]
                )
                if annotation == "∙":
                    y = 1.09
                elif annotation == "**":
                    y = 1.073
                elif annotation == "*":
                    y = 1.073
                elif annotation == "":
                    y = 1.073
                ax.annotate(annotation, xy=(x, y), ha="center", va="center")
            except KeyError:
                continue
        ax.text(
            -0.0, 1.07, panel, fontsize=12, fontweight="heavy", transform=ax.transAxes
        )
    ax.set_xticks(lib.project_style.SAMPLE_TYPE_INDEX)
    ax.set_xticklabels(
        lib.project_style.SAMPLE_TYPE_INDEX.index.to_series().map(
            lib.project_style.SAMPLE_TYPE_LABELS
        )
    )
    #     lib.plot.rotate_xticklabels(ax=ax)
    ax.set_ylim(-0.05, 1.2)
    ax.set_xlim(-0.5, 8.5)
    ax.set_yticks(np.linspace(0, 1, num=6))
    ax.set_xlabel("Time-point")

for taxon_class, ax in zip(lib.engraftment.TAXON_CLASS_ORDER, axs[:, 0]):
    ax.set_ylabel(taxon_class)
fig.tight_layout()
fig.savefig("fig/engraftment_profiles_by_treatment.pdf", bbox_inches="tight")


# Legend
fig, ax = plt.subplots(figsize=(1, 1))
for colorby_feat in colorby_order:
    ax.plot(
        [],
        [],
        marker="o",
        markersize=8,
        markeredgecolor="none",
        lw=2,
        color=colorby_pallete[colorby_feat],
        label=colorby_feat,
    )
ax.legend()
ax.legend(ncol=1)
for spines in ["top", "right", "bottom", "left"]:
    ax.spines[spines].set_visible(False)
ax.set_xticks([])
ax.set_yticks([])
fig.savefig("fig/treatment_legend.pdf", bbox_inches="tight")
```

In [ ]:

```
d = sotu_taxon_class_rabund.assign(subject_any=lambda x: x.shared + x.subject)
m = sample.join(recipient, on="subject_id")
d.join(m).groupby(["sample_type"])[d.columns].median()
```

Out[ ]:

|  | donor | other\_donor | other\_nodonor | shared | subject | subject\_any |
| --- | --- | --- | --- | --- | --- | --- |
| sample\_type |  |  |  |  |  |  |
| baseline | 0.000001 | 0.000001 | 0.000002 | 0.124142 | 0.875854 | 0.999996 |
| followup\_1 | 0.573073 | 0.008348 | 0.107059 | 0.039751 | 0.117334 | 0.231476 |
| followup\_2 | 0.503559 | 0.013374 | 0.138167 | 0.065910 | 0.199691 | 0.289163 |
| followup\_3 | 0.375309 | 0.024521 | 0.089730 | 0.082067 | 0.428373 | 0.510440 |
| post\_antibiotic | 0.016345 | 0.008508 | 0.439794 | 0.014645 | 0.092036 | 0.287428 |
| pre\_maintenance\_1 | 0.736017 | 0.003868 | 0.077342 | 0.059619 | 0.113698 | 0.199914 |
| pre\_maintenance\_2 | 0.634010 | 0.013747 | 0.068618 | 0.044342 | 0.121294 | 0.200998 |
| pre\_maintenance\_3 | 0.659148 | 0.004945 | 0.066568 | 0.064082 | 0.200454 | 0.230168 |
| pre\_maintenance\_4 | 0.698530 | 0.008971 | 0.061878 | 0.064689 | 0.098940 | 0.192335 |
| pre\_maintenance\_5 | 0.657255 | 0.004589 | 0.089448 | 0.037387 | 0.130316 | 0.249832 |
| pre\_maintenance\_6 | 0.563631 | 0.007399 | 0.073038 | 0.050460 | 0.184037 | 0.292069 |

In [ ]:

```
d = sotu_taxon_class_rabund.assign(subject_any=lambda x: x.shared + x.subject)
m = sample.join(recipient, on="subject_id")
d.join(m).groupby(["antibiotics_", "sample_type"])[d.columns].median()
```

Out[ ]:

|  |  | donor | other\_donor | other\_nodonor | shared | subject | subject\_any |
| --- | --- | --- | --- | --- | --- | --- | --- |
| antibiotics\_ | sample\_type |  |  |  |  |  |  |
| ABX+ | baseline | 0.000001 | 8.613767e-07 | 0.000002 | 0.121894 | 0.878103 | 0.999996 |
|  | followup\_1 | 0.584727 | 6.101573e-03 | 0.103991 | 0.047047 | 0.028298 | 0.071290 |
|  | followup\_2 | 0.538584 | 1.046737e-02 | 0.127422 | 0.066486 | 0.179356 | 0.277382 |
|  | post\_antibiotic | 0.016345 | 8.507932e-03 | 0.439794 | 0.014645 | 0.092036 | 0.287428 |
|  | pre\_maintenance\_1 | 0.797612 | 4.636043e-03 | 0.085436 | 0.059743 | 0.030948 | 0.161594 |
|  | pre\_maintenance\_2 | 0.768067 | 2.675684e-03 | 0.015198 | 0.097352 | 0.016321 | 0.141368 |
|  | pre\_maintenance\_3 | 0.831755 | 3.104671e-03 | 0.027245 | 0.066055 | 0.024230 | 0.139969 |
|  | pre\_maintenance\_4 | 0.746950 | 5.601914e-03 | 0.048544 | 0.098094 | 0.015666 | 0.192204 |
|  | pre\_maintenance\_5 | 0.682994 | 3.219126e-03 | 0.048396 | 0.038015 | 0.021901 | 0.058125 |
|  | pre\_maintenance\_6 | 0.688694 | 7.245964e-03 | 0.075167 | 0.093186 | 0.098544 | 0.217582 |
| ABX- | baseline | 0.000001 | 1.180739e-06 | 0.000002 | 0.124142 | 0.875854 | 0.999996 |
|  | followup\_1 | 0.473159 | 8.879006e-03 | 0.107059 | 0.039751 | 0.304288 | 0.383810 |
|  | followup\_2 | 0.409232 | 1.337362e-02 | 0.138167 | 0.019260 | 0.285950 | 0.435131 |
|  | followup\_3 | 0.375309 | 2.452114e-02 | 0.089730 | 0.082067 | 0.428373 | 0.510440 |
|  | pre\_maintenance\_1 | 0.402161 | 3.099753e-03 | 0.069247 | 0.059495 | 0.272270 | 0.294273 |
|  | pre\_maintenance\_2 | 0.580231 | 1.407375e-02 | 0.078773 | 0.036525 | 0.239896 | 0.273050 |
|  | pre\_maintenance\_3 | 0.398771 | 6.816927e-03 | 0.175819 | 0.050291 | 0.395121 | 0.544502 |
|  | pre\_maintenance\_4 | 0.427103 | 1.185157e-02 | 0.074513 | 0.021300 | 0.380733 | 0.503213 |
|  | pre\_maintenance\_5 | 0.314598 | 5.958889e-03 | 0.095579 | 0.023217 | 0.492749 | 0.547414 |
|  | pre\_maintenance\_6 | 0.563631 | 1.010409e-02 | 0.073038 | 0.017455 | 0.309883 | 0.360344 |

In [ ]:

```
fig, axs = plt.subplots(2, 2, figsize=(8, 5), sharex=True, sharey=True)
pvalue_for = "donor_subject_id"
colorby = "donor_subject_id"
colorby_order = donor_order
colorby_pallete = subject_color_palette

for (score, (score_title, d0)), ax_col in zip(
    dict(
        taxon_class_rabund=("Relative Abundance", sotu_taxon_class_rabund),
        braycurtis=("Bray-Curtis Dissimilarity", 1 - sotu_braycurtis),
    ).items(),
    axs.T,
):
    d1 = (
        d0.join(sample)
        .join(subject, on="subject_id")
        .join(lib.project_style.SAMPLE_TYPE_INDEX.rename("x"), on="sample_type")
    )
    ax_col[0].set_title(score_title)
    for taxon_class, ax in zip(lib.engraftment.TAXON_CLASS_ORDER, ax_col):
        for colorby_feat, d2 in d1.groupby(colorby):
            for subject_id, d3 in d2.groupby("subject_id"):
                if taxon_class not in d3:
                    ax.set_visible(False)
                    continue
                ax.plot(
                    "x",
                    taxon_class,
                    data=d3.sort_values("x"),
                    color=colorby_pallete[colorby_feat],
                    label="__nolegend__",
                )
        for sample_type, x in lib.project_style.SAMPLE_TYPE_INDEX.to_dict().items():
            try:
                annotation = lib.project_style.pvalue_to_annotation(
                    sotu_taxon_class_pvalues[(pvalue_for, taxon_class, score, sample_type)]
                )
                if annotation == "∙":
                    y = 1.101
                elif annotation == "**":
                    y = 1.08
                elif annotation == "*":
                    y = 1.08
                elif annotation == "":
                    y = 1.08
                ax.annotate(annotation, xy=(x, y), ha="center", va="center")
            except KeyError:
                continue
    ax.set_xticks(
        lib.project_style.SAMPLE_TYPE_INDEX,
    )
    ax.set_xticklabels(lib.project_style.SAMPLE_TYPE_INDEX.index)
    lib.plot.rotate_xticklabels(ax=ax)
    ax.set_ylim(-0.05, 1.22)

for taxon_class, ax in zip(lib.engraftment.TAXON_CLASS_ORDER, axs[:, 0]):
    ax.set_ylabel(taxon_class)

ax = axs[0, -1]
for colorby_feat in colorby_order:
    ax.plot([], [], color=colorby_pallete[colorby_feat], label=colorby_feat)
ax.legend(bbox_to_anchor=(1, 1))

fig.savefig("fig/engraftment_profile_donor.pdf", dpi=250, bbox_inches="tight")
```

In [ ]:

```
fig, axs = plt.subplots(2, 2, figsize=(8, 5), sharex=True, sharey=True)
pvalue_for = "remission"
colorby = "remission"
colorby_order = [False, True]
colorby_pallete = lib.project_style.DEFAULT_COLOR_PALETTE

for (score, (score_title, d0)), ax_col in zip(
    dict(
        taxon_class_rabund=("Relative Abundance", sotu_taxon_class_rabund),
        braycurtis=("Bray-Curtis Dissimilarity", sotu_braycurtis),
    ).items(),
    axs.T,
):
    d1 = (
        d0.join(sample)
        .join(subject, on="subject_id")
        .join(lib.project_style.SAMPLE_TYPE_INDEX.rename("x"), on="sample_type")
    )
    ax_col[0].set_title(score_title)
    for taxon_class, ax in zip(lib.engraftment.TAXON_CLASS_ORDER, ax_col):
        for colorby_feat, d2 in d1.groupby(colorby):
            for subject_id, d3 in d2.groupby("subject_id"):
                if taxon_class not in d3:
                    ax.set_visible(False)
                    continue
                ax.plot(
                    "x",
                    taxon_class,
                    data=d3.sort_values("x"),
                    color=lib.project_style.DEFAULT_COLOR_PALETTE[colorby_feat],
                    label="__nolegend__",
                )
        for sample_type, x in lib.project_style.SAMPLE_TYPE_INDEX.to_dict().items():
            try:
                annotation = lib.project_style.pvalue_to_annotation(
                    sotu_taxon_class_pvalues[(pvalue_for, taxon_class, score, sample_type)]
                )
                if annotation == "∙":
                    y = 1.101
                elif annotation == "**":
                    y = 1.08
                elif annotation == "*":
                    y = 1.08
                elif annotation == "":
                    y = 1.08
                ax.annotate(annotation, xy=(x, y), ha="center", va="center")
            except KeyError:
                continue
    ax.set_xticks(
        lib.project_style.SAMPLE_TYPE_INDEX,
    )
    ax.set_xticklabels(lib.project_style.SAMPLE_TYPE_INDEX.index)
    lib.plot.rotate_xticklabels(ax=ax)
    ax.set_ylim(-0.05, 1.22)

for taxon_class, ax in zip(lib.engraftment.TAXON_CLASS_ORDER, axs[:, 0]):
    ax.set_ylabel(taxon_class)

ax = axs[0, -1]
for colorby_feat in colorby_order:
    ax.plot([], [], color=colorby_pallete[colorby_feat], label=colorby_feat)
ax.legend(bbox_to_anchor=(1, 1))
```

Out[ ]:

```
<matplotlib.legend.Legend at 0x13521c5e0>
```

##### All levels combined¶

In [ ]:

```
# TODO: Plot all five taxon classes profiles over time-points

pvalue_for = "antibiotics_"
colorby = "arm"
colorby_order = lib.project_style.ARM_ORDER
colorby_pallete = lib.project_style.DEFAULT_COLOR_PALETTE

taxon_classification_levels = [
    (
        'ASV',
        rotu_taxon_class_rabund,
        rotu_taxon_class_pvalues,
    ),
    (
        'Species',
        motu_taxon_class_rabund,
        motu_taxon_class_pvalues,
    ),
    (
        'Strain',
        sotu_taxon_class_rabund,
        sotu_taxon_class_pvalues,
    ),
]

ncol = len(taxon_classification_levels)
nrow = len(lib.engraftment.TAXON_CLASS_ORDER)


fig, axs = plt.subplots(nrow, ncol, figsize=(3*ncol, 2*nrow), sharex=True, sharey=True)
# axs = axs.reshape((nrow, -1))


for (taxon_level_name, d0, _taxon_class_pvalues), ax_col in zip(taxon_classification_levels, axs.T):
    d1 = (
        d0.join(sample)
        .join(subject, on="subject_id")
        .join(lib.project_style.SAMPLE_TYPE_INDEX.rename("x"), on="sample_type")
    )
    ax_col[0].set_title(taxon_level_name)
    for taxon_class, ax in zip(
        lib.engraftment.TAXON_CLASS_ORDER, ax_col,
    ):
        for colorby_feat, d2 in d1.groupby(colorby):
            for subject_id, d3 in d2.groupby("subject_id"):
                # if taxon_class not in d3:
                #     ax.set_visible(False)
                #     continue
                ax.plot(
                    "x",
                    taxon_class,
                    data=d3.sort_values("x"),
                    color=lib.project_style.DEFAULT_COLOR_PALETTE[colorby_feat],
                    label="__nolegend__",
                )
        for sample_type, x in lib.project_style.SAMPLE_TYPE_INDEX.to_dict().items():
            try:
                annotation = lib.project_style.pvalue_to_annotation(
                    _taxon_class_pvalues[(pvalue_for, taxon_class, 'taxon_class_rabund', sample_type)]
                )
                if annotation == "∙":
                    y = 1.101
                elif annotation == "**":
                    y = 1.08
                elif annotation == "*":
                    y = 1.08
                elif annotation == "":
                    y = 1.08
                ax.annotate(annotation, xy=(x, y), ha="center", va="center")
            except KeyError:
                pass

        ax.set_xticks(lib.project_style.SAMPLE_TYPE_INDEX)
        ax.set_xticklabels(
            lib.project_style.SAMPLE_TYPE_INDEX.index.to_series().map(
                lib.project_style.SAMPLE_TYPE_LABELS
            )
        )

for taxon_class, ax_row in zip(lib.engraftment.TAXON_CLASS_ORDER, axs):
    ax_row[0].set_ylabel(taxon_class)
    ax_row[0].set_ylim(-0.05, 1.2)


# for taxon_class, ax in zip(lib.engraftment.TAXON_CLASS_ORDER, axs[:, 0]):
#     ax.set_ylabel(taxon_class)

ax = axs[2, 2]
for colorby_feat in colorby_order:
    ax.plot([], [], color=colorby_pallete[colorby_feat], label=colorby_feat)
ax.legend()

fig.tight_layout()
# fig.savefig('fig/engraftment_profile_antibiotics.png', dpi=250, bbox_inches='tight')
fig.savefig('doc/static/engraftment_extended_figure.svg', dpi=250, bbox_inches='tight')
```

In [ ]:

```
d0 = sotu_taxon_class_rabund.assign(other=lambda x: x.other_donor + x.other_nodonor)
d1 = (
    d0.join(sample)
    .join(subject, on="subject_id")
    .join(lib.project_style.SAMPLE_TYPE_INDEX.rename("x"), on="sample_type")
)
d2 = d1.loc[lambda x: x.sample_type == 'followup_1'].set_index('subject_id')['other']
print(d2.sort_values())
print(d2.median())
print(d2.quantile([0.9]))
```

```
subject_id
S0004    0.048893
S0013    0.090213
S0021    0.102436
S0007    0.115407
S0027    0.116531
S0055    0.124251
S0041    0.140742
S0001    0.143032
S0024    0.172143
S0056    0.185607
S0008    0.360152
S0047    0.612340
S0053    0.705289
Name: other, dtype: float64
0.14074218694141724
0.9    0.561902
Name: other, dtype: float64
```

In [ ]:

```
d0 = sotu_taxon_class_rabund
d1 = (
    d0.join(sample)
    .join(subject, on="subject_id")
    .join(lib.project_style.SAMPLE_TYPE_INDEX.rename("x"), on="sample_type")
)
d2 = d1.loc[lambda x: x.sample_type == 'followup_1'].set_index('subject_id')['other_donor']
print(d2.sort_values())
print(d2.median())
```

```
subject_id
S0004    0.001648
S0027    0.002909
S0056    0.003478
S0053    0.004127
S0013    0.005133
S0021    0.008076
S0007    0.008348
S0008    0.008879
S0055    0.024112
S0001    0.027512
S0041    0.042944
S0024    0.110069
S0047    0.348534
Name: other_donor, dtype: float64
0.008348017125288307
```

##### Assess potential false positive rate of assigning strain "transfer" events¶

In [ ]:

```
# Let's build venn-diagrams of strains across the three donors' mean samples
from matplotlib_venn import venn3

thresh = 1e-4

fig, axs = plt.subplots(1, 3, figsize=(8, 4))
span = 25

venn_list = []
for (name, d), ax in zip(
    {
        "ASV (16S)": sample_x_rotu,
        "Species": sample_x_motu,
        "Strain": sample_x_sotu,
    }.items(),
    axs,
):
    d0 = (d.loc[donor_means_list] > thresh).T
    d0 = d0[d0.sum(1) >= 1]
    d0 = d0.rename(columns=lambda s: s[:-5])

    if name == "ASV (16S)":
        set_labels = d0.columns
    else:
        set_labels = None
    v = venn3(
        [set(idxwhere(d0[col])) for col in d0],
        set_labels=set_labels,
        set_colors=[subject_color_palette[col] for col in d0],
        normalize_to=d0.sum().sum(),
        ax=ax,
    )
    ax.set_title(name, fontsize=12)
    ax.set_ylim(-span, span)
    ax.set_xlim(-span, span)
    venn_list.append(v)

for v in venn_list[1:]:
    v.set_labels = None

fig.tight_layout(pad=0.98)
fig.savefig("fig/taxa_venn_diagram_donors.pdf", bbox_inches="tight")
```

In [ ]:

```
sample_x_sotu.groupby([sample.subject_id, sample.sample_type]).apply(len).loc[
    ["D0044", "D0097", "D0485"]
]
```

Out[ ]:

```
subject_id  sample_type  
D0044       donor_enema       5
            donor_initial     5
            donor_mean        1
D0097       donor_enema      25
            donor_initial     6
            donor_mean        1
D0485       donor_initial     1
            donor_mean        1
dtype: int64
```

##### Strain transfer differs over phyla¶

In [ ]:

```
d0 = sample_x_sotu
thresh = 1e-4
baseline_sample_type = "baseline"

strain_transfer = {}
for subject_id in recipient_has_mgen_order:
    donor_subject_id = subject.loc[
        subject_id
    ].donor_subject_id  # TODO: Use incorrect donor matching to check FPs
    donor_sample_id = donor_subject_id + "_mean"

    baseline_sample_id = idxwhere(
        (sample.sample_class == baseline_sample_type)
        & (sample.subject_id == subject_id)
    )
    assert len(baseline_sample_id) == 1
    if not baseline_sample_id[0] in d0.index:
        warn(
            f"No {baseline_sample_type} sample for {subject_id}. Skipping this subject."
        )
        continue
    # TODO; if subject_id == 'S0047': skip because this person received feces from a diff donor.
    baseline_sample_id = baseline_sample_id[0]

    taxon_class = lib.engraftment.classify_taxon_sets(
        d0, baseline_sample_id, donor_sample_id, thresh=thresh
    )

    for followup_sample_type in lib.project_data.SAMPLE_TYPE_ORDER_SIMPLE[1:]:
        followup_sample_id = idxwhere(
            (sample.sample_type == followup_sample_type)
            & (sample.subject_id == subject_id)
        )
        if not (followup_sample_id and (followup_sample_id[0] in d0.index)):
            warn(
                f"No {followup_sample_type} sample for {subject_id}. Skipping this sample."
            )
            continue
        #        assert (len(followup_sample_id) == 1)
        followup_sample_id = followup_sample_id[0]
        strain_transfer[(followup_sample_type, subject_id)] = (
            d0.loc[followup_sample_id, taxon_class == "donor"] > thresh
        ).astype(float)

subject_x_strain_transfer = pd.DataFrame(strain_transfer).rename_axis(
    columns=("sample_type", "subject_id")
)
```

```
/var/folders/vv/8wpk7cln4lgb9qyqchjvcsmh0000gq/T/ipykernel_17922/490653557.py:35: UserWarning: No pre_maintenance_2 sample for S0047. Skipping this sample.
  warn(
/var/folders/vv/8wpk7cln4lgb9qyqchjvcsmh0000gq/T/ipykernel_17922/490653557.py:35: UserWarning: No pre_maintenance_6 sample for S0047. Skipping this sample.
  warn(
/var/folders/vv/8wpk7cln4lgb9qyqchjvcsmh0000gq/T/ipykernel_17922/490653557.py:35: UserWarning: No pre_maintenance_5 sample for S0004. Skipping this sample.
  warn(
/var/folders/vv/8wpk7cln4lgb9qyqchjvcsmh0000gq/T/ipykernel_17922/490653557.py:35: UserWarning: No pre_maintenance_6 sample for S0021. Skipping this sample.
  warn(
/var/folders/vv/8wpk7cln4lgb9qyqchjvcsmh0000gq/T/ipykernel_17922/490653557.py:35: UserWarning: No pre_maintenance_1 sample for S0027. Skipping this sample.
  warn(
/var/folders/vv/8wpk7cln4lgb9qyqchjvcsmh0000gq/T/ipykernel_17922/490653557.py:35: UserWarning: No pre_maintenance_1 sample for S0007. Skipping this sample.
  warn(
/var/folders/vv/8wpk7cln4lgb9qyqchjvcsmh0000gq/T/ipykernel_17922/490653557.py:35: UserWarning: No pre_maintenance_2 sample for S0007. Skipping this sample.
  warn(
/var/folders/vv/8wpk7cln4lgb9qyqchjvcsmh0000gq/T/ipykernel_17922/490653557.py:35: UserWarning: No pre_maintenance_1 sample for S0008. Skipping this sample.
  warn(
```

In [ ]:

```
taxon_level = "p__"
sample_type = "followup_1"

opportunities = (
    (subject_x_strain_transfer.notna())
    .groupby(sotu_to_taxonomy[taxon_level])
    .sum()
    .groupby(level="sample_type", axis="columns")
    .sum()
)
transfers = (
    (subject_x_strain_transfer)
    .groupby(sotu_to_taxonomy[taxon_level])
    .sum()
    .groupby(level="sample_type", axis="columns")
    .sum()
)

d = (
    opportunities[sample_type]
    .to_frame(name="opportunities")
    .assign(transfers=transfers[sample_type])
    .assign(transfer_rate=lambda x: x.transfers / x.opportunities)
    .sort_values("transfer_rate", ascending=False)
)
d
```

Out[ ]:

|  | opportunities | transfers | transfer\_rate |
| --- | --- | --- | --- |
| p\_\_ |  |  |  |
| d\_\_Bacteria;p\_\_Desulfobacterota; | 13 | 8.0 | 0.615385 |
| d\_\_Archaea;p\_\_Euryarchaeota; | 14 | 7.0 | 0.500000 |
| d\_\_Bacteria;p\_\_Firmicutes\_C; | 30 | 13.0 | 0.433333 |
| d\_\_Bacteria;p\_\_Bacteroidota; | 1062 | 362.0 | 0.340866 |
| d\_\_Bacteria;p\_\_Firmicutes; | 219 | 71.0 | 0.324201 |
| d\_\_Bacteria;p\_\_Actinobacteriota; | 339 | 98.0 | 0.289086 |
| d\_\_Bacteria;p\_\_Firmicutes\_A; | 4305 | 1138.0 | 0.264344 |
| d\_\_Bacteria;p\_\_Proteobacteria; | 110 | 29.0 | 0.263636 |
| d\_\_Bacteria;p\_\_Verrucomicrobiota; | 35 | 1.0 | 0.028571 |

In [ ]:

```
taxon_level = "p__"
sample_type = "followup_1"

opportunities = (
    (subject_x_strain_transfer.notna())
    .groupby(sotu_to_taxonomy[taxon_level])
    .sum()
    .groupby(level="sample_type", axis="columns")
    .sum()
)
transfers = (
    (subject_x_strain_transfer)
    .groupby(sotu_to_taxonomy[taxon_level])
    .sum()
    .groupby(level="sample_type", axis="columns")
    .sum()
)

d = (
    opportunities[sample_type]
    .to_frame(name="opportunities")
    .assign(transfers=transfers[sample_type])
    .assign(transfer_rate=lambda x: x.transfers / x.opportunities)
    .sort_values("transfer_rate", ascending=False)
)
d[d.index.str.startswith("d__Bacteria;p__Verrucomicrobiota;")].T
```

Out[ ]:

| p\_\_ | d\_\_Bacteria;p\_\_Verrucomicrobiota; |
| --- | --- |
| opportunities | 35.000000 |
| transfers | 1.000000 |
| transfer\_rate | 0.028571 |

In [ ]:

```
from scipy.stats import fisher_exact

table = []
for t1, t2 in product(idxwhere(d.opportunities > 20), repeat=2):
    table.append(
        (t1, t2, fisher_exact(d.loc[[t1, t2], ["opportunities", "transfers"]])[1])
    )

print(
    pd.DataFrame(table, columns=["t1", "t2", "pvalue"])
    .set_index(["t1", "t2"])
    .pvalue.unstack()
)
```

```
t2                                 d__Bacteria;p__Actinobacteriota;  \
t1                                                                    
d__Bacteria;p__Actinobacteriota;                           1.000000   
d__Bacteria;p__Bacteroidota;                               0.228376   
d__Bacteria;p__Firmicutes;                                 0.531234   
d__Bacteria;p__Firmicutes_A;                               0.464049   
d__Bacteria;p__Firmicutes_C;                               0.257248   
d__Bacteria;p__Proteobacteria;                             0.726380   
d__Bacteria;p__Verrucomicrobiota;                          0.002486   

t2                                 d__Bacteria;p__Bacteroidota;  \
t1                                                                
d__Bacteria;p__Actinobacteriota;                       0.228376   
d__Bacteria;p__Bacteroidota;                           1.000000   
d__Bacteria;p__Firmicutes;                             0.767225   
d__Bacteria;p__Firmicutes_A;                           0.000312   
d__Bacteria;p__Firmicutes_C;                           0.479715   
d__Bacteria;p__Proteobacteria;                         0.259920   
d__Bacteria;p__Verrucomicrobiota;                      0.000640   

t2                                 d__Bacteria;p__Firmicutes;  \
t1                                                              
d__Bacteria;p__Actinobacteriota;                     0.531234   
d__Bacteria;p__Bacteroidota;                         0.767225   
d__Bacteria;p__Firmicutes;                           1.000000   
d__Bacteria;p__Firmicutes_A;                         0.160105   
d__Bacteria;p__Firmicutes_C;                         0.452547   
d__Bacteria;p__Proteobacteria;                       0.464641   
d__Bacteria;p__Verrucomicrobiota;                    0.001218   

t2                                 d__Bacteria;p__Firmicutes_A;  \
t1                                                                
d__Bacteria;p__Actinobacteriota;                       0.464049   
d__Bacteria;p__Bacteroidota;                           0.000312   
d__Bacteria;p__Firmicutes;                             0.160105   
d__Bacteria;p__Firmicutes_A;                           1.000000   
d__Bacteria;p__Firmicutes_C;                           0.135089   
d__Bacteria;p__Proteobacteria;                         1.000000   
d__Bacteria;p__Verrucomicrobiota;                      0.003470   

t2                                 d__Bacteria;p__Firmicutes_C;  \
t1                                                                
d__Bacteria;p__Actinobacteriota;                       0.257248   
d__Bacteria;p__Bacteroidota;                           0.479715   
d__Bacteria;p__Firmicutes;                             0.452547   
d__Bacteria;p__Firmicutes_A;                           0.135089   
d__Bacteria;p__Firmicutes_C;                           1.000000   
d__Bacteria;p__Proteobacteria;                         0.217802   
d__Bacteria;p__Verrucomicrobiota;                      0.002113   

t2                                 d__Bacteria;p__Proteobacteria;  \
t1                                                                  
d__Bacteria;p__Actinobacteriota;                         0.726380   
d__Bacteria;p__Bacteroidota;                             0.259920   
d__Bacteria;p__Firmicutes;                               0.464641   
d__Bacteria;p__Firmicutes_A;                             1.000000   
d__Bacteria;p__Firmicutes_C;                             0.217802   
d__Bacteria;p__Proteobacteria;                           1.000000   
d__Bacteria;p__Verrucomicrobiota;                        0.011100   

t2                                 d__Bacteria;p__Verrucomicrobiota;  
t1                                                                    
d__Bacteria;p__Actinobacteriota;                            0.002486  
d__Bacteria;p__Bacteroidota;                                0.000640  
d__Bacteria;p__Firmicutes;                                  0.001218  
d__Bacteria;p__Firmicutes_A;                                0.003470  
d__Bacteria;p__Firmicutes_C;                                0.002113  
d__Bacteria;p__Proteobacteria;                              0.011100  
d__Bacteria;p__Verrucomicrobiota;                           1.000000
```

##### Strain co-existence within species¶

In [ ]:

```
thresh = 1e-6


sample_x_motu_x_taxon_class = (
    pd.merge(
        sample[['subject_id']],
        sotu_taxon_class.unstack('subject_id').T,
        left_on='subject_id',
        right_index=True
    )
    .drop(columns='subject_id')
    .rename_axis(columns='sotu_id')
    .stack()
    .to_frame(name='taxon_class')
    .join(sample_x_sotu.stack().rename('rabund'))
    .dropna()
    [lambda x: x.rabund > thresh]
    .join(sotu_to_taxonomy.motu_id)
    .set_index(['motu_id', 'taxon_class'], append=True)
    .groupby(level=['sample_id', 'motu_id', 'taxon_class'])
    .apply(len)
    # .unstack()
)

assert sample_x_motu_x_taxon_class.index.is_unique
```

In [ ]:

```
donor_recipient_coexist = (
    sample_x_motu_x_taxon_class
    .unstack(fill_value=0)
    .assign(donor_recipient_coexist=lambda x: (x.donor>0) & (x.subject>0))
    .donor_recipient_coexist
    .astype(int)
    .unstack('motu_id')
    .reindex(idxwhere(sample.sample_type == 'followup_1'))
    .rename(sample.subject_id)
    .reindex(recipient_has_mgen_data_list)
    .fillna(0)
    .assign(arm=subject['arm'], donor_subject_id=subject['donor_subject_id'])
    .sort_values('arm')
    .set_index(['arm', 'donor_subject_id'], append=True)
    .T
    .assign(g__=motu_to_taxonomy['g__'])
    .sort_values('g__')
    .set_index(['g__'], append=True)
    .T
)

fig, ax = plt.subplots(figsize=(15, 5))
ax = sns.heatmap(donor_recipient_coexist.loc[:, donor_recipient_coexist.sum() > 0], xticklabels=1, yticklabels=1, ax=ax, cbar=False)#, figsize=(20, 15))
ax.set_xticklabels(ax.get_xmajorticklabels(), fontdict=dict(rotation=-90, ha='center'))

fig.savefig('doc/static/strain_coexistence_figure.svg', bbox_inches='tight')
```

#### Subsection: Antibiotics and donors may modulate effects of FMT on the microbiome¶

##### GEE on donor/subject similarity profiles¶

In [ ]:

```
# TODO: Figure out how to use weeks_since_intiial_fmt as a term in the covariance specification but NOT in the model.
```

###### Load/Shape Data¶

In [ ]:

```
# TODO: Create dataframe with either (bc-dissilarity to donor) or (fraction donor strains)
# across time for each subject.

import statsmodels.formula.api as smf
import statsmodels.genmod.cov_struct as cov_struct
import statsmodels.genmod.families as families


engraftment_gee_data = (
    sample.assign(
        donor_bc=sotu_braycurtis["donor"],
        baseline_bc=sotu_braycurtis["subject"],
        subject_rabund=sotu_taxon_class_rabund["subject"],
        donor_rabund=sotu_taxon_class_rabund["donor"],
        other_rabund=sotu_taxon_class_rabund["other_nodonor"] + sotu_taxon_class_rabund["other_donor"],
    )
    .join(subject, on="subject_id")[
        lambda x: x.sample_type.isin(lib.project_data.SAMPLE_TYPE_ORDER_SIMPLE)
        & ~x.sample_type.isin(["baseline", "post_antibiotic"])
    ]
    .assign(
        remission=lambda x: x.remission.astype(float),
        responder=lambda x: x.responder.astype(float),
        mayo_endo_improved=lambda x: x.mayo_endo_improved.astype(float),
    )
    .drop(columns=["treatment_abx_pre", "withdrawal_due_to_failure"])
    .sort_values(["subject_id", "days_post_fmt"])
    .assign(weeks_post_fmt=lambda x: x.days_post_fmt / 7)
)
```

In [ ]:

```
%%R -i engraftment_gee_data

library("geepack")

variables = c("weeks_post_fmt", "antibiotics_", "donor_subject_id", "maintenance_", "responder")
engraftment_gee_data$subject_id <- as.factor(engraftment_gee_data$subject_id)

engraftment_gee_data_dr <- engraftment_gee_data[complete.cases(engraftment_gee_data[c('donor_rabund', variables)]),]
engraftment_gee_data_sr <- engraftment_gee_data[complete.cases(engraftment_gee_data[c('subject_rabund', variables)]),]
engraftment_gee_data_db <- engraftment_gee_data[complete.cases(engraftment_gee_data[c('donor_bc', variables)]),]
engraftment_gee_data_sb <- engraftment_gee_data[complete.cases(engraftment_gee_data[c('baseline_bc', variables)]),]
engraftment_gee_data_or <- engraftment_gee_data[complete.cases(engraftment_gee_data[c('other_rabund', variables)]),]
```

###### Null Model¶

In [ ]:

```
%%R

# WARNING: d$subject_id (or whatever "id" is, must be sorted)

lm0_dr <- geeglm(
    donor_rabund ~ weeks_post_fmt,
    id=subject_id,
    data=engraftment_gee_data_dr,
    corstr="ar1",
    waves=engraftment_gee_data_dr$weeks_post_fmt,
)
print(summary(lm0_dr))
print(confint.geeglm(lm0_dr))
print('--------')

lm0_sr <- geeglm(
    subject_rabund ~ weeks_post_fmt,
    id=subject_id,
    data=engraftment_gee_data_sr,
    corstr="ar1",
    waves=engraftment_gee_data_dr$weeks_post_fmt
)
print(summary(lm0_sr))
print(confint.geeglm(lm0_sr))
print('--------')

lm0_db <- geeglm(
    donor_bc ~ weeks_post_fmt,
    id=subject_id,
    data=engraftment_gee_data_db,
    corstr="ar1",
    waves=engraftment_gee_data_dr$weeks_post_fmt
)
print(summary(lm0_db))
print(confint.geeglm(lm0_db))
print('--------')

lm0_sb <- geeglm(
    baseline_bc ~ weeks_post_fmt,
    id=subject_id,
    data=engraftment_gee_data_sb,
    corstr="ar1",
    waves=engraftment_gee_data_dr$weeks_post_fmt
)
print(summary(lm0_sb))
print(confint.geeglm(lm0_sb))
print('--------')

lm0_or <- geeglm(
    other_rabund ~ weeks_post_fmt,
    id=subject_id,
    data=engraftment_gee_data_sb,
    corstr="ar1",
    waves=engraftment_gee_data_dr$weeks_post_fmt
)
print(summary(lm0_sb))
print(confint.geeglm(lm0_sb))
print('--------')
```

```
Call:
geeglm(formula = donor_rabund ~ weeks_post_fmt, data = engraftment_gee_data_dr, 
    id = subject_id, waves = engraftment_gee_data_dr$weeks_post_fmt, 
    corstr = "ar1")

 Coefficients:
               Estimate  Std.err  Wald Pr(>|W|)    
(Intercept)     0.59633  0.07485 63.47  1.7e-15 ***
weeks_post_fmt -0.00715  0.00622  1.32     0.25    
---
Signif. codes:  0 ‘***’ 0.001 ‘**’ 0.01 ‘*’ 0.05 ‘.’ 0.1 ‘ ’ 1

Correlation structure = ar1 
Estimated Scale Parameters:

            Estimate Std.err
(Intercept)   0.0665  0.0143
  Link = identity 

Estimated Correlation Parameters:
      Estimate Std.err
alpha    0.973  0.0124
Number of clusters:   13  Maximum cluster size: 8 
                   lwr     upr
(Intercept)     0.4496 0.74304
weeks_post_fmt -0.0193 0.00505
[1] "--------"

Call:
geeglm(formula = subject_rabund ~ weeks_post_fmt, data = engraftment_gee_data_sr, 
    id = subject_id, waves = engraftment_gee_data_dr$weeks_post_fmt, 
    corstr = "ar1")

 Coefficients:
               Estimate Std.err  Wald Pr(>|W|)    
(Intercept)     0.20156 0.05337 14.26  0.00016 ***
weeks_post_fmt  0.00151 0.00363  0.17  0.67809    
---
Signif. codes:  0 ‘***’ 0.001 ‘**’ 0.01 ‘*’ 0.05 ‘.’ 0.1 ‘ ’ 1

Correlation structure = ar1 
Estimated Scale Parameters:

            Estimate Std.err
(Intercept)   0.0362 0.00747
  Link = identity 

Estimated Correlation Parameters:
      Estimate Std.err
alpha    0.983 0.00462
Number of clusters:   13  Maximum cluster size: 8 
                    lwr     upr
(Intercept)     0.09696 0.30616
weeks_post_fmt -0.00561 0.00862
[1] "--------"

Call:
geeglm(formula = donor_bc ~ weeks_post_fmt, data = engraftment_gee_data_db, 
    id = subject_id, waves = engraftment_gee_data_dr$weeks_post_fmt, 
    corstr = "ar1")

 Coefficients:
               Estimate Std.err   Wald Pr(>|W|)    
(Intercept)     0.69138 0.04637 222.30   <2e-16 ***
weeks_post_fmt  0.01704 0.00573   8.86   0.0029 ** 
---
Signif. codes:  0 ‘***’ 0.001 ‘**’ 0.01 ‘*’ 0.05 ‘.’ 0.1 ‘ ’ 1

Correlation structure = ar1 
Estimated Scale Parameters:

            Estimate Std.err
(Intercept)   0.0121 0.00309
  Link = identity 

Estimated Correlation Parameters:
      Estimate Std.err
alpha     1.48  0.0195
Number of clusters:   13  Maximum cluster size: 8 
                   lwr    upr
(Intercept)    0.60050 0.7823
weeks_post_fmt 0.00582 0.0283
[1] "--------"

Call:
geeglm(formula = baseline_bc ~ weeks_post_fmt, data = engraftment_gee_data_sb, 
    id = subject_id, waves = engraftment_gee_data_dr$weeks_post_fmt, 
    corstr = "ar1")

 Coefficients:
               Estimate Std.err  Wald Pr(>|W|)    
(Intercept)     0.84920 0.04191 410.6   <2e-16 ***
weeks_post_fmt  0.00188 0.00424   0.2     0.66    
---
Signif. codes:  0 ‘***’ 0.001 ‘**’ 0.01 ‘*’ 0.05 ‘.’ 0.1 ‘ ’ 1

Correlation structure = ar1 
Estimated Scale Parameters:

            Estimate Std.err
(Intercept)   0.0174 0.00452
  Link = identity 

Estimated Correlation Parameters:
      Estimate Std.err
alpha     1.31  0.0141
Number of clusters:   13  Maximum cluster size: 8 
                    lwr    upr
(Intercept)     0.76707 0.9313
weeks_post_fmt -0.00642 0.0102
[1] "--------"

Call:
geeglm(formula = baseline_bc ~ weeks_post_fmt, data = engraftment_gee_data_sb, 
    id = subject_id, waves = engraftment_gee_data_dr$weeks_post_fmt, 
    corstr = "ar1")

 Coefficients:
               Estimate Std.err  Wald Pr(>|W|)    
(Intercept)     0.84920 0.04191 410.6   <2e-16 ***
weeks_post_fmt  0.00188 0.00424   0.2     0.66    
---
Signif. codes:  0 ‘***’ 0.001 ‘**’ 0.01 ‘*’ 0.05 ‘.’ 0.1 ‘ ’ 1

Correlation structure = ar1 
Estimated Scale Parameters:

            Estimate Std.err
(Intercept)   0.0174 0.00452
  Link = identity 

Estimated Correlation Parameters:
      Estimate Std.err
alpha     1.31  0.0141
Number of clusters:   13  Maximum cluster size: 8 
                    lwr    upr
(Intercept)     0.76707 0.9313
weeks_post_fmt -0.00642 0.0102
[1] "--------"
```

###### Antibiotics¶

In [ ]:

```
%%R

lm_abx_dr <- geeglm(
    donor_rabund ~ weeks_post_fmt + antibiotics_,
    id=subject_id,
    data=engraftment_gee_data_dr,
    corstr="ar1",
    waves=engraftment_gee_data_dr$weeks_post_fmt,
)
print(summary(lm_abx_dr))
print(confint.geeglm(lm_abx_dr))
print(anova(lm0_dr, lm_abx_dr))
print('--------')

lm_abx_sr <- geeglm(
    subject_rabund ~ weeks_post_fmt + antibiotics_,
    id=subject_id,
    data=engraftment_gee_data_sr,
    corstr="ar1",
    waves=engraftment_gee_data_sr$weeks_post_fmt
)
print(summary(lm_abx_sr))
print(confint.geeglm(lm_abx_sr))
print(anova(lm0_sr, lm_abx_sr))
print('--------')

lm_abx_db <- geeglm(
    donor_bc ~ weeks_post_fmt + antibiotics_,
    id=subject_id,
    data=engraftment_gee_data_db,
    corstr="ar1",
    waves=engraftment_gee_data_db$weeks_post_fmt
)
print(summary(lm_abx_db))
print(confint.geeglm(lm_abx_db))
print(anova(lm0_db, lm_abx_db))
print('--------')

lm_abx_sb <- geeglm(
    baseline_bc ~ weeks_post_fmt + antibiotics_,
    id=subject_id,
    data=engraftment_gee_data_sb,
    corstr="ar1",
    waves=engraftment_gee_data_db$weeks_post_fmt
)
print(summary(lm_abx_sb))
print(confint.geeglm(lm_abx_sb))
print(anova(lm0_sb, lm_abx_sb))
print('--------')


lm_abx_or <- geeglm(
    other_rabund ~ weeks_post_fmt + antibiotics_,
    id=subject_id,
    data=engraftment_gee_data_sb,
    corstr="ar1",
    waves=engraftment_gee_data_db$weeks_post_fmt
)
print(summary(lm_abx_or))
print(confint.geeglm(lm_abx_or))
print(anova(lm0_or, lm_abx_or))
print('--------')
```

```
Call:
geeglm(formula = donor_rabund ~ weeks_post_fmt + antibiotics_, 
    data = engraftment_gee_data_dr, id = subject_id, waves = engraftment_gee_data_dr$weeks_post_fmt, 
    corstr = "ar1")

 Coefficients:
                 Estimate Std.err  Wald Pr(>|W|)    
(Intercept)        0.6801  0.1382 24.20  8.7e-07 ***
weeks_post_fmt    -0.0434  0.0183  5.59    0.018 *  
antibiotics_ABX+   0.2632  0.1077  5.98    0.015 *  
---
Signif. codes:  0 ‘***’ 0.001 ‘**’ 0.01 ‘*’ 0.05 ‘.’ 0.1 ‘ ’ 1

Correlation structure = ar1 
Estimated Scale Parameters:

            Estimate Std.err
(Intercept)   0.0718  0.0199
  Link = identity 

Estimated Correlation Parameters:
      Estimate Std.err
alpha      1.2  0.0111
Number of clusters:   13  Maximum cluster size: 8 
                     lwr      upr
(Intercept)       0.4091  0.95101
weeks_post_fmt   -0.0793 -0.00743
antibiotics_ABX+  0.0522  0.47423
Analysis of 'Wald statistic' Table

Model 1 donor_rabund ~ weeks_post_fmt + antibiotics_ 
Model 2 donor_rabund ~ weeks_post_fmt
  Df   X2 P(>|Chi|)  
1  1 5.98     0.015 *
---
Signif. codes:  0 ‘***’ 0.001 ‘**’ 0.01 ‘*’ 0.05 ‘.’ 0.1 ‘ ’ 1
[1] "--------"

Call:
geeglm(formula = subject_rabund ~ weeks_post_fmt + antibiotics_, 
    data = engraftment_gee_data_sr, id = subject_id, waves = engraftment_gee_data_sr$weeks_post_fmt, 
    corstr = "ar1")

 Coefficients:
                  Estimate   Std.err  Wald Pr(>|W|)    
(Intercept)       0.318930  0.079035 16.28  5.5e-05 ***
weeks_post_fmt    0.000694  0.005429  0.02  0.89826    
antibiotics_ABX+ -0.257585  0.072683 12.56  0.00039 ***
---
Signif. codes:  0 ‘***’ 0.001 ‘**’ 0.01 ‘*’ 0.05 ‘.’ 0.1 ‘ ’ 1

Correlation structure = ar1 
Estimated Scale Parameters:

            Estimate Std.err
(Intercept)   0.0241 0.00496
  Link = identity 

Estimated Correlation Parameters:
      Estimate Std.err
alpha      1.6  0.0233
Number of clusters:   13  Maximum cluster size: 8 
                      lwr     upr
(Intercept)       0.16402  0.4738
weeks_post_fmt   -0.00995  0.0113
antibiotics_ABX+ -0.40004 -0.1151
Analysis of 'Wald statistic' Table

Model 1 subject_rabund ~ weeks_post_fmt + antibiotics_ 
Model 2 subject_rabund ~ weeks_post_fmt
  Df   X2 P(>|Chi|)    
1  1 12.6   0.00039 ***
---
Signif. codes:  0 ‘***’ 0.001 ‘**’ 0.01 ‘*’ 0.05 ‘.’ 0.1 ‘ ’ 1
[1] "--------"

Call:
geeglm(formula = donor_bc ~ weeks_post_fmt + antibiotics_, data = engraftment_gee_data_db, 
    id = subject_id, waves = engraftment_gee_data_db$weeks_post_fmt, 
    corstr = "ar1")

 Coefficients:
                 Estimate  Std.err   Wald Pr(>|W|)    
(Intercept)       0.72328  0.05080 202.69   <2e-16 ***
weeks_post_fmt    0.01953  0.00649   9.06   0.0026 ** 
antibiotics_ABX+ -0.10146  0.04162   5.94   0.0148 *  
---
Signif. codes:  0 ‘***’ 0.001 ‘**’ 0.01 ‘*’ 0.05 ‘.’ 0.1 ‘ ’ 1

Correlation structure = ar1 
Estimated Scale Parameters:

            Estimate Std.err
(Intercept)   0.0118 0.00338
  Link = identity 

Estimated Correlation Parameters:
      Estimate Std.err
alpha     1.31  0.0142
Number of clusters:   13  Maximum cluster size: 8 
                      lwr     upr
(Intercept)       0.62371  0.8229
weeks_post_fmt    0.00681  0.0322
antibiotics_ABX+ -0.18304 -0.0199
Analysis of 'Wald statistic' Table

Model 1 donor_bc ~ weeks_post_fmt + antibiotics_ 
Model 2 donor_bc ~ weeks_post_fmt
  Df   X2 P(>|Chi|)  
1  1 5.94     0.015 *
---
Signif. codes:  0 ‘***’ 0.001 ‘**’ 0.01 ‘*’ 0.05 ‘.’ 0.1 ‘ ’ 1
[1] "--------"

Call:
geeglm(formula = baseline_bc ~ weeks_post_fmt + antibiotics_, 
    data = engraftment_gee_data_sb, id = subject_id, waves = engraftment_gee_data_db$weeks_post_fmt, 
    corstr = "ar1")

 Coefficients:
                 Estimate Std.err   Wald Pr(>|W|)    
(Intercept)       0.76763 0.04387 306.15   <2e-16 ***
weeks_post_fmt    0.00113 0.00261   0.19     0.66    
antibiotics_ABX+  0.18558 0.04206  19.46    1e-05 ***
---
Signif. codes:  0 ‘***’ 0.001 ‘**’ 0.01 ‘*’ 0.05 ‘.’ 0.1 ‘ ’ 1

Correlation structure = ar1 
Estimated Scale Parameters:

            Estimate Std.err
(Intercept)  0.00987 0.00288
  Link = identity 

Estimated Correlation Parameters:
      Estimate Std.err
alpha     1.63   0.024
Number of clusters:   13  Maximum cluster size: 8 
                      lwr     upr
(Intercept)       0.68164 0.85362
weeks_post_fmt   -0.00398 0.00625
antibiotics_ABX+  0.10314 0.26802
Analysis of 'Wald statistic' Table

Model 1 baseline_bc ~ weeks_post_fmt + antibiotics_ 
Model 2 baseline_bc ~ weeks_post_fmt
  Df   X2 P(>|Chi|)    
1  1 19.5     1e-05 ***
---
Signif. codes:  0 ‘***’ 0.001 ‘**’ 0.01 ‘*’ 0.05 ‘.’ 0.1 ‘ ’ 1
[1] "--------"

Call:
geeglm(formula = other_rabund ~ weeks_post_fmt + antibiotics_, 
    data = engraftment_gee_data_sb, id = subject_id, waves = engraftment_gee_data_db$weeks_post_fmt, 
    corstr = "ar1")

 Coefficients:
                 Estimate Std.err Wald Pr(>|W|)  
(Intercept)       0.10692 0.04948 4.67    0.031 *
weeks_post_fmt    0.00865 0.00520 2.77    0.096 .
antibiotics_ABX+  0.00415 0.05236 0.01    0.937  
---
Signif. codes:  0 ‘***’ 0.001 ‘**’ 0.01 ‘*’ 0.05 ‘.’ 0.1 ‘ ’ 1

Correlation structure = ar1 
Estimated Scale Parameters:

            Estimate Std.err
(Intercept)   0.0182 0.00605
  Link = identity 

Estimated Correlation Parameters:
      Estimate Std.err
alpha    0.907  0.0432
Number of clusters:   13  Maximum cluster size: 8 
                      lwr    upr
(Intercept)       0.00994 0.2039
weeks_post_fmt   -0.00153 0.0188
antibiotics_ABX+ -0.09847 0.1068
Analysis of 'Wald statistic' Table

Model 1 other_rabund ~ weeks_post_fmt + antibiotics_ 
Model 2 other_rabund ~ weeks_post_fmt
  Df      X2 P(>|Chi|)
1  1 0.00629      0.94
[1] "--------"
```

###### Maintenance Delivery¶

In [ ]:

```
%%R

lm_mnt_dr <- geeglm(
    donor_rabund ~ weeks_post_fmt + maintenance_,
    id=subject_id,
    data=engraftment_gee_data_dr,
    corstr="ar1",
    waves=engraftment_gee_data_dr$weeks_post_fmt,
)
print(summary(lm_mnt_dr))
print(confint.geeglm(lm_mnt_dr))
print(anova(lm0_dr, lm_mnt_dr))
# print(aov(lm_mnt_dr))
print('--------')

lm_mnt_sr <- geeglm(
    subject_rabund ~ weeks_post_fmt + maintenance_,
    id=subject_id,
    data=engraftment_gee_data_sr,
    corstr="ar1",
    waves=engraftment_gee_data_sr$weeks_post_fmt
)
print(summary(lm_mnt_sr))
print(confint.geeglm(lm_mnt_sr))
print(anova(lm0_sr, lm_mnt_sr))
print('--------')

lm_mnt_db <- geeglm(
    donor_bc ~ weeks_post_fmt + maintenance_,
    id=subject_id,
    data=engraftment_gee_data_db,
    corstr="ar1",
    waves=engraftment_gee_data_db$weeks_post_fmt
)
print(summary(lm_mnt_db))
print(confint.geeglm(lm_mnt_db))
print(anova(lm0_db, lm_mnt_db))
print('--------')

lm_mnt_sb <- geeglm(
    baseline_bc ~ weeks_post_fmt + maintenance_,
    id=subject_id,
    data=engraftment_gee_data_sb,
    corstr="ar1",
    waves=engraftment_gee_data_db$weeks_post_fmt
)
print(summary(lm_mnt_sb))
print(confint.geeglm(lm_mnt_sb))
print(anova(lm0_sb, lm_mnt_sb))
print('--------')
```

```
Call:
geeglm(formula = donor_rabund ~ weeks_post_fmt + maintenance_, 
    data = engraftment_gee_data_dr, id = subject_id, waves = engraftment_gee_data_dr$weeks_post_fmt, 
    corstr = "ar1")

 Coefficients:
                 Estimate  Std.err  Wald Pr(>|W|)    
(Intercept)       0.54242  0.08806 37.94  7.3e-10 ***
weeks_post_fmt   -0.00695  0.00621  1.25     0.26    
maintenance_ENMA  0.11274  0.10502  1.15     0.28    
---
Signif. codes:  0 ‘***’ 0.001 ‘**’ 0.01 ‘*’ 0.05 ‘.’ 0.1 ‘ ’ 1

Correlation structure = ar1 
Estimated Scale Parameters:

            Estimate Std.err
(Intercept)   0.0657  0.0137
  Link = identity 

Estimated Correlation Parameters:
      Estimate Std.err
alpha    0.974  0.0122
Number of clusters:   13  Maximum cluster size: 8 
                     lwr     upr
(Intercept)       0.3698 0.71501
weeks_post_fmt   -0.0191 0.00523
maintenance_ENMA -0.0931 0.31857
Analysis of 'Wald statistic' Table

Model 1 donor_rabund ~ weeks_post_fmt + maintenance_ 
Model 2 donor_rabund ~ weeks_post_fmt
  Df   X2 P(>|Chi|)
1  1 1.15      0.28
[1] "--------"

Call:
geeglm(formula = subject_rabund ~ weeks_post_fmt + maintenance_, 
    data = engraftment_gee_data_sr, id = subject_id, waves = engraftment_gee_data_sr$weeks_post_fmt, 
    corstr = "ar1")

 Coefficients:
                 Estimate  Std.err  Wald Pr(>|W|)    
(Intercept)       0.21595  0.06234 12.00  0.00053 ***
weeks_post_fmt    0.00148  0.00364  0.17  0.68359    
maintenance_ENMA -0.03058  0.08250  0.14  0.71090    
---
Signif. codes:  0 ‘***’ 0.001 ‘**’ 0.01 ‘*’ 0.05 ‘.’ 0.1 ‘ ’ 1

Correlation structure = ar1 
Estimated Scale Parameters:

            Estimate Std.err
(Intercept)   0.0359 0.00732
  Link = identity 

Estimated Correlation Parameters:
      Estimate Std.err
alpha    0.983  0.0046
Number of clusters:   13  Maximum cluster size: 8 
                      lwr     upr
(Intercept)       0.09376 0.33815
weeks_post_fmt   -0.00565 0.00862
maintenance_ENMA -0.19227 0.13112
Analysis of 'Wald statistic' Table

Model 1 subject_rabund ~ weeks_post_fmt + maintenance_ 
Model 2 subject_rabund ~ weeks_post_fmt
  Df    X2 P(>|Chi|)
1  1 0.137      0.71
[1] "--------"

Call:
geeglm(formula = donor_bc ~ weeks_post_fmt + maintenance_, data = engraftment_gee_data_db, 
    id = subject_id, waves = engraftment_gee_data_db$weeks_post_fmt, 
    corstr = "ar1")

 Coefficients:
                 Estimate  Std.err   Wald Pr(>|W|)    
(Intercept)       0.82272  0.03010 747.22   <2e-16 ***
weeks_post_fmt    0.00337  0.00186   3.29    0.070 .  
maintenance_ENMA -0.06299  0.03129   4.05    0.044 *  
---
Signif. codes:  0 ‘***’ 0.001 ‘**’ 0.01 ‘*’ 0.05 ‘.’ 0.1 ‘ ’ 1

Correlation structure = ar1 
Estimated Scale Parameters:

            Estimate Std.err
(Intercept)  0.00993 0.00189
  Link = identity 

Estimated Correlation Parameters:
      Estimate Std.err
alpha    0.955   0.013
Number of clusters:   13  Maximum cluster size: 8 
                      lwr      upr
(Intercept)       0.76373  0.88171
weeks_post_fmt   -0.00027  0.00702
maintenance_ENMA -0.12433 -0.00166
Analysis of 'Wald statistic' Table

Model 1 donor_bc ~ weeks_post_fmt + maintenance_ 
Model 2 donor_bc ~ weeks_post_fmt
  Df   X2 P(>|Chi|)  
1  1 4.05     0.044 *
---
Signif. codes:  0 ‘***’ 0.001 ‘**’ 0.01 ‘*’ 0.05 ‘.’ 0.1 ‘ ’ 1
[1] "--------"

Call:
geeglm(formula = baseline_bc ~ weeks_post_fmt + maintenance_, 
    data = engraftment_gee_data_sb, id = subject_id, waves = engraftment_gee_data_db$weeks_post_fmt, 
    corstr = "ar1")

 Coefficients:
                 Estimate  Std.err   Wald Pr(>|W|)    
(Intercept)      0.846472 0.059857 199.99   <2e-16 ***
weeks_post_fmt   0.002589 0.004121   0.39     0.53    
maintenance_ENMA 0.000134 0.067197   0.00     1.00    
---
Signif. codes:  0 ‘***’ 0.001 ‘**’ 0.01 ‘*’ 0.05 ‘.’ 0.1 ‘ ’ 1

Correlation structure = ar1 
Estimated Scale Parameters:

            Estimate Std.err
(Intercept)   0.0175  0.0047
  Link = identity 

Estimated Correlation Parameters:
      Estimate Std.err
alpha     1.26  0.0128
Number of clusters:   13  Maximum cluster size: 8 
                      lwr    upr
(Intercept)       0.72916 0.9638
weeks_post_fmt   -0.00549 0.0107
maintenance_ENMA -0.13157 0.1318
Analysis of 'Wald statistic' Table

Model 1 baseline_bc ~ weeks_post_fmt + maintenance_ 
Model 2 baseline_bc ~ weeks_post_fmt
  Df       X2 P(>|Chi|)
1  1 3.96e-06         1
[1] "--------"
```

###### Donor Subject ID¶

In [ ]:

```
%%R

lm_dnr_dr <- geeglm(
    donor_rabund ~ weeks_post_fmt + donor_subject_id,
    id=subject_id,
    data=engraftment_gee_data_dr,
    corstr="ar1",
    waves=engraftment_gee_data_dr$weeks_post_fmt,
)
print(summary(lm_dnr_dr))
print(confint.geeglm(lm_dnr_dr))
print(anova(lm0_dr, lm_dnr_dr))
# print(aov(lm_dnr_dr))
print('--------')

lm_dnr_sr <- geeglm(
    subject_rabund ~ weeks_post_fmt + donor_subject_id,
    id=subject_id,
    data=engraftment_gee_data_sr,
    corstr="ar1",
    waves=engraftment_gee_data_sr$weeks_post_fmt
)
print(summary(lm_dnr_sr))
print(confint.geeglm(lm_dnr_sr))
print(anova(lm0_sr, lm_dnr_sr))
print('--------')

lm_dnr_db <- geeglm(
    donor_bc ~ weeks_post_fmt + donor_subject_id,
    id=subject_id,
    data=engraftment_gee_data_db,
    corstr="ar1",
    waves=engraftment_gee_data_db$weeks_post_fmt
)
print(summary(lm_dnr_db))
print(confint.geeglm(lm_dnr_db))
print(anova(lm0_db, lm_dnr_db))
print('--------')

lm_dnr_sb <- geeglm(
    baseline_bc ~ weeks_post_fmt + donor_subject_id,
    id=subject_id,
    data=engraftment_gee_data_sb,
    corstr="ar1",
    waves=engraftment_gee_data_db$weeks_post_fmt
)
print(summary(lm_dnr_sb))
print(confint.geeglm(lm_dnr_sb))
print(anova(lm0_sb, lm_dnr_sb))
print('--------')
```

```
Call:
geeglm(formula = donor_rabund ~ weeks_post_fmt + donor_subject_id, 
    data = engraftment_gee_data_dr, id = subject_id, waves = engraftment_gee_data_dr$weeks_post_fmt, 
    corstr = "ar1")

 Coefficients:
                      Estimate  Std.err  Wald Pr(>|W|)    
(Intercept)            0.60708  0.07856 59.71  1.1e-14 ***
weeks_post_fmt        -0.00688  0.00640  1.16     0.28    
donor_subject_idD0097  0.08970  0.08787  1.04     0.31    
donor_subject_idD0485 -0.34602  0.07343 22.20  2.5e-06 ***
---
Signif. codes:  0 ‘***’ 0.001 ‘**’ 0.01 ‘*’ 0.05 ‘.’ 0.1 ‘ ’ 1

Correlation structure = ar1 
Estimated Scale Parameters:

            Estimate Std.err
(Intercept)   0.0423  0.0095
  Link = identity 

Estimated Correlation Parameters:
      Estimate Std.err
alpha    0.958  0.0207
Number of clusters:   13  Maximum cluster size: 8 
                          lwr      upr
(Intercept)            0.4531  0.76107
weeks_post_fmt        -0.0194  0.00566
donor_subject_idD0097 -0.0825  0.26192
donor_subject_idD0485 -0.4899 -0.20209
Analysis of 'Wald statistic' Table

Model 1 donor_rabund ~ weeks_post_fmt + donor_subject_id 
Model 2 donor_rabund ~ weeks_post_fmt
  Df   X2 P(>|Chi|)    
1  2 45.7   1.2e-10 ***
---
Signif. codes:  0 ‘***’ 0.001 ‘**’ 0.01 ‘*’ 0.05 ‘.’ 0.1 ‘ ’ 1
[1] "--------"

Call:
geeglm(formula = subject_rabund ~ weeks_post_fmt + donor_subject_id, 
    data = engraftment_gee_data_sr, id = subject_id, waves = engraftment_gee_data_sr$weeks_post_fmt, 
    corstr = "ar1")

 Coefficients:
                      Estimate Std.err  Wald Pr(>|W|)   
(Intercept)            0.17140 0.06307  7.38   0.0066 **
weeks_post_fmt         0.00187 0.00374  0.25   0.6177   
donor_subject_idD0097  0.00584 0.07705  0.01   0.9396   
donor_subject_idD0485  0.19036 0.05872 10.51   0.0012 **
---
Signif. codes:  0 ‘***’ 0.001 ‘**’ 0.01 ‘*’ 0.05 ‘.’ 0.1 ‘ ’ 1

Correlation structure = ar1 
Estimated Scale Parameters:

            Estimate Std.err
(Intercept)   0.0258 0.00645
  Link = identity 

Estimated Correlation Parameters:
      Estimate Std.err
alpha    0.988  0.0068
Number of clusters:   13  Maximum cluster size: 8 
                           lwr    upr
(Intercept)            0.04778 0.2950
weeks_post_fmt        -0.00546 0.0092
donor_subject_idD0097 -0.14517 0.1569
donor_subject_idD0485  0.07527 0.3055
Analysis of 'Wald statistic' Table

Model 1 subject_rabund ~ weeks_post_fmt + donor_subject_id 
Model 2 subject_rabund ~ weeks_post_fmt
  Df   X2 P(>|Chi|)    
1  2 15.1   0.00052 ***
---
Signif. codes:  0 ‘***’ 0.001 ‘**’ 0.01 ‘*’ 0.05 ‘.’ 0.1 ‘ ’ 1
[1] "--------"

Call:
geeglm(formula = donor_bc ~ weeks_post_fmt + donor_subject_id, 
    data = engraftment_gee_data_db, id = subject_id, waves = engraftment_gee_data_db$weeks_post_fmt, 
    corstr = "ar1")

 Coefficients:
                      Estimate  Std.err   Wald Pr(>|W|)    
(Intercept)            0.71817  0.06339 128.35   <2e-16 ***
weeks_post_fmt         0.01421  0.00642   4.90   0.0268 *  
donor_subject_idD0097 -0.06467  0.04543   2.03   0.1546    
donor_subject_idD0485  0.11980  0.03923   9.33   0.0023 ** 
---
Signif. codes:  0 ‘***’ 0.001 ‘**’ 0.01 ‘*’ 0.05 ‘.’ 0.1 ‘ ’ 1

Correlation structure = ar1 
Estimated Scale Parameters:

            Estimate Std.err
(Intercept)  0.00871 0.00274
  Link = identity 

Estimated Correlation Parameters:
      Estimate Std.err
alpha     1.38  0.0163
Number of clusters:   13  Maximum cluster size: 8 
                           lwr    upr
(Intercept)            0.59392 0.8424
weeks_post_fmt         0.00163 0.0268
donor_subject_idD0097 -0.15372 0.0244
donor_subject_idD0485  0.04292 0.1967
Analysis of 'Wald statistic' Table

Model 1 donor_bc ~ weeks_post_fmt + donor_subject_id 
Model 2 donor_bc ~ weeks_post_fmt
  Df   X2 P(>|Chi|)    
1  2 36.7   1.1e-08 ***
---
Signif. codes:  0 ‘***’ 0.001 ‘**’ 0.01 ‘*’ 0.05 ‘.’ 0.1 ‘ ’ 1
[1] "--------"

Call:
geeglm(formula = baseline_bc ~ weeks_post_fmt + donor_subject_id, 
    data = engraftment_gee_data_sb, id = subject_id, waves = engraftment_gee_data_db$weeks_post_fmt, 
    corstr = "ar1")

 Coefficients:
                       Estimate   Std.err   Wald Pr(>|W|)    
(Intercept)            0.894548  0.042620 440.54   <2e-16 ***
weeks_post_fmt        -0.000697  0.001051   0.44    0.507    
donor_subject_idD0097 -0.008047  0.060843   0.02    0.895    
donor_subject_idD0485 -0.168866  0.067113   6.33    0.012 *  
---
Signif. codes:  0 ‘***’ 0.001 ‘**’ 0.01 ‘*’ 0.05 ‘.’ 0.1 ‘ ’ 1

Correlation structure = ar1 
Estimated Scale Parameters:

            Estimate Std.err
(Intercept)   0.0123 0.00376
  Link = identity 

Estimated Correlation Parameters:
      Estimate Std.err
alpha    0.982 0.00777
Number of clusters:   13  Maximum cluster size: 8 
                           lwr      upr
(Intercept)            0.81102  0.97808
weeks_post_fmt        -0.00276  0.00136
donor_subject_idD0097 -0.12730  0.11120
donor_subject_idD0485 -0.30041 -0.03733
Analysis of 'Wald statistic' Table

Model 1 baseline_bc ~ weeks_post_fmt + donor_subject_id 
Model 2 baseline_bc ~ weeks_post_fmt
  Df   X2 P(>|Chi|)  
1  2 7.68     0.021 *
---
Signif. codes:  0 ‘***’ 0.001 ‘**’ 0.01 ‘*’ 0.05 ‘.’ 0.1 ‘ ’ 1
[1] "--------"
```

###### Antibiotics compared to alternative null¶

In [ ]:

```
%%R

lm_abx_dnr_dr <- geeglm(
    donor_rabund ~ weeks_post_fmt + antibiotics_ + donor_subject_id,
    id=subject_id,
    data=engraftment_gee_data_dr,
    corstr="ar1",
    waves=engraftment_gee_data_dr$weeks_post_fmt,
)
print(summary(lm_abx_dnr_dr))
print(confint.geeglm(lm_abx_dnr_dr))
print(anova(lm_dnr_dr, lm_abx_dnr_dr))
# print(aov(lm_abx_dnr_dr))
print('--------')

lm_abx_dnr_sr <- geeglm(
    subject_rabund ~ weeks_post_fmt + antibiotics_ + donor_subject_id,
    id=subject_id,
    data=engraftment_gee_data_sr,
    corstr="ar1",
    waves=engraftment_gee_data_sr$weeks_post_fmt
)
print(summary(lm_abx_dnr_sr))
print(confint.geeglm(lm_abx_dnr_sr))
print(anova(lm_dnr_sr, lm_abx_dnr_sr))
print('--------')

lm_abx_dnr_db <- geeglm(
    donor_bc ~ weeks_post_fmt + antibiotics_ + donor_subject_id,
    id=subject_id,
    data=engraftment_gee_data_db,
    corstr="ar1",
    waves=engraftment_gee_data_db$weeks_post_fmt
)
print(summary(lm_abx_dnr_db))
print(confint.geeglm(lm_abx_dnr_db))
print(anova(lm_dnr_db, lm_abx_dnr_db))
print('--------')

lm_abx_dnr_sb <- geeglm(
    baseline_bc ~ weeks_post_fmt + antibiotics_ + donor_subject_id,
    id=subject_id,
    data=engraftment_gee_data_sb,
    corstr="ar1",
    waves=engraftment_gee_data_db$weeks_post_fmt
)
print(summary(lm_abx_dnr_sb))
print(confint.geeglm(lm_abx_dnr_sb))
print(anova(lm_dnr_sb, lm_abx_dnr_sb))
print('--------')
```

```
Call:
geeglm(formula = donor_rabund ~ weeks_post_fmt + antibiotics_ + 
    donor_subject_id, data = engraftment_gee_data_dr, id = subject_id, 
    waves = engraftment_gee_data_dr$weeks_post_fmt, corstr = "ar1")

 Coefficients:
                      Estimate  Std.err  Wald Pr(>|W|)    
(Intercept)            0.59117  0.09525 38.52  5.4e-10 ***
weeks_post_fmt        -0.00658  0.00643  1.05  0.30589    
antibiotics_ABX+       0.02600  0.09375  0.08  0.78154    
donor_subject_idD0097  0.07990  0.08762  0.83  0.36187    
donor_subject_idD0485 -0.32541  0.09744 11.15  0.00084 ***
---
Signif. codes:  0 ‘***’ 0.001 ‘**’ 0.01 ‘*’ 0.05 ‘.’ 0.1 ‘ ’ 1

Correlation structure = ar1 
Estimated Scale Parameters:

            Estimate Std.err
(Intercept)   0.0417 0.00805
  Link = identity 

Estimated Correlation Parameters:
      Estimate Std.err
alpha    0.959  0.0192
Number of clusters:   13  Maximum cluster size: 8 
                          lwr      upr
(Intercept)            0.4045  0.77786
weeks_post_fmt        -0.0192  0.00602
antibiotics_ABX+      -0.1578  0.20976
donor_subject_idD0097 -0.0918  0.25163
donor_subject_idD0485 -0.5164 -0.13444
Analysis of 'Wald statistic' Table

Model 1 donor_rabund ~ weeks_post_fmt + antibiotics_ + donor_subject_id 
Model 2 donor_rabund ~ weeks_post_fmt + donor_subject_id
  Df     X2 P(>|Chi|)
1  1 0.0769      0.78
[1] "--------"

Call:
geeglm(formula = subject_rabund ~ weeks_post_fmt + antibiotics_ + 
    donor_subject_id, data = engraftment_gee_data_sr, id = subject_id, 
    waves = engraftment_gee_data_sr$weeks_post_fmt, corstr = "ar1")

 Coefficients:
                      Estimate  Std.err  Wald Pr(>|W|)    
(Intercept)            0.27238  0.08934  9.30  0.00230 ** 
weeks_post_fmt        -0.00611  0.01404  0.19  0.66378    
antibiotics_ABX+      -0.21736  0.07766  7.83  0.00513 ** 
donor_subject_idD0097  0.00739  0.07120  0.01  0.91733    
donor_subject_idD0485  0.38803  0.10538 13.56  0.00023 ***
---
Signif. codes:  0 ‘***’ 0.001 ‘**’ 0.01 ‘*’ 0.05 ‘.’ 0.1 ‘ ’ 1

Correlation structure = ar1 
Estimated Scale Parameters:

            Estimate Std.err
(Intercept)   0.0214  0.0039
  Link = identity 

Estimated Correlation Parameters:
      Estimate Std.err
alpha      1.1 0.00894
Number of clusters:   13  Maximum cluster size: 8 
                          lwr     upr
(Intercept)            0.0973  0.4475
weeks_post_fmt        -0.0336  0.0214
antibiotics_ABX+      -0.3696 -0.0651
donor_subject_idD0097 -0.1322  0.1469
donor_subject_idD0485  0.1815  0.5946
Analysis of 'Wald statistic' Table

Model 1 subject_rabund ~ weeks_post_fmt + antibiotics_ + donor_subject_id 
Model 2 subject_rabund ~ weeks_post_fmt + donor_subject_id
  Df   X2 P(>|Chi|)   
1  1 7.83    0.0051 **
---
Signif. codes:  0 ‘***’ 0.001 ‘**’ 0.01 ‘*’ 0.05 ‘.’ 0.1 ‘ ’ 1
[1] "--------"

Call:
geeglm(formula = donor_bc ~ weeks_post_fmt + antibiotics_ + donor_subject_id, 
    data = engraftment_gee_data_db, id = subject_id, waves = engraftment_gee_data_db$weeks_post_fmt, 
    corstr = "ar1")

 Coefficients:
                      Estimate  Std.err    Wald Pr(>|W|)    
(Intercept)            0.82089  0.02481 1094.34   <2e-16 ***
weeks_post_fmt         0.00358  0.00194    3.42   0.0644 .  
antibiotics_ABX+      -0.01624  0.02891    0.32   0.5741    
donor_subject_idD0097 -0.07535  0.02872    6.88   0.0087 ** 
donor_subject_idD0485  0.05539  0.02911    3.62   0.0571 .  
---
Signif. codes:  0 ‘***’ 0.001 ‘**’ 0.01 ‘*’ 0.05 ‘.’ 0.1 ‘ ’ 1

Correlation structure = ar1 
Estimated Scale Parameters:

            Estimate Std.err
(Intercept)  0.00718 0.00148
  Link = identity 

Estimated Correlation Parameters:
      Estimate Std.err
alpha    0.924  0.0116
Number of clusters:   13  Maximum cluster size: 8 
                            lwr      upr
(Intercept)            0.772258  0.86953
weeks_post_fmt        -0.000215  0.00738
antibiotics_ABX+      -0.072899  0.04041
donor_subject_idD0097 -0.131629 -0.01906
donor_subject_idD0485 -0.001677  0.11245
Analysis of 'Wald statistic' Table

Model 1 donor_bc ~ weeks_post_fmt + antibiotics_ + donor_subject_id 
Model 2 donor_bc ~ weeks_post_fmt + donor_subject_id
  Df    X2 P(>|Chi|)
1  1 0.316      0.57
[1] "--------"

Call:
geeglm(formula = baseline_bc ~ weeks_post_fmt + antibiotics_ + 
    donor_subject_id, data = engraftment_gee_data_sb, id = subject_id, 
    waves = engraftment_gee_data_db$weeks_post_fmt, corstr = "ar1")

 Coefficients:
                      Estimate  Std.err   Wald Pr(>|W|)    
(Intercept)            0.83665  0.04287 380.91   <2e-16 ***
weeks_post_fmt        -0.00112  0.00103   1.18   0.2773    
antibiotics_ABX+       0.14978  0.04904   9.33   0.0023 ** 
donor_subject_idD0097 -0.04987  0.04522   1.22   0.2701    
donor_subject_idD0485 -0.09767  0.06970   1.96   0.1611    
---
Signif. codes:  0 ‘***’ 0.001 ‘**’ 0.01 ‘*’ 0.05 ‘.’ 0.1 ‘ ’ 1

Correlation structure = ar1 
Estimated Scale Parameters:

            Estimate Std.err
(Intercept)    0.008 0.00215
  Link = identity 

Estimated Correlation Parameters:
      Estimate Std.err
alpha    0.975 0.00896
Number of clusters:   13  Maximum cluster size: 8 
                           lwr      upr
(Intercept)            0.75263 0.920668
weeks_post_fmt        -0.00313 0.000898
antibiotics_ABX+       0.05367 0.245896
donor_subject_idD0097 -0.13850 0.038765
donor_subject_idD0485 -0.23428 0.038927
Analysis of 'Wald statistic' Table

Model 1 baseline_bc ~ weeks_post_fmt + antibiotics_ + donor_subject_id 
Model 2 baseline_bc ~ weeks_post_fmt + donor_subject_id
  Df   X2 P(>|Chi|)   
1  1 9.33    0.0023 **
---
Signif. codes:  0 ‘***’ 0.001 ‘**’ 0.01 ‘*’ 0.05 ‘.’ 0.1 ‘ ’ 1
[1] "--------"
```

###### Maintenance method compared to alternative null¶

In [ ]:

```
%%R

lm_mnt_dnr_dr <- geeglm(
    donor_rabund ~ weeks_post_fmt + maintenance_ + donor_subject_id,
    id=subject_id,
    data=engraftment_gee_data_dr,
    corstr="ar1",
    waves=engraftment_gee_data_dr$weeks_post_fmt,
)
print(summary(lm_mnt_dnr_dr))
print(confint.geeglm(lm_mnt_dnr_dr))
print(anova(lm_dnr_dr, lm_mnt_dnr_dr))
# print(aov(lm_mnt_dnr_dr))
print('--------')

lm_mnt_dnr_sr <- geeglm(
    subject_rabund ~ weeks_post_fmt + maintenance_ + donor_subject_id,
    id=subject_id,
    data=engraftment_gee_data_sr,
    corstr="ar1",
    waves=engraftment_gee_data_sr$weeks_post_fmt
)
print(summary(lm_mnt_dnr_sr))
print(confint.geeglm(lm_mnt_dnr_sr))
print(anova(lm_dnr_sr, lm_mnt_dnr_sr))
print('--------')

lm_mnt_dnr_db <- geeglm(
    donor_bc ~ weeks_post_fmt + maintenance_ + donor_subject_id,
    id=subject_id,
    data=engraftment_gee_data_db,
    corstr="ar1",
    waves=engraftment_gee_data_db$weeks_post_fmt
)
print(summary(lm_mnt_dnr_db))
print(confint.geeglm(lm_mnt_dnr_db))
print(anova(lm_dnr_db, lm_mnt_dnr_db))
print('--------')

lm_mnt_dnr_sb <- geeglm(
    baseline_bc ~ weeks_post_fmt + maintenance_ + donor_subject_id,
    id=subject_id,
    data=engraftment_gee_data_sb,
    corstr="ar1",
    waves=engraftment_gee_data_db$weeks_post_fmt
)
print(summary(lm_mnt_dnr_sb))
print(confint.geeglm(lm_mnt_dnr_sb))
print(anova(lm_dnr_sb, lm_mnt_dnr_sb))
print('--------')
```

```
Call:
geeglm(formula = donor_rabund ~ weeks_post_fmt + maintenance_ + 
    donor_subject_id, data = engraftment_gee_data_dr, id = subject_id, 
    waves = engraftment_gee_data_dr$weeks_post_fmt, corstr = "ar1")

 Coefficients:
                      Estimate  Std.err   Wald Pr(>|W|)    
(Intercept)            0.62832  0.06041 108.17  < 2e-16 ***
weeks_post_fmt        -0.00708  0.00643   1.21     0.27    
maintenance_ENMA      -0.09407  0.13586   0.48     0.49    
donor_subject_idD0097  0.15015  0.13623   1.21     0.27    
donor_subject_idD0485 -0.36778  0.06119  36.13  1.8e-09 ***
---
Signif. codes:  0 ‘***’ 0.001 ‘**’ 0.01 ‘*’ 0.05 ‘.’ 0.1 ‘ ’ 1

Correlation structure = ar1 
Estimated Scale Parameters:

            Estimate Std.err
(Intercept)   0.0395 0.00579
  Link = identity 

Estimated Correlation Parameters:
      Estimate Std.err
alpha    0.952  0.0213
Number of clusters:   13  Maximum cluster size: 8 
                          lwr      upr
(Intercept)            0.5099  0.74672
weeks_post_fmt        -0.0197  0.00551
maintenance_ENMA      -0.3603  0.17222
donor_subject_idD0097 -0.1169  0.41716
donor_subject_idD0485 -0.4877 -0.24785
Analysis of 'Wald statistic' Table

Model 1 donor_rabund ~ weeks_post_fmt + maintenance_ + donor_subject_id 
Model 2 donor_rabund ~ weeks_post_fmt + donor_subject_id
  Df    X2 P(>|Chi|)
1  1 0.479      0.49
[1] "--------"

Call:
geeglm(formula = subject_rabund ~ weeks_post_fmt + maintenance_ + 
    donor_subject_id, data = engraftment_gee_data_sr, id = subject_id, 
    waves = engraftment_gee_data_sr$weeks_post_fmt, corstr = "ar1")

 Coefficients:
                      Estimate  Std.err  Wald Pr(>|W|)    
(Intercept)            0.16415  0.06356  6.67   0.0098 ** 
weeks_post_fmt        -0.00540  0.00528  1.05   0.3059    
maintenance_ENMA       0.14567  0.14488  1.01   0.3147    
donor_subject_idD0097 -0.12006  0.14509  0.68   0.4080    
donor_subject_idD0485  0.41335  0.05104 65.58  5.6e-16 ***
---
Signif. codes:  0 ‘***’ 0.001 ‘**’ 0.01 ‘*’ 0.05 ‘.’ 0.1 ‘ ’ 1

Correlation structure = ar1 
Estimated Scale Parameters:

            Estimate Std.err
(Intercept)    0.022 0.00413
  Link = identity 

Estimated Correlation Parameters:
      Estimate Std.err
alpha     1.57  0.0224
Number of clusters:   13  Maximum cluster size: 8 
                          lwr     upr
(Intercept)            0.0396 0.28872
weeks_post_fmt        -0.0157 0.00494
maintenance_ENMA      -0.1383 0.42962
donor_subject_idD0097 -0.4044 0.16431
donor_subject_idD0485  0.3133 0.51339
Analysis of 'Wald statistic' Table

Model 1 subject_rabund ~ weeks_post_fmt + maintenance_ + donor_subject_id 
Model 2 subject_rabund ~ weeks_post_fmt + donor_subject_id
  Df   X2 P(>|Chi|)
1  1 1.01      0.31
[1] "--------"

Call:
geeglm(formula = donor_bc ~ weeks_post_fmt + maintenance_ + donor_subject_id, 
    data = engraftment_gee_data_db, id = subject_id, waves = engraftment_gee_data_db$weeks_post_fmt, 
    corstr = "ar1")

 Coefficients:
                      Estimate  Std.err   Wald Pr(>|W|)    
(Intercept)            0.68800  0.06054 129.15  < 2e-16 ***
weeks_post_fmt         0.01693  0.00719   5.54  0.01861 *  
maintenance_ENMA       0.06857  0.04627   2.20  0.13830    
donor_subject_idD0097 -0.10826  0.04658   5.40  0.02011 *  
donor_subject_idD0485  0.13627  0.03556  14.68  0.00013 ***
---
Signif. codes:  0 ‘***’ 0.001 ‘**’ 0.01 ‘*’ 0.05 ‘.’ 0.1 ‘ ’ 1

Correlation structure = ar1 
Estimated Scale Parameters:

            Estimate Std.err
(Intercept)   0.0095 0.00343
  Link = identity 

Estimated Correlation Parameters:
      Estimate Std.err
alpha     1.27  0.0129
Number of clusters:   13  Maximum cluster size: 8 
                           lwr    upr
(Intercept)            0.56935  0.807
weeks_post_fmt         0.00283  0.031
maintenance_ENMA      -0.02211  0.159
donor_subject_idD0097 -0.19955 -0.017
donor_subject_idD0485  0.06656  0.206
Analysis of 'Wald statistic' Table

Model 1 donor_bc ~ weeks_post_fmt + maintenance_ + donor_subject_id 
Model 2 donor_bc ~ weeks_post_fmt + donor_subject_id
  Df  X2 P(>|Chi|)
1  1 2.2      0.14
[1] "--------"

Call:
geeglm(formula = baseline_bc ~ weeks_post_fmt + maintenance_ + 
    donor_subject_id, data = engraftment_gee_data_sb, id = subject_id, 
    waves = engraftment_gee_data_db$weeks_post_fmt, corstr = "ar1")

 Coefficients:
                       Estimate   Std.err    Wald Pr(>|W|)    
(Intercept)            0.921112  0.023244 1570.31  < 2e-16 ***
weeks_post_fmt        -0.000487  0.001006    0.23  0.62811    
maintenance_ENMA      -0.137938  0.059015    5.46  0.01942 *  
donor_subject_idD0097  0.080625  0.059087    1.86  0.17240    
donor_subject_idD0485 -0.202620  0.055171   13.49  0.00024 ***
---
Signif. codes:  0 ‘***’ 0.001 ‘**’ 0.01 ‘*’ 0.05 ‘.’ 0.1 ‘ ’ 1

Correlation structure = ar1 
Estimated Scale Parameters:

            Estimate Std.err
(Intercept)  0.00975 0.00219
  Link = identity 

Estimated Correlation Parameters:
      Estimate Std.err
alpha    0.973  0.0125
Number of clusters:   13  Maximum cluster size: 8 
                           lwr      upr
(Intercept)            0.87555  0.96667
weeks_post_fmt        -0.00246  0.00148
maintenance_ENMA      -0.25361 -0.02227
donor_subject_idD0097 -0.03518  0.19643
donor_subject_idD0485 -0.31075 -0.09449
Analysis of 'Wald statistic' Table

Model 1 baseline_bc ~ weeks_post_fmt + maintenance_ + donor_subject_id 
Model 2 baseline_bc ~ weeks_post_fmt + donor_subject_id
  Df   X2 P(>|Chi|)  
1  1 5.46     0.019 *
---
Signif. codes:  0 ‘***’ 0.001 ‘**’ 0.01 ‘*’ 0.05 ‘.’ 0.1 ‘ ’ 1
[1] "--------"
```

###### Responder Status¶

In [ ]:

```
%%R

lm_rsp_dr <- geeglm(
    donor_rabund ~ weeks_post_fmt + responder,
    id=subject_id,
    data=engraftment_gee_data_dr,
    corstr="ar1",
    waves=engraftment_gee_data_dr$weeks_post_fmt,
)
print(summary(lm_rsp_dr))
print(confint.geeglm(lm_rsp_dr))
print(anova(lm0_dr, lm_rsp_dr))
# print(aov(lm_rsp_dr))
print('--------')

lm_rsp_sr <- geeglm(
    subject_rabund ~ weeks_post_fmt + responder,
    id=subject_id,
    data=engraftment_gee_data_sr,
    corstr="ar1",
    waves=engraftment_gee_data_sr$weeks_post_fmt
)
print(summary(lm_rsp_sr))
print(confint.geeglm(lm_rsp_sr))
print(anova(lm0_sr, lm_rsp_sr))
print('--------')

lm_rsp_db <- geeglm(
    donor_bc ~ weeks_post_fmt + responder,
    id=subject_id,
    data=engraftment_gee_data_db,
    corstr="ar1",
    waves=engraftment_gee_data_db$weeks_post_fmt
)
print(summary(lm_rsp_db))
print(confint.geeglm(lm_rsp_db))
print(anova(lm0_db, lm_rsp_db))
print('--------')

lm_rsp_sb <- geeglm(
    baseline_bc ~ weeks_post_fmt + responder,
    id=subject_id,
    data=engraftment_gee_data_sb,
    corstr="ar1",
    waves=engraftment_gee_data_db$weeks_post_fmt
)
print(summary(lm_rsp_sb))
print(confint.geeglm(lm_rsp_sb))
print(anova(lm0_sb, lm_rsp_sb))
print('--------')
```

```
Call:
geeglm(formula = donor_rabund ~ weeks_post_fmt + responder, data = engraftment_gee_data_dr, 
    id = subject_id, waves = engraftment_gee_data_dr$weeks_post_fmt, 
    corstr = "ar1")

 Coefficients:
               Estimate  Std.err  Wald Pr(>|W|)    
(Intercept)     0.63378  0.09020 49.37  2.1e-12 ***
weeks_post_fmt -0.00707  0.00624  1.28     0.26    
responder      -0.08103  0.10688  0.57     0.45    
---
Signif. codes:  0 ‘***’ 0.001 ‘**’ 0.01 ‘*’ 0.05 ‘.’ 0.1 ‘ ’ 1

Correlation structure = ar1 
Estimated Scale Parameters:

            Estimate Std.err
(Intercept)   0.0661  0.0142
  Link = identity 

Estimated Correlation Parameters:
      Estimate Std.err
alpha    0.972  0.0125
Number of clusters:   13  Maximum cluster size: 8 
                   lwr     upr
(Intercept)     0.4570 0.81056
weeks_post_fmt -0.0193 0.00516
responder      -0.2905 0.12845
Analysis of 'Wald statistic' Table

Model 1 donor_rabund ~ weeks_post_fmt + responder 
Model 2 donor_rabund ~ weeks_post_fmt
  Df    X2 P(>|Chi|)
1  1 0.575      0.45
[1] "--------"

Call:
geeglm(formula = subject_rabund ~ weeks_post_fmt + responder, 
    data = engraftment_gee_data_sr, id = subject_id, waves = engraftment_gee_data_sr$weeks_post_fmt, 
    corstr = "ar1")

 Coefficients:
               Estimate Std.err Wald Pr(>|W|)   
(Intercept)     0.18758 0.06452 8.45   0.0036 **
weeks_post_fmt  0.00165 0.00365 0.20   0.6513   
responder       0.03340 0.08186 0.17   0.6832   
---
Signif. codes:  0 ‘***’ 0.001 ‘**’ 0.01 ‘*’ 0.05 ‘.’ 0.1 ‘ ’ 1

Correlation structure = ar1 
Estimated Scale Parameters:

            Estimate Std.err
(Intercept)   0.0364  0.0074
  Link = identity 

Estimated Correlation Parameters:
      Estimate Std.err
alpha    0.985 0.00455
Number of clusters:   13  Maximum cluster size: 8 
                    lwr     upr
(Intercept)     0.06113 0.31404
weeks_post_fmt -0.00551 0.00881
responder      -0.12703 0.19383
Analysis of 'Wald statistic' Table

Model 1 subject_rabund ~ weeks_post_fmt + responder 
Model 2 subject_rabund ~ weeks_post_fmt
  Df    X2 P(>|Chi|)
1  1 0.167      0.68
[1] "--------"

Call:
geeglm(formula = donor_bc ~ weeks_post_fmt + responder, data = engraftment_gee_data_db, 
    id = subject_id, waves = engraftment_gee_data_db$weeks_post_fmt, 
    corstr = "ar1")

 Coefficients:
               Estimate Std.err   Wald Pr(>|W|)    
(Intercept)     0.66504 0.05945 125.15   <2e-16 ***
weeks_post_fmt  0.02043 0.00688   8.83    0.003 ** 
responder       0.01449 0.04985   0.08    0.771    
---
Signif. codes:  0 ‘***’ 0.001 ‘**’ 0.01 ‘*’ 0.05 ‘.’ 0.1 ‘ ’ 1

Correlation structure = ar1 
Estimated Scale Parameters:

            Estimate Std.err
(Intercept)   0.0135 0.00386
  Link = identity 

Estimated Correlation Parameters:
      Estimate Std.err
alpha     1.28  0.0132
Number of clusters:   13  Maximum cluster size: 8 
                    lwr    upr
(Intercept)     0.54852 0.7816
weeks_post_fmt  0.00695 0.0339
responder      -0.08321 0.1122
Analysis of 'Wald statistic' Table

Model 1 donor_bc ~ weeks_post_fmt + responder 
Model 2 donor_bc ~ weeks_post_fmt
  Df     X2 P(>|Chi|)
1  1 0.0845      0.77
[1] "--------"

Call:
geeglm(formula = baseline_bc ~ weeks_post_fmt + responder, data = engraftment_gee_data_sb, 
    id = subject_id, waves = engraftment_gee_data_db$weeks_post_fmt, 
    corstr = "ar1")

 Coefficients:
                Estimate   Std.err   Wald Pr(>|W|)    
(Intercept)     0.839346  0.042924 382.36   <2e-16 ***
weeks_post_fmt -0.000567  0.007482   0.01     0.94    
responder       0.058628  0.076901   0.58     0.45    
---
Signif. codes:  0 ‘***’ 0.001 ‘**’ 0.01 ‘*’ 0.05 ‘.’ 0.1 ‘ ’ 1

Correlation structure = ar1 
Estimated Scale Parameters:

            Estimate Std.err
(Intercept)   0.0176 0.00451
  Link = identity 

Estimated Correlation Parameters:
      Estimate Std.err
alpha     1.09 0.00889
Number of clusters:   13  Maximum cluster size: 8 
                   lwr    upr
(Intercept)     0.7552 0.9235
weeks_post_fmt -0.0152 0.0141
responder      -0.0921 0.2094
Analysis of 'Wald statistic' Table

Model 1 baseline_bc ~ weeks_post_fmt + responder 
Model 2 baseline_bc ~ weeks_post_fmt
  Df    X2 P(>|Chi|)
1  1 0.581      0.45
[1] "--------"
```

##### Fraction of donor strains transferred as a function of treatment¶

In [ ]:

```
subject_x_strain_transfer.stack("sample_type")
```

Out[ ]:

|  | subject\_id | S0001 | S0004 | S0007 | S0008 | S0013 | S0021 | S0024 | S0027 | S0041 | S0047 | S0053 | S0055 | S0056 |
| --- | --- | --- | --- | --- | --- | --- | --- | --- | --- | --- | --- | --- | --- | --- |
| sotu\_id | sample\_type |  |  |  |  |  |  |  |  |  |  |  |  |  |
| 100002-s001 | followup\_1 | NaN | 1.0 | NaN | NaN | 0.0 | 1.0 | 0.0 | 1.0 | NaN | NaN | NaN | NaN | NaN |
|  | followup\_2 | NaN | 1.0 | NaN | NaN | 0.0 | 0.0 | 0.0 | 1.0 | NaN | NaN | NaN | NaN | NaN |
|  | pre\_maintenance\_1 | NaN | 1.0 | NaN | NaN | 0.0 | 0.0 | 0.0 | NaN | NaN | NaN | NaN | NaN | NaN |
|  | pre\_maintenance\_2 | NaN | 0.0 | NaN | NaN | 0.0 | 1.0 | 0.0 | 1.0 | NaN | NaN | NaN | NaN | NaN |
|  | pre\_maintenance\_3 | NaN | 1.0 | NaN | NaN | 0.0 | 1.0 | 0.0 | 1.0 | NaN | NaN | NaN | NaN | NaN |
| ... | ... | ... | ... | ... | ... | ... | ... | ... | ... | ... | ... | ... | ... | ... |
| 104593-s020 | pre\_maintenance\_2 | NaN | NaN | NaN | 0.0 | NaN | NaN | NaN | NaN | NaN | NaN | NaN | NaN | NaN |
|  | pre\_maintenance\_3 | NaN | NaN | 0.0 | 0.0 | NaN | NaN | NaN | NaN | NaN | NaN | NaN | NaN | NaN |
|  | pre\_maintenance\_4 | NaN | NaN | 0.0 | 0.0 | NaN | NaN | NaN | NaN | NaN | NaN | NaN | NaN | NaN |
|  | pre\_maintenance\_5 | NaN | NaN | 0.0 | 0.0 | NaN | NaN | NaN | NaN | NaN | NaN | NaN | NaN | NaN |
|  | pre\_maintenance\_6 | NaN | NaN | 0.0 | 0.0 | NaN | NaN | NaN | NaN | NaN | NaN | NaN | NaN | NaN |

9956 rows × 13 columns

In [ ]:

```
taxon_level = "d__"
taxon_order = ["d__Bacteria;"]
comparison = "antibiotics_"
sample_type = "followup_1"

opportunities = (
    (subject_x_strain_transfer.notna()).groupby(sotu_to_taxonomy[taxon_level]).sum()
)
transfers = (subject_x_strain_transfer).groupby(sotu_to_taxonomy[taxon_level]).sum()
no_transfers = opportunities - transfers
d = (
    (transfers / opportunities)
    .unstack()
    .rename("transfer_index")
    .reset_index()
    .rename(columns={"level_1": "subject_id", "level_0": "sample_type"})
    .join(subject, on="subject_id")
    .sort_values(taxon_level)[lambda x: x.sample_type == sample_type]
)

fig, ax = plt.subplots(figsize=(14, 5))
lib.plot.boxplot_with_points(
    y="transfer_index",
    x=taxon_level,
    hue=comparison,
    palette=lib.project_style.DEFAULT_COLOR_PALETTE,
    order=taxon_order,
    data=d,
    ax=ax,
    dodge=True,
    dist_kwargs={"boxprops": {"alpha": 0.5}},
)
lib.plot.rotate_xticklabels(ax, rotation=90)
ax.legend(bbox_to_anchor=(1, 1))

print(
    d[lambda x: x[taxon_level].isin(taxon_order)]
    .groupby(taxon_level)
    .apply(
        lambda d: lib.stats.mannwhitneyu(
            comparison, "transfer_index", data=d, na_action="drop"
        )
    )
    .loc[taxon_order]
)
```

```
d__
d__Bacteria;    (0.7619047619047619, 0.13752913752913754)
dtype: object
```

In [ ]:

```
taxon_level = "d__"
taxon_order = ["d__Bacteria;"]
comparison = "maintenance_"
sample_type = "followup_1"

opportunities = (
    (subject_x_strain_transfer.notna()).groupby(sotu_to_taxonomy[taxon_level]).sum()
)
transfers = (subject_x_strain_transfer).groupby(sotu_to_taxonomy[taxon_level]).sum()
no_transfers = opportunities - transfers
d = (
    (transfers / opportunities)
    .unstack()
    .rename("transfer_index")
    .reset_index()
    .rename(columns={"level_1": "subject_id", "level_0": "sample_type"})
    .join(subject, on="subject_id")
    .sort_values(taxon_level)[lambda x: x.sample_type == sample_type]
)

fig, ax = plt.subplots(figsize=(14, 5))
lib.plot.boxplot_with_points(
    y="transfer_index",
    x=taxon_level,
    hue=comparison,
    palette=lib.project_style.DEFAULT_COLOR_PALETTE,
    order=taxon_order,
    data=d,
    ax=ax,
    dodge=True,
    dist_kwargs={"boxprops": {"alpha": 0.5}},
)
lib.plot.rotate_xticklabels(ax, rotation=90)
ax.legend(bbox_to_anchor=(1, 1))

print(
    d[lambda x: x[taxon_level].isin(taxon_order)]
    .groupby(taxon_level)
    .apply(
        lambda d: lib.stats.mannwhitneyu(
            comparison, "transfer_index", data=d, na_action="drop"
        )
    )
    .loc[taxon_order]
)
```

```
d__
d__Bacteria;    (0.7619047619047619, 0.13752913752913754)
dtype: object
```

In [ ]:

```
taxon_level = "d__"
taxon_order = ["d__Bacteria;"]
comparison = "donor_subject_id"
sample_type = "followup_1"
pallete = subject_color_palette


opportunities = (
    (subject_x_strain_transfer.notna()).groupby(sotu_to_taxonomy[taxon_level]).sum()
)
transfers = (subject_x_strain_transfer).groupby(sotu_to_taxonomy[taxon_level]).sum()
no_transfers = opportunities - transfers
d = (
    (transfers / opportunities)
    .unstack()
    .rename("transfer_index")
    .reset_index()
    .rename(columns={"level_1": "subject_id", "level_0": "sample_type"})
    .join(subject, on="subject_id")
    .sort_values(taxon_level)[lambda x: x.sample_type == sample_type]
)

fig, ax = plt.subplots(figsize=(14, 5))
lib.plot.boxplot_with_points(
    y="transfer_index",
    x=taxon_level,
    hue=comparison,
    palette=pallete,
    order=taxon_order,
    data=d,
    ax=ax,
    dodge=True,
    dist_kwargs={"boxprops": {"alpha": 0.5}},
)
lib.plot.rotate_xticklabels(ax, rotation=90)
ax.legend(bbox_to_anchor=(1, 1))

print(
    d[lambda x: x[taxon_level].isin(taxon_order)]
    .groupby(taxon_level)
    .apply(
        lambda d: lib.stats.mannwhitneyu(
            comparison,
            "transfer_index",
            data=d[d.donor_subject_id.isin(["D0044", "D0097"])],
            na_action="drop",
        )
    )
    .loc[taxon_order]
)
```

```
d__
d__Bacteria;    (0.8333333333333334, 0.08225108225108226)
dtype: object
```

In [ ]:

```
taxon_level = "d__"
taxon_order = ["d__Bacteria;"]
comparison = "responder"
sample_type = "followup_1"

opportunities = (
    (subject_x_strain_transfer.notna()).groupby(sotu_to_taxonomy[taxon_level]).sum()
)
transfers = (subject_x_strain_transfer).groupby(sotu_to_taxonomy[taxon_level]).sum()
no_transfers = opportunities - transfers
d = (
    (transfers / opportunities)
    .unstack()
    .rename("transfer_index")
    .reset_index()
    .rename(columns={"level_1": "subject_id", "level_0": "sample_type"})
    .join(subject, on="subject_id")
    .sort_values(taxon_level)[lambda x: x.sample_type == sample_type]
)

fig, ax = plt.subplots(figsize=(14, 5))
lib.plot.boxplot_with_points(
    y="transfer_index",
    x=taxon_level,
    hue=comparison,
    palette=lib.project_style.DEFAULT_COLOR_PALETTE,
    order=taxon_order,
    data=d,
    ax=ax,
    dodge=True,
    dist_kwargs={"boxprops": {"alpha": 0.5}},
)
lib.plot.rotate_xticklabels(ax, rotation=90)
ax.legend(bbox_to_anchor=(1, 1))

print(
    d[lambda x: x[taxon_level].isin(taxon_order)]
    .groupby(taxon_level)
    .apply(
        lambda d: lib.stats.mannwhitneyu(
            comparison, "transfer_index", data=d, na_action="drop"
        )
    )
    .loc[taxon_order]
)
```

```
d__
d__Bacteria;    (0.7142857142857143, 0.23426573426573427)
dtype: object
```

In [ ]:

```
sample_x_sotu.loc[sample.sample_type.isin(["donor_mean", "baseline"])].groupby(
    sotu_to_taxonomy.p__, axis="columns"
).sum().mean().sort_values().tail(6)
```

Out[ ]:

```
p__
d__Bacteria;p__Firmicutes_C;        0.028479
d__Bacteria;p__Proteobacteria;      0.036457
d__Bacteria;p__Firmicutes;          0.081450
d__Bacteria;p__Actinobacteriota;    0.103703
d__Bacteria;p__Bacteroidota;        0.183250
d__Bacteria;p__Firmicutes_A;        0.539989
dtype: float64
```

In [ ]:

```
top_phylum_donor_median_abundance = (
    sample_x_phylum.loc[["D0044_mean", "D0097_mean", "D0485_mean"]]
    .median()
    .sort_values(ascending=False)
    .head(6)
)
top_phylum_donor_median_abundance
```

Out[ ]:

```
p__
d__Bacteria;p__Firmicutes_A;        0.730048
d__Bacteria;p__Bacteroidota;        0.230452
d__Bacteria;p__Actinobacteriota;    0.038871
d__Bacteria;p__Firmicutes;          0.014149
d__Bacteria;p__Proteobacteria;      0.010157
d__Bacteria;p__Firmicutes_C;        0.008239
dtype: float64
```

In [ ]:

```
taxon_level = "p__"
taxon_order = top_phylum_donor_median_abundance.index
comparison = "antibiotics_"
sample_type = "followup_1"

opportunities = (
    (subject_x_strain_transfer.notna()).groupby(sotu_to_taxonomy[taxon_level]).sum()
)
transfers = (subject_x_strain_transfer).groupby(sotu_to_taxonomy[taxon_level]).sum()
no_transfers = opportunities - transfers
d = (
    (transfers / opportunities)
    .unstack()
    .rename("transfer_index")
    .reset_index()
    .rename(columns={"level_1": "subject_id", "level_0": "sample_type"})
    .join(subject, on="subject_id")
    .sort_values(taxon_level)[lambda x: x.sample_type == sample_type]
)

fig, ax = plt.subplots(figsize=(14, 5))
lib.plot.boxplot_with_points(
    y="transfer_index",
    x=taxon_level,
    hue=comparison,
    palette=lib.project_style.DEFAULT_COLOR_PALETTE,
    order=taxon_order,
    data=d,
    ax=ax,
    dodge=True,
    dist_kwargs={"boxprops": {"alpha": 0.5}},
)
lib.plot.rotate_xticklabels(ax, rotation=90)
ax.legend(bbox_to_anchor=(1, 1))

print(
    d[lambda x: x[taxon_level].isin(taxon_order)]
    .groupby(taxon_level)
    .apply(
        lambda d: lib.stats.mannwhitneyu(
            comparison, "transfer_index", data=d, na_action="drop"
        )
    )
    .loc[taxon_order]
)
```

```
p__
d__Bacteria;p__Firmicutes_A;        (0.16666666666666666, 0.05128205128205128)
d__Bacteria;p__Bacteroidota;          (0.6428571428571429, 0.4452214452214452)
d__Bacteria;p__Actinobacteriota;    (0.23809523809523808, 0.13254585342192138)
d__Bacteria;p__Firmicutes;          (0.8690476190476191, 0.031652756841673554)
d__Bacteria;p__Proteobacteria;      (0.23809523809523808, 0.12825007684924755)
d__Bacteria;p__Firmicutes_C;                                        (0.5, 1.0)
dtype: object
```

In [ ]:

```
def simplify_tax_string(s, prefix):
    return s.split(";")[-2].replace("_", " ")[len(prefix) :]


taxon_level = "p__"
taxon_order = sorted(top_phylum_donor_median_abundance.index)
comparison = "antibiotics_"
comparison_order = ["ABX-", "ABX+"]
sample_type = "followup_1"

opportunities = (
    (subject_x_strain_transfer.notna()).groupby(sotu_to_taxonomy[taxon_level]).sum()
)
transfers = (subject_x_strain_transfer).groupby(sotu_to_taxonomy[taxon_level]).sum()
no_transfers = opportunities - transfers
d0 = (
    (transfers / opportunities)
    .unstack()
    .rename("transfer_index")
    .reset_index()
    .rename(columns={"level_1": "subject_id", "level_0": "sample_type"})
    .join(subject, on="subject_id")
    .sort_values(taxon_level)[lambda x: x.sample_type == sample_type]
)

pvalues = (
    d0[lambda x: x[taxon_level].isin(taxon_order)]
    .groupby(taxon_level)
    .apply(
        lambda d: lib.stats.mannwhitneyu(
            comparison, "transfer_index", data=d, na_action="drop"
        )[1]
    )
    .loc[taxon_order]
)

fig, axs = plt.subplots(
    1,
    len(top_phylum_donor_median_abundance.index),
    figsize=(10, 4),
    sharex=True,
    sharey=True,
)

for tax, ax in zip(taxon_order, axs):
    d1 = d0[d0[taxon_level] == tax]
    lib.plot.boxplot_with_points(
        y="transfer_index",
        x=comparison,
        order=comparison_order,
        palette=lib.project_style.DEFAULT_COLOR_PALETTE,
        data=d1,
        ax=ax,
        dodge=True,
        dist_kwargs={"boxprops": {"alpha": 0.5}},
    )
    ax.set_title(simplify_tax_string(tax, taxon_level))
    ax.set_ylabel("")
    ax.set_xlabel("")
    ax.legend_.set_visible(False)
    annotation = lib.project_style.pvalue_to_annotation(pvalues[tax])
    if annotation == "∙":
        y = 1.161
    elif annotation == "**":
        y = 1.133
    elif annotation == "*":
        y = 1.133
    elif annotation == "":
        y = 1.133
    ax.annotate(annotation, xy=(0.5, y), fontsize=16, ha="center", va="center")
    ax.set_ylim(-0.05, 1.25)
    ax.set_xticklabels(["–", "+"])
    ax.set_yticks(np.linspace(0, 1, num=5))
#     lib.plot.rotate_xticklabels(ax, rotation=-45, ha='left')
# ax.legend(bbox_to_anchor=(1, 1))

axs[0].set_ylabel("Fraction Donor Strains")
ax.text(0.1, 0.072, "ABX →", fontsize=8, transform=fig.transFigure)


pvalues
```

Out[ ]:

```
p__
d__Bacteria;p__Actinobacteriota;    0.132546
d__Bacteria;p__Bacteroidota;        0.445221
d__Bacteria;p__Firmicutes;          0.031653
d__Bacteria;p__Firmicutes_A;        0.051282
d__Bacteria;p__Firmicutes_C;        1.000000
d__Bacteria;p__Proteobacteria;      0.128250
dtype: float64
```

In [ ]:

```
top_phylum_donor_median_abundance
```

Out[ ]:

```
p__
d__Bacteria;p__Firmicutes_A;        0.730048
d__Bacteria;p__Bacteroidota;        0.230452
d__Bacteria;p__Actinobacteriota;    0.038871
d__Bacteria;p__Firmicutes;          0.014149
d__Bacteria;p__Proteobacteria;      0.010157
d__Bacteria;p__Firmicutes_C;        0.008239
dtype: float64
```

In [ ]:

```
def simplify_tax_string(s, prefix):
    return s.split(";")[-2].replace("_", " ")[len(prefix) :]


taxon_level = "p__"
taxon_order = sorted(top_phylum_donor_median_abundance.index)
comparison = "antibiotics_"
comparison_order = ["ABX-", "ABX+"]
sample_type = "followup_1"

opportunities = (
    (subject_x_strain_transfer.notna()).groupby(sotu_to_taxonomy[taxon_level]).sum()
)
transfers = (subject_x_strain_transfer).groupby(sotu_to_taxonomy[taxon_level]).sum()
no_transfers = opportunities - transfers
d0 = (
    (transfers / opportunities)
    .unstack()
    .rename("transfer_index")
    .reset_index()
    .rename(columns={"level_1": "subject_id", "level_0": "sample_type"})
    .join(subject, on="subject_id")
    .sort_values(taxon_level)[lambda x: x.sample_type == sample_type]
)
jitter_width = 0.15
d0["jitter"] = np.random.uniform(
    -jitter_width,
    jitter_width,
    size=d0.shape[0],
)
comparison_x = pd.Series(range(len(comparison_order)), index=comparison_order)

pvalues = (
    d0[lambda x: x[taxon_level].isin(taxon_order)]
    .groupby(taxon_level)
    .apply(
        lambda d: lib.stats.mannwhitneyu(
            comparison, "transfer_index", data=d, na_action="drop"
        )[1]
    )
    .loc[taxon_order]
)

fig, axs = plt.subplots(
    # fig, ax = plt.subplots(
    nrows=1,
    #     ncols=1,
    ncols=len(top_phylum_donor_median_abundance.index),
    figsize=(9, 4),
    sharex=True,
    sharey=True,
)

# tax = 'd__Bacteria;p__Firmicutes;'
for tax, ax in zip(taxon_order, axs):
    d1 = d0[d0[taxon_level] == tax]

    for antibiotics_, d2 in d1.groupby("antibiotics_"):
        for maintenance_, d3 in d2.groupby("maintenance_"):
            arm = antibiotics_ + "/" + maintenance_
            ax.scatter(
                d3.jitter + comparison_x[antibiotics_],
                d3.transfer_index,
                c=lib.project_style.DEFAULT_COLOR_PALETTE[arm],
                edgecolor="grey",
                #                 edgecolor='none',
                s=50,
                zorder=1,
                alpha=0.8,
            )
        ax.boxplot(
            d2.transfer_index,
            positions=[comparison_x[antibiotics_]],
            widths=jitter_width * 2.5,
            boxprops=dict(lw=1),
            medianprops=dict(lw=1, linestyle="--", color="k"),
            whis=0,
            showfliers=False,
            zorder=0,
        )
    ax.set_title(simplify_tax_string(tax, taxon_level))
    ax.set_ylabel("")
    ax.set_xlabel("")
    # ax.legend_.set_visible(False)
    annotation = lib.project_style.pvalue_to_annotation(pvalues[tax])
    if annotation == "∙":
        y = 1.161
    elif annotation == "**":
        y = 1.133
    elif annotation == "*":
        y = 1.133
    elif annotation == "":
        y = 1.133
    ax.annotate(annotation, xy=(0.5, y), fontsize=16, ha="center", va="center")
    ax.set_ylim(-0.05, 1.25)
    ax.set_xlim(-2 * jitter_width, 1 + 2 * jitter_width)
    ax.set_xticks([0, 1])
    ax.set_xticklabels(["–", "+"])
    ax.set_yticks(np.linspace(0, 1, num=5))

axs[0].set_ylabel("Fraction Donor Strains")
ax.text(0.1, 0.072, "ABX →", fontsize=8, transform=fig.transFigure)

axs[0].text(
    -0.3, 1.05, "E", fontsize=12, fontweight="heavy", transform=axs[0].transAxes
)

fig.savefig("fig/engraftment_rates_by_phylum.pdf", bbox_inches="tight")
# pvalues
```

In [ ]:

```
taxon_level = "p__"
taxon_order = top_phylum_donor_median_abundance.index
comparison = "maintenance_"
sample_type = "followup_1"

opportunities = (
    (subject_x_strain_transfer.notna()).groupby(sotu_to_taxonomy[taxon_level]).sum()
)
transfers = (subject_x_strain_transfer).groupby(sotu_to_taxonomy[taxon_level]).sum()
no_transfers = opportunities - transfers
d = (
    (transfers / opportunities)
    .unstack()
    .rename("transfer_index")
    .reset_index()
    .rename(columns={"level_1": "subject_id", "level_0": "sample_type"})
    .join(subject, on="subject_id")
    .sort_values(taxon_level)[lambda x: x.sample_type == sample_type]
)

fig, ax = plt.subplots(figsize=(14, 5))
lib.plot.boxplot_with_points(
    y="transfer_index",
    x=taxon_level,
    hue=comparison,
    palette=lib.project_style.DEFAULT_COLOR_PALETTE,
    order=taxon_order,
    data=d,
    ax=ax,
    dodge=True,
    dist_kwargs={"boxprops": {"alpha": 0.5}},
)
lib.plot.rotate_xticklabels(ax, rotation=90)
ax.legend(bbox_to_anchor=(1, 1))

print(
    d[lambda x: x[taxon_level].isin(taxon_order)]
    .groupby(taxon_level)
    .apply(
        lambda d: lib.stats.mannwhitneyu(
            comparison, "transfer_index", data=d, na_action="drop"
        )
    )
    .loc[taxon_order]
)
```

```
p__
d__Bacteria;p__Firmicutes_A;         (0.7380952380952381, 0.18065268065268064)
d__Bacteria;p__Bacteroidota;         (0.2857142857142857, 0.23426573426573427)
d__Bacteria;p__Actinobacteriota;    (0.36904761904761907, 0.47383319059283024)
d__Bacteria;p__Firmicutes;           (0.39285714285714285, 0.5666413565522703)
d__Bacteria;p__Proteobacteria;                                      (0.5, 1.0)
d__Bacteria;p__Firmicutes_C;          (0.4642857142857143, 0.8821455812960055)
dtype: object
```

In [ ]:

```
taxon_level = "p__"
taxon_order = top_phylum_donor_median_abundance.index
comparison = "donor_subject_id"
sample_type = "followup_1"
pallete = subject_color_palette

opportunities = (
    (subject_x_strain_transfer.notna()).groupby(sotu_to_taxonomy[taxon_level]).sum()
)
transfers = (subject_x_strain_transfer).groupby(sotu_to_taxonomy[taxon_level]).sum()
no_transfers = opportunities - transfers
d = (
    (transfers / opportunities)
    .unstack()
    .rename("transfer_index")
    .reset_index()
    .rename(columns={"level_1": "subject_id", "level_0": "sample_type"})
    .join(subject, on="subject_id")
    .sort_values(taxon_level)[lambda x: x.sample_type == sample_type]
)

fig, ax = plt.subplots(figsize=(14, 5))
lib.plot.boxplot_with_points(
    y="transfer_index",
    x=taxon_level,
    hue=comparison,
    palette=pallete,
    order=taxon_order,
    data=d,
    ax=ax,
    dodge=True,
    dist_kwargs={"boxprops": {"alpha": 0.5}},
)
lib.plot.rotate_xticklabels(ax, rotation=90)
ax.legend(bbox_to_anchor=(1, 1))

print(
    d[lambda x: x[taxon_level].isin(taxon_order)]
    .groupby(taxon_level)
    .apply(
        lambda d: lib.stats.mannwhitneyu(
            comparison,
            "transfer_index",
            data=d[d.donor_subject_id.isin(["D0044", "D0097"])],
            na_action="drop",
        )
    )
    .loc[taxon_order]
)
```

```
p__
d__Bacteria;p__Firmicutes_A;         (0.8333333333333334, 0.08225108225108226)
d__Bacteria;p__Bacteroidota;         (0.7666666666666667, 0.17748917748917747)
d__Bacteria;p__Actinobacteriota;    (0.16666666666666666, 0.08213856110808998)
d__Bacteria;p__Firmicutes;           (0.7333333333333333, 0.23426713617079375)
d__Bacteria;p__Proteobacteria;       (0.8166666666666667, 0.09570475117488142)
d__Bacteria;p__Firmicutes_C;          (0.3333333333333333, 0.3970009440149582)
dtype: object
```

In [ ]:

```
taxon_level = "p__"
taxon_order = top_phylum_donor_median_abundance.index
comparison = "responder"
sample_type = "followup_1"

opportunities = (
    (subject_x_strain_transfer.notna()).groupby(sotu_to_taxonomy[taxon_level]).sum()
)
transfers = (subject_x_strain_transfer).groupby(sotu_to_taxonomy[taxon_level]).sum()
no_transfers = opportunities - transfers
d = (
    (transfers / opportunities)
    .unstack()
    .rename("transfer_index")
    .reset_index()
    .rename(columns={"level_1": "subject_id", "level_0": "sample_type"})
    .join(subject, on="subject_id")
    .sort_values(taxon_level)[lambda x: x.sample_type == sample_type]
)

fig, ax = plt.subplots(figsize=(14, 5))
lib.plot.boxplot_with_points(
    y="transfer_index",
    x=taxon_level,
    hue=comparison,
    palette=lib.project_style.DEFAULT_COLOR_PALETTE,
    order=taxon_order,
    data=d,
    ax=ax,
    dodge=True,
    dist_kwargs={"boxprops": {"alpha": 0.5}},
)
lib.plot.rotate_xticklabels(ax, rotation=90)
ax.legend(bbox_to_anchor=(1, 1))

print(
    d[lambda x: x[taxon_level].isin(taxon_order)]
    .groupby(taxon_level)
    .apply(
        lambda d: lib.stats.mannwhitneyu(
            comparison, "transfer_index", data=d, na_action="drop"
        )
    )
    .loc[taxon_order]
)
```

```
p__
d__Bacteria;p__Firmicutes_A;         (0.6428571428571429, 0.4452214452214452)
d__Bacteria;p__Bacteroidota;        (0.7857142857142857, 0.10139860139860139)
d__Bacteria;p__Actinobacteriota;      (0.6785714285714286, 0.315977335831649)
d__Bacteria;p__Firmicutes;          (0.42857142857142855, 0.7202474654940236)
d__Bacteria;p__Proteobacteria;      (0.42857142857142855, 0.7172383586012208)
d__Bacteria;p__Firmicutes_C;         (0.42857142857142855, 0.710917113388619)
dtype: object
```

In [ ]:

```
top_family_donor_median_abundance = (
    sample_x_family.loc[["D0044_mean", "D0097_mean", "D0485_mean"]]
    .median()
    .sort_values(ascending=False)
    .head(10)
)
top_family_donor_median_abundance
```

Out[ ]:

```
f__
d__Bacteria;p__Firmicutes_A;c__Clostridia;o__Lachnospirales;f__Lachnospiraceae;                0.320751
d__Bacteria;p__Firmicutes_A;c__Clostridia;o__Oscillospirales;f__Ruminococcaceae;               0.210927
d__Bacteria;p__Bacteroidota;c__Bacteroidia;o__Bacteroidales;f__Bacteroidaceae;                 0.136459
d__Bacteria;p__Firmicutes_A;c__Clostridia;o__Oscillospirales;f__DTU089;                        0.050016
d__Bacteria;p__Firmicutes_A;c__Clostridia;o__Oscillospirales;f__Oscillospiraceae;              0.042416
d__Bacteria;p__Bacteroidota;c__Bacteroidia;o__Bacteroidales;f__Rikenellaceae;                  0.031743
d__Bacteria;p__Actinobacteriota;c__Actinobacteria;o__Actinomycetales;f__Bifidobacteriaceae;    0.030945
d__Bacteria;p__Bacteroidota;c__Bacteroidia;o__Bacteroidales;f__Tannerellaceae;                 0.012909
d__Bacteria;p__Firmicutes;c__Bacilli;o__Erysipelotrichales;f__Erysipelatoclostridiaceae;       0.010586
d__Bacteria;p__Actinobacteriota;c__Coriobacteriia;o__Coriobacteriales;f__Eggerthellaceae;      0.009423
dtype: float64
```

In [ ]:

```
taxon_level = "f__"
taxon_order = top_family_donor_median_abundance.index
comparison = "antibiotics_"
sample_type = "followup_1"

opportunities = (
    (subject_x_strain_transfer.notna()).groupby(sotu_to_taxonomy[taxon_level]).sum()
)
transfers = (subject_x_strain_transfer).groupby(sotu_to_taxonomy[taxon_level]).sum()
no_transfers = opportunities - transfers
d = (
    (transfers / opportunities)
    .unstack()
    .rename("transfer_index")
    .reset_index()
    .rename(columns={"level_1": "subject_id", "level_0": "sample_type"})
    .join(subject, on="subject_id")
    .sort_values(taxon_level)[lambda x: x.sample_type == sample_type]
)

fig, ax = plt.subplots(figsize=(14, 5))
lib.plot.boxplot_with_points(
    y="transfer_index",
    x=taxon_level,
    hue=comparison,
    palette=lib.project_style.DEFAULT_COLOR_PALETTE,
    order=taxon_order,
    data=d,
    ax=ax,
    dodge=True,
    dist_kwargs={"boxprops": {"alpha": 0.5}},
)
lib.plot.rotate_xticklabels(ax, rotation=90)
ax.legend(bbox_to_anchor=(1, 1))

print(
    d[lambda x: x[taxon_level].isin(taxon_order)]
    .groupby(taxon_level)
    .apply(
        lambda d: lib.stats.mannwhitneyu(
            comparison, "transfer_index", data=d, na_action="drop"
        )
    )
    .loc[taxon_order]
)
```

```
f__
d__Bacteria;p__Firmicutes_A;c__Clostridia;o__Lachnospirales;f__Lachnospiraceae;                 (0.8333333333333334, 0.05128205128205128)
d__Bacteria;p__Firmicutes_A;c__Clostridia;o__Oscillospirales;f__Ruminococcaceae;                 (0.5714285714285714, 0.7307692307692307)
d__Bacteria;p__Bacteroidota;c__Bacteroidia;o__Bacteroidales;f__Bacteroidaceae;                  (0.42857142857142855, 0.7307692307692307)
d__Bacteria;p__Firmicutes_A;c__Clostridia;o__Oscillospirales;f__DTU089;                          (0.4166666666666667, 0.6678058142504256)
d__Bacteria;p__Firmicutes_A;c__Clostridia;o__Oscillospirales;f__Oscillospiraceae;                (0.6904761904761905, 0.2948717948717949)
d__Bacteria;p__Bacteroidota;c__Bacteroidia;o__Bacteroidales;f__Rikenellaceae;                   (0.6666666666666666, 0.34910823732597074)
d__Bacteria;p__Actinobacteriota;c__Actinobacteria;o__Actinomycetales;f__Bifidobacteriaceae;    (0.16666666666666666, 0.07934368319771502)
d__Bacteria;p__Bacteroidota;c__Bacteroidia;o__Bacteroidales;f__Tannerellaceae;                    (0.5238095238095238, 0.942095857778815)
d__Bacteria;p__Firmicutes;c__Bacilli;o__Erysipelotrichales;f__Erysipelatoclostridiaceae;                      (0.25, 0.14515427904479308)
d__Bacteria;p__Actinobacteriota;c__Coriobacteriia;o__Coriobacteriales;f__Eggerthellaceae;       (0.30952380952380953, 0.2819848280938406)
dtype: object
```

In [ ]:

```
taxon_level = "f__"
taxon_order = top_family_donor_median_abundance.index
comparison = "maintenance_"
sample_type = "followup_1"

opportunities = (
    (subject_x_strain_transfer.notna()).groupby(sotu_to_taxonomy[taxon_level]).sum()
)
transfers = (subject_x_strain_transfer).groupby(sotu_to_taxonomy[taxon_level]).sum()
no_transfers = opportunities - transfers
d = (
    (transfers / opportunities)
    .unstack()
    .rename("transfer_index")
    .reset_index()
    .rename(columns={"level_1": "subject_id", "level_0": "sample_type"})
    .join(subject, on="subject_id")
    .sort_values(taxon_level)[lambda x: x.sample_type == sample_type]
)

fig, ax = plt.subplots(figsize=(14, 5))
lib.plot.boxplot_with_points(
    y="transfer_index",
    x=taxon_level,
    hue=comparison,
    palette=lib.project_style.DEFAULT_COLOR_PALETTE,
    order=taxon_order,
    data=d,
    ax=ax,
    dodge=True,
    dist_kwargs={"boxprops": {"alpha": 0.5}},
)
lib.plot.rotate_xticklabels(ax, rotation=90)
ax.legend(bbox_to_anchor=(1, 1))

print(
    d[lambda x: x[taxon_level].isin(taxon_order)]
    .groupby(taxon_level)
    .apply(
        lambda d: lib.stats.ttest(
            comparison, "transfer_index", data=d, na_action="drop", equal_var=False
        )
    )
    .loc[taxon_order]
)
```

```
f__
d__Bacteria;p__Firmicutes_A;c__Clostridia;o__Lachnospirales;f__Lachnospiraceae;                  (-0.0441883550641489, 0.10138021371948537)
d__Bacteria;p__Firmicutes_A;c__Clostridia;o__Oscillospirales;f__Ruminococcaceae;               (-0.031729800624144135, 0.22525226946319699)
d__Bacteria;p__Bacteroidota;c__Bacteroidia;o__Bacteroidales;f__Bacteroidaceae;                  (0.017881524626337207, 0.46945960978174905)
d__Bacteria;p__Firmicutes_A;c__Clostridia;o__Oscillospirales;f__DTU089;                        (-0.0010533809246115894, 0.9656863994849314)
d__Bacteria;p__Firmicutes_A;c__Clostridia;o__Oscillospirales;f__Oscillospiraceae;                (0.013923939227668084, 0.5705670737737062)
d__Bacteria;p__Bacteroidota;c__Bacteroidia;o__Bacteroidales;f__Rikenellaceae;                    (0.012388267746704304, 0.6133146780094056)
d__Bacteria;p__Actinobacteriota;c__Actinobacteria;o__Actinomycetales;f__Bifidobacteriaceae;     (0.0012715098992452335, 0.9704275113421342)
d__Bacteria;p__Bacteroidota;c__Bacteroidia;o__Bacteroidales;f__Tannerellaceae;                   (0.004983508594510846, 0.8380367369095271)
d__Bacteria;p__Firmicutes;c__Bacilli;o__Erysipelotrichales;f__Erysipelatoclostridiaceae;        (-0.013066612523248195, 0.5948776145995525)
d__Bacteria;p__Actinobacteriota;c__Coriobacteriia;o__Coriobacteriales;f__Eggerthellaceae;        (0.028557640504266135, 0.2579896867610607)
dtype: object
```

In [ ]:

```
taxon_level = "f__"
taxon_order = top_family_donor_median_abundance.index
comparison = "donor_subject_id"
sample_type = "followup_1"
palette = subject_color_palette

opportunities = (
    (subject_x_strain_transfer.notna()).groupby(sotu_to_taxonomy[taxon_level]).sum()
)
transfers = (subject_x_strain_transfer).groupby(sotu_to_taxonomy[taxon_level]).sum()
no_transfers = opportunities - transfers
d = (
    (transfers / opportunities)
    .unstack()
    .rename("transfer_index")
    .reset_index()
    .rename(columns={"level_1": "subject_id", "level_0": "sample_type"})
    .join(subject, on="subject_id")
    .sort_values(taxon_level)[lambda x: x.sample_type == sample_type]
)

fig, ax = plt.subplots(figsize=(14, 5))
lib.plot.boxplot_with_points(
    y="transfer_index",
    x=taxon_level,
    hue=comparison,
    palette=palette,
    order=taxon_order,
    data=d,
    ax=ax,
    dodge=True,
    dist_kwargs={"boxprops": {"alpha": 0.5}},
)
lib.plot.rotate_xticklabels(ax, rotation=90)
ax.legend(bbox_to_anchor=(1, 1))

print(
    d[lambda x: x[taxon_level].isin(taxon_order)]
    .groupby(taxon_level)
    .apply(
        lambda d: lib.stats.ttest(
            comparison,
            "transfer_index",
            data=d[d.donor_subject_id.isin(["D0044", "D0097"])],
            na_action="drop",
            equal_var=False,
        )
    )
    .loc[taxon_order]
)
```

```
f__
d__Bacteria;p__Firmicutes_A;c__Clostridia;o__Lachnospirales;f__Lachnospiraceae;                (-0.1275591623440232, 0.010191198120217623)
d__Bacteria;p__Firmicutes_A;c__Clostridia;o__Oscillospirales;f__Ruminococcaceae;               (-0.002011356334610464, 0.9533788725422642)
d__Bacteria;p__Bacteroidota;c__Bacteroidia;o__Bacteroidales;f__Bacteroidaceae;                   (0.03951030159849838, 0.2674118772211199)
d__Bacteria;p__Firmicutes_A;c__Clostridia;o__Oscillospirales;f__DTU089;                          (-0.02348700355352938, 0.510081599124989)
d__Bacteria;p__Firmicutes_A;c__Clostridia;o__Oscillospirales;f__Oscillospiraceae;                (0.01875557505811198, 0.5892931811533324)
d__Bacteria;p__Bacteroidota;c__Bacteroidia;o__Bacteroidales;f__Rikenellaceae;                  (-0.0028656487468788922, 0.934527887007324)
d__Bacteria;p__Actinobacteriota;c__Actinobacteria;o__Actinomycetales;f__Bifidobacteriaceae;      (0.008960527611399104, 0.794650728972796)
d__Bacteria;p__Bacteroidota;c__Bacteroidia;o__Bacteroidales;f__Tannerellaceae;                  (-0.0388915510038534, 0.27445438916594683)
d__Bacteria;p__Firmicutes;c__Bacilli;o__Erysipelotrichales;f__Erysipelatoclostridiaceae;         (-0.0261999241251296, 0.4524509366586349)
d__Bacteria;p__Actinobacteriota;c__Coriobacteriia;o__Coriobacteriales;f__Eggerthellaceae;      (0.043349374112670494, 0.23476677219095773)
dtype: object
```

In [ ]:

```
taxon_level = "f__"
taxon_order = top_family_donor_median_abundance.index
comparison = "responder"
sample_type = "followup_1"

opportunities = (
    (subject_x_strain_transfer.notna()).groupby(sotu_to_taxonomy[taxon_level]).sum()
)
transfers = (subject_x_strain_transfer).groupby(sotu_to_taxonomy[taxon_level]).sum()
no_transfers = opportunities - transfers
d = (
    (transfers / opportunities)
    .unstack()
    .rename("transfer_index")
    .reset_index()
    .rename(columns={"level_1": "subject_id", "level_0": "sample_type"})
    .join(subject, on="subject_id")
    .sort_values(taxon_level)[lambda x: x.sample_type == sample_type]
)

fig, ax = plt.subplots(figsize=(14, 5))
lib.plot.boxplot_with_points(
    y="transfer_index",
    x=taxon_level,
    hue=comparison,
    palette=lib.project_style.DEFAULT_COLOR_PALETTE,
    order=taxon_order,
    data=d,
    ax=ax,
    dodge=True,
    dist_kwargs={"boxprops": {"alpha": 0.5}},
)
lib.plot.rotate_xticklabels(ax, rotation=90)
ax.legend(bbox_to_anchor=(1, 1))

print(
    d[lambda x: x[taxon_level].isin(taxon_order)]
    .groupby(taxon_level)
    .apply(
        lambda d: lib.stats.ttest(
            comparison, "transfer_index", data=d, na_action="drop", equal_var=False
        )
    )
    .loc[taxon_order]
)
```

```
f__
d__Bacteria;p__Firmicutes_A;c__Clostridia;o__Lachnospirales;f__Lachnospiraceae;                (-0.018556263305716313, 0.45225212983551866)
d__Bacteria;p__Firmicutes_A;c__Clostridia;o__Oscillospirales;f__Ruminococcaceae;               (-0.08682611970398871, 0.004214813531685746)
d__Bacteria;p__Bacteroidota;c__Bacteroidia;o__Bacteroidales;f__Bacteroidaceae;                   (-0.02276136172370172, 0.3606358960860395)
d__Bacteria;p__Firmicutes_A;c__Clostridia;o__Oscillospirales;f__DTU089;                          (0.004253019537100326, 0.8614881852258324)
d__Bacteria;p__Firmicutes_A;c__Clostridia;o__Oscillospirales;f__Oscillospiraceae;                (0.015879353205430376, 0.5217057200527233)
d__Bacteria;p__Bacteroidota;c__Bacteroidia;o__Bacteroidales;f__Rikenellaceae;                  (-0.029665783813836426, 0.24009195219157117)
d__Bacteria;p__Actinobacteriota;c__Actinobacteria;o__Actinomycetales;f__Bifidobacteriaceae;     (-0.018695077529913593, 0.5911526052524638)
d__Bacteria;p__Bacteroidota;c__Bacteroidia;o__Bacteroidales;f__Tannerellaceae;                  (0.037261710708708644, 0.15170187764331558)
d__Bacteria;p__Firmicutes;c__Bacilli;o__Erysipelotrichales;f__Erysipelatoclostridiaceae;       (-0.017287466567304392, 0.48444368936405235)
d__Bacteria;p__Actinobacteriota;c__Coriobacteriia;o__Coriobacteriales;f__Eggerthellaceae;       (-0.03498509479025929, 0.17080428723573063)
dtype: object
```

#### Subsection: Broad correlations between datasets¶

In [ ]:

```
import skbio as skb

m = (
    sample.join(subject, on="subject_id", lsuffix="_")
    .assign(subject_type=lambda x: x.recipient.map({True: "recipient", False: "donor"}))
    .assign(zorder=lambda x: x.recipient.map({True: 0, False: 1}))
)

d = sample_x_rotu_cvrg.rename(columns=lambda s: s.replace("_", " "))

tree = skb.io.read(
    "sraw/2020-12-30-Dose_Finding_Study_Box_mirror/16S data files/dada2_optim_tree_OTUtips_FMT_VED_May2019.tre",
    into=skb.TreeNode,
).root_at_midpoint()


motu_unifrac_dmat = skb.diversity.beta_diversity(
    "weighted_unifrac",
    counts=d,
    ids=d.index,
    otu_ids=d.columns,
    tree=tree,
    normalized=True,
).to_data_frame()
```

In [ ]:

```
# TODO: Only calculate for reasonable data subset

datasets = (
    ("subject", sample[["subject_id"]], lambda x, y: x != y),
    ("time", sample[["days_post_fmt"]], "cityblock"),
    ("rotu", sample_x_rotu, "braycurtis"),
    ("motu", sample_x_motu, "braycurtis"),
    ("motu_bact", sample_x_motu_bacteroidota, "braycurtis"),
    ("motu_firm_A", sample_x_motu_firmicutes_A, "braycurtis"),
    ("motu_firm", sample_x_motu_firmicutes, "braycurtis"),
    ("motu_firm_C", sample_x_motu_firmicutes_C, "braycurtis"),
    ("motu_actino", sample_x_motu_actinobacteria, "braycurtis"),
    ("motu_proteo", sample_x_motu_proteobacteria, "braycurtis"),
    ("sotu_bact", sample_x_sotu_bacteroidota, "braycurtis"),
    ("sotu_firm_A", sample_x_sotu_firmicutes_A, "braycurtis"),
    ("sotu_firm_C", sample_x_sotu_firmicutes_C, "braycurtis"),
    ("sotu_firm", sample_x_sotu_firmicutes, "braycurtis"),
    ("sotu_actino", sample_x_sotu_actinobacteria, "braycurtis"),
    ("sotu_proteo", sample_x_sotu_proteobacteria, "braycurtis"),
    ("sotu", sample_x_sotu, "braycurtis"),
    ("family", sample_x_family, "braycurtis"),
    ("family_bact", sample_x_family_bacteroidota, "braycurtis"),
    ("family_firm_A", sample_x_family_firmicutes_A, "braycurtis"),
    ("family_firm", sample_x_family_firmicutes, "braycurtis"),
    ("family_firm_C", sample_x_family_firmicutes_C, "braycurtis"),
    ("family_actino", sample_x_family_actinobacteria, "braycurtis"),
    ("family_proteo", sample_x_family_proteobacteria, "braycurtis"),
    #     ('eggnog', sample_x_eggnog_cvrg, 'cosine'),
    ("ko", sample_x_ko_cvrg, "cosine"),
    #     ('keggmodule', sample_x_keggmodule_cvrg, 'cosine'),
    #     ('chem', sample_x_chem_std2, 'cosine'),
    ("chem_ba", sample_x_chem_ba_std2, "cosine"),
)

dmatrices = {}
for dataset_name, _data, metric in tqdm(datasets, ascii=True):
    dmatrices[dataset_name] = dmatrix(_data, metric=metric)

dmatrices["sqrt_time"] = np.sqrt(dmatrices["time"])
dmatrices["log_time"] = np.log(dmatrices["time"])
dmatrices["cbrt_time"] = np.cbrt(dmatrices["time"])
dmatrices["rotu_unifrac"] = motu_unifrac_dmat
```

```
100%|########################################################################################################################| 26/26 [00:01<00:00, 19.24it/s]
```

In [ ]:

```
nrows, ncols = 9, 3
fig, axs = plt.subplots(nrows=nrows, ncols=ncols, figsize=(ncols * 5, nrows * 5))

for dataset_name, ax in zip(dmatrices, axs.flatten()):
    _time, _subject, _x = align_dmatrices(
        dmatrices["time"], dmatrices["subject"], dmatrices[dataset_name]
    )
    if dataset_name in ["time", "sqrt_time", "log_time", "cbrt_time"]:
        vmax = None
    else:
        vmax = 1
    sns.heatmap(
        data=_x,
        mask=_subject.astype(bool),
        cmap="magma",
        cbar=False,
        ax=ax,
        xticklabels=0,
        yticklabels=0,
        vmin=0,
        vmax=vmax,
    )
    ax.set_facecolor("grey")
    ax.set_ylabel("")
    ax.set_xlabel("")
    ax.set_title(dataset_name)
```

In [ ]:

```
drop = idxwhere(
    sample.sample_type.isin(
        [
            "post_antibiotic",
            "donor_mean",
            "donor_initial",
            "donor_capsule",
            "donor_enema",
        ]
    )
)

for xname, yname in [
    #     ('eggnog', 'time'),
    #     ('eggnog', 'sqrt_time'),
    #     ('eggnog', 'cbrt_time'),
    #     ('eggnog', 'log_time'),
    ("ko", "time"),
    ("ko", "sqrt_time"),
    ("ko", "cbrt_time"),
    ("ko", "log_time"),
    #     ('keggmodule', 'time'),
    #     ('keggmodule', 'sqrt_time'),
    #     ('keggmodule', 'cbrt_time'),
    #     ('keggmodule', 'log_time'),
    ("sotu", "time"),
    ("sotu", "sqrt_time"),
    ("sotu", "cbrt_time"),
    ("sotu", "log_time"),
    ("motu", "time"),
    ("motu", "sqrt_time"),
    ("motu", "cbrt_time"),
    ("motu", "log_time"),
    ("rotu", "time"),
    ("rotu", "sqrt_time"),
    ("rotu", "cbrt_time"),
    ("rotu", "log_time"),
    ("family", "time"),
    ("family", "sqrt_time"),
    ("family", "cbrt_time"),
    ("family", "log_time"),
    #     ('chem', 'time'),
    #     ('chem', 'sqrt_time'),
    #     ('chem', 'cbrt_time'),
    #     ('chem', 'log_time'),
    ("chem_ba", "time"),
    ("chem_ba", "sqrt_time"),
    ("chem_ba", "cbrt_time"),
    ("chem_ba", "log_time"),
]:
    t, m, x, y = align_dmatrices(
        dmatrices["time"],
        dmatrices["subject"],
        dmatrices[xname],
        dmatrices[yname],
        drop_labels=drop,
    )
    t, x, y = [mask_dmatrix(d, m.astype(bool)) for d in [t, x, y]]
    s = sample.subject_id.loc[m.index]
    print(
        xname, yname, *mantel_test(x, y, strata=s, permutations=999, method="pearson")
    )
```

```
ko time 0.155226166677554 0.014
ko sqrt_time 0.16638781039562173 0.01
ko cbrt_time 0.16822947860585036 0.008
ko log_time 0.16856134780441823 0.003
sotu time 0.2879527761385169 0.001
sotu sqrt_time 0.33262023377638555 0.001
sotu cbrt_time 0.34552316259172045 0.001
sotu log_time 0.3659097351434171 0.001
motu time 0.2435144357294096 0.001
motu sqrt_time 0.2772146281019003 0.001
motu cbrt_time 0.28690999480614593 0.001
motu log_time 0.3021864209875207 0.001
rotu time 0.2907227225790046 0.001
rotu sqrt_time 0.3297764016531522 0.001
rotu cbrt_time 0.3422005183094123 0.001
rotu log_time 0.36305251525228666 0.001
family time 0.1401223377703447 0.007
family sqrt_time 0.1614155811150649 0.004
family cbrt_time 0.16726683760358835 0.002
family log_time 0.17603195328981672 0.001
chem_ba time 0.1724687585393343 0.014
chem_ba sqrt_time 0.18262410454246497 0.009
chem_ba cbrt_time 0.1854607811941613 0.005
chem_ba log_time 0.18924717372039265 0.004
```

In [ ]:

```
drop = idxwhere(
    sample.sample_type.isin(
        [
            "post_antibiotic",
            "donor_mean",
            "donor_initial",
            "donor_capsule",
            "donor_enema",
        ]
    )
)

for xname, yname, zname in [
    #     ('eggnog', 'sotu', 'sqrt_time'),
    #     ('eggnog', 'motu', 'sqrt_time'),
    #     ('eggnog', 'rotu', 'sqrt_time'),
    #     ('eggnog', 'sotu', 'motu'),
    ("ko", "sotu", "sqrt_time"),
    ("ko", "motu", "sqrt_time"),
    ("ko", "rotu", "sqrt_time"),
    ("ko", "sotu", "motu"),
    #     ('chem', 'sotu', 'sqrt_time'),
    #     ('chem', 'motu', 'sqrt_time'),
    #     ('chem', 'rotu', 'sqrt_time'),
    #     ('chem', 'ko', 'sqrt_time'),
    ("chem_ba", "family", "sqrt_time"),
    ("chem_ba", "family_bact", "sqrt_time"),
    ("chem_ba", "family_firm", "sqrt_time"),
    ("chem_ba", "family_firm_A", "sqrt_time"),
    ("chem_ba", "family_firm_C", "sqrt_time"),
    ("chem_ba", "family_actino", "sqrt_time"),
    ("chem_ba", "family_proteo", "sqrt_time"),
    ("chem_ba", "motu", "sqrt_time"),
    ("chem_ba", "motu_bact", "sqrt_time"),
    ("chem_ba", "motu_firm", "sqrt_time"),
    ("chem_ba", "motu_firm_A", "sqrt_time"),
    ("chem_ba", "motu_firm_C", "sqrt_time"),
    ("chem_ba", "motu_actino", "sqrt_time"),
    ("chem_ba", "motu_proteo", "sqrt_time"),
    ("chem_ba", "sotu", "sqrt_time"),
    ("chem_ba", "sotu_bact", "sqrt_time"),
    ("chem_ba", "sotu_firm", "sqrt_time"),
    ("chem_ba", "sotu_firm_A", "sqrt_time"),
    ("chem_ba", "sotu_firm_C", "sqrt_time"),
    ("chem_ba", "sotu_actino", "sqrt_time"),
    ("chem_ba", "sotu_proteo", "sqrt_time"),
    ("chem_ba", "sotu", "motu"),
    ("chem_ba", "sotu", "rotu"),
    ("chem_ba", "sotu", "ko"),
    ("chem_ba", "sotu_firm", "motu_firm"),
    ("chem_ba", "sotu_firm_A", "motu_firm_A"),
    ("chem_ba", "sotu_firm_C", "motu_firm_C"),
    ("chem_ba", "sotu_actino", "motu_actino"),
    ("chem_ba", "sotu_bact", "motu_bact"),
    ("chem_ba", "ko", "sqrt_time"),
    ("chem_ba", "ko", "sqrt_time"),
    ("chem_ba", "rotu", "sqrt_time"),
    ("chem_ba", "family", "sqrt_time"),
    ("chem_ba", "rotu", "sotu"),
    ("chem_ba", "rotu", "motu"),
]:
    t, m, x, y, z = align_dmatrices(
        dmatrices["time"],
        dmatrices["subject"],
        dmatrices[xname],
        dmatrices[yname],
        dmatrices[zname],
        drop_labels=drop,
    )
    t, x, y, z = [mask_dmatrix(d, m.astype(bool)) for d in [t, x, y, z]]
    s = sample.subject_id.loc[m.index]
    print(
        xname,
        yname,
        zname,
        *partial_mantel_test(x, y, z, strata=s, permutations=999, method="pearson"),
    )
```

```
ko sotu sqrt_time 0.5619669102291576 0.001
ko motu sqrt_time 0.7249648496501688 0.001
ko rotu sqrt_time 0.4442590436857634 0.001
ko sotu motu -0.2614348321514883 1.0
chem_ba family sqrt_time 0.18013627426520712 0.001
chem_ba family_bact sqrt_time 0.1888176989126553 0.017
chem_ba family_firm sqrt_time -0.02641651888241617 0.329
chem_ba family_firm_A sqrt_time 0.1389311528491224 0.013
chem_ba family_firm_C sqrt_time 0.2238514921124214 0.003
chem_ba family_actino sqrt_time -0.020544766890465666 0.097
chem_ba family_proteo sqrt_time 0.006999701695863209 0.186
chem_ba motu sqrt_time 0.29086684050610595 0.001
chem_ba motu_bact sqrt_time 0.27539810127829073 0.001
chem_ba motu_firm sqrt_time -0.03670518579333471 0.527
chem_ba motu_firm_A sqrt_time 0.33109330136627024 0.001
chem_ba motu_firm_C sqrt_time 0.25753300752918545 0.001
chem_ba motu_actino sqrt_time 0.07463254572004246 0.045
chem_ba motu_proteo sqrt_time 0.014428652391759892 0.025
chem_ba sotu sqrt_time 0.27929820262502847 0.001
chem_ba sotu_bact sqrt_time 0.23234949916188796 0.001
chem_ba sotu_firm sqrt_time 0.03089664765748308 0.326
chem_ba sotu_firm_A sqrt_time 0.3083271670864226 0.001
chem_ba sotu_firm_C sqrt_time 0.21515765881686566 0.003
chem_ba sotu_actino sqrt_time 0.12584093503958768 0.009
chem_ba sotu_proteo sqrt_time -0.02924800396588947 0.005
chem_ba sotu motu 0.06949482138730546 0.117
chem_ba sotu rotu 0.0819701285448154 0.135
chem_ba sotu ko 0.31204282480940115 0.001
chem_ba sotu_firm motu_firm 0.10640665196691391 0.143
chem_ba sotu_firm_A motu_firm_A 0.05813796189656106 0.158
chem_ba sotu_firm_C motu_firm_C 0.05407548195091993 0.065
chem_ba sotu_actino motu_actino 0.12498535866331952 0.034
chem_ba sotu_bact motu_bact 0.05471102071291598 0.077
chem_ba ko sqrt_time 0.0906291965905439 0.031
chem_ba ko sqrt_time 0.0906291965905439 0.059
chem_ba rotu sqrt_time 0.30572326620271684 0.001
chem_ba family sqrt_time 0.18013627426520712 0.001
chem_ba rotu sotu 0.16112798051648866 0.003
chem_ba rotu motu 0.14157742479419116 0.005
```

In [ ]:

```
drop = idxwhere(
    sample.sample_type.isin(
        [
            "post_antibiotic",
            "donor_mean",
            "donor_initial",
            "donor_capsule",
            "donor_enema",
        ]
    )
)

for xname, yname in [
    #     ('eggnog', 'sqrt_time'),
    #     ('eggnog', 'sotu'),
    #     ('eggnog', 'motu'),
    #     ('eggnog', 'rotu'),
    #     ('eggnog', 'family'),
    ("ko", "sqrt_time"),
    ("ko", "sotu"),
    ("ko", "motu"),
    ("ko", "rotu"),
    ("ko", "family"),
    ("ko", "sqrt_time"),
    ("ko", "sotu"),
    ("ko", "motu"),
    ("ko", "rotu"),
    ("ko", "family"),
    #     ('chem', 'sqrt_time'),
    #     ('chem', 'sotu'),
    #     ('chem', 'motu'),
    #     ('chem', 'rotu'),
    #     ('chem', 'family'),
    #     ('chem', 'eggnog'),
    #     ('chem', 'ko'),
    ("chem_ba", "sqrt_time"),
    ("chem_ba", "family"),
    ("chem_ba", "family_bact"),
    ("chem_ba", "family_firm"),
    ("chem_ba", "family_firm_A"),
    ("chem_ba", "family_firm_C"),
    ("chem_ba", "family_actino"),
    ("chem_ba", "family_proteo"),
    #     ('chem_ba', 'eggnog'),
    ("chem_ba", "ko"),
    ("chem_ba", "motu"),
    ("chem_ba", "motu_bact"),
    ("chem_ba", "motu_firm"),
    ("chem_ba", "motu_firm_A"),
    ("chem_ba", "motu_firm_C"),
    ("chem_ba", "motu_actino"),
    ("chem_ba", "motu_proteo"),
    ("chem_ba", "sotu"),
    ("chem_ba", "sotu_bact"),
    ("chem_ba", "sotu_firm"),
    ("chem_ba", "sotu_firm_A"),
    ("chem_ba", "sotu_firm_C"),
    ("chem_ba", "sotu_actino"),
    ("chem_ba", "sotu_proteo"),
]:
    t, m, x, y = align_dmatrices(
        dmatrices["time"],
        dmatrices["subject"],
        dmatrices[xname],
        dmatrices[yname],
        drop_labels=drop,
    )
    t, x, y = [mask_dmatrix(d, m.astype(bool)) for d in [t, x, y]]
    s = sample.subject_id.loc[m.index]
    print(
        xname, yname, *mantel_test(x, y, strata=s, permutations=999, method="spearman")
    )
```

```
ko sqrt_time 0.1743185030096906 0.003
ko sotu 0.6397464423950604 0.001
ko motu 0.7917612516527884 0.001
ko rotu 0.5631032806962835 0.001
ko family 0.8585165326549447 0.001
ko sqrt_time 0.1743185030096906 0.004
ko sotu 0.6397464423950604 0.001
ko motu 0.7917612516527884 0.001
ko rotu 0.5631032806962835 0.001
ko family 0.8585165326549447 0.001
chem_ba sqrt_time 0.18718179613569222 0.001
chem_ba family 0.24262043299202782 0.001
chem_ba family_bact 0.11932004171423047 0.011
chem_ba family_firm 0.035323043790380475 0.099
chem_ba family_firm_A 0.153923565194393 0.001
chem_ba family_firm_C 0.21736158701811834 0.01
chem_ba family_actino -0.020338346825213818 0.144
chem_ba family_proteo 0.0483247199318189 0.046
chem_ba ko 0.19704367703177544 0.001
chem_ba motu 0.3293988740695961 0.001
chem_ba motu_bact 0.26629136813467413 0.001
chem_ba motu_firm 0.04131107051338199 0.143
chem_ba motu_firm_A 0.3202539010843692 0.001
chem_ba motu_firm_C 0.26893146446697586 0.004
chem_ba motu_actino 0.11931839649374694 0.009
chem_ba motu_proteo 0.05259088774356658 0.006
chem_ba sotu 0.3037114142139268 0.001
chem_ba sotu_bact 0.23358451471408367 0.001
chem_ba sotu_firm 0.0659142753814146 0.141
chem_ba sotu_firm_A 0.32312432610628444 0.001
chem_ba sotu_firm_C 0.23192815570299477 0.001
chem_ba sotu_actino 0.1625958395348608 0.002
chem_ba sotu_proteo 0.03498190050363374 0.001
```

In [ ]:

```
from itertools import combinations

drop = idxwhere(
    sample.sample_type.isin(
        [
            "post_antibiotic",
            "donor_mean",
            "donor_initial",
            "donor_capsule",
            "donor_enema",
        ]
    )
)


dmats_to_compare = [
    "rotu",
    "family",
    "rotu_unifrac",
    "motu",  #'motu_bact', 'motu_firm', 'motu_actino', 'motu_proteo',
    "sotu",  #'sotu_bact', 'sotu_firm', 'sotu_actino', 'sotu_proteo',
    #     'eggnog',
    "ko",
    #     'keggmodule',
    #     'chem',
    "chem_ba",
]

mantel_results = []
mantel_partial_results = []
for xname, yname in tqdm(list(combinations(dmats_to_compare, r=2)), ascii=True):
    zname = "sqrt_time"

    t, m, x, y, z = align_dmatrices(
        dmatrices["time"],
        dmatrices["subject"],
        dmatrices[xname],
        dmatrices[yname],
        dmatrices[zname],
        drop_labels=drop,
    )
    t, x, y, z = [mask_dmatrix(d, m.astype(bool)) for d in [t, x, y, z]]
    s = sample.subject_id.loc[m.index]
    mantel_partial_results.append(
        (
            xname,
            yname,
            *partial_mantel_test(x, y, z, strata=s, permutations=999, method="pearson"),
        )
    )
    mantel_results.append(
        (xname, yname, *mantel_test(x, y, strata=s, permutations=999, method="pearson"))
    )

mantel_partial_results = pd.DataFrame(
    mantel_partial_results, columns=["X", "Y", "r", "p"]
).set_index(["X", "Y"])
mantel_results = pd.DataFrame(mantel_results, columns=["X", "Y", "r", "p"]).set_index(
    ["X", "Y"]
)

mantel_partial_r_matrix = (
    mantel_partial_results["r"]
    .unstack()
    .reindex(index=dmats_to_compare, columns=dmats_to_compare)
)
mantel_partial_p_matrix = (
    mantel_partial_results["p"]
    .unstack()
    .reindex(index=dmats_to_compare, columns=dmats_to_compare)
)

mantel_r_matrix = (
    mantel_results["r"]
    .unstack()
    .reindex(index=dmats_to_compare, columns=dmats_to_compare)
)
mantel_p_matrix = (
    mantel_results["p"]
    .unstack()
    .reindex(index=dmats_to_compare, columns=dmats_to_compare)
)
```

```
100%|########################################################################################################################| 21/21 [00:24<00:00,  1.15s/it]
```

In [ ]:

```
# dist = squareform(1 - d)
# linkage = scipy.cluster.hierarchy.linkage(y=dist, method='single', optimal_ordering=True)
order = [
    #     'motu_proteo', 'sotu_proteo',
    #     'motu_actino', 'sotu_actino',
    #     'motu_bact', 'sotu_bact',
    #     'motu_firm', 'sotu_firm',
    #    'rotu',
    "family",
    "rotu",
    "rotu_unifrac",
    "motu",
    "sotu",
    #     'eggnog',
    "ko",
    #     'keggmodule',
    #     'chem',
    "chem_ba",
]

d = mantel_r_matrix.fillna(mantel_partial_r_matrix.T).fillna(1)
p = mantel_p_matrix.fillna(mantel_partial_p_matrix.T)

d = d.loc[order, order]
p = p.loc[order, order]

sns.heatmap(
    d,
    annot=p.applymap(lambda x: {True: "ø", False: ""}[x > 0.05]),
    fmt="",
    vmin=0,
    vmax=1,
)
```

Out[ ]:

```
<AxesSubplot:xlabel='Y', ylabel='X'>
```

In [ ]:

```
from itertools import combinations, product


def make_xy_coord_tables(d):
    return (
        pd.DataFrame(
            np.asarray([range(d.shape[1]) for _ in range(d.shape[0])]),
            index=d.index,
            columns=d.columns,
        ),
        pd.DataFrame(
            np.asarray([range(d.shape[0]) for _ in range(d.shape[1])]).T,
            index=d.index,
            columns=d.columns,
        ),
    )
```

In [ ]:

```
drop = idxwhere(
    sample.sample_type.isin(
        [
            "post_antibiotic",
            "donor_mean",
            "donor_initial",
            "donor_capsule",
            "donor_enema",
        ]
    )
)

xlist = ["ko", "chem_ba"]
ylist = ["ko", "family", "motu", "sotu", "rotu"]
mantel_results_overall = []
for xname, yname in tqdm(list(product(xlist, ylist)), ascii=True):
    zname = "sqrt_time"
    t, m, x, y, z = align_dmatrices(
        dmatrices["time"],
        dmatrices["subject"],
        dmatrices[xname],
        dmatrices[yname],
        dmatrices[zname],
        drop_labels=drop,
    )
    t, x, y, z = [mask_dmatrix(d, m.astype(bool)) for d in [t, x, y, z]]
    s = sample.subject_id.loc[m.index]
    mantel_results_overall.append(
        (
            xname,
            yname,
            *partial_mantel_test(
                x, y, z, strata=s, permutations=9999, method="pearson"
            ),
        )
    )

mantel_results_overall = pd.DataFrame(
    mantel_results_overall, columns=["xname", "yname", "r", "p"]
).set_index(["xname", "yname"])
mantel_results_overall_r = mantel_results_overall["r"].unstack().loc[xlist, ylist]
mantel_results_overall_p = mantel_results_overall["p"].unstack().loc[xlist, ylist]
```

```
100%|########################################################################################################################| 10/10 [00:45<00:00,  4.58s/it]
```

In [ ]:

```
rr = mantel_results_overall_r
pp = mantel_results_overall_p
xx, yy = make_xy_coord_tables(rr)

fig, ax = plt.subplots(figsize=(5, 2.5))
plot = sns.heatmap(
    rr,
    fmt="",
    vmin=0,
    vmax=1,
    ax=ax,
    cbar=False,
)


for _, (p, x, y) in pd.DataFrame(
    dict(p=pp.unstack(), x=xx.unstack(), y=yy.unstack())
).iterrows():
    if p > 0.05:
        ax.add_patch(
            mpl.patches.Rectangle(
                (x, y), 1, 1, hatch="//", edgecolor="lightgrey", linewidth=0, fill=False
            )
        )
```

In [ ]:

```
rr = mantel_results_overall_r
pp = mantel_results_overall_p
xx, yy = make_xy_coord_tables(rr)

fig, ax = plt.subplots(figsize=(5, 2.5))
plot = sns.heatmap(
    rr,
    fmt="",
    vmin=0,
    vmax=1,
    ax=ax,
    cbar=False,
)

for _, (p, x, y) in pd.DataFrame(
    dict(p=pp.unstack(), x=xx.unstack(), y=yy.unstack())
).iterrows():
    if p > 0.05:
        ax.add_patch(
            mpl.patches.Rectangle(
                (x, y), 1, 1, hatch="//", edgecolor="lightgrey", linewidth=0, fill=False
            )
        )

# Remove chrome
ax.set_xticks([])
ax.set_xlabel(None)
ax.set_yticks([])
ax.set_ylabel(None)

fig.savefig("fig/mantel_multiomics_heatmap_nolabels.pdf", bbox_inches="tight")

# Color-bar
fig, ax = plt.subplots(figsize=(0.5, 3))
sns.heatmap(
    [[]],
    vmin=0,
    vmax=1,
    ax=ax,
    cbar=True,
    cbar_kws={"fraction": 1.0, "label": "Correlation"},
)
for spines in ["top", "right", "bottom", "left"]:
    ax.spines[spines].set_visible(False)
ax.set_xticks([])
ax.set_yticks([])
fig.savefig("fig/heatmap_colorbar.pdf", bbox_inches="tight")
```

```
/Users/byronsmith/anaconda3/envs/ucfmt2/lib/python3.9/site-packages/seaborn/matrix.py:305: UserWarning: Attempting to set identical left == right == 0 results in singular transformations; automatically expanding.
  ax.set(xlim=(0, self.data.shape[1]), ylim=(0, self.data.shape[0]))
```

In [ ]:

```
print(mantel_results_overall_r)
print(mantel_results_overall_p)
```

```
yname          ko    family      motu      sotu      rotu
xname                                                    
ko       1.000000  0.832267  0.724965  0.561967  0.444259
chem_ba  0.090629  0.180136  0.290867  0.279298  0.305723
yname        ko  family    motu    sotu    rotu
xname                                          
ko       0.0001  0.0001  0.0001  0.0001  0.0001
chem_ba  0.0416  0.0001  0.0001  0.0001  0.0001
```

In [ ]:

```
from itertools import combinations, product

drop = idxwhere(
    sample.sample_type.isin(
        [
            "post_antibiotic",
            "donor_mean",
            "donor_initial",
            "donor_capsule",
            "donor_enema",
        ]
    )
)

mantel_results_split = []
for tax_level, fraction in tqdm(
    list(
        product(
            ["family", "motu", "sotu"],
            ["", "_bact", "_firm", "_firm_A", "_firm_C", "_actino", "_proteo"],
        )
    ),
    ascii=True,
):
    zname = "sqrt_time"
    xname = "chem_ba"
    yname = tax_level + fraction
    t, m, x, y, z = align_dmatrices(
        dmatrices["time"],
        dmatrices["subject"],
        dmatrices[xname],
        dmatrices[yname],
        dmatrices[zname],
        drop_labels=drop,
    )
    t, x, y, z = [mask_dmatrix(d, m.astype(bool)) for d in [t, x, y, z]]
    s = sample.subject_id.loc[m.index]
    mantel_results_split.append(
        (
            tax_level,
            fraction,
            *partial_mantel_test(
                x, y, z, strata=s, permutations=9999, method="pearson"
            ),
        )
    )

mantel_results_split = pd.DataFrame(
    mantel_results_split, columns=["tax_level", "fraction", "r", "p"]
).set_index(["fraction", "tax_level"])
mantel_results_split_r = mantel_results_split["r"].unstack()
mantel_results_split_p = mantel_results_split["p"].unstack()
```

```
100%|########################################################################################################################| 21/21 [01:55<00:00,  5.50s/it]
```

In [ ]:

```
rr = mantel_results_split_r
pp = mantel_results_split_p
xx, yy = make_xy_coord_tables(rr)

fig, ax = plt.subplots(figsize=(3, 3))
plot = sns.heatmap(
    rr,
    #     annot=pp.applymap(lambda x: {True: '.', False: ''}[x > 0.05]),
    fmt="",
    vmin=0,
    vmax=1,
    ax=ax,
    cbar=False,
)


for _, (p, x, y) in pd.DataFrame(
    dict(p=pp.unstack(), x=xx.unstack(), y=yy.unstack())
).iterrows():
    if p > 0.05:
        ax.add_patch(
            mpl.patches.Rectangle(
                (x, y), 1, 1, hatch="//", edgecolor="lightgrey", linewidth=0, fill=False
            )
        )
```

In [ ]:

```
rr = mantel_results_split_r
pp = mantel_results_split_p
xx, yy = make_xy_coord_tables(rr)

fig, ax = plt.subplots(figsize=(3, 3))
plot = sns.heatmap(
    rr,
    #     annot=pp.applymap(lambda x: {True: '.', False: ''}[x > 0.05]),
    fmt="",
    vmin=0,
    vmax=1,
    ax=ax,
    cbar=False,
)
# Remove chrome
ax.set_xticks([])
ax.set_xlabel(None)
ax.set_yticks([])
ax.set_ylabel(None)

for _, (p, x, y) in pd.DataFrame(
    dict(p=pp.unstack(), x=xx.unstack(), y=yy.unstack())
).iterrows():
    if p > 0.05:
        ax.add_patch(
            mpl.patches.Rectangle(
                (x, y), 1, 1, hatch="//", edgecolor="lightgrey", linewidth=0, fill=False
            )
        )

fig.savefig("fig/mantel_taxonomic_bileacids_heatmap_nolabels.pdf", bbox_inches="tight")
```

In [ ]:

```
print(mantel_results_split_r)
print("----------")
print(mantel_results_split_p)
```

```
tax_level    family      motu      sotu
fraction                               
           0.180136  0.290867  0.279298
_actino   -0.020545  0.074633  0.125841
_bact      0.188818  0.275398  0.232349
_firm     -0.026417 -0.036705  0.030897
_firm_A    0.138931  0.331093  0.308327
_firm_C    0.223851  0.257533  0.215158
_proteo    0.007000  0.014429 -0.029248
----------
tax_level  family    motu    sotu
fraction                         
           0.0001  0.0001  0.0001
_actino    0.1129  0.0468  0.0101
_bact      0.0105  0.0001  0.0004
_firm      0.3061  0.5353  0.3249
_firm_A    0.0154  0.0001  0.0002
_firm_C    0.0047  0.0024  0.0022
_proteo    0.1903  0.0270  0.0035
```
