## Supplementary material for "Strain-resolved analysis in a randomized trial of antibiotic pretreatment and maintenance dose delivery mode with fecal microbiota transplant for ulcerative colitis": Self Sampling Instructions

**Fecal Microbiota Transplant for Treatment for Ulcerative Colitis**
**Stool sample collection instructions**

*You are receiving this sample collection pack because you have indicated your willingness to participate in our study investigating the long-term outcomes of patients receiving treatment for Ulcerative Colitis. To ensure that we can use your sample for our studies, please make sure you have completed and signed a study consent.*
***Thank you for your participation in our study.***

**Kit content:**
1 Paper stool sample collector
1 Stool sample collection tube
1 Biohazard bag
1 Ziploc bag
1 Insulated envelope
1 Ice pack
1 Pre-paid mailer

### General instructions

1. Place the ice pack provided in your freezer ***at least one day prior*** to sample collection.
2. Collect a sample of the ***first*** bowel movement of the day.
3. Collect and send samples ***only*** from Monday to Wednesday.

### Stool sample collection

1. Stick the paper sample collector onto toilet seat according to instructions printed on paper collection device (***do not urinate on collection device***).
2. Following bowel movement, unscrew the lid of the sample collection tube – the lid has a scoop attached, using the scoop, collect a ***large*** scoopful of stool from the collection hat.
3. Re-screw the lid tightly on the sample collection tube.
4. Remaining stool and paper collection device may be flushed down the toilet (see instructions on collection device).
5. Wash your hands with soap and warm water for at least 30 seconds.
6. Please write the ***date of sample collection*** on the label of the sample collection tube.

### Shipping of samples

1. Following sample collection, place the stool collection tube in the ***biosafety bag provided*** and place these into the ***ziploc bag provided***, seal both bags.
2. Place the ziploc bag, frozen ***ice pack, and absorbent sheet*** into the silver ***insulated envelope,*** seal the envelope.
3. Place ***the insulated envelope containing the samples into the pre-paid mailer,*** seal the mailer.
4. Place the pre-paid mailer containing the samples in ***any FedEx*** dropbox ***as soon as possible*** following sample collection to ensure ***next day delivery to our*** laboratory.

***Please remember: In order to avoid sample delivery on a weekend, please only collect and mail samples from Monday through Wednesday.***
